# Genetic mechanisms of 184 neuro-related proteins in human plasma

**DOI:** 10.1101/2023.02.10.23285650

**Authors:** Linda Repetto, Jiantao Chen, Zhijian Yang, Ranran Zhai, Paul R. H. J. Timmers, Ting Li, Emma L. Twait, Sebastian May-Wilson, Marisa D. Muckian, Bram P. Prins, Grace Png, Charles Kooperberg, Åsa Johansson, Robert F. Hillary, Eleanor Wheeler, Lu Pan, Yazhou He, Sofia Klasson, Shahzad Ahmad, James E. Peters, Arthur Gilly, Maria Karaleftheri, Emmanouil Tsafantakis, Jeffrey Haessler, Ulf Gyllensten, Sarah E. Harris, Nicholas J. Wareham, Andreas Göteson, Cecilia Lagging, Mohammad Arfan Ikram, Cornelia M. van Duijn, Christina Jern, Mikael Landén, Claudia Langenberg, Ian J. Deary, Riccardo E. Marioni, Stefan Enroth, Alexander P. Reiner, George Dedoussis, Eleftheria Zeggini, Adam S. Butterworth, Anders Mälarstig, James F. Wilson, Pau Navarro, Xia Shen, the SCALLOP Consortium

## Abstract

Understanding the genetic basis of neuro-related proteins is essential for dissecting the disease etiology of neuropsychiatric disorders and other complex traits and diseases. Here, the SCALLOP Consortium conducted a genome-wide association meta-analysis of over 12,500 individuals for 184 neuro-related proteins in human plasma. The analysis identified 117 cis-regulatory protein quantitative trait loci (cis-pQTL) and 166 trans-pQTL. The mapped pQTL capture on average 50% of each protein’s heritability. Mendelian randomization analyses revealed multiple proteins showing potential causal effects on neuro-related traits as well as complex diseases such as hypertension, high cholesterol, immune-related disorders, and psychiatric disorders. Integrating with established drug information, we validated 13 combinations of protein targets and diseases or side effects with available drugs, while suggesting hundreds of re-purposing and new therapeutic targets for diseases and comorbidities. This consortium effort provides a large-scale proteogenomic resource for biomedical research.

---

Neuropsychiatric disorders are among the leading causes of life-long disability globally, affecting around 800 million people ^1, 2^. As of 2022, mental health remains a global crisis and priority brought to the forefront of public health discussions anew, after the impact of COVID-19 on people’s lives, where stressors such as isolation, significant changes in habits, and global enhanced mortality and fear of contracting the disease have had severe consequences on mental well-being^3–5^. Such disorders represent a significant challenge for medical research due to the high complexity of their neurobiological mechanisms and heterogeneity of symptoms which often overlap with other neurological, psychiatric, and non-psychiatric disorders^6–8^.

In the past decade, genome-wide association studies (GWAS) have been successful in identifying numerous genetic variants that can partially account for variation in complex traits and diseases ^9, 10^. However, the effect of a genetic variant such as a single nucleotide polymorphism (SNP) on a complex disease is usually very small and often does not provide information on the phenotype’s molecular architecture. Measuring proteins may overcome this obstacle as proteins are the product of translated DNA and functional elements that bridge the genetic codes and disease outcomes. Circulating proteins in blood plasma originate from various organ tissues and cell types in the human body and have fundamental roles in different biological processes^**?**, 11, 12^. Thus, such proteins are often used in clinical practice as disease biomarkers. Circulating neurology-related proteins have the potential to provide insight into the pathophysiology of neurological and mental disorders and the genetic architecture of their molecular pathways, setting the basis for the improvement of diagnostic instruments and targeted therapy^13^.

Protein levels are more linked to variation in cognitive function than genetic variants alone. Current studies on neurology-related proteins either focussed on neurodegenerative disorders or cognitive function specifically or had a limited sample size ^14–19^. In a recent study, neurology-related proteins were associated with general fluid cognitive abilities in late life, and a portion of these was observed to be mediated by brain volume, measured as a structural brain variable ^17^.

The field of proteomics has been rapidly expanding in recent years and produced results that have played a fundamental role in the decoding process of molecular mechanisms involved in several traits and diseases, from cardiovascular disease to general health ^16, 20–23^. The genomic studies of the human proteome have benefited from various high-throughput measurement techniques, such as mass spectrometry^24, 25^, aptamer-based assays^26^, and antibody-based assays^12^. Among these, the antibody-based Proximity Extension Assay ^27^ has high measurement precision, especially for many functional but low-abundant proteins.

This study aims to identify genetic variants associated with 184 neurology-related blood circulating proteins via a large-scale genome-wide association meta-analysis (GWAMA) and investigate the proteins’ genetic and potential causal relationships with common psychiatric disorders and comorbidities. We provide an atlas for the genetic architecture of these proteins as a resource for future biological and drug target investigations.

## Results

### GWAMA identified 283 loci associated with 184 neuro-related proteins

In the discovery phase, we conducted a GWAMA using data from up to 12,176 individuals (mean age = 61.9, percentage females = 44.6%) for 92 proteins in the Olink©Neurology panel, and up to 5013 individuals (mean age = 49.6, percentage females = 56.1%, see Supplementary Tables 11-23 for details) for 92 proteins in the Olink©Neuro-Exploratory panel, from a total of twelve participating cohorts (Supplementary Tables 12-23). Overall, we identified 284 top variants distributed across a total of 117 cis-pQTL and 166 trans-pQTL with the significance threshold of *P* < 5 × 10^−8^ for the cisloci and to *P* < 1.76 × 10^−10^ for the trans-loci (Supplementary Table 1, Supplementary Fig. 9-10). Out of the 137 proteins with detected pQTL, 68 proteins had significantly associated variants both in cis- and trans-regulatory loci.

As expected, the identified trans-pQTL, in general, were more weakly associated than the cis-pQTL, nevertheless, we found that 24 proteins shared a total of 14 trans-pQTL. For example, well-known pleiotropic loci such as the *HLA* region and the *ABO* locus showed trans-regulatory effects across a number of plasma proteins (**Fig. 1a**). For instance, 19 proteins showed significant trans-pQTL at the *ABO* locus, nevertheless, the associations were not completely due to the same causal variants (Supplementary Fig. 3). Most of the mapped pQTL were also found to be expression QTL (eQTL) significantly associated with the expressions of the corresponding/nearest genes, however, compared to trans-pQTL, cis-pQTL were much more likely to colocalize with eQTL, in terms of the underlying genetic regulation (Supplementary Fig. 1-2). The lead variants of the cis-pQTL were also more centered around the transcription start sites (TSS) of the corresponding coding genes, compared to those of the trans-pQTL around the TSS of the nearest coding genes (Fig. 1b). The cis-pQTL also had stronger effects, less correlated with the minor allele frequencies (MAFs), compared to the trans-pQTL (**Fig. 1c-d**).

**Figure 1:**
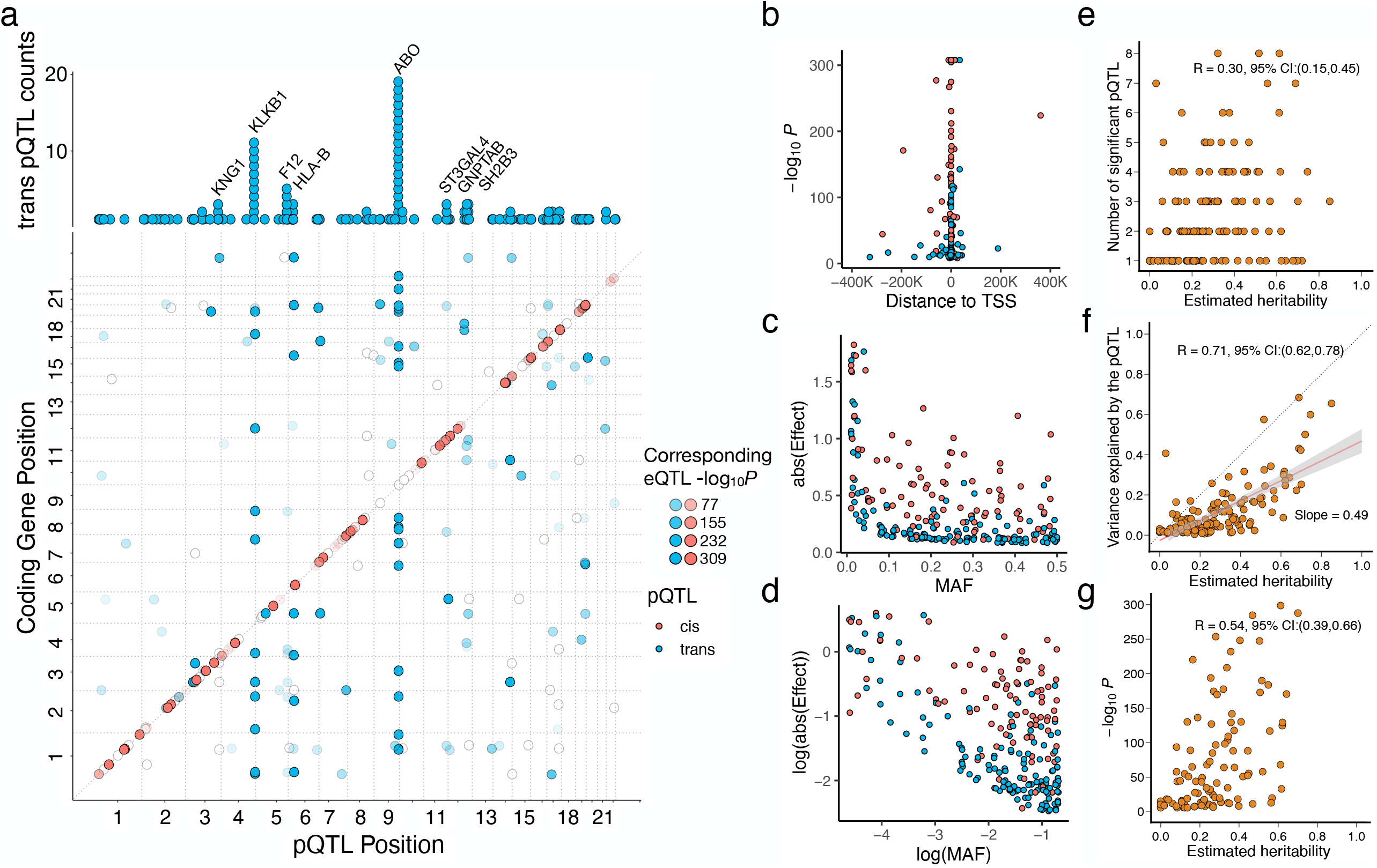
Overview of the mapped protein quantitative trait loci (pQTL). **a**. Pleiotropic trans-pQTL counts and overlap of the mapped pQTL with existing eQTL. The upper barplot shows the number of proteins share trans-pQTL (gene annotations based on gene closest to the trans-pQTL). The scatterplot shows the genomic location of significant cis-pQTL in red(*P* < 5×10^−8^), significant trans-pQTL in blue (*P* < 5 × 10^−8^*/*184), and the shading within the dots indicates significance of the corresponding/nearest cis-eQTL for the respective protein. **b**. Scatterplot of the pQTL lead variants association signals v.s. their distance to the transcription start site (TSS) of the corresponding/nearest coding genes. **c**. Scatterplot of the absolute estimated genetic effects of the pQTL lead variants v.s. their minor allele frequencies (MAFs). **d**. The scatterplot in **c** shown in logarithm scale. **e**. Number of mapped pQTL per protein v.s. the linear mixed model estimated heritability in the ORCADES cohort. **f**. The variance explained by the mapped pQTL summed up for each protein v.s. the estimated heritability. **g**. For the proteins with significant cis-pQTL mapped, the lead variant signal strength v.s. the estimated heritability of each protein.

The fact that the trans-pQTL were not colocalized with eQTL could be partly due to the weaker signals of the trans-pQTL than those of the cis-pQTL. However, we hypothesized that the trans-pQTL may not necessarily reflect the biological regulatory mechanisms of the corresponding proteins, but rather driven by underlying features of the blood samples, due to their influence on the immuno-reaction of the Olink assay. For example, the pleiotropic trans-pQTL across the proteins highlight major blood coagulation and clotting factors such as KLKB1 (Plasma kallikrein), KNG1 (Kininogen-1), and F12 (Coagulation factor XII), as well as glycosylation locus *ST3GAL4*. We thus also looked into the functional pathways and gene sets that involve the closest genes to our transpQTL, using the gene set enrichment analyses (Supplementary Fig. 6). With a false discovery rate < 5%, 997 significant pathways were found to be enriched for the genes of our trans loci, of which 443 (44.4%) were driven or partly driven by the HLA genes. Most top enriched pathways were clustered into inflammatory and immune responses, coagulation processes, cell-to-cell signaling and adhesion, and protein glycosylation (Supplementary Table 8). Particularly, the trans-pQTL were found to be enriched in 1) established GWAS traits such as blood protein levels, platelet count, and platelet crit; 2) GO pathways such as biological adhesion, wound healing, coagulation, and glycosylation; 3) Hallmark gene sets including coagulation; 4) Reactome pathways including hemostasis and clotting formation; 5) microRNA targets and Wiki pathways for blood clotting cascade.

We assessed the overall heritabilities across the 184 analyzed plasma proteins. Summary statisticsbased methods have been developed to infer heritability and genetic correlation parameters for complex traits with GWAS results; however, consistent estimates can only be obtained for genetic correlations ^28–30^. Thus, we used a standard polygenic mixed model on the individual-level data collected in the ORCADES cohort to assess the narrow-sense heritability for each protein ^31^. Across the analyzed proteins, we found that the higher the protein’s heritability, the more pQTL detected for the protein (**Fig. 1e**), the stronger the cis-pQTL effects are (**Fig. 1g**), and the higher amount of phenotypic variance captured by the detected pQTL (**Fig. 1f**). On average, the mapped pQTL together explain 49% of the proteins’ heritability. This indicates that proteins as molecular phenotypes have strong major regulatory loci. Nevertheless, their genetic effects can still be widespread across the genome, having a polygenic genetic architecture.

Using data from the ORCADES cohort, we found TDGF1 (Teratocarcinoma-Derived Growth Factor 1) to have the highest heritability (*h*^2^ = 0.85), followed by MDGA1 (MAM Domain-Containing Glycosylphosphatidylinositol Anchor Protein 1, *h*^2^ = 0.75), CLM1 (CD300 Molecule Like Family Member F, *h*^2^ = 0.72), and LAIR2 (Leukocyte Associated Immunoglobulin Like Receptor 2, *h*^2^ = 0.70). In contrast, CTF1 (Cardiotrophin 1), EPHA10 (Ephrin Type-A Receptor 10), GSTP1 (Glutathione S-Transferase Pi 1), HSP90B1 (Heat Shock Protein 90 Beta Family Member 1), IFI30 (Gamma-Interferon-Inducible Lysosomal Thiol Reductase), NDRG1 (N-Myc Downstream Regulated 1) and SFRP1 (Secreted Frizzled Related Protein 1) all had an estimated *h*^2^ value of 0, while having at least one pQTL.

We used the PhenoScanner pQTL database ^32, 33^ to determine whether the pQTL sentinel variants or variants in linkage disequilibrium (LD) with them (*r*^2^ > 0.8) that we identified had been previously found to be significantly associated with the corresponding proteins (Supplementary Table 2). 113 of our discovered loci were already discovered in previous studies. We also checked whether the hits from the meta-analysis were significant in the individual cohorts and observed that 73 of the sentinel variants were found to be statistically significant only in the meta-analysis. We also extracted the established associations between our mapped cis-pQTL and complex traits from the PhenoScanner database (Supplementary Table 3). At a 5% false discovery rate, 39 cis-pQTL showed significant association with both complex traits and other proteins (mostly based on an aptamer-based assay). We found that the level of pleiotropy at the protein level, i.e., being trans-pQTL for other proteins, is associated with the level of pleiotropy on the complex traits (Supplementary Fig. 4).

We performed linkage disequilibrium (LD) pruning(*r*^2^ < 0.001) to identify secondary independent associations at the cis-pQTL. We identified a total of 769 additional variants across all the 117 proteins with cis-pQTL mapped (Supplementary Table 4). Furthermore, we cross-referenced the significant loci discovered in the meta-analysis with the currently available pQTL data from the UK Biobank Pharma Proteomics Project^34^. We observed that 174 out of our 283 discovered loci were not reported in the UK Biobank significant results, including loci found in both cis- and trans-regions of the respective proteins (Supplementary Table 3).

### Mendelian randomization analysis identifies plausible causal protein markers for neuro-related phenotypes

In order to make statements on potential causality from the proteins to complex traits and diseases, we focused on the genetic associations at the cis-pQTL, which provide strong and most likely valid genetic instruments in Mendelian randomization (MR) analysis. We first considered the 152 neuro-related traits whose GWAS summary statistics are available through LD-Hub^35^ as the outcome data. We performed an inverse-variance weighted (IVW) two-sample MR analysis using the 886 LD-pruned genetic instruments across the 117 cis-pQTL on the 152 phenotypes. With a false discovery rate 5% threshold, we obtained 24 significant potential causal associations for 13 proteins on 22 traits, where three proteins are currently druggable targets (**Fig. 2**, Supplementary Table 5).

**Figure 2:**
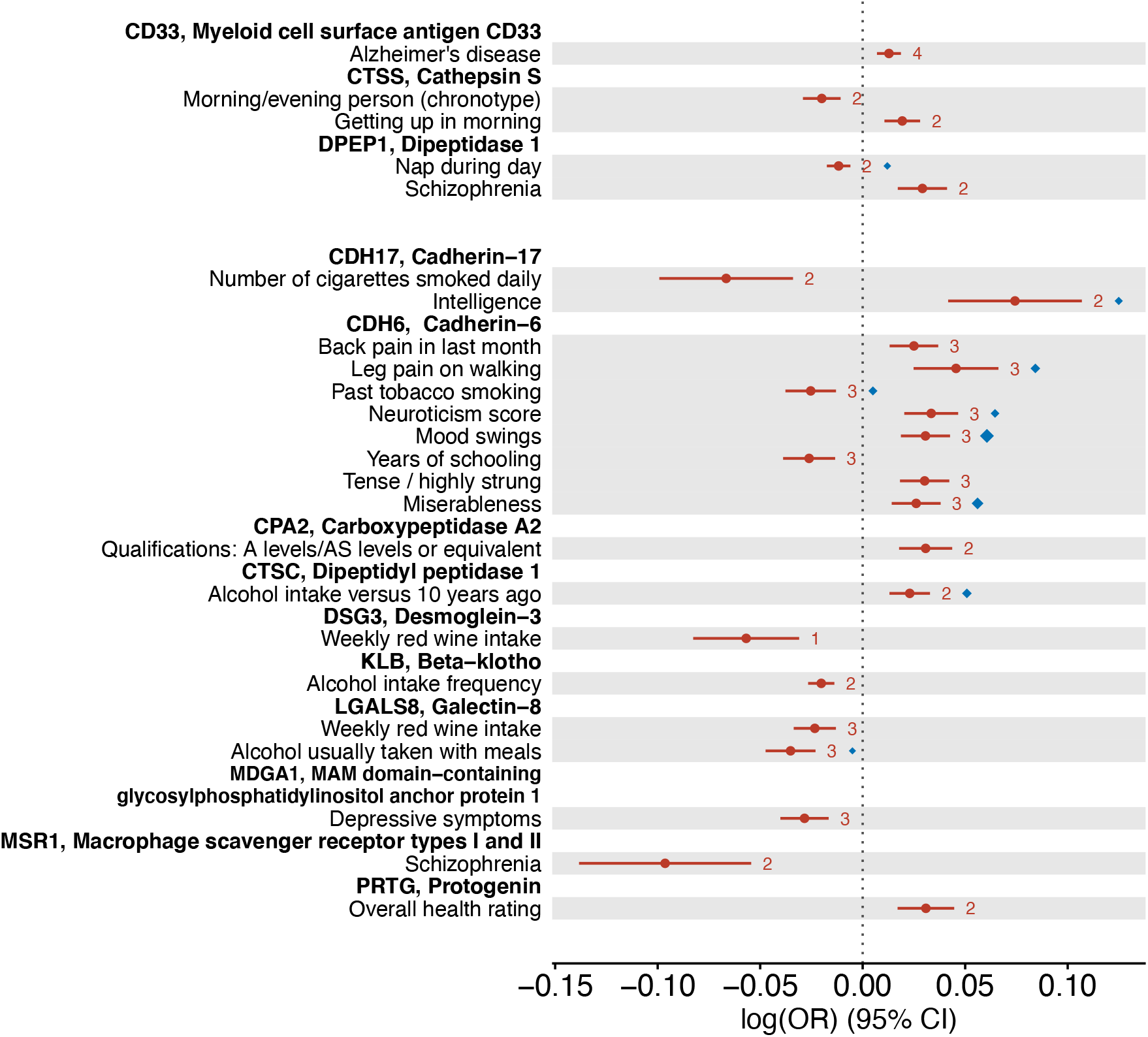
Causality between the proteins and neuro-related phenotypes inferred by Mendelian randomization (MR) analyses. The forest plot shows the significant MR results (false discovery rate < 0.05) based on LD-pruned (*r*^2^ < 0.001) instrumental variants within each cis-pQTL. Inversevariance weighted (IVW) estimates are provided as the solid round dots, and the whiskers indicate 95% confidence intervals. The numbers of instrumental variants in the cis-pQTL are given to the right of the whiskers. As a colocalization measure, the HEIDI (heterogeneity in dependent instruments) test evidence (*p >* 0.05) are given as the diamonds, where the largest diamonds correspond to a p-value of 1. The upper part of the plot shows the results where the proteins are known druggable targets, while the lower part shows the results for new protein targets.

In order to control for false positive inference due to LD, we adopted the HEIDI (heterogeneity in dependent instruments)^36^ test statistic to examine the colocalization between each pQTL and its association with the corresponding downstream outcome phenotypes. Nine out of the 24 plausible causal associations had colocalization support by HEIDI (*p* > 0.05) (**Fig. 2-3**, Supplementary Table 5). Among these, the single protein CDH6 showed a potential causal effect on neurological and behavioral traits including mood swings, miserableness, leg pain, smoking, and neuroticism, where the effect on smoking had a different direction compared to on the others. CTSC and LGALS8 were both plausible causal markers for alcohol intake but with opposite effects directions. CDH17 showed an positive effect on intelligence. DPEP1 showed a negative effect on napping, while as a druggable target it also showed a potential risk-increasing effect on schizophrenia.

**Figure 3:**
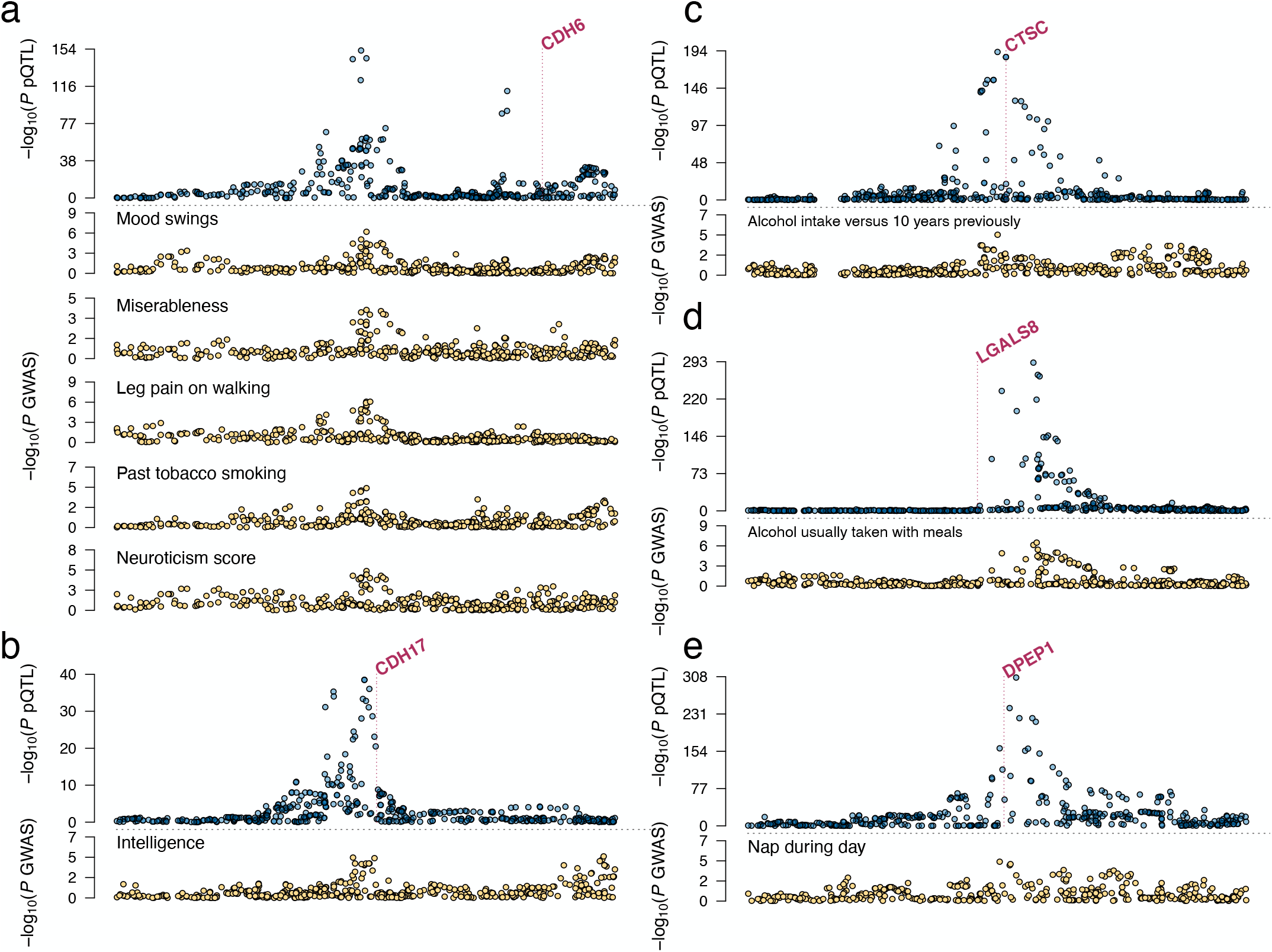
Regional association patterns of the pQTL and the colocalized neuro-related complex traits. The displayed protein-trait pairs correspond to the Mendelian randomization discoveries in Figure 2 with the HEIDI p-value > 0.05. Each subfigure shows the pQTL region of 1Mb centered at the lead variant. The vertical dashed line in each subfigure marks the transcription start site of the corresponding protein’s coding gene.

### Mendelian randomization analysis provides evidence for the proteins’ causal effects on other complex diseases

Expanding our cis-pQTL-based MR analysis to a broader range of complex traits, we used the UK Biobank GWAS summary-level data for 4,085 phenotypes by the Neale’s lab (see Data Availability) as the outcome data. We performed the same analysis procedure as above, and with a false discovery rate 5% threshold, the analysis yielded in 472 significant potential causal associations for 82 proteins on 221 traits. Among these discoveries, 59 were for 47 diseases with 33 plausible causal protein markers.

Again, we utilized the HEIDI test statistic to examine the colocalization between each pQTL and the disease genetic associations. 29 out of the 59 plausible causal associations with disease outcomes showed colocalization supported by HEIDI (*p* > 0.05) (**Fig. 4**, Supplementary Table 6), including 8 druggable protein targets and 14 new targets.

**Figure 4:**
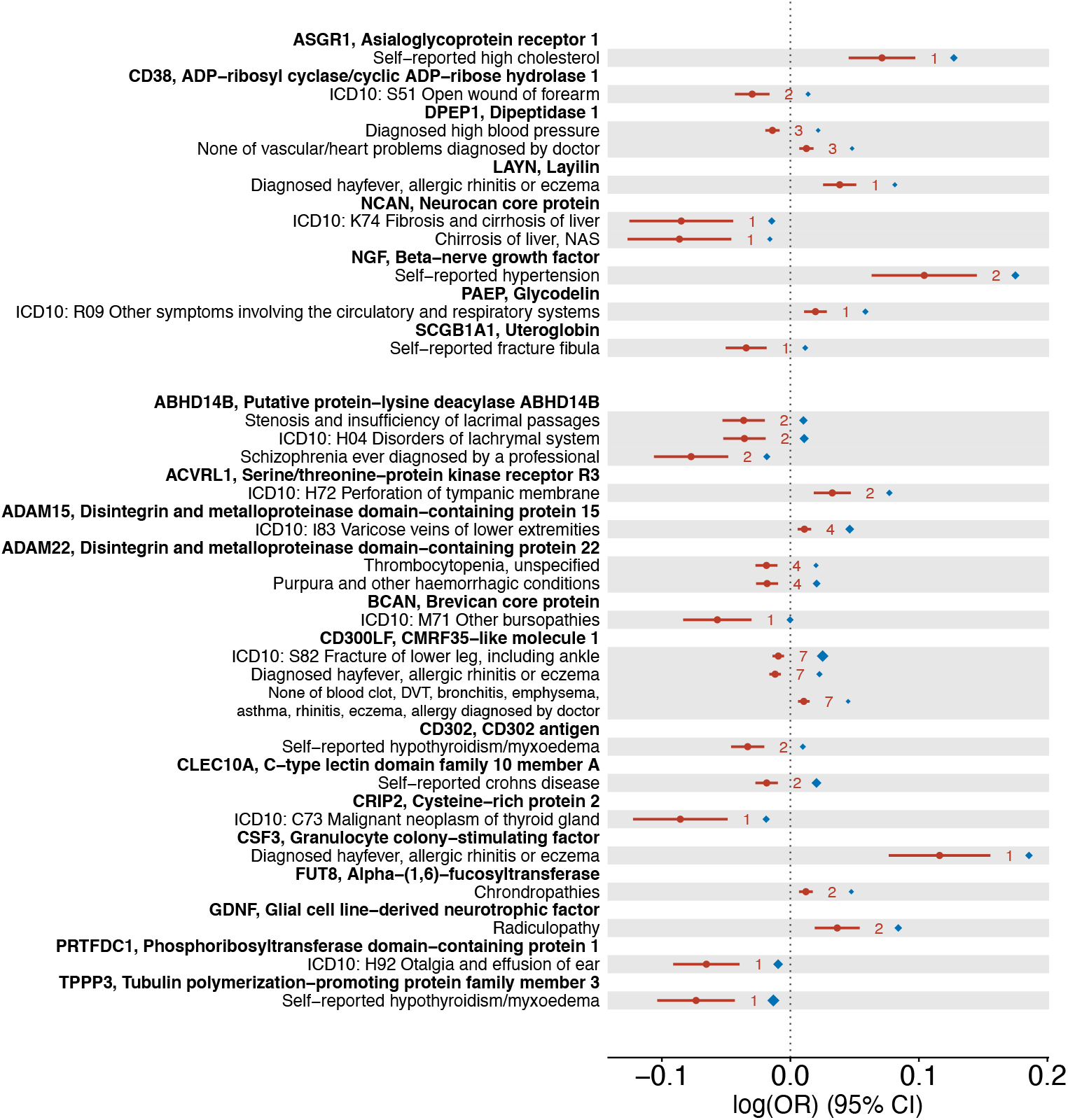
Causality between the proteins and UK Biobank disease phenotypes inferred by Mendelian randomization (MR) analyses. The forest plot shows the significant MR results (false discovery rate < 0.05) based on LD-pruned (*r*^2^ < 0.001) instrumental variants within each cis-pQTL. Inversevariance weighted (IVW) estimates are provided as the solid round dots, and the whiskers indicate 95% confidence intervals. The numbers of instrumental variants in the cis-pQTL are given to the right of the whiskers. As a colocalization measure, the HEIDI (heterogeneity in dependent instruments) test evidence (*p* > 0.05) are given as the diamonds, where the largest diamonds correspond to a p-value of 1. The upper part of the plot shows the results where the proteins are known druggable targets, while the lower part shows the results for new protein targets.

Except for the effect of TPPP3 (tubulin polymerization−promoting protein family member 3) on hypothyroidism/myxoedema, reverse generalized summary-statistics-based MR (GSMR)^37^ did not show evidence for reverse causality of the other significant MR discoveries on the complex diseases. In general, the MR estimated odds ratios (FDR < 0.05) were found to be ranging from 0.49 to 2.48, consistent with previous studies evaluating the causal effects of blood circulating proteins on other complex traits ^12, 38^.

### Mendelian randomization reveals established, re-purposing, and new drug targets for complex diseases and comorbidities

Based on the MR causal inference, we systematically investigated the protein markers in the DrugBank database (see Data Availability). There were 13 protein-trait combinations from the significant MR discoveries that matched established drugs. We found that for all the 13 established drug targets (**Fig. 5a-b**), the MR-inferred causal effects directions matched the corresponding targeting drugs’ pharmacological effects (including side effects) (**Fig. 5c**). For instance, hyaluronic acid is a liver disease biomarker, the protein NCAN binds with hyaluronic acid thus reduces liver chirrosis. Gemtuzumab ozogamicin is a monoclonal anti−CD33 antibody, reducing white blood cell count. Benralizumab is an antibody for IL5RA, treating eosinophilic asthma by affecting its causal effect on eosinophill counts. Overdosed acetaminophen increases the mean corpuscular volume and mean corpuscular haemoglobin, due to the insufficient enzyme activity of Glutathione S-transferase P (GSTP1).

**Figure 5:**
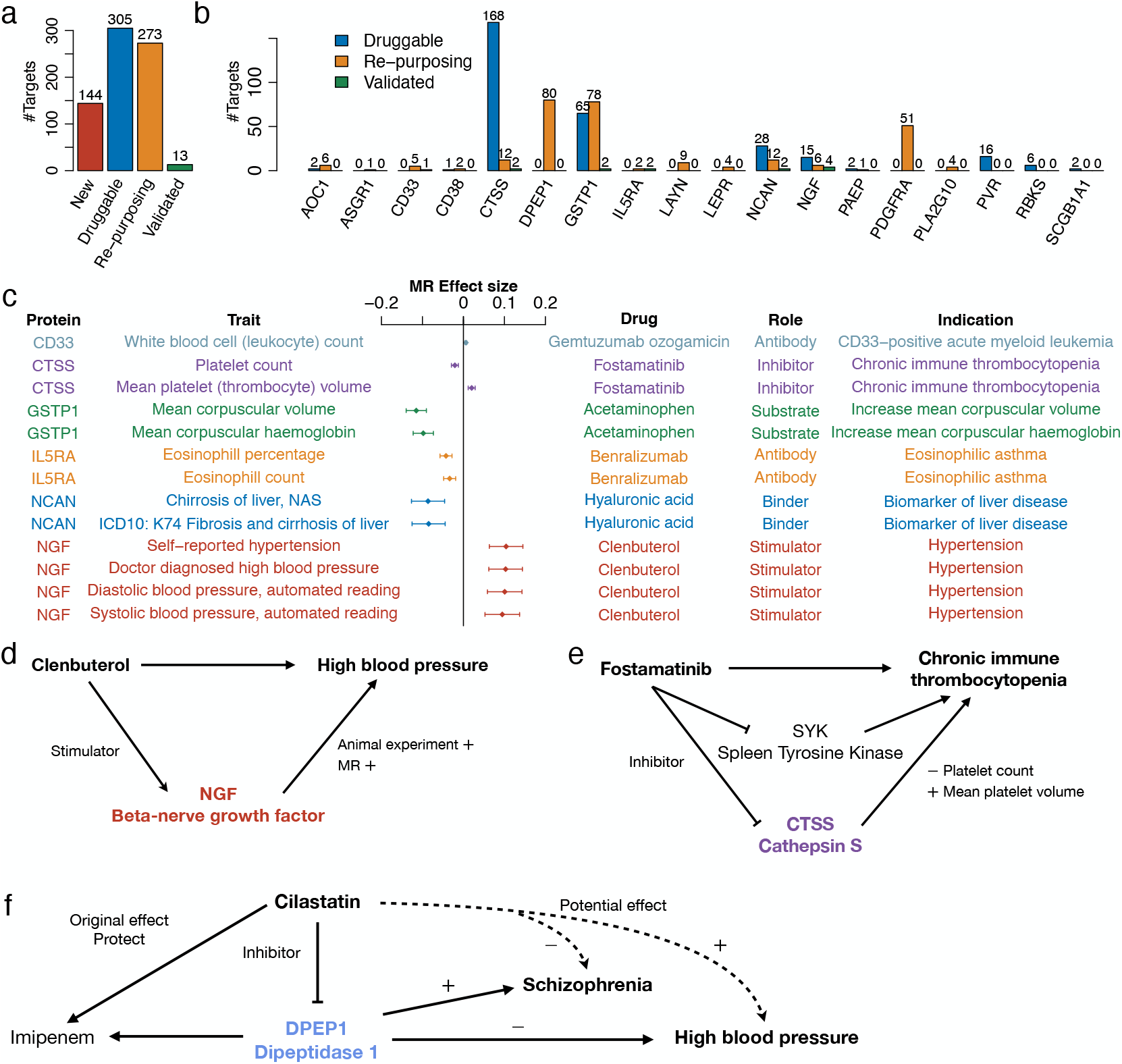
Drug targets revealed by Mendelian randomization (MR) analyses. The MR results with 5% false discovery rate are considered. **a**. The number of MR inferred pairs of proteins and traits split into four categories: new (drug) targets, druggable targets that have drugs with unclear clinical function, re-purposing targets that have established drugs but for different diseases, and validated known targets where the established drugs have pharmacological effects that match the MR results. **b**. Numbers of re-purposing and validated drug targets per protein analysed. **c**. The validated known drug targets, the description of the drugs, and the corresponding consistent MR estimated effects. **d**. Potential mechanism of the adverse effect of Clenbuterol that targets NGF. **e**. Potential mechanism of Fostamatinib treating Chronic immune thrombocytopenia through CTSS. **f**. Potential pharmacology of DPEP1’s re-purposing drug on schizophrenia.

Clenbuterol was used as a bronchodilator in the treatment of asthma patients. But it can cause long and short-term side effects, including hypertension. Our MR analysis showed that the increased level of beta-nerve growth factor (beta-NGF), which could be caused by Clenbuterol, could lead to a higher risk of hypertension (**Fig. 5d**).

The MR analysis reveals that protein CTSS (cathepsin S) can increase platelets in the blood and reduce mean platelet volume. Fostamatinib can inhibit the protein CTSS, known as an approved medication for chronic immune thrombocytopenia (ITP) by inhibiting the spleen tyrosine kinase (SYK). It indicates that fostamatinib treats ITP via both protein SYK and CTSS (**Fig. 5e**).

Cilastatin is a dehydropeptidase 1 (DPEP1) inhibitor used to prevent degradation of imipenem, both were used together to treat infections. We found that inhibiting DPEP1 can increase the risk of high blood pressure, while decrease the risk of schizophrenia (**Fig. 5f**). This indicates clinical re-purposing potential of Cilastatin, and other DPEP1 inhibitors, as treatments for schizophrenia, though further investigations are needed.

Overall, besides the validated targets, we also identified 273 suggestive drug re-purposing target-disease pairs for 18 proteins (**Fig. 5a-b**, Supplementary Table 9). There already exist established drugs for these protein targets, making these drugs potentially useful upon further clinical trials. At last, 144 new target-disease combinations were suggested, based on our causal inference (Supplementary Table 10).

## Discussion

We identified novel pQTL for 137 of 184 neuro-related proteins, provided insights into their molecular mechanisms and effects on complex diseases and traits, and highlighted useful therapeutic targets with established drugs. On average, we identified half of the genetic architecture underlying the concentration of these proteins. We provide a well powered genetic landscape for these proteins with large-scale summary-level data for future research.

This meta-analysis within our SCALLOP collaborative framework is a follow-up of a previous study on the proteins from the Olink Neurology and Neuro-exploratory panels, where data were collected from the two Greek cohorts that we included in this study^39^. Our results replicated over 90% of the established loci, including the previous main discoveries of the cis-pQTL for CD33, GP-NMB, and MSR1.

Although the proteins were found to have small effects individually in the MR analysis, our results indicated that for most of the identified proteins, having low levels in plasma leads to a higher chance of having poorer health conditions (Supplementary Fig. 5). These conditions include both deterioration of mental health and related non-neurological comorbidities. Such results on the neuro-related proteins are consistent with the notion that psychiatric and neurological disorders are multi-factorial and not limited to the central nervous system, but rather are products of interactions among multiple systems within the organism^40–43^. The intertwining of neuropsychiatric, inflammatory, and cardiovascular disorders has long presented a challenge in clinical research due to the difficulties in discerning the relationships among them^44, 45^. Our results suggest that these disorders may share molecular mechanisms and pathways and provide the basis for developing new diagnostic tools and treatment strategies. We also reported a large number of drug re-purposing targets, suggesting the potential use of established drugs in new clinical trials for treatment of different symptoms and disorders.

Regarding the MR methodology, we found that the MR analysis with a single genetic instrument at the cis-pQTL tended to generate stronger estimated causal effect (**Fig. 4**). This indicates: 1) a single genetic instrument analysis may be more prone to winner’s curse, i.e., more likely to detect an overestimated effect; 2) using multiple independent instruments within a locus may not only improve power but also control false discoveries due to winner’s curse.

As expected, the mapped trans-pQTL did not show good colocalization with nearby genes, and they were enriched in blood clotting and coagulation pathways. For instance, a blood clotting factor *KLKB1* appeared to be a trans-regulatory hub for multiple proteins. We thus infer that some of the trans-pQTL discovered are not directly involved in the genetic mechanisms of the correspond-Ing proteins, but rather they regulate blood characteristics that affect the performance of the antibody-based assays. This is an important discovery for biotechnological development in proteomics, suggesting that the features of the plasma samples could be non-negligible factors in circulating protein quantification.

In summary, this study provides fundamental new knowledge of the genetics of many neuro-related proteins, moving towards drug discovery with new protein and pathway targets. Future work can include results from more cohorts and the UK Biobank Pharma Proteomics Project. An even larger multi-cohort meta-analysis and replication analysis can seek insights into the secondary signals in the discovered pQTL. From our pQTL discovery and causal inference with disease outcomes, clinical studies can investigate actionable drug targets and integrate them into multi-omics analyses.

## Methods

### Proteins

This study focussed on proteins from the Olink Neurology and Olink Neuro-exploratory panels. Circulating protein levels were quantified using Proximity Extension Assay technology, consisting of pairs of oligonucleotides-labelled antibodies to bind target proteins and hybridize to have their sequence extended and amplified through polymerase chain reaction (PCR). The level of amplified DNA is then quantified by microfluidic qPCR^27^.

Proteins were selected by a panel of experts to include protein biomarkers that are known to be associated with neurological disorders and conditions through existing literature. The functions of these proteins comprise axonal development, metabolism, immune response, and cell-to-cell communication. The proteins have been included in their respective panel on the basis of their observed involvement in neurological conditions and disorders, as well as the general performance of the assay.

### Cohorts and data collection

We obtained summary statistics from the GWAS analyses performed on the Olink Neurology proteins from 10 cohorts and the Olink Neuro-exploratory proteins from 6 cohorts. Cohorts comprised population-based and case-control studies. The summary statistics information for each cohort can be found in Supplementary Tables 11-25. The total sample size for the Neurology panel meta-analysis was 12,176, whereas the Neuro-exploratory panel meta-analysis included up to 5,013 individuals. The participating cohorts used whole-genome sequencing data or imputed data using the 1000 Genomes Project (Phase1 and Phase3) or the Haplotype Reference Consortium (HRC) as reference panels. An average of 14.5 million SNPs were tested per protein, and the lowest per-SNP filter imputation quality ranged from 0.4 to 0.3 depending on the cohort. Each cohort carried out quality control according to their study design, as reported in Supplementary Table 11.

Data below the Olink limit of detection (LOD) is calculated based on the negative controls included in each PCR run. Data below the LOD was available only for some cohorts participating in the meta-analysis. As the proteins were quantified at different times across cohorts, not all studies have data on all proteins in the two Olink panels.

### Genome-wide association analysis of the proteins

The Normalized Protein expression values (NPX), Olink’s unit of protein abundance level on a log2 scale ^27^, were rank-based inverse normal transformed before running the per-protein GWAS analyses. Genotypic data were the allelic dosages resulting from imputation using the Haplotype reference consortium (HRC) or the 1000 genomes data as reference panel. Monomorphic SNPs were excluded. The genotype-phenotype association analysis was performed using regression models adjusting for sex, age, plate number, plate column, plate row, sample time in storage, season of sample collection, population structure (when appropriate), and other study-specific covariates.

### Meta-analysis

The summary association statistics from each participating cohort were uploaded through a secured FTP channel to the University of Edinburgh’s ECDF Eddie Mark 3 cluster. The meta-analysis was run per protein in METAL (version 2018-08-28)^46^ using the inverse variance weighted method. We defined cis-pQTL to be 500kb upstream or downstream of the gene coding for the respective protein and set the trans-pQTL window to be 1Mb around the top variants that were found outside the defined cis-window. A 1% MAF filter was applied to the meta-analysis summary statistics for subsequent analyses. The variants that existed in only one participating cohort were also removed before subsequent analyses. The significance threshold was set to be 5 × 10^−8^ for the top variants of cis-regulatory variants and 5×10^−8^*/*184 = 2.73×10^−10^ for the variants in trans-regions.

### Heritability analysis

We used a standard polygenic mixed model implemented in GenABEL^31^ on the individual-level data collected in the ORCADES cohort to assess the narrow-sense heritability for each protein. The heritability captured by each pQTL is calculated as 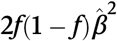, where *f* and 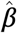 are the coding allele frequency and estimated genetic effect, respectively, assuming Hardy-Weinberg equilibrium.

### Established genetic associations

We used PhenoScanner v2^32, 33^ to cross-reference the lead (most significant) genetic variants in the cis-pQTL from our meta-analysis with other phenotypes. PhenoScanner is an extensive database of over 65 billion associations from publicly available GWAS studies. We used the lead variants of our cis-loci as input without the additional option of using proxy markers. When checking the novelty of our mapped cis-pQTL, we consider established pQTL associations with *P* < 5 × 10^−6^ as known. When extracting the established complex traits associations, we set the p-value threshold to 1 to include all possible associations. Thereafter, results with false discovery rate less than 0.05 are considered. We excluded the studies with non-European ancestry.

### Gene set enrichment and functional annotation of GWAS trans loci

We performed our gene set enrichment analyses using the GENE2FUNC in FUMA v1.3.7 ^47, 48^, which returns functional annotation to ENSEMBL v92 gene models for the submitted list in a biological context. We identified the genes closest to the top SNPs in our trans loci using the locuszoom v0.12^49, 50^ database and then submitted the list of genes to the FUMA website. We selected all types of genes to use as background for this analysis, including over 57,000 genetic elements. We set the maximum FDR adjusted p-value for gene set association to 1.

### Mendelian randomization analysis

We performed a two-sample Mendelian randomization (MR) analysis using the inverse-variance weighted (IVW) method to evaluate causal effects between the proteins with genome-wide significant cis-pQTL and the traits from the UK Biobank GWAS results by the Neale’s lab. Multiple sentinel variants of our cis-pQTL after LD pruning (*r*^2^ < 0.001) were used jointly as instrumental variables. We report the significant discoveries at a level of 5% false discovery rate, for which we also performed a reverse generalized summary-statistics-based MR (GSMR) from the complex trait exposures to protein outcomes.

### Colocalization analysis

For the MR-positive discoveries, the pQTL-complex-trait colocalization analysis was performed using the SMR/HEIDI tool in the GCTA software^36^. We considered a pair of QTL associations to be colocalized if the HEIDI test p-value was greater than 0.05.

For eQTL-pQTL colocalization analysis, we adopted the v7 release of both the GTEx eQTL and eQTLGen summary-level data. We used the Bayesian colocalization analysis tool `coloc`, with the posterior probabilities testing the H4 colocalization hypothesis, which tests for one shared variant between the pair of corresponding eQTL and pQTL^51^. For each cis-pQTL, we tested colocalization with the cis-eQTL of the corresponding coding gene in each tissue. For each trans-pQTL, we tested colocalization with the cis-eQTL of the nearest coding gene.

### Drug target investigation

For the protein markers from IVW MR results with false discovery rate less than 5%, we systematically investigated available drugs targetting these markers using the Drug-Bank database. We considered a drug target validated if an MR discovery between the protein marker and the trait/disease suggested the same effect direction as the drug’s effect on the protein target. The protein targets that have available drugs but not directly related to the MR discovered outcomes were regarded as re-purposing targets. The remaining MR discoveries were reported as new targets.

## Data Availability

The full genome-wide summary association statistics for the 184 proteins will be made publicly available upon publication of the paper; GTEx data: https://gtexportal.org/home/datasets; 1000 Genomes phase 3 genotype data: https://www.cog-genomics.org/plink/2.0/resources#phase3_1kg; Neale lab UK Biobank round2 GWAS summary-level data: http://www.nealelab.is/uk-biobank; DrugBank: https://www.drugbank.com.

## Code availability

METAL: https://genome.sph.umich.edu/wiki/METAL_Documentation;PLINK: https://www.cog-genomics.org/plink/; GCTA-GSMR: https://yanglab.westlake.edu.cn/software/gcta/#GSMR; PhenoScanner: http://www.phenoscanner.medschl.cam.ac.uk; SMR & HEIDI: https://yanglab.westlake.edu.cn/software/smr/#SMR&HEIDIanalysis;FUMA: https://fuma.ctglab.nl.

## Data availability

The full genome-wide summary association statistics for the 184 proteins will be made publicly available **upon publication of the paper**; GTEx data: https://gtexportal.org/home/datasets; 1000 Genomes phase 3 genotype data: https://www.cog-genomics.org/plink/2.0/resources#phase3_1kg; Neale’s lab UK Biobank round2 GWAS summary-level data: http://www.nealelab.is/uk-biobank; DrugBank: https://www.drugbank.com.

## Acknowledgements

X.S. was in receipt of Swedish Research Council (Vetenskapsrådet) grants (No. 2017-02543 & No. 2022-01309), a National Natural Science Foundation of China (NSFC) grant (No. 12171495), a Natural Science Foundation of Guangdong Province grant (No. 2114050001435), and a National Key Research and Development Program grant (No. 2022YFF1202105). P.R.H.J.T. and J.F.W. acknowledge support from the Medical Research Council Human Genetics Unit program grant “Quantitative Traits in Health and Disease” (U. MC_UU_00007/10). The work from C.K. and A.P.R. is supported in part by NIH grant R01-HL136574.

We thank the members of the cited consortia of genome-wide association studies for making their data available. Cohort-specific acknowledgements are given in the **Supplementary Information**.

## Author contributions

X.S., P.N., and J.F.W. initiated and coordinated the study. L.R., J.C., Z.Y., and R.Z. performed data analysis. L.R., P.R.H.J.T., E.L.T., P.N., and X.S. contributed to the analysis pipeline. S.M.W., M.D.M., B.P.P., A.J., R.F.H., E.W., S.K., S.A., L.P., S.B., Y.H., G.P., C.K., J.E.P., U.G., S.E.H., N.J.W., C.L., M.A.T., A.G., A.G., M.K., E.T., J.H., A.P.R., G.D., E.Z., M.L., C.M.V.D., C.J., C.L., I.J.D., R.E.M., S.E., A.S.B., and A.M. contributed to the cohort-level analysis. L.R., J.C., Z.Y., R.Z., P.N., and X.S. wrote the paper. All authors approved the submitted version of the paper.

