## Supplementary Information for "Genetic mechanisms of 184 neuro-related proteins in human plasma"

#### Contents

**1 Cohort-specific Acknowledgements** **2**

**2 Supplementary Tables** **5**

**3 Supplementary Figures** **13**

### 1 Cohort-specific Acknowledgements

**HELIC MANOLIS & HELIC POMAK** This work was funded by the Wellcome Trust (098051) and the European Research Council (ERC-2011-StG 280559-SEPI). The MANOLIS cohort is named in honour of Manolis Giannakakis, 1978-2010. We thank the residents of the Mylopotamos villages, and of the Pomak villages, for taking part. The HELIC study has been supported by many individuals who have contributed to sample collection (including A. Athanasiadis, O. Balafouti, C. Batzaki, G. Daskalakis, E. Emmanouil, C. Giannakaki, M. Giannakopoulou, A. Kaparou, V. Kariakli, S. Koinaki, D. Kokori, M. Konidari, H. Koundouraki, D. Koutoukidis, V. Mamakou, E. Mamalaki, E. Mpamiaki, M. Tsoukana, D. Tzakou, K. Vosdogianni, N. Xenaki, E. Zengini), data entry (T. Antonos, D. Papagrigoriou, B. Spiliopoulou), sample logistics (S. Edkins, E. Gray), genotyping (R. Andrews, H. Blackburn, D. Simpkin, S. Whitehead), research administration (A. Kolb-Kokocinski, S. Smee, D. Walker) and informatics (M. Pollard, J. Randall)."

**WHI** The WHI program is funded by the National Heart, Lung, and Blood Institute, National Institutes of Health, U.S. Department of Health and Human Services through contracts 75N92021D00001, 75N92021D00002, 75N92021D00003, 75N92021D00004, 75N92021D00005. C.K. and A.P.R. are supported in part by NIH grant R01-HL136574.

**SAHLSIS** The Sahlgrenska Academy Study on Ischemic Stroke (SAHLSIS) study was supported by the Swedish Research Council (2021-01114), the Swedish State under the agreement between the Swedish Government and the County Councils, the ALF- agreement (ALFGBG-965328), the Swedish Heart and Lung Foundation (20220184), the Swedish Stroke Association, the Gothenburg Foundation for Neurological Research, and the King Gustaf V's and Queen Victoria's Freemasons' Foundation. We thank research nurse Ingrid Eriksson for her excellent assistance in recruiting the study participants and for conducting the follow-up study. Furthermore, we are grateful to our study participants without whom this work would not have been possible.

**SWEBIC** The Swedish bipolar collection (SWEBIC) study was funded by the Stanley Center for Psychiatric Research, Broad Institute from a grant from Stanley Medical Research Institute (Subaward No. 5710002350). This research was supported by grants from the Swedish Medical Research Council (2018-02653), the Swedish Foundation for Strategic Research (KF10-0039), the Swedish Brain Foundation (FO2020-0261), and the Swedish Society for Medical Research. We are deeply grateful for the participation of all individuals contributing to this research. We would also like to thank specific staff who worked with the collection: Marie Lundin, Radja

Satgunanthan-Dawoud, Malin Rådström, Birgitta Ohlander, and Leila Nyrén. Bozenna Iliadou and Mathias Kardell are acknowledged for statistical and database support. Finally, we wish to thank the KI Biobank at Karolinska Institutet for professional biobank service.

**FENLAND** The Fenland Study (10.22025/2017.10.101.00001) is funded by the Medical Research Council (MC_UU_12015/1). We further acknowledge support for genomics and metabolomics from the Medical Research Council (MC_PC_13046). We are grateful to all the volunteers and to the General Practitioners and practice staff for assistance with recruitment. We thank the Fenland Study Investigators, Fenland Study Co-ordination team and the Epidemiology Field, Data and Laboratory teams.

**NSPHS** The Northern Swedish Population Health Study (NSPHS) has been funded by the Swedish Medical Research Council (K2007-66X-20270-01-3, 2011-5252, 2012-2884 and 2011-2354), and the Foundation for Strategic Research (SSF). NSPHS as part of European Special Populations Research Network (EUROSPAN) was also supported by the European Commission FP6 STRP (01947, LSHG-CT-2006-01947). Whole-genome sequencing was funded by SciLifeLab. Computations were performed on resources provided by SNIC through Uppsala Multidisciplinary Center for Advanced Computational Science (UPPMAX) under project sens2016007.

**INTERVAL** Participants in the INTERVAL randomised controlled trial were recruited with the active collaboration of NHS Blood and Transplant England (www.nhsbt.nhs.uk), which has supported field work and other elements of the trial. DNA extraction and genotyping was co-funded by the National Institute for Health Research (NIHR), the NIHR BioResource (http://bioresource.nihr.ac.uk) and the NIHR Cambridge Biomedical Research Centre (BRC) [*]. The academic coordinating centre for INTERVAL was supported by core funding from: NIHR Blood and Transplant Research Unit in Donor Health and Genomics (NIHR BTRU-2014-10024), UK Medical Research Council (MR/L003120/1), British Heart Foundation (SP/09/002; RG/13/13/30194; RG/18/13/33946) and the NIHR Cambridge BRC [*]. A complete list of the investigators and contributors to the INTERVAL trial is provided in reference 58. The academic coordinating centre would like to thank blood donor centre staff and blood donors for participating in the INTERVAL trial.

This work was supported by Health Data Research UK, which is funded by the UK Medical Research Council, Engineering and Physical Sciences Research Council, Economic and Social Research Council, Department of Health and Social Care (England), Chief Scientist Office of the Scottish Government Health and Social Care Directorates, Health and Social Care Research and Development Division (Welsh Government), Public Health Agency (Northern Ireland), British Heart Foundation and Wellcome.

*The views expressed are those of the author(s) and not necessarily those of the NIHR or the Department of Health and Social Care.

**LBC1936** The authors thank all LBC1936 study participants and research team members who have contributed, and continue to contribute, to ongoing studies. LBC1936 is supported by the Biotechnology and Biological Sciences Research Council, and the Economic and Social Research Council [BB/W008793/1], Age UK (Disconnected Mind project), and the University of Edinburgh. Genotyping was funded by the BBSRC (BB/F019394/1). The Olink® Neurology Proteomics assay was supported by a National Institutes of Health (NIH) research grant R01AG054628.

#### 2 Supplementary Tables

**Supplementary Table 1. List of top significant SNPs across both cis- and trans-protein quantitative trait loci (pQTL).**

**Panel** Olink panel name

**Protein** Protein name

**rsID** rs ID from dbSNP

**SNPID** SNP ID of the variant in terms of its chromosome and base pair position

**cis/trans** The SNP can be either found in the protein-encoding gene plus or minus 500kb (cis), or in other genomic regions (trans)

**A1** Coding allele

**A2** Reference allele

**Freq** Frequency of the coding allele

**Effect** Genetic effect size as standard deviations of protein concentration per coding allele

**StdErr** Standard error of the genetic effect estimate

**P-value** P-value of the meta-analysis as calculated by the software METAL

**N** Total sample size in the meta-analysis

**Direction** Cohort-specific estimated genetic effect directions, where +, -, and ? refer to positive, negative, and missing, respectively. The order of the cohorts is: For NEU: ORCADES, INTERVAL, HELIC, WHI_EA, NSPHS, LBC1936, FENLAND, SAHLSIS_Gothenburg, RS; For NEX: ORCADES, HELIC, FENLAND, STANLEY LAH1, STANLEY SWE6

**HetPVal** P-value for estimated effects heterogeneity across cohorts via a Chi-squared test

**N_Genes** Number of genes within the ±500kb window of the SNP

**Closest_Gene** The closest gene to the SNP

**Distance** Distance from the SNP to the closest gene

**Region** Genomic region of the locus

[See the Excel File]

**Supplementary Table 2. Information on the novelty of the discovered protein quantitative trait loci (pQTL) in comparison with the established pQTL in the PhenoScanner database.**

**Protein** Protein name of the meta-analysis phenotype

**ref_rsid** Proxies were requested as part of the analysis, therefore this is the input SNP

**snpid** SNP ID in terms of chromosome and base pair position

**panel** Olink panel in which the meta-analysis protein belongs to

**chr** chromosome of the variant

**pos** hg19 base pair position of the variant

**a1** Effect allele (aligned to the + strand) for the variant

**a2** Non-effect allele (aligned to the + strand) for the variant

**n** Sample size in the meta-analysis

**freq1** Frequency of the a1 allele

**beta1** Effect of the a1 allele

**se** Standard error of beta1

**p** P-value from the meta-analysis

**Direction** Cohort-specific estimated genetic effect directions, where +, -, and ? refer to positive, negative, and missing, respectively. The order of the cohorts is: For NEU: ORCADES, INTERVAL, HELIC, WHI_EA, NSPHS, LBC1936, FENLAND, SAHLSIS_Gothenburg, RS; For NEX: ORCADES, HELIC, FENLAND, STANLEY LAH1, STANLEY SWE6

**cis** The SNP can be either found in the protein encoding gene plus or minus 5e5b (cis), or in other genomic regions (trans)

**ref_hg19_coordinates** The hg19 chromosome position for the 'rsid' column

**ref_hg38_coordinates** The hg38 chromosome position for the 'rsid' column

**phenoscanner_a1** Effect allele in PhenoScanner

**phenoscanner_a2** Non-effect allele in PhenoScanner

**proxy** An indicator variable which equals 0 if the proxy SNP is the input SNP and 1 otherwise

**r2** The r2 statistic between the input SNP and the proxy SNP based on the phased haplotypes from 1000 Genomes

**dprime** The D' statistic between the input SNP and the proxy SNP based on the phased haplotypes from 1000 Genomes

**trait** Protein phenotype from the PhenoScanner database

**efo** The EFO ontology term for the protein phenotype

**study** The name of the consortium/lead author of the study

**pmid** PubMed ID of the study

**ancestry** The ancestry of the study population

**year** The year the study was published

**phenoscanner_beta** Effect in PhenoScanner for the association between the protein phenotype and the SNP expressed per additional copy of the effect allele

**phenoscanner_se** Standard error of phenoscanner_beta

**phenoscanner_p** P-value from PhenoScanner

**direction** PhenoScanner direction of association with respect to the effect allele

**phenoscanner_n** Number of individuals in the study from the PhenoScanner database

**same_target** An indicator variable that shows whether the PhenoScanner_pQTL_trait corresponds to the protein in which the meta-analysis pQTL was reported (yes), or otherwise (no)

[See the Excel File]

**Supplementary Table 3. Established complex traits associations of the mapped cis-pQTL lead variants in the PhenoScanner database.**

**Protein** Protein name of the meta-analysis phenotype

**rsid** rsid of the variant

**snpid** SNP ID in terms of chromosome and base pair position

**hg19_coordinates** The hg19 chromosome position for the 'rsid' column

**hg38_coordinates** The hg38 chromosome position for the 'rsid' column

**a1** Effect allele for the variant

**a2** Non-effect allele for the variant

**trait** Protein phenotype from the PhenoScanner database

**efo** The EFO ontology term for the protein phenotype

**study** The name of the consortium/lead author of the study

**pmid** PubMed ID of the study

**ancestry** The ancestry of the study population

**year** The year the study was published

**beta** Effect in PhenoScanner for the association between the protein phenotype and the SNP expressed per additional copy of the effect allele

**se** Standard error of phenoscanner_beta

**p** P-value from PhenoScanner

**direction** PhenoScanner direction of association with respect to the effect allele

**n** Number of individuals in the study from the PhenoScanner database

**n_cases** Number of cases in the study from the PhenoScanner database

**n_controls** Number of controls in the study from the PhenoScanner database

**unit** Phenotype unit

**FDR** False discovery rate adjusted from the p-value

[See the Excel File]

**Supplementary Table 4. List of the LD-pruned (r2 < .001) genome-wide significant variants of each cis-pQTL.** The column names are described as in Supplementary Table 1.

[See the Excel File]

**Supplementary Table 5. Mendelian Randomisation (MR) causal inference results between the analyzed proteins and 152 neuro-related phenotypes.** **Name** Protein name

**Gene** Gene name

**Protein_full_name** Full name of the protein

**Trait** IDs of traits

**Description** Descriptions of the traits

**Name in figure** Trait name used in figures

**SNP num** Number of independent SNPs used as instruments

**Effect** Size of the causal effect estimate

**SE** Standard error of the effect

**P-value** P-value testing the estimated effect

**SMR p-value** P-value based on the SMR method

**HEIDI p-value** P-value based on the HEIDI method for colocalization

[See the Excel File]

**Supplementary Table 6. Mendelian Randomisation (MR) causal inference results between the analyzed proteins and UK Biobank disease phenotypes.** **Name** Protein name

**Gene** Gene name

**Protein_full_name** Full name of the protein

**Trait** IDs of traits

**Description** Descriptions of the traits

**Name in figure** Trait name used in figures

**SNP num** Number of independent SNPs used as instruments

**Effect** Size of the causal effect estimate

**SE** Standard error of the effect

**P-value** P-value testing the estimated effect

**SMR p-value** P-value based on the SMR method

**HEIDI p-value** P-value based on the HEIDI method for colocalization

[See the Excel File]

**Supplementary Table 7. Results of the eQTL-pQTL genetic association colocalization analysis using coloc.** Two sources of cis-eQTL information were used, GTEx tissues and eQTLGen. For the cis-pQTL, we tested the colocalization between the circulating protein levels and the expressions of their encoding genes. For the trans-pQTL, we tested the colocalization between the circulating protein levels and the expression of each gene within 2Mb of the pQTL lead variant. The eQTL association summary-level data for different tissues were obtained from the GTEx portal. The table shows all the results with colocalization posterior probability (PP4) $> 0.8$.

[See the Excel File]

**Supplementary Table 8. Gene set enrichment analysis results from the FUMA GENE2FUNC module.**

**Category** Category from the MsigDB database

**GeneSet** Name of gene set as provided by MsigDB

**N_genes** Number of genes in a gene set

**N_overlap** Number of input genes (most proximal genes to meta-analysis trans-pQTLs) overlapping with the gene set

**P-value** P-value calculated on FUMA

**Adj_P-value** FDR adjusted P-value

**Genes** Overlapping genes that are most proximal to the trans-pQTL found in the meta-analysis, and submitted to FUMA

**link** Link to the MsigDB database of the GeneSet used for the enrichment analysis

[See the Excel File]

**Supplementary Table 9. DrugBank known (Re-purposing and Validated) drug targets for protein-trait IVW MR discoveries (FDR $< 0.05$).** *: the Category column, 0 is the druggable target that has drugs with unclear clinical function, 1 is the validated known target where the established drug has the pharmacological effect that matches the MR result, and 2 is the re-purposing target that has established drugs but for different diseases.

[See the Excel File]

**Supplementary Table 10. New potential drug targets suggested by protein-trait IVW MR discoveries (FDR $< 0.05$).**

[See the Excel File]

**Supplementary Table 11. Information on the cohorts participating in the meta-analysis and replication.**

[See the Excel File]

**Supplementary Table 12. Summary data information of the ORCADES cohort.**

[See the Excel File]

**Supplementary Table 13. Summary data information of the INTERVAL cohort.**

[See the Excel File]

**Supplementary Table 14. Summary data information of the NSPHS cohort.**

[See the Excel File]

**Supplementary Table 15. Summary data information of the LBC1936 cohort.**

[See the Excel File]

**Supplementary Table 16. Summary data information of the Fenland cohort.**

[See the Excel File]

**Supplementary Table 17. Summary data information of the SAHLSIS Gothenburg cohort.**

[See the Excel File]

**Supplementary Table 18. Summary data information of the Rotterdam Study cohort.**

[See the Excel File]

**Supplementary Table 19. Summary data information of the STANLEY LAH1 cohort.**

[See the Excel File]

**Supplementary Table 20. Summary data information of the STANLEY SWE6 cohort.**

[See the Excel File]

**Supplementary Table 21. Summary data information of the HELIC Manolis cohort.**

[See the Excel File]

**Supplementary Table 22. Summary data information of the HELIC Pomak cohort.**

[See the Excel File]

**Supplementary Table 23. Summary data information of the WHI European American cohort.**

[See the Excel File]

**Supplementary Table 24. Summary data information of the WHI Hispanic American cohort.**

[See the Excel File]

**Supplementary Table 25. Summary data information of the WHI African American cohort.**

[See the Excel File]

### 3 Supplementary Figures

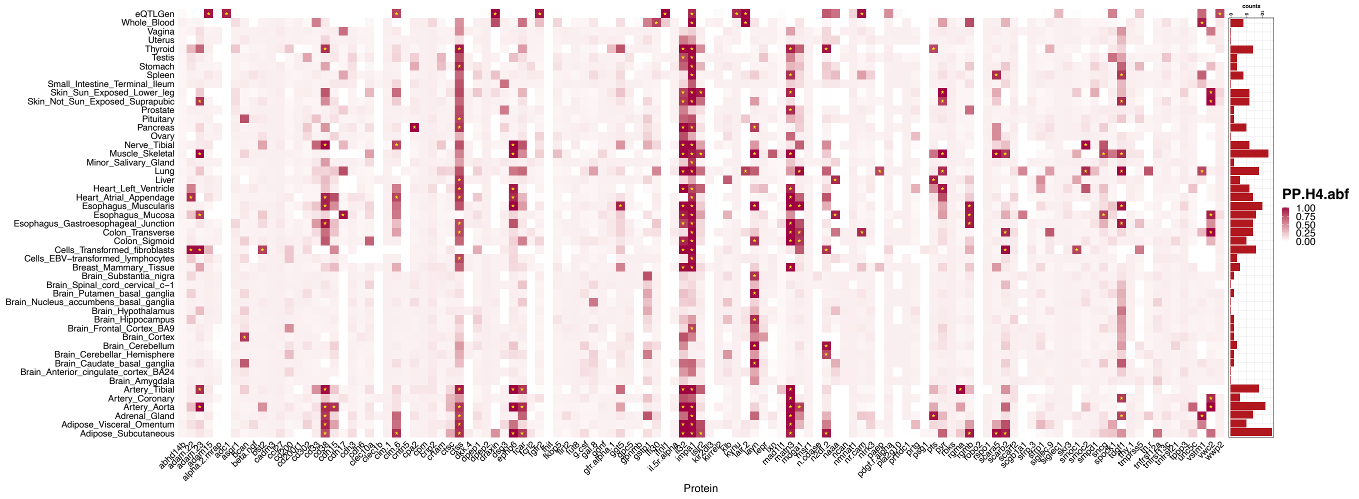


**Supplementary Figure 1. Posterior probabilities of the colocalization between the detected cis-pQTL and the cis-eQTL of the corresponding coding gene in the GTEx data.** The probability was calculated using `coloc` testing the H4 colocalization hypothesis.

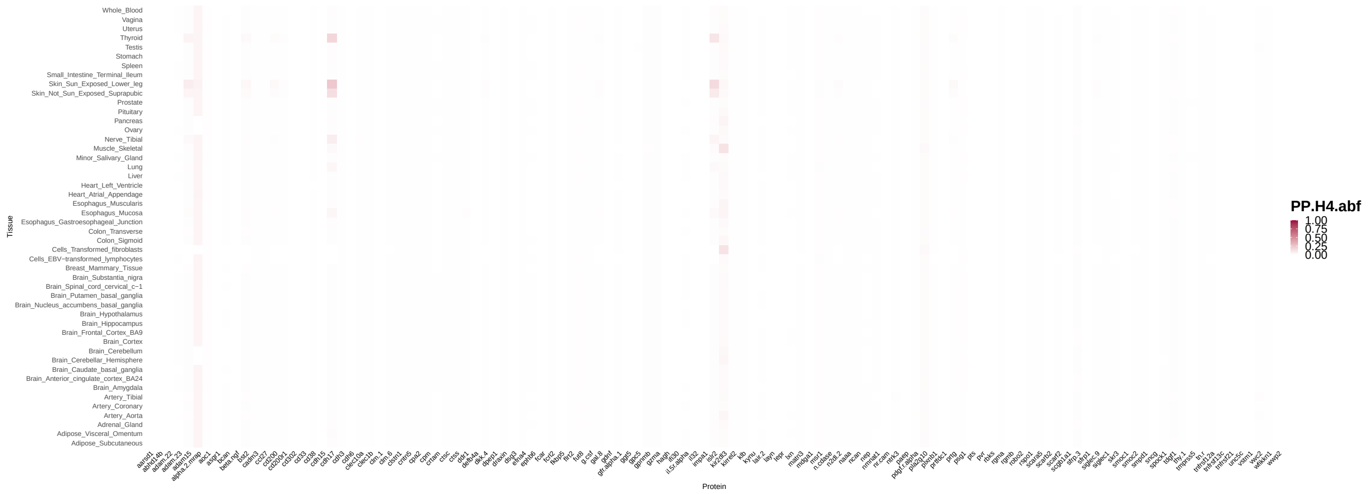


**Supplementary Figure 2. Posterior probabilities of the colocalization between the detected trans-pQTL and the cis-eQTL of the nearest coding gene in the GTEx data.** The probability was calculated using `coloc` testing the H4 colocalization hypothesis.

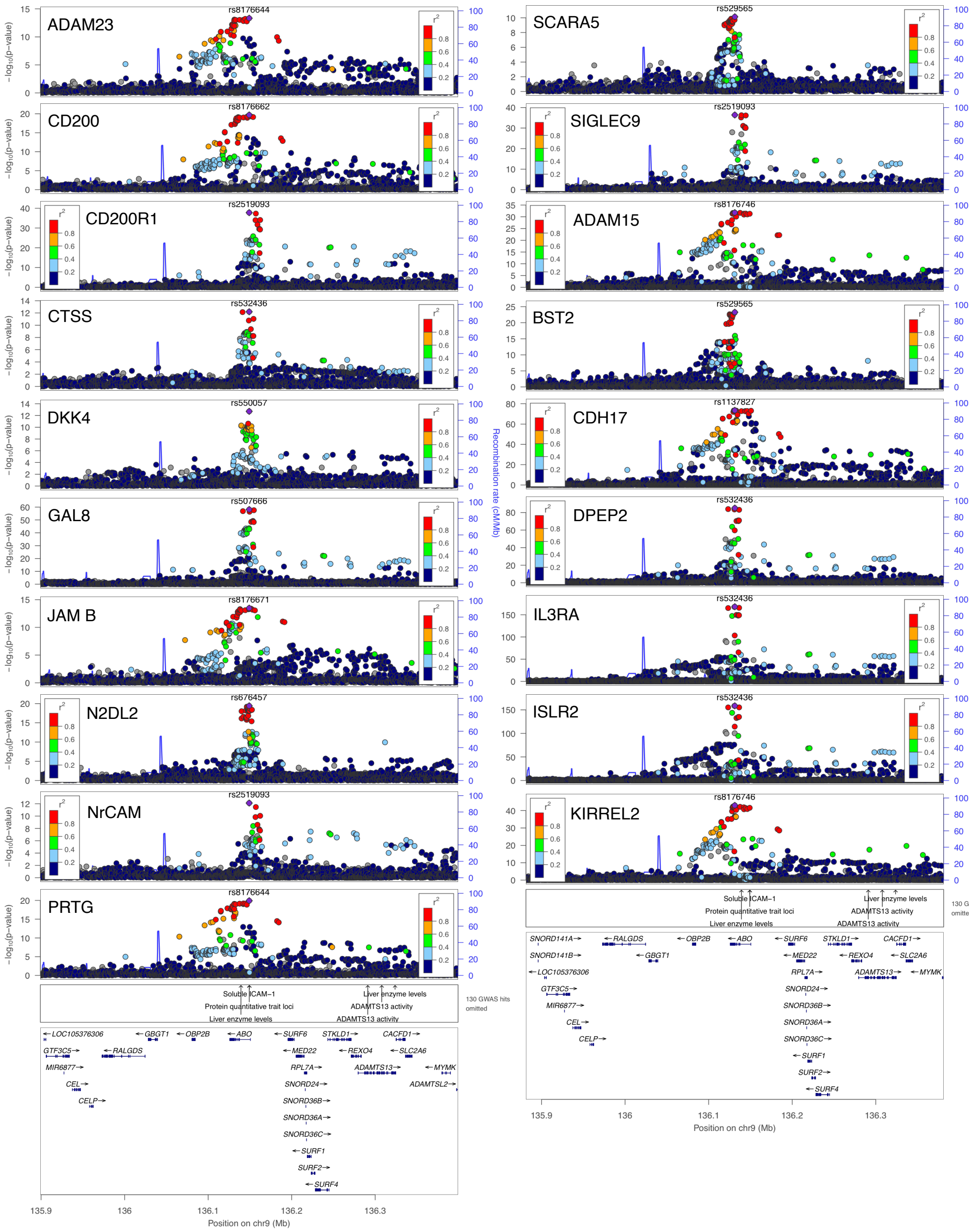


**Supplementary Figure 3: The *ABO* locus as significant trans-pQTL for 19 proteins.** The $r^2$ values are the squared linkage disequilibrium correlation coefficients between the plotted variants and the leading variant (purple dot). The bottom panel marks the established associations at this locus according to the GWAS catalog ($p < 5 \times 10^{-8}$) and annotates the positions of the nearby genes.

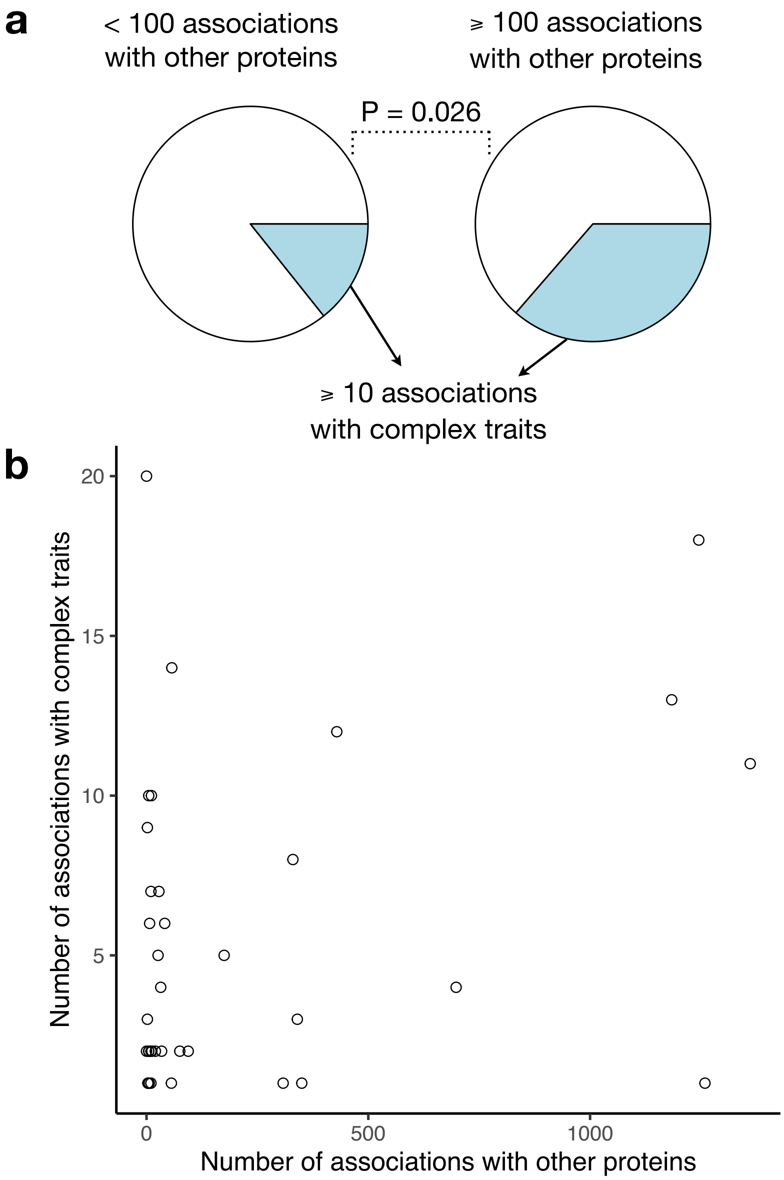


**Supplementary Figure 4: Number of trans-pQTL associations and associated complex traits in PhenoScanner for 39 cis-pQTL.** (a) Significant enrichment of the cis-pQTL associated with more than 10 complex traits when the cis-pQTL has more than 100 trans-pQTL association records. (b) The scatter plot of the number of trans-pQTL associations v.s. the number of associated complex traits.

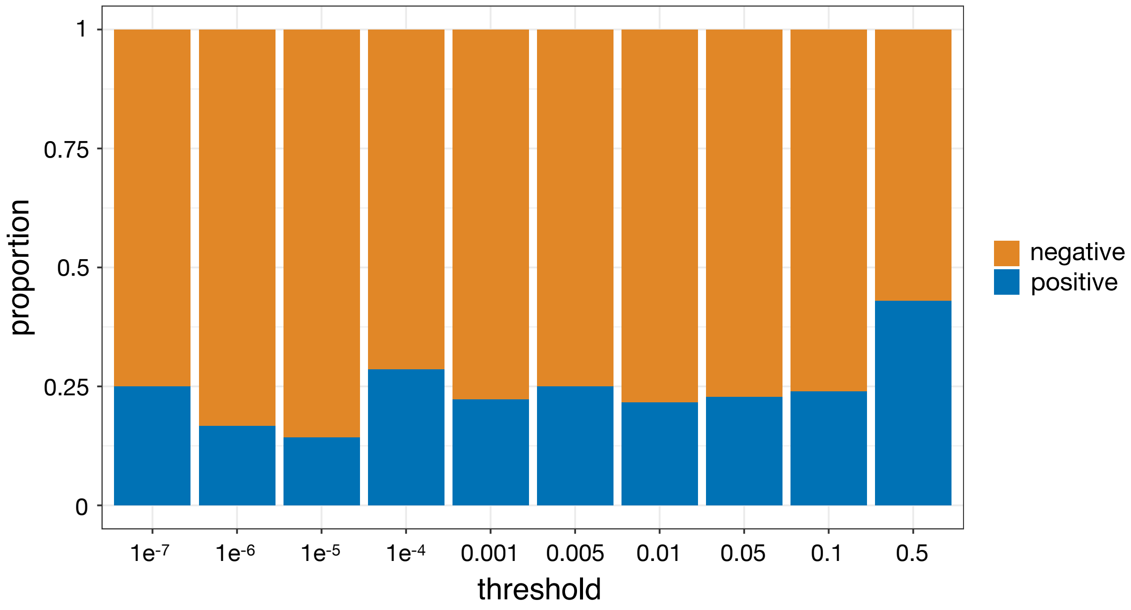


**Supplementary Figure 5. Proportion of negative v.s. positive estimated causal effects based on Mendelian randomization given different significance thresholds.**

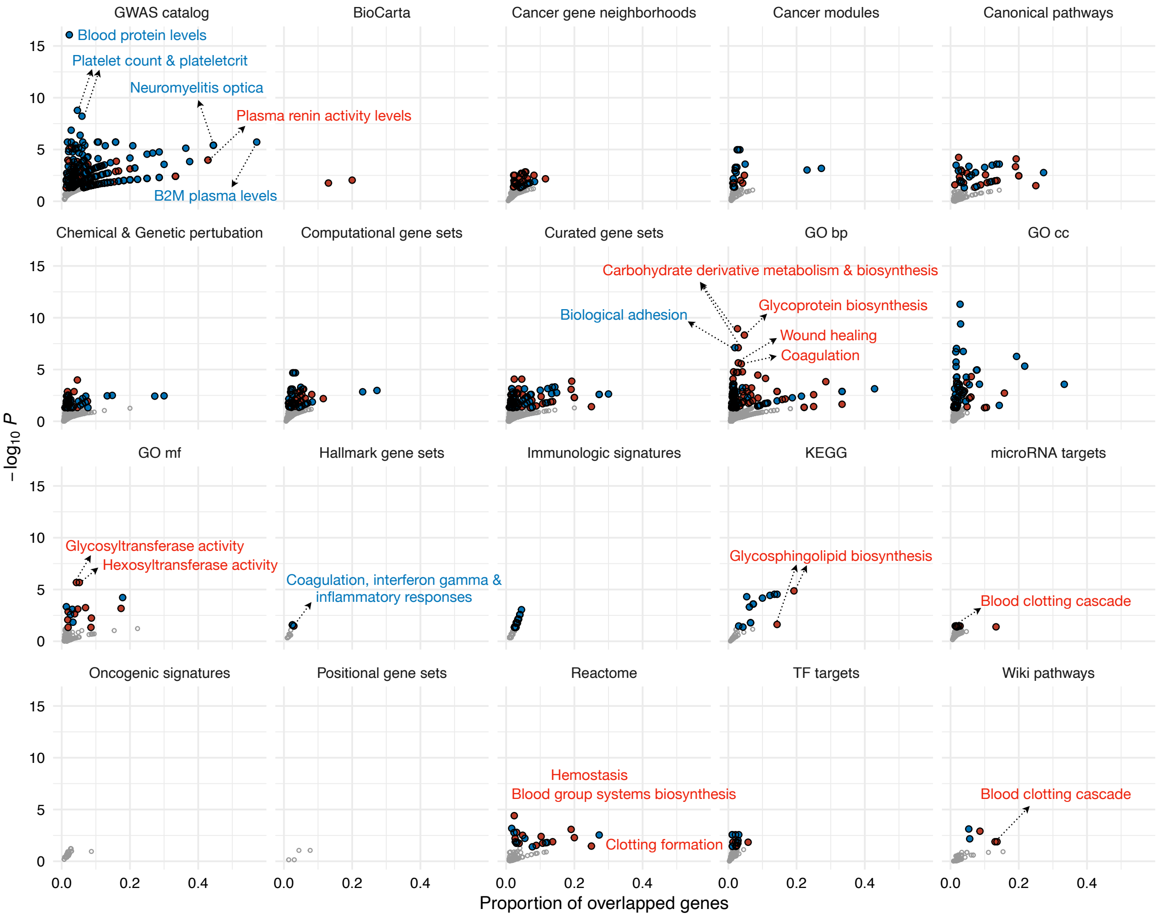


**Supplementary Figure 6: Gene set enrichment analysis reveals that the discovered trans-pQTL are involved in blood-related pathways.** Significant enrichment results are highlighted in blue and red dots, corresponding to the gene sets involving and not involving the HLA genes, respectively. Gene set enrichment analyses were conducted using FUMA's GENE2FUNC module. B2M: Beta-2 microglubulin.

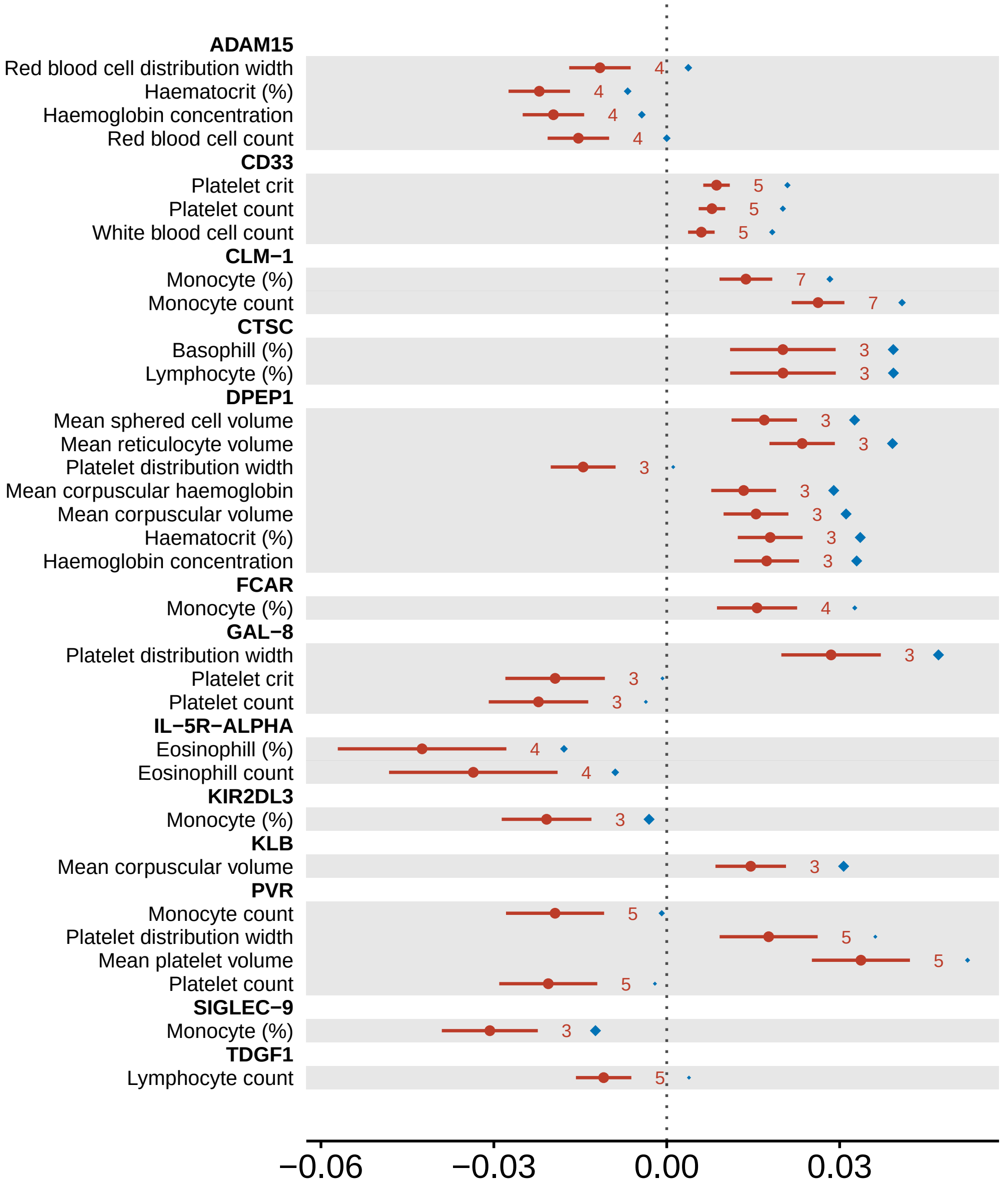


**Supplementary Figure 7: Causality between the proteins and UK Biobank complete blood count (CBC) phenotypes inferred by Mendelian randomisation (MR) analyses.** The forest plot shows the significant MR results (false discovery rate $< 0.05$) based on LD-pruned ($r^2 < 0.001$) instrumental variants within each cis-pQTL. Inverse-variance weighted (IVW) estimates are provided as the solid round dots, and the whiskers indicate standard errors. The numbers of instrumental variants in the cis-pQTL are given to the right of the whiskers. For each cis-pQTL with at least three instrumental variants, a rank-based correlation coefficient was calculated between the genetic effects on the protein and those on the trait. As a colocalisation measure, the squared correlations are given as the diamonds, where the largest diamonds correspond to a value of 1. Results with colocalisation posterior probability from the coloc tool greater than 0.8 are marked with a yellow star.

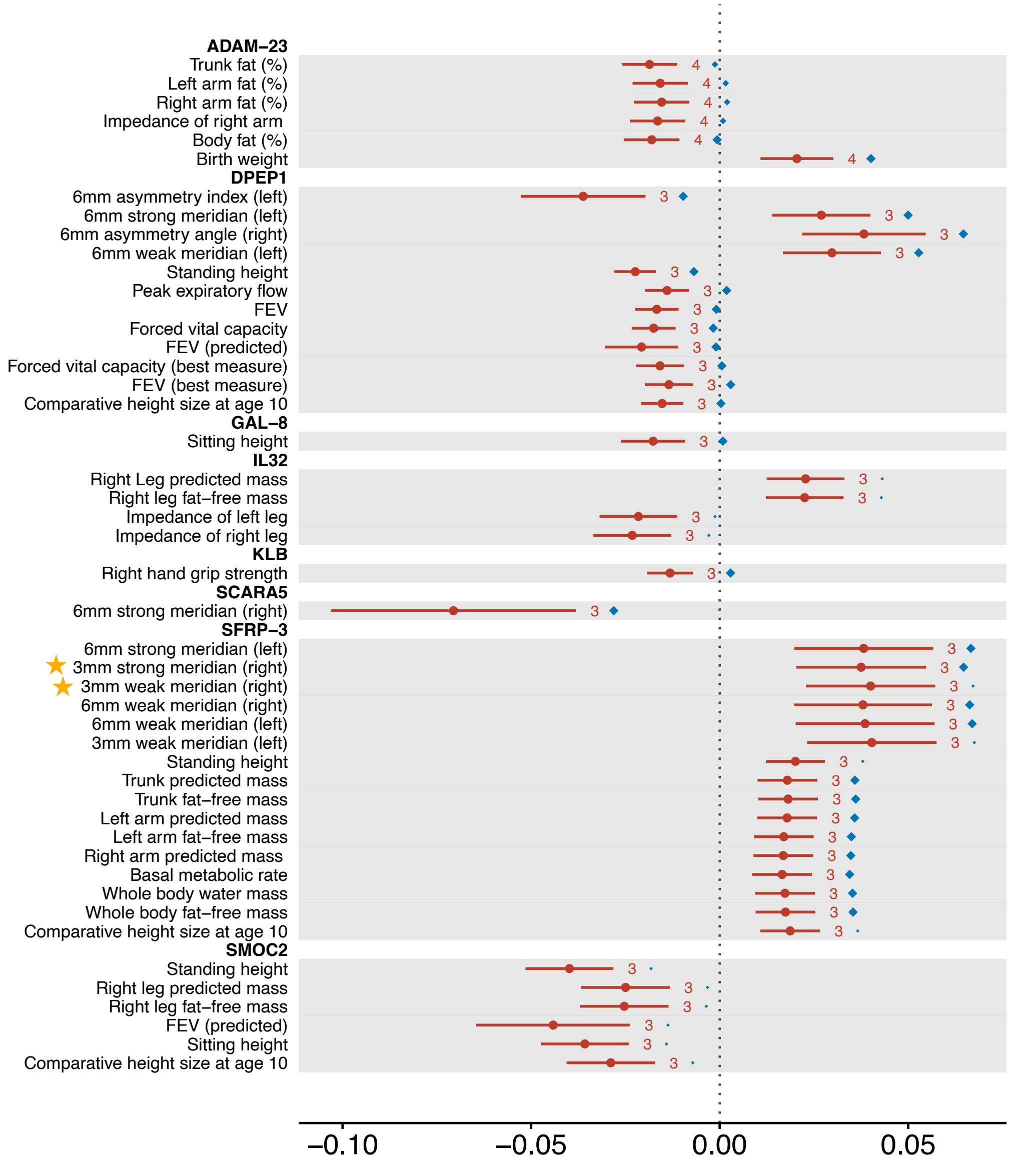


**Supplementary Figure 8: Causality between the proteins and UK Biobank anthropometrical phenotypes inferred by Mendelian randomisation (MR) analyses.** The forest plot shows the significant MR results (false discovery rate < 0.05) based on LD-pruned ($r^2 < 0.001$) instrumental variants within each cis-pQTL. Inverse-variance weighted (IVW) estimates are provided as the solid round dots, and the whiskers indicate standard errors. The numbers of instrumental variants in the cis-pQTL are given to the right of the whiskers. For each cis-pQTL with at least three instrumental variants, a rank-based correlation coefficient was calculated between the genetic effects on the protein and those on the trait. As a colocalisation measure, the squared correlations are given as the diamonds, where the largest diamonds correspond to a value of 1. Results with colocalisation posterior probability from the coloc tool greater than 0.8 are marked with a yellow star.

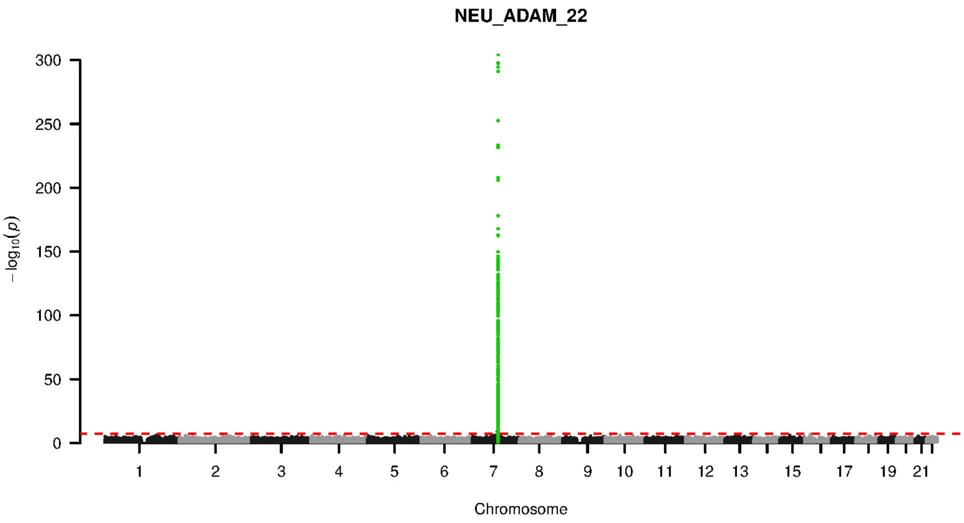


**Supplementary Figure 9-1. Manhattan plot and pQTL associations of the genome-wide meta-analysis for protein ADAM 22.** The horizontal dashed line corresponds to the genome-wide significance threshold of $p = 5 \times 10^{-8}$. The variants in the cis-regulatory region (±500kb of the coding gene region) of the protein-coding gene are highlighted in green.

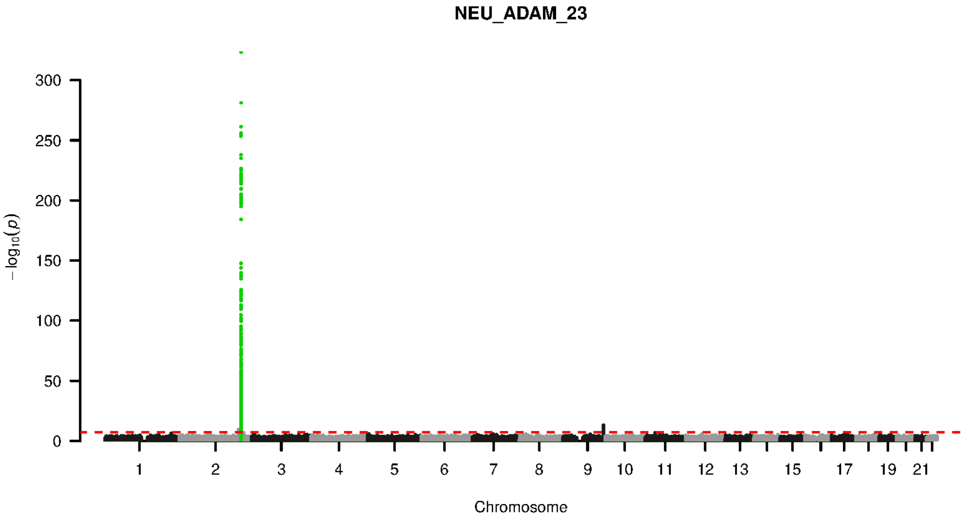


**Supplementary Figure 9-2. Manhattan plot and pQTL associations of the genome-wide meta-analysis for protein ADAM 23.** The horizontal dashed line corresponds to the genome-wide significance threshold of $p = 5 \times 10^{-8}$. The variants in the cis-regulatory region (±500kb of the coding gene region) of the protein-coding gene are highlighted in green.

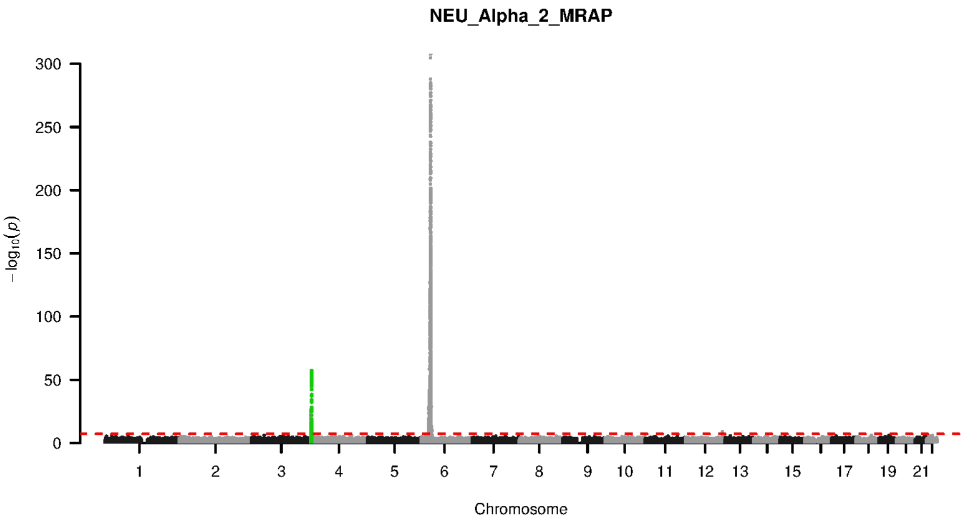


**Supplementary Figure 9-3. Manhattan plot and pQTL associations of the genome-wide meta-analysis for protein ALPHA 2 MRAP.** The horizontal dashed line corresponds to the genome-wide significance threshold of $p = 5 \times 10^{-8}$. The variants in the cis-regulatory region ($\pm$500kb of the coding gene region) of the protein-coding gene are highlighted in green.

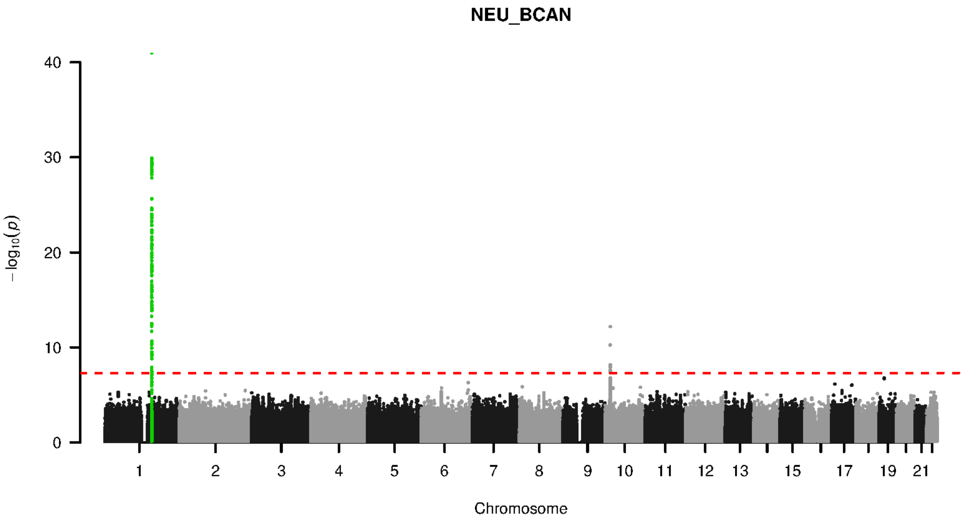


**Supplementary Figure 9-4. Manhattan plot and pQTL associations of the genome-wide meta-analysis for protein BCAN.** The horizontal dashed line corresponds to the genome-wide significance threshold of $p = 5 \times 10^{-8}$. The variants in the cis-regulatory region ($\pm$500kb of the coding gene region) of the protein-coding gene are highlighted in green.

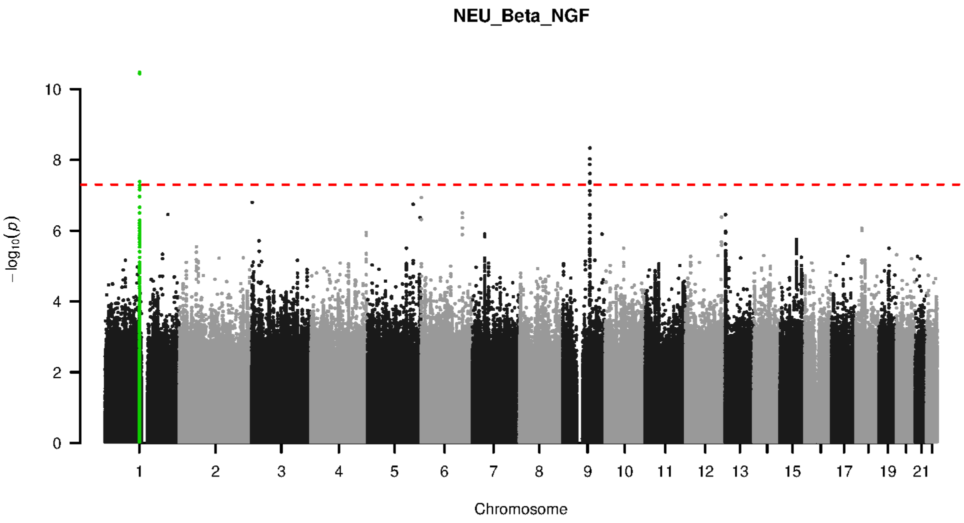


**Supplementary Figure 9-5. Manhattan plot and pQTL associations of the genome-wide meta-analysis for protein BETA NGF.** The horizontal dashed line corresponds to the genome-wide significance threshold of $p = 5 \times 10^{-8}$. The variants in the cis-regulatory region (±500kb of the coding gene region) of the protein-coding gene are highlighted in green.

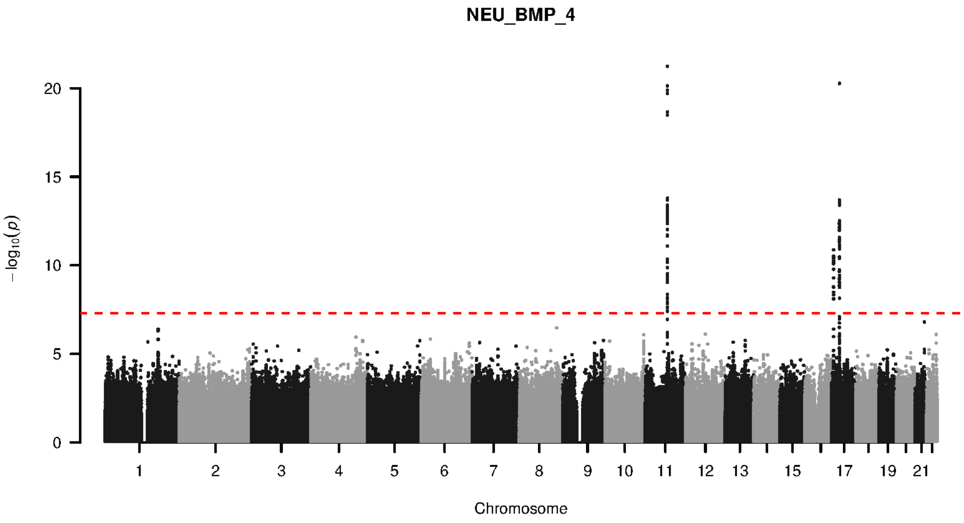


**Supplementary Figure 9-6. Manhattan plot and pQTL associations of the genome-wide meta-analysis for protein BMP 4.** The horizontal dashed line corresponds to the genome-wide significance threshold of $p = 5 \times 10^{-8}$. The variants in the cis-regulatory region (±500kb of the coding gene region) of the protein-coding gene are highlighted in green.

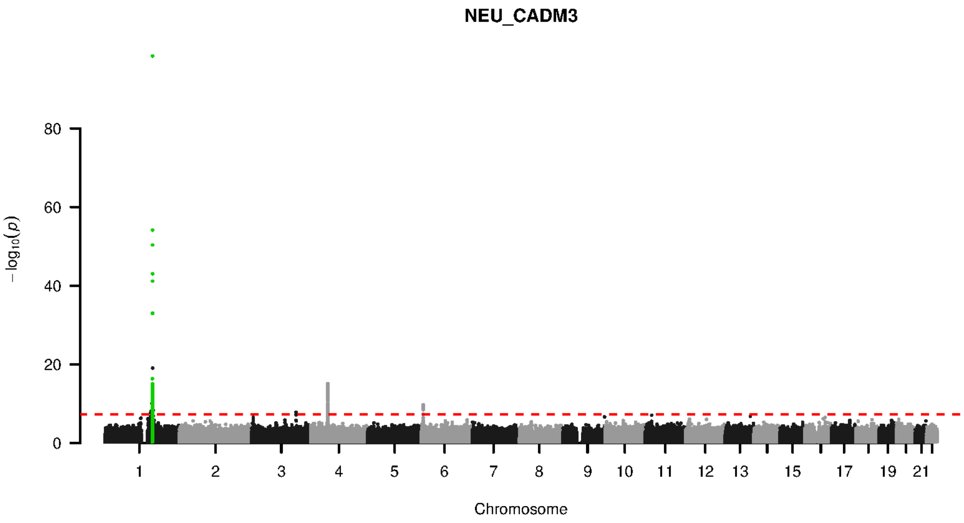


**Supplementary Figure 9-7. Manhattan plot and pQTL associations of the genome-wide meta-analysis for protein CADM3.** The horizontal dashed line corresponds to the genome-wide significance threshold of $p = 5 \times 10^{-8}$. The variants in the cis-regulatory region ($\pm$500kb of the coding gene region) of the protein-coding gene are highlighted in green.

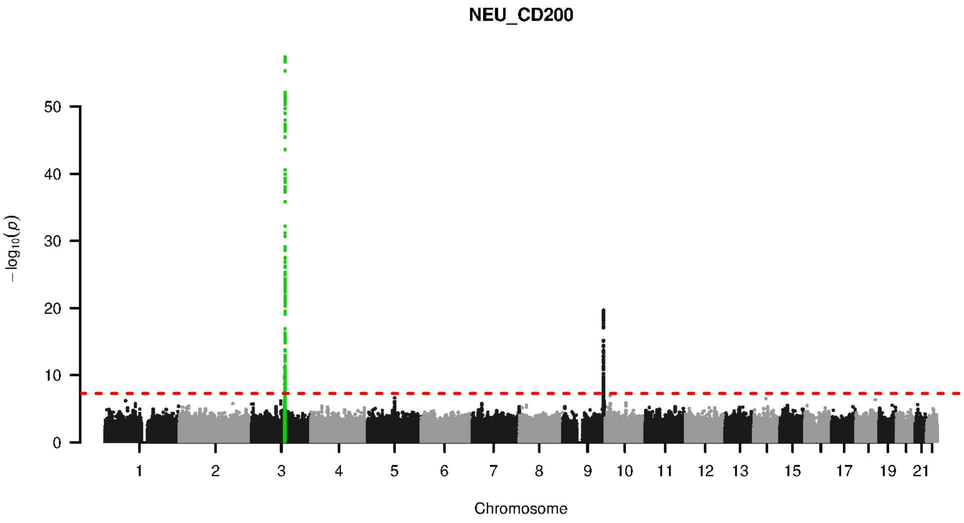


**Supplementary Figure 9-8. Manhattan plot and pQTL associations of the genome-wide meta-analysis for protein CD200.** The horizontal dashed line corresponds to the genome-wide significance threshold of $p = 5 \times 10^{-8}$. The variants in the cis-regulatory region ($\pm$500kb of the coding gene region) of the protein-coding gene are highlighted in green.

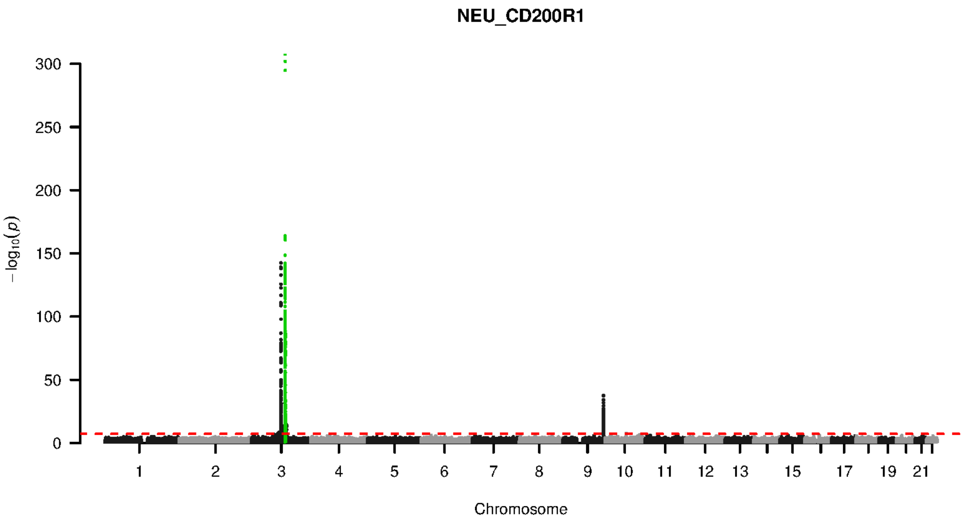


**Supplementary Figure 9-9. Manhattan plot and pQTL associations of the genome-wide meta-analysis for protein CD200R1.** The horizontal dashed line corresponds to the genome-wide significance threshold of $p = 5 \times 10^{-8}$. The variants in the cis-regulatory region (±500kb of the coding gene region) of the protein-coding gene are highlighted in green.

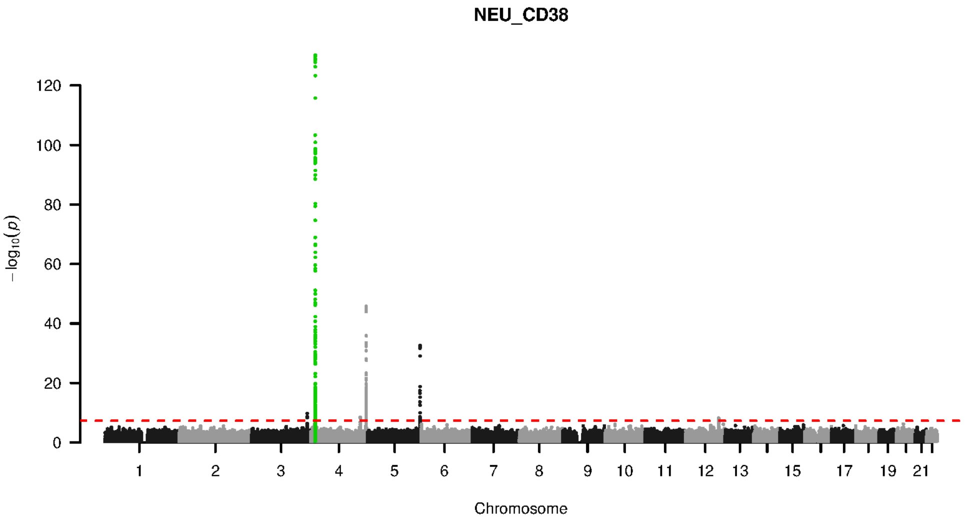


**Supplementary Figure 9-10. Manhattan plot and pQTL associations of the genome-wide meta-analysis for protein CD38.** The horizontal dashed line corresponds to the genome-wide significance threshold of $p = 5 \times 10^{-8}$. The variants in the cis-regulatory region (±500kb of the coding gene region) of the protein-coding gene are highlighted in green.

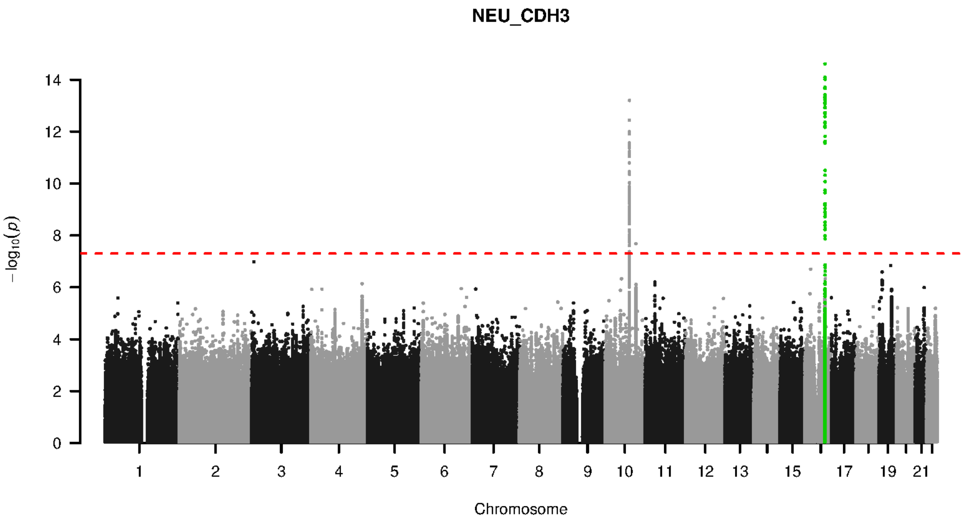


**Supplementary Figure 9-11. Manhattan plot and pQTL associations of the genome-wide meta-analysis for protein CDH3.** The horizontal dashed line corresponds to the genome-wide significance threshold of $p = 5 \times 10^{-8}$. The variants in the cis-regulatory region (±500kb of the coding gene region) of the protein-coding gene are highlighted in green.

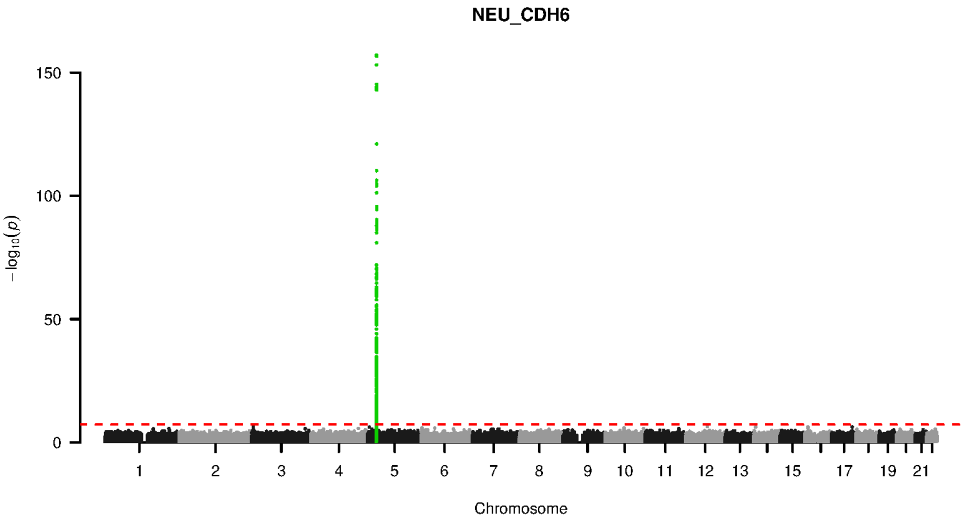


**Supplementary Figure 9-12. Manhattan plot and pQTL associations of the genome-wide meta-analysis for protein CDH6.** The horizontal dashed line corresponds to the genome-wide significance threshold of $p = 5 \times 10^{-8}$. The variants in the cis-regulatory region (±500kb of the coding gene region) of the protein-coding gene are highlighted in green.

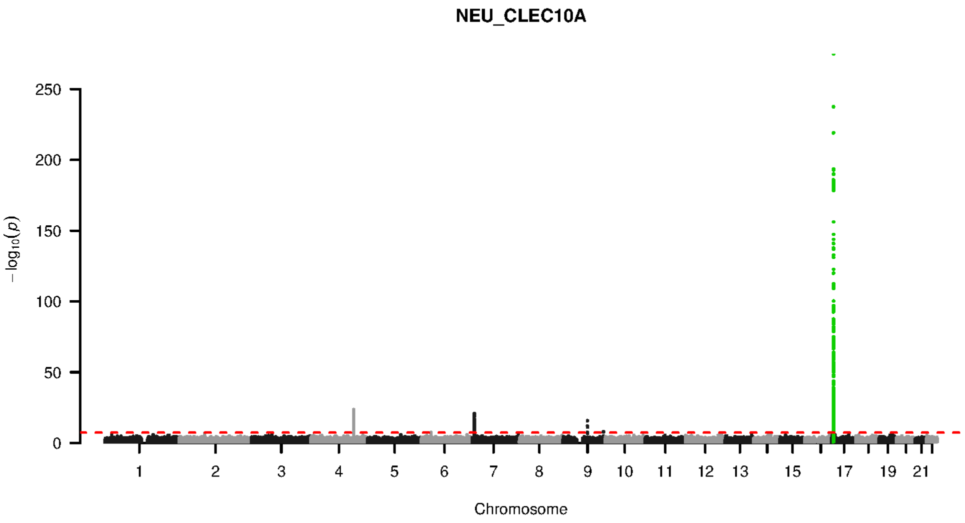


**Supplementary Figure 9-13. Manhattan plot and pQTL associations of the genome-wide meta-analysis for protein CLEC10A.** The horizontal dashed line corresponds to the genome-wide significance threshold of $p = 5 \times 10^{-8}$. The variants in the cis-regulatory region (±500kb of the coding gene region) of the protein-coding gene are highlighted in green.

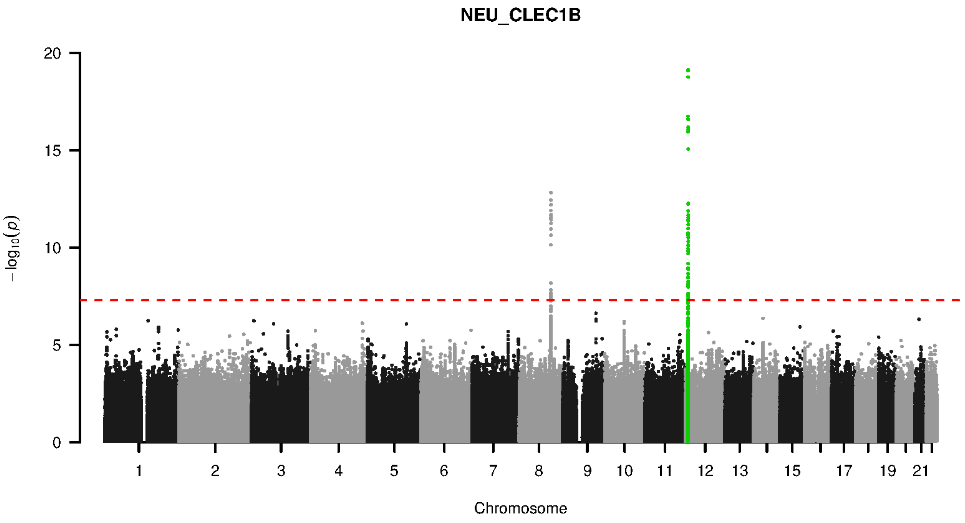


**Supplementary Figure 9-14. Manhattan plot and pQTL associations of the genome-wide meta-analysis for protein CLEC1B.** The horizontal dashed line corresponds to the genome-wide significance threshold of $p = 5 \times 10^{-8}$. The variants in the cis-regulatory region (±500kb of the coding gene region) of the protein-coding gene are highlighted in green.

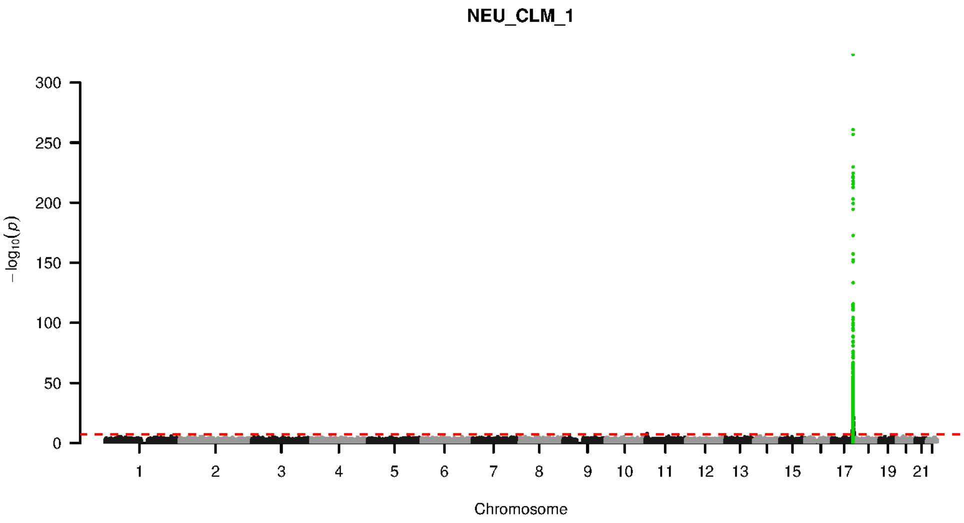


**Supplementary Figure 9-15. Manhattan plot and pQTL associations of the genome-wide meta-analysis for protein CLM 1.** The horizontal dashed line corresponds to the genome-wide significance threshold of $p = 5 \times 10^{-8}$. The variants in the cis-regulatory region (±500kb of the coding gene region) of the protein-coding gene are highlighted in green.

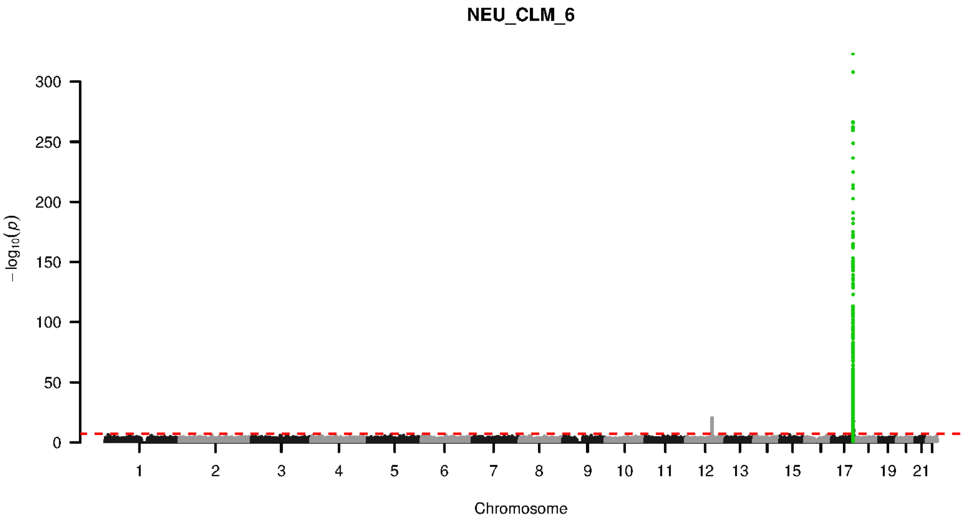


**Supplementary Figure 9-16. Manhattan plot and pQTL associations of the genome-wide meta-analysis for protein CLM 6.** The horizontal dashed line corresponds to the genome-wide significance threshold of $p = 5 \times 10^{-8}$. The variants in the cis-regulatory region (±500kb of the coding gene region) of the protein-coding gene are highlighted in green.

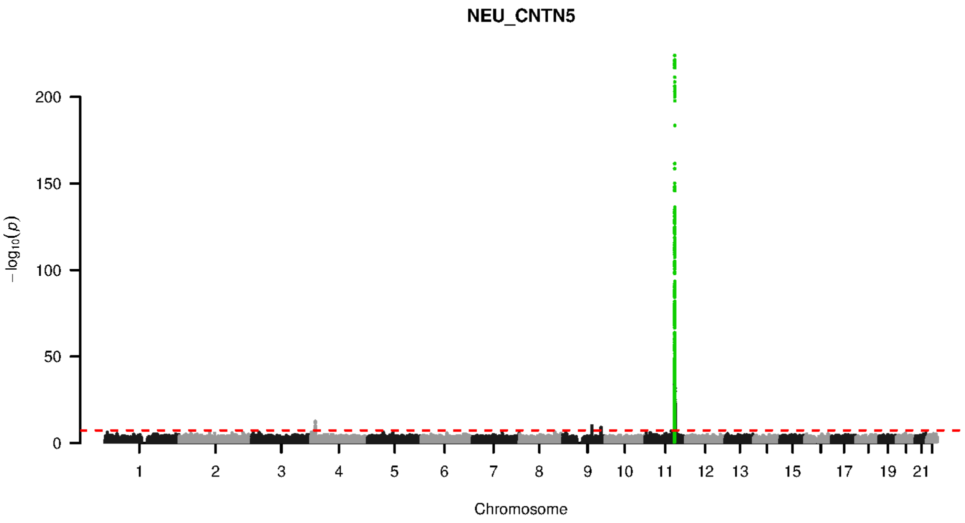


**Supplementary Figure 9-17. Manhattan plot and pQTL associations of the genome-wide meta-analysis for protein CNTN5.** The horizontal dashed line corresponds to the genome-wide significance threshold of $p = 5 \times 10^{-8}$. The variants in the cis-regulatory region (±500kb of the coding gene region) of the protein-coding gene are highlighted in green.

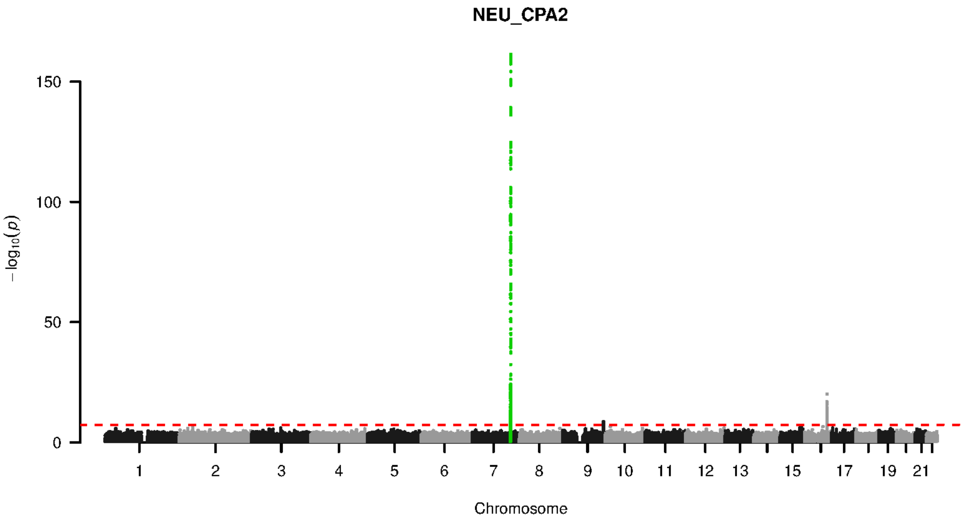


**Supplementary Figure 9-18. Manhattan plot and pQTL associations of the genome-wide meta-analysis for protein CPA2.** The horizontal dashed line corresponds to the genome-wide significance threshold of $p = 5 \times 10^{-8}$. The variants in the cis-regulatory region (±500kb of the coding gene region) of the protein-coding gene are highlighted in green.

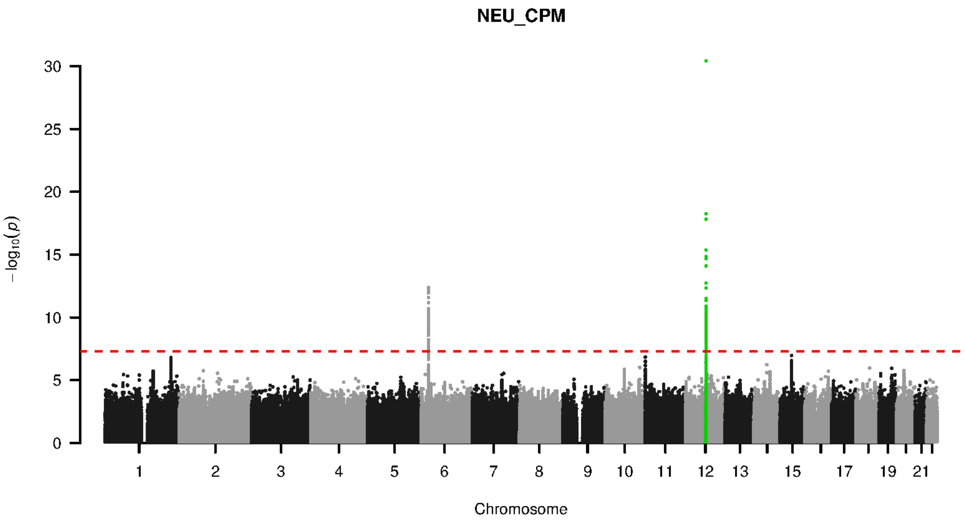


**Supplementary Figure 9-19. Manhattan plot and pQTL associations of the genome-wide meta-analysis for protein CPM.** The horizontal dashed line corresponds to the genome-wide significance threshold of $p = 5 \times 10^{-8}$. The variants in the cis-regulatory region ($\pm$500kb of the coding gene region) of the protein-coding gene are highlighted in green.

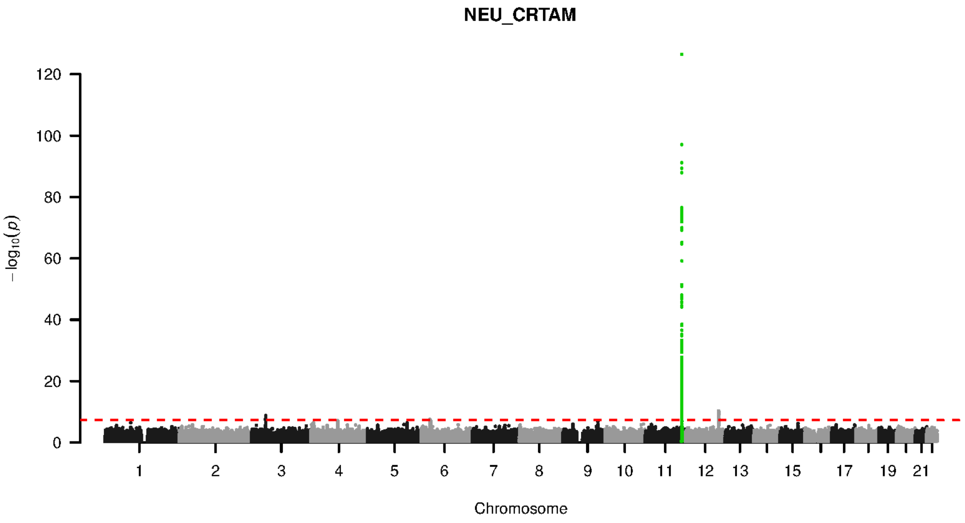


**Supplementary Figure 9-20. Manhattan plot and pQTL associations of the genome-wide meta-analysis for protein CRTAM.** The horizontal dashed line corresponds to the genome-wide significance threshold of $p = 5 \times 10^{-8}$. The variants in the cis-regulatory region ($\pm$500kb of the coding gene region) of the protein-coding gene are highlighted in green.

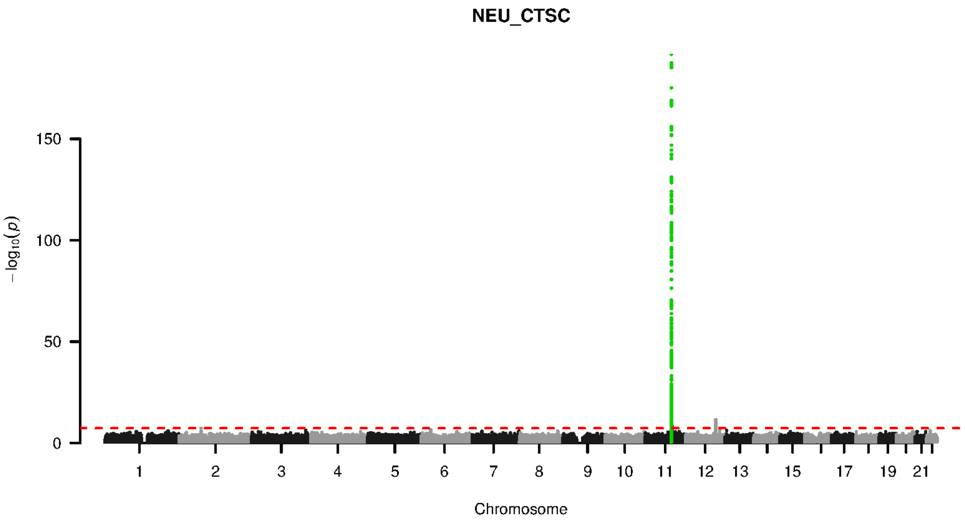


**Supplementary Figure 9-21. Manhattan plot and pQTL associations of the genome-wide meta-analysis for protein CTSC.** The horizontal dashed line corresponds to the genome-wide significance threshold of $p = 5 \times 10^{-8}$. The variants in the cis-regulatory region (±500kb of the coding gene region) of the protein-coding gene are highlighted in green.

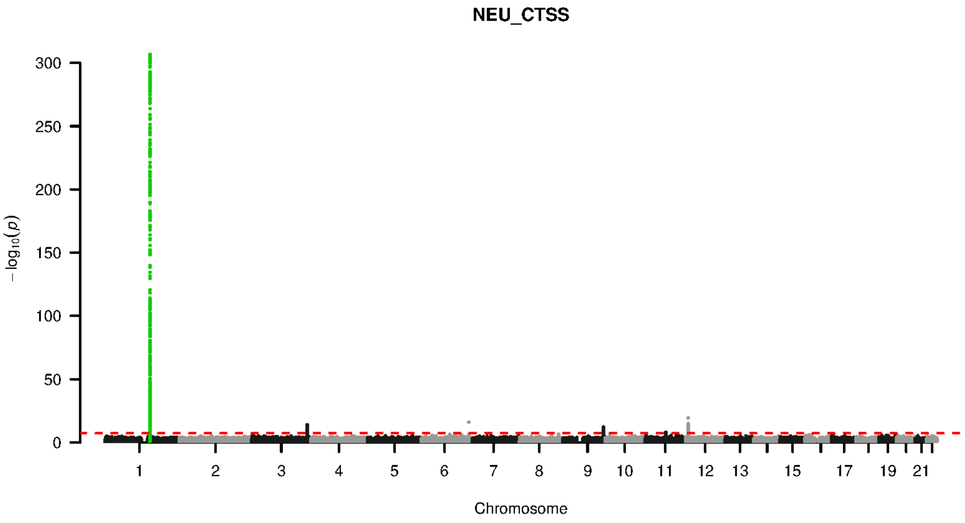


**Supplementary Figure 9-22. Manhattan plot and pQTL associations of the genome-wide meta-analysis for protein CTSS.** The horizontal dashed line corresponds to the genome-wide significance threshold of $p = 5 \times 10^{-8}$. The variants in the cis-regulatory region (±500kb of the coding gene region) of the protein-coding gene are highlighted in green.

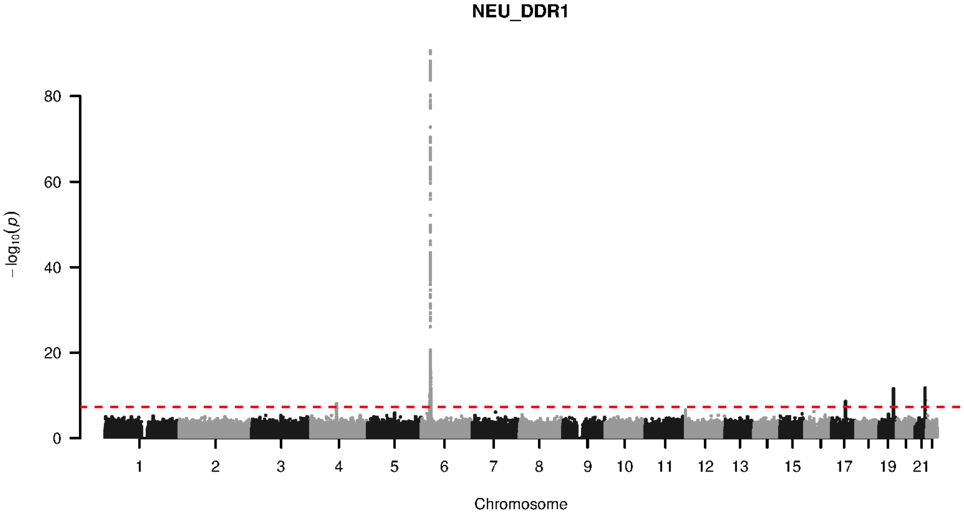


**Supplementary Figure 9-23. Manhattan plot and pQTL associations of the genome-wide meta-analysis for protein DDR1.** The horizontal dashed line corresponds to the genome-wide significance threshold of $p = 5 \times 10^{-8}$. The variants in the cis-regulatory region (±500kb of the coding gene region) of the protein-coding gene are highlighted in green.

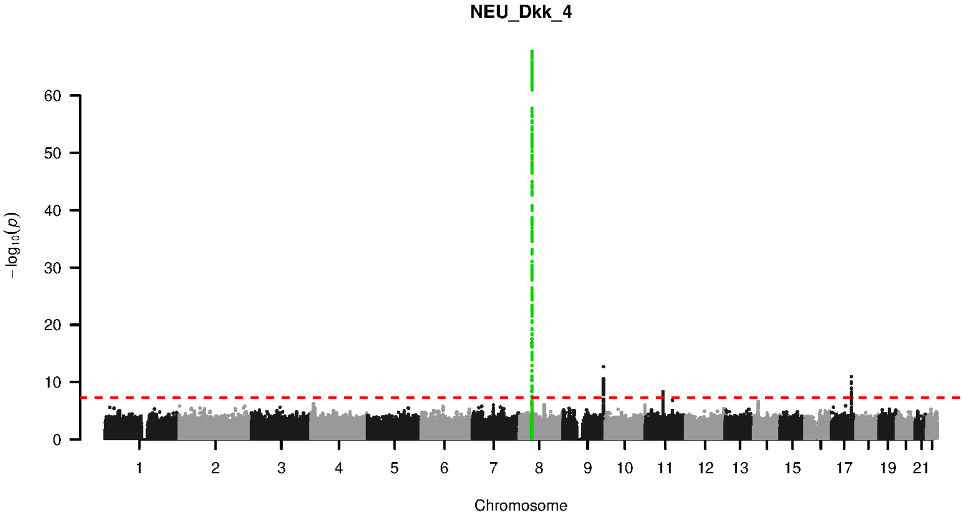


**Supplementary Figure 9-24. Manhattan plot and pQTL associations of the genome-wide meta-analysis for protein DKK 4.** The horizontal dashed line corresponds to the genome-wide significance threshold of $p = 5 \times 10^{-8}$. The variants in the cis-regulatory region (±500kb of the coding gene region) of the protein-coding gene are highlighted in green.

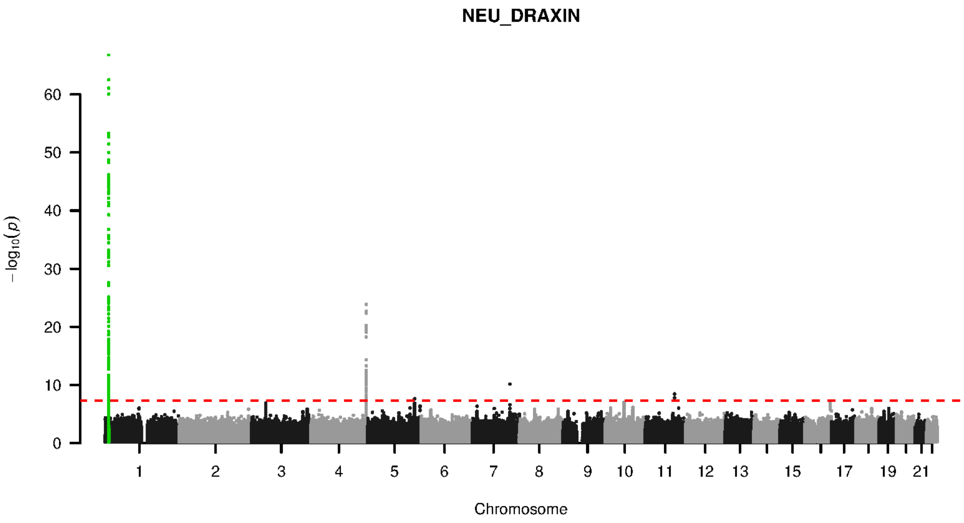


**Supplementary Figure 9-25. Manhattan plot and pQTL associations of the genome-wide meta-analysis for protein DRAXIN.** The horizontal dashed line corresponds to the genome-wide significance threshold of $p = 5 \times 10^{-8}$. The variants in the cis-regulatory region (±500kb of the coding gene region) of the protein-coding gene are highlighted in green.

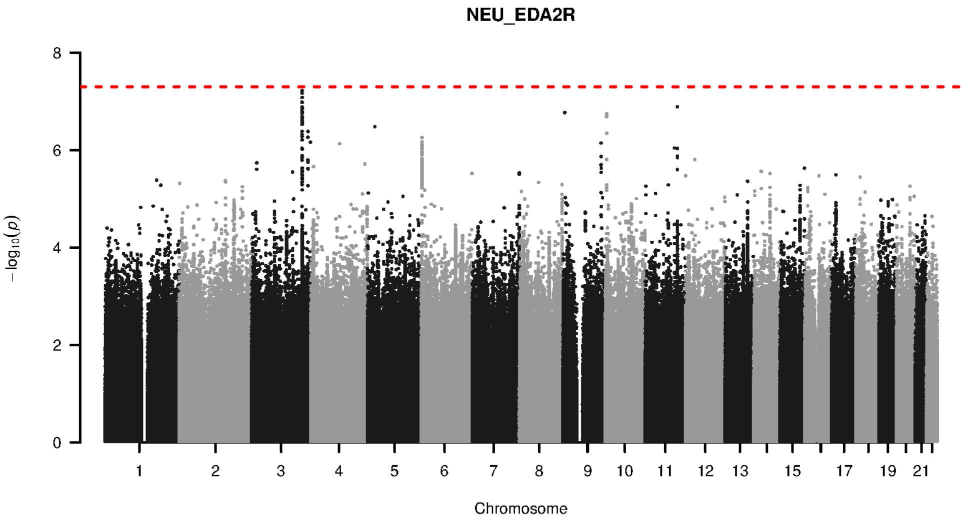


**Supplementary Figure 9-26. Manhattan plot and pQTL associations of the genome-wide meta-analysis for protein EDA2R.** The horizontal dashed line corresponds to the genome-wide significance threshold of $p = 5 \times 10^{-8}$. The variants in the cis-regulatory region (±500kb of the coding gene region) of the protein-coding gene are highlighted in green.

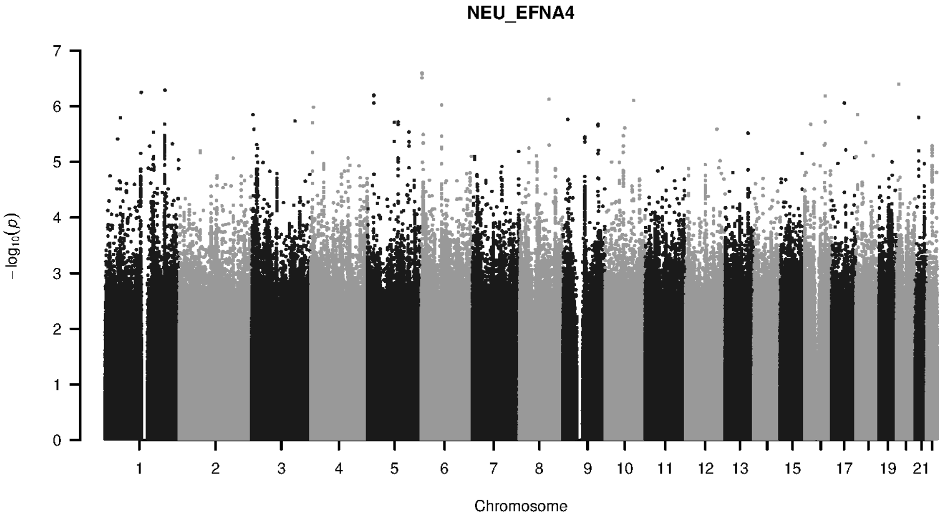


**Supplementary Figure 9-27. Manhattan plot and pQTL associations of the genome-wide meta-analysis for protein EFNA4.** The horizontal dashed line corresponds to the genome-wide significance threshold of $p = 5 \times 10^{-8}$. The variants in the cis-regulatory region (±500kb of the coding gene region) of the protein-coding gene are highlighted in green.

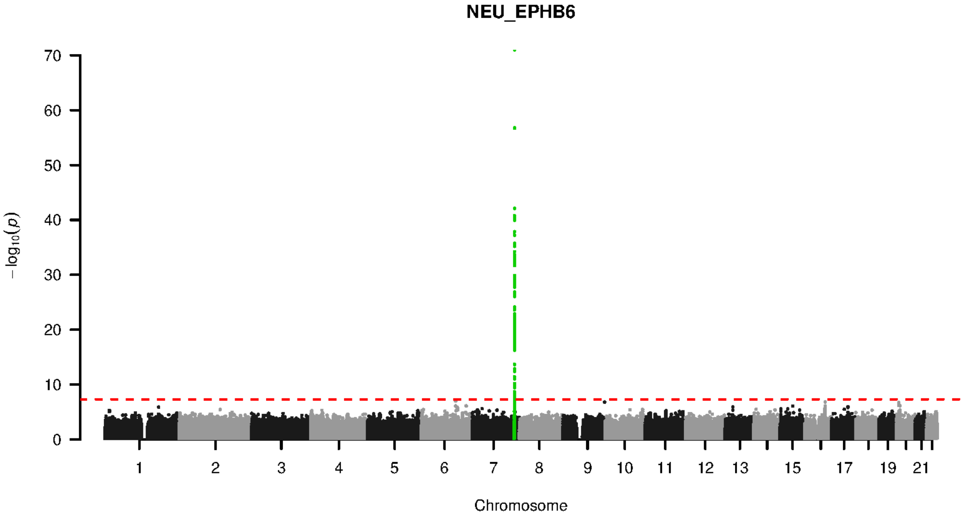


**Supplementary Figure 9-28. Manhattan plot and pQTL associations of the genome-wide meta-analysis for protein EPHB6.** The horizontal dashed line corresponds to the genome-wide significance threshold of $p = 5 \times 10^{-8}$. The variants in the cis-regulatory region (±500kb of the coding gene region) of the protein-coding gene are highlighted in green.

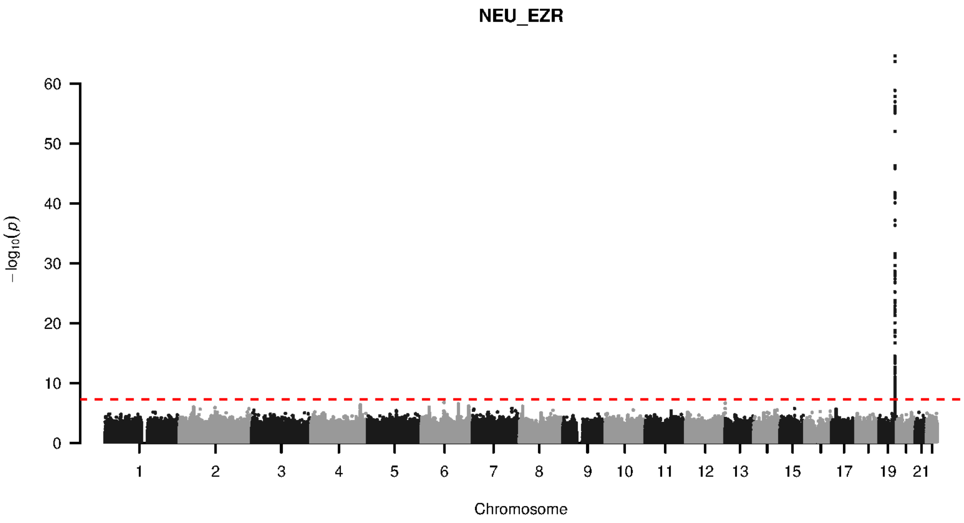


**Supplementary Figure 9-29. Manhattan plot and pQTL associations of the genome-wide meta-analysis for protein EZR.** The horizontal dashed line corresponds to the genome-wide significance threshold of $p = 5 \times 10^{-8}$. The variants in the cis-regulatory region (±500kb of the coding gene region) of the protein-coding gene are highlighted in green.

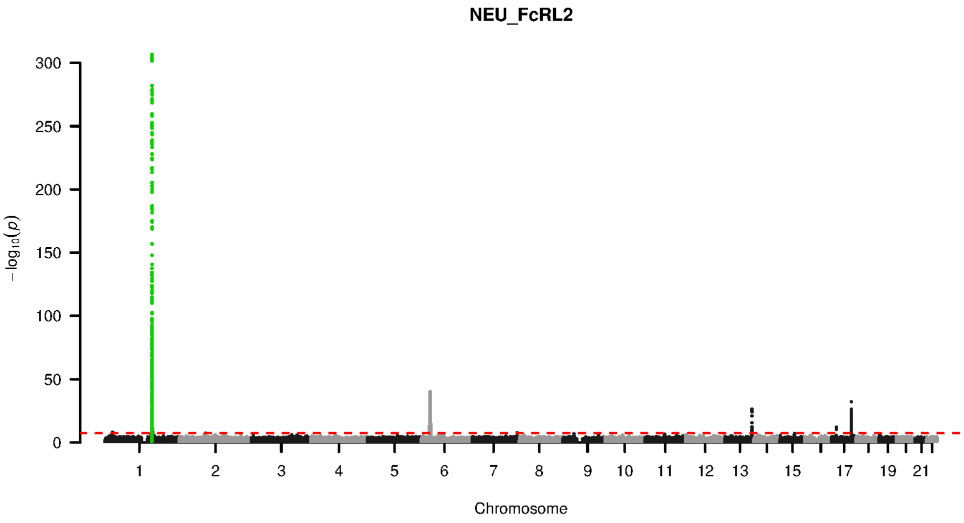


**Supplementary Figure 9-30. Manhattan plot and pQTL associations of the genome-wide meta-analysis for protein FCRL2.** The horizontal dashed line corresponds to the genome-wide significance threshold of $p = 5 \times 10^{-8}$. The variants in the cis-regulatory region (±500kb of the coding gene region) of the protein-coding gene are highlighted in green.

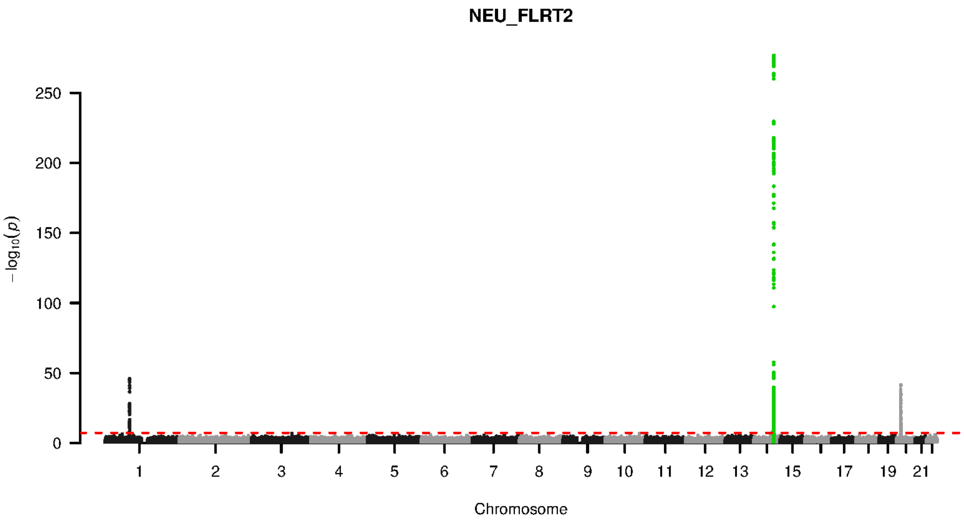


**Supplementary Figure 9-31. Manhattan plot and pQTL associations of the genome-wide meta-analysis for protein FLRT2.** The horizontal dashed line corresponds to the genome-wide significance threshold of $p = 5 \times 10^{-8}$. The variants in the cis-regulatory region (±500kb of the coding gene region) of the protein-coding gene are highlighted in green.

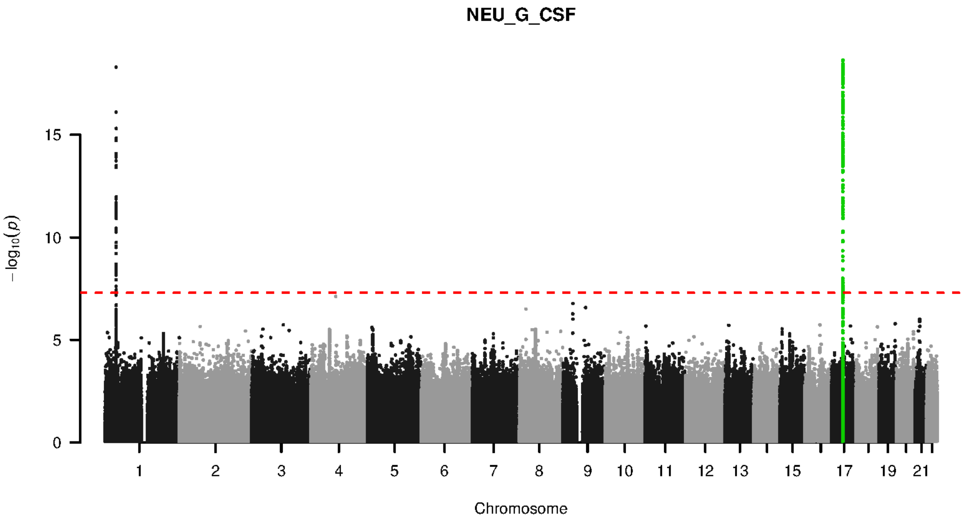


**Supplementary Figure 9-32. Manhattan plot and pQTL associations of the genome-wide meta-analysis for protein G CSF.** The horizontal dashed line corresponds to the genome-wide significance threshold of $p = 5 \times 10^{-8}$. The variants in the cis-regulatory region (±500kb of the coding gene region) of the protein-coding gene are highlighted in green.

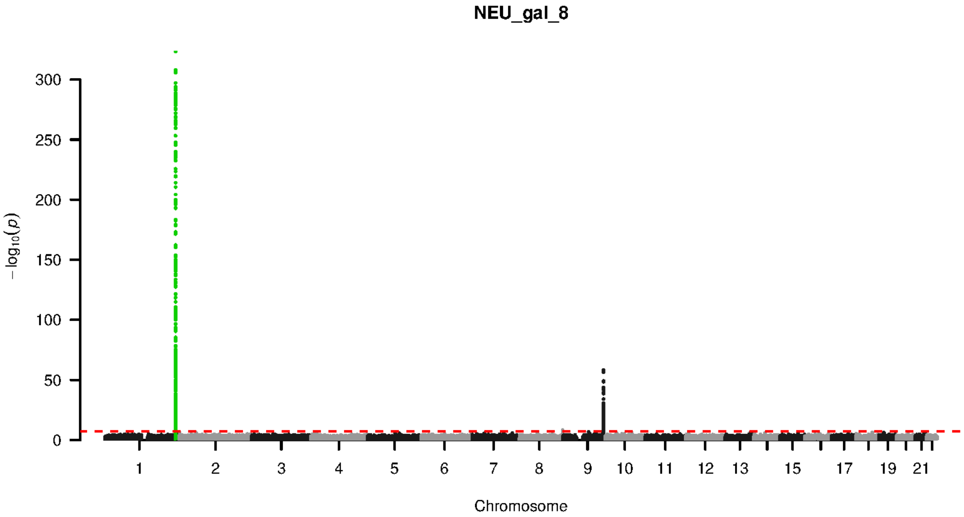


**Supplementary Figure 9-33. Manhattan plot and pQTL associations of the genome-wide meta-analysis for protein GAL 8.** The horizontal dashed line corresponds to the genome-wide significance threshold of $p = 5 \times 10^{-8}$. The variants in the cis-regulatory region (±500kb of the coding gene region) of the protein-coding gene are highlighted in green.

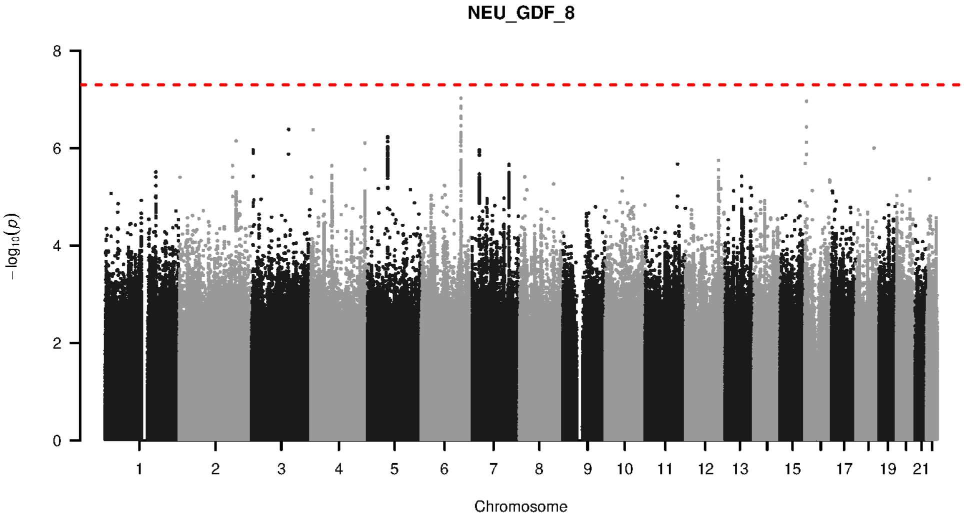


**Supplementary Figure 9-34. Manhattan plot and pQTL associations of the genome-wide meta-analysis for protein GDF 8.** The horizontal dashed line corresponds to the genome-wide significance threshold of $p = 5 \times 10^{-8}$. The variants in the cis-regulatory region (±500kb of the coding gene region) of the protein-coding gene are highlighted in green.

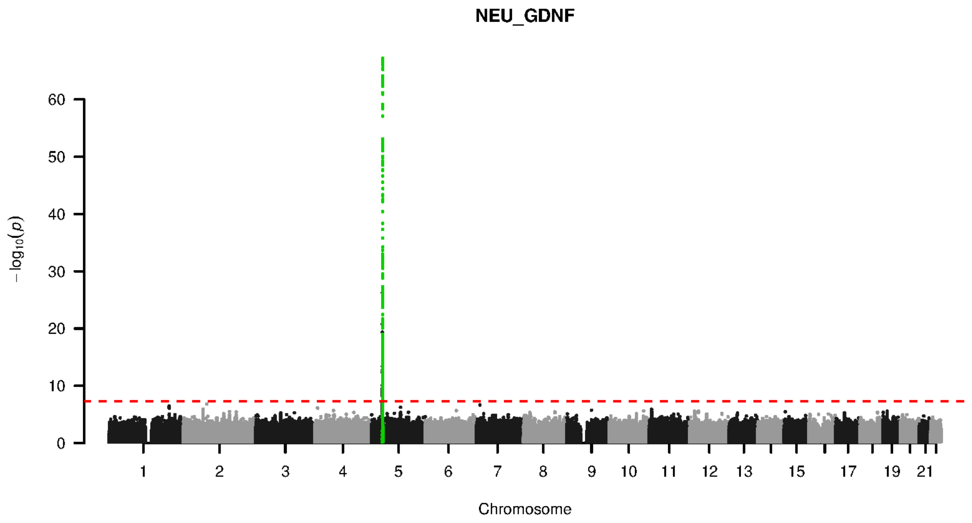


**Supplementary Figure 9-35. Manhattan plot and pQTL associations of the genome-wide meta-analysis for protein GDNF.** The horizontal dashed line corresponds to the genome-wide significance threshold of $p = 5 \times 10^{-8}$. The variants in the cis-regulatory region (±500kb of the coding gene region) of the protein-coding gene are highlighted in green.

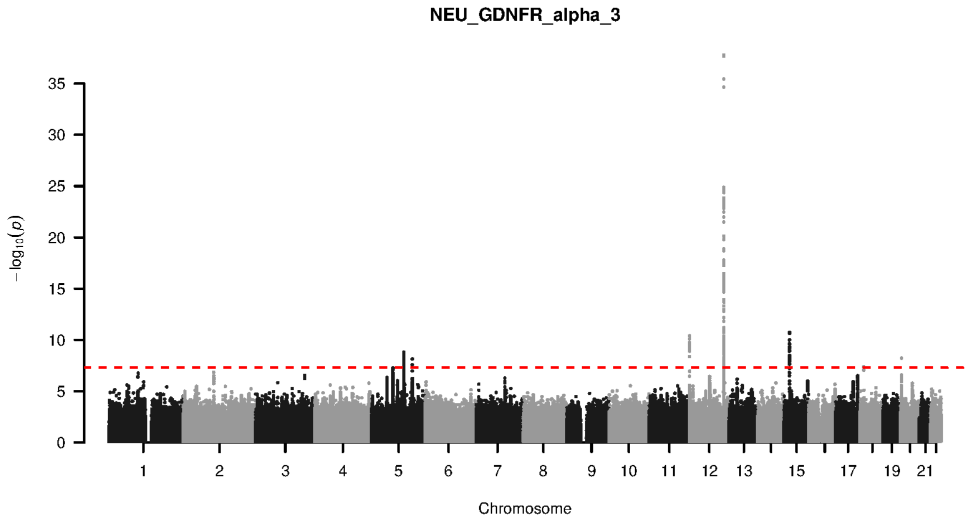


**Supplementary Figure 9-36. Manhattan plot and pQTL associations of the genome-wide meta-analysis for protein GDNFR ALPHA 3.** The horizontal dashed line corresponds to the genome-wide significance threshold of $p = 5 \times 10^{-8}$. The variants in the cis-regulatory region (±500kb of the coding gene region) of the protein-coding gene are highlighted in green.

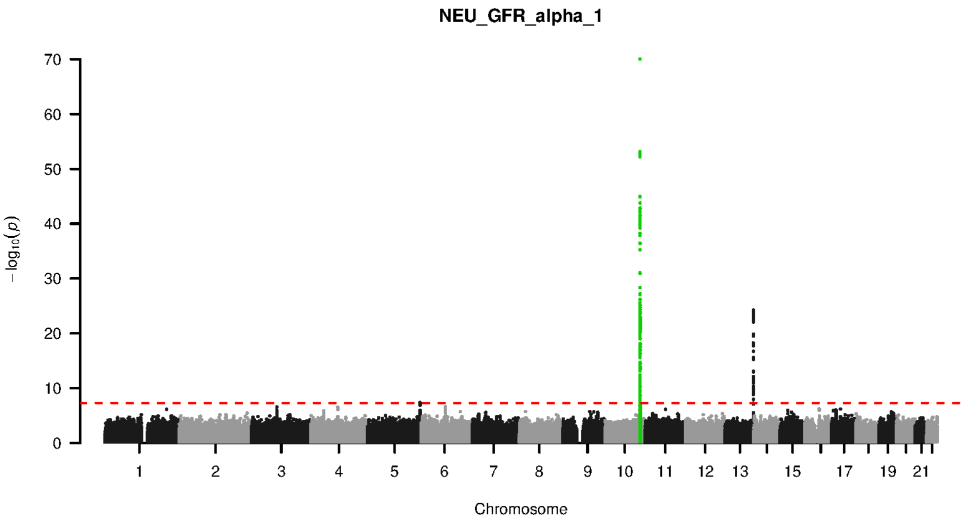


**Supplementary Figure 9-37. Manhattan plot and pQTL associations of the genome-wide meta-analysis for protein GFR ALPHA 1.** The horizontal dashed line corresponds to the genome-wide significance threshold of $p = 5 \times 10^{-8}$. The variants in the cis-regulatory region (±500kb of the coding gene region) of the protein-coding gene are highlighted in green.

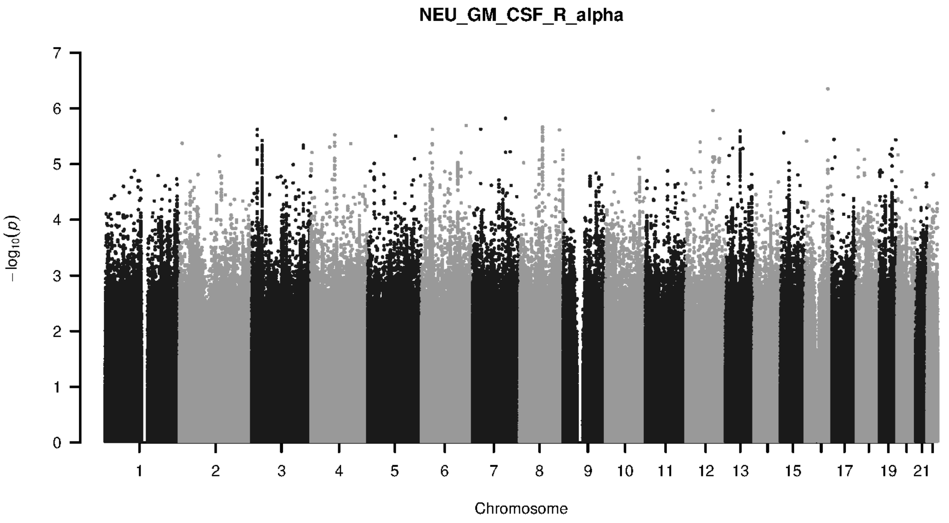


**Supplementary Figure 9-38. Manhattan plot and pQTL associations of the genome-wide meta-analysis for protein GM CSF R ALPHA.** The horizontal dashed line corresponds to the genome-wide significance threshold of $p = 5 \times 10^{-8}$. The variants in the cis-regulatory region (±500kb of the coding gene region) of the protein-coding gene are highlighted in green.

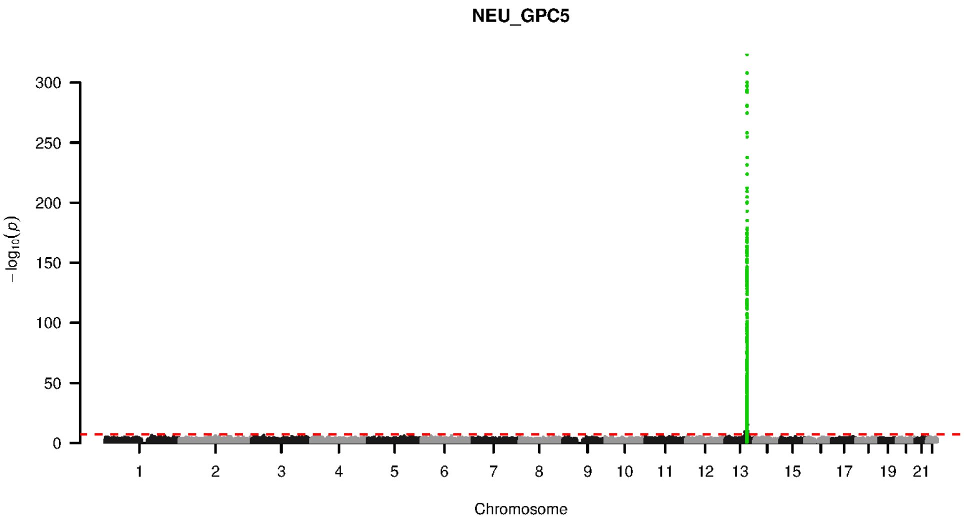


**Supplementary Figure 9-39. Manhattan plot and pQTL associations of the genome-wide meta-analysis for protein GPC5.** The horizontal dashed line corresponds to the genome-wide significance threshold of $p = 5 \times 10^{-8}$. The variants in the cis-regulatory region (±500kb of the coding gene region) of the protein-coding gene are highlighted in green.

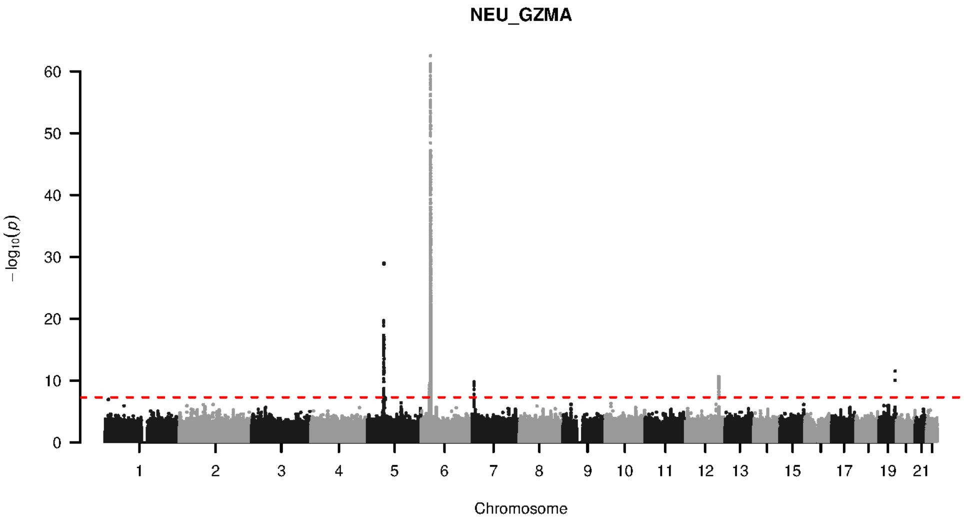


**Supplementary Figure 9-40. Manhattan plot and pQTL associations of the genome-wide meta-analysis for protein GZMA.** The horizontal dashed line corresponds to the genome-wide significance threshold of $p = 5 \times 10^{-8}$. The variants in the cis-regulatory region (±500kb of the coding gene region) of the protein-coding gene are highlighted in green.

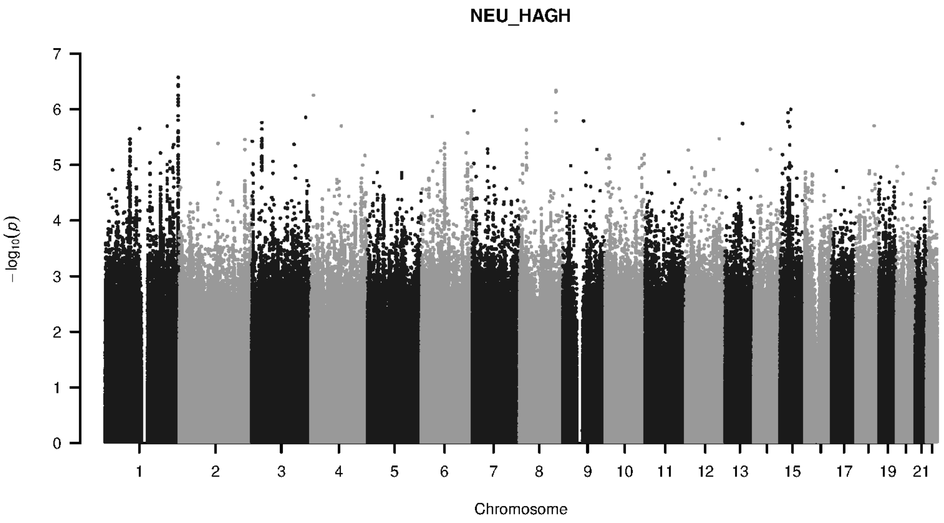


**Supplementary Figure 9-41. Manhattan plot and pQTL associations of the genome-wide meta-analysis for protein HAGH.** The horizontal dashed line corresponds to the genome-wide significance threshold of $p = 5 \times 10^{-8}$. The variants in the cis-regulatory region (±500kb of the coding gene region) of the protein-coding gene are highlighted in green.

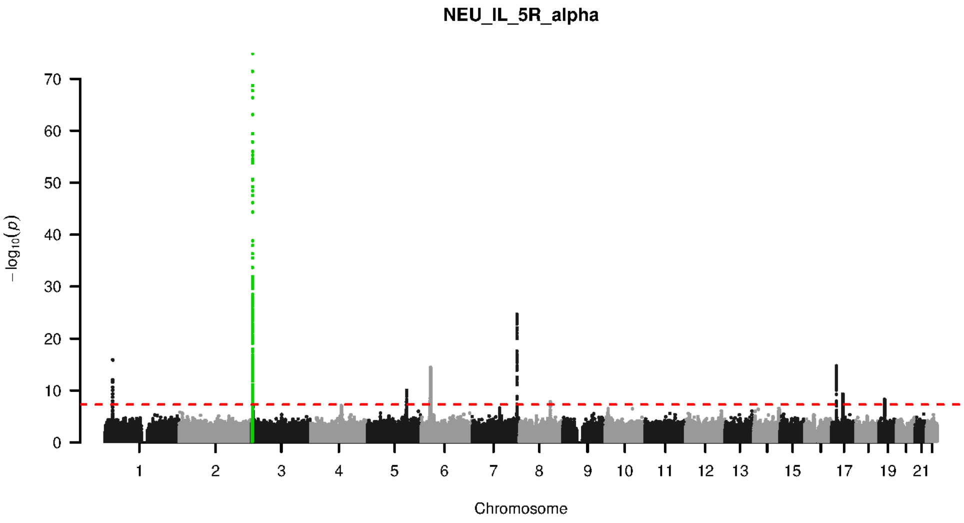


**Supplementary Figure 9-42. Manhattan plot and pQTL associations of the genome-wide meta-analysis for protein IL 5R ALPHA.** The horizontal dashed line corresponds to the genome-wide significance threshold of $p = 5 \times 10^{-8}$. The variants in the cis-regulatory region (±500kb of the coding gene region) of the protein-coding gene are highlighted in green.

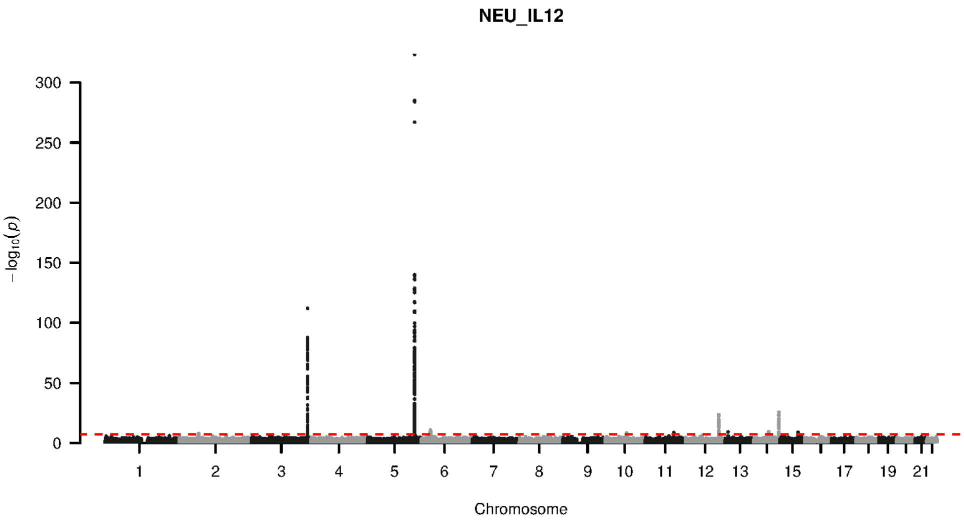


**Supplementary Figure 9-43. Manhattan plot and pQTL associations of the genome-wide meta-analysis for protein IL12.** The horizontal dashed line corresponds to the genome-wide significance threshold of $p = 5 \times 10^{-8}$. The variants in the cis-regulatory region (±500kb of the coding gene region) of the protein-coding gene are highlighted in green.

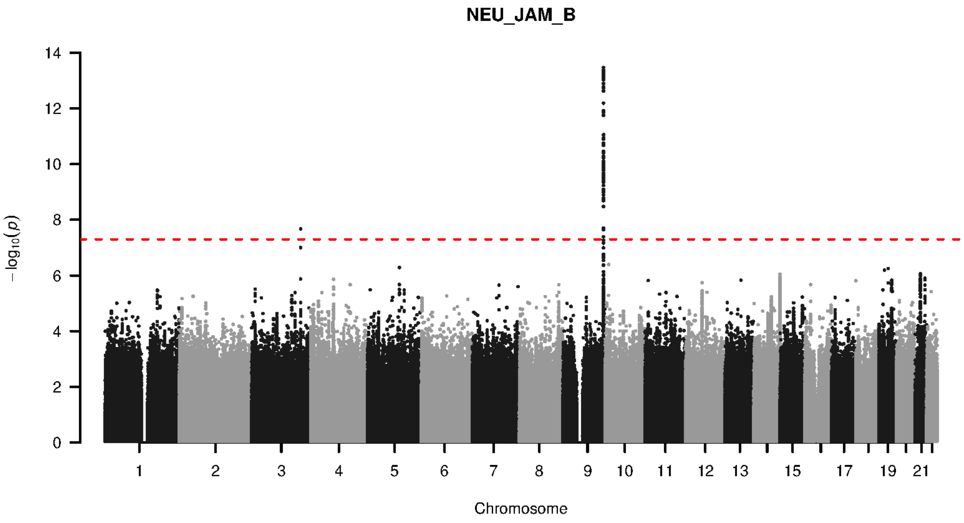


**Supplementary Figure 9-44. Manhattan plot and pQTL associations of the genome-wide meta-analysis for protein JAM B.** The horizontal dashed line corresponds to the genome-wide significance threshold of $p = 5 \times 10^{-8}$. The variants in the cis-regulatory region (±500kb of the coding gene region) of the protein-coding gene are highlighted in green.

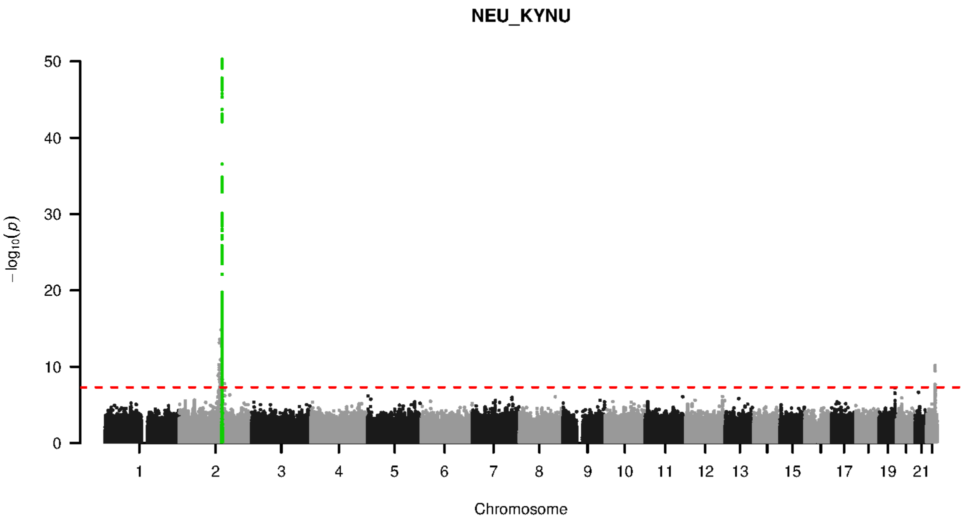


**Supplementary Figure 9-45. Manhattan plot and pQTL associations of the genome-wide meta-analysis for protein KYNU.** The horizontal dashed line corresponds to the genome-wide significance threshold of $p = 5 \times 10^{-8}$. The variants in the cis-regulatory region (±500kb of the coding gene region) of the protein-coding gene are highlighted in green.

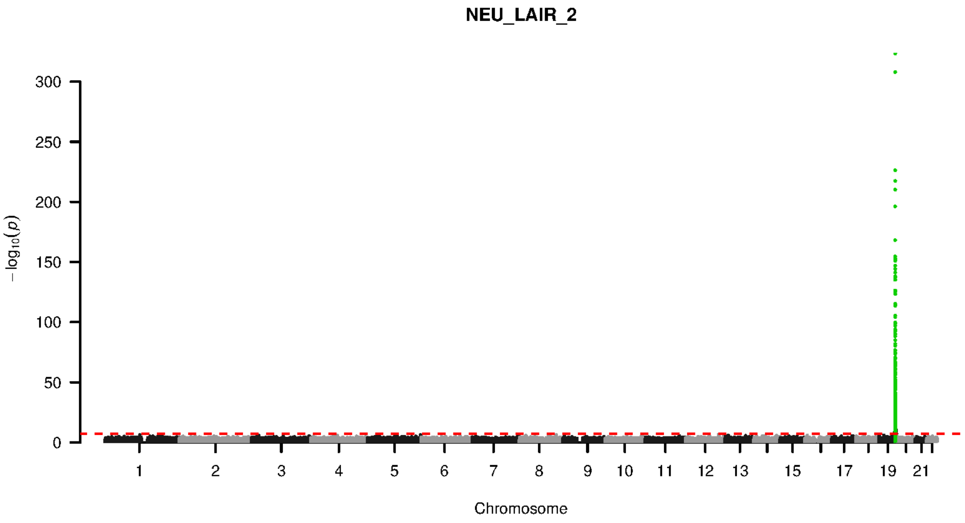


**Supplementary Figure 9-46. Manhattan plot and pQTL associations of the genome-wide meta-analysis for protein LAIR 2.** The horizontal dashed line corresponds to the genome-wide significance threshold of $p = 5 \times 10^{-8}$. The variants in the cis-regulatory region (±500kb of the coding gene region) of the protein-coding gene are highlighted in green.

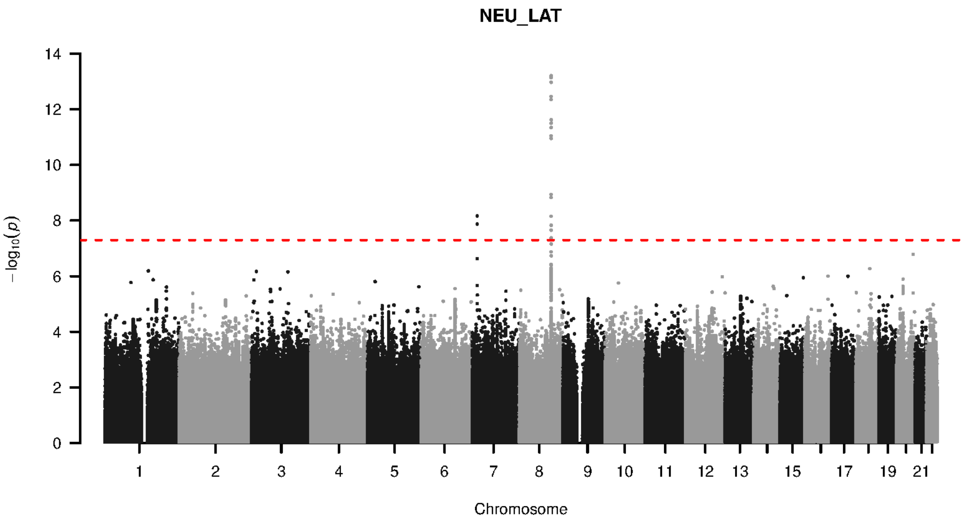


**Supplementary Figure 9-47. Manhattan plot and pQTL associations of the genome-wide meta-analysis for protein LAT.** The horizontal dashed line corresponds to the genome-wide significance threshold of $p = 5 \times 10^{-8}$. The variants in the cis-regulatory region (±500kb of the coding gene region) of the protein-coding gene are highlighted in green.

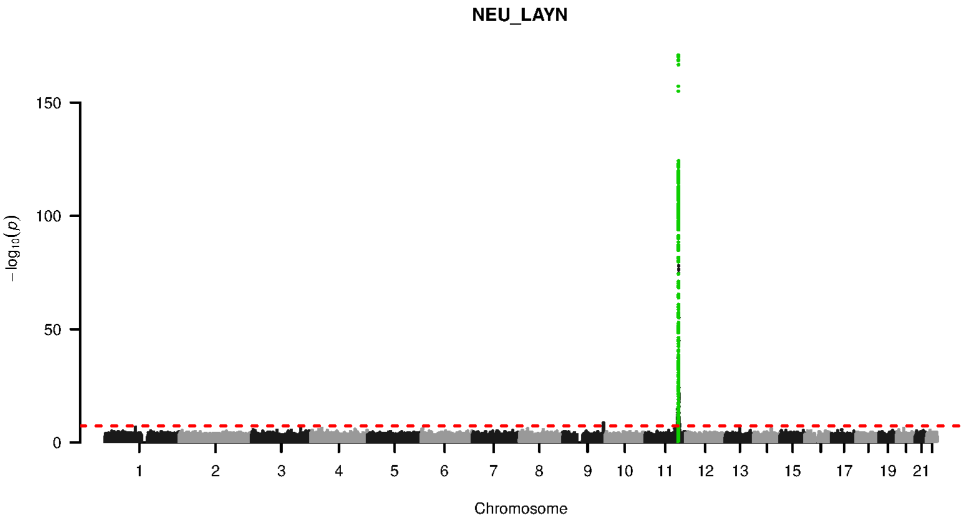


**Supplementary Figure 9-48. Manhattan plot and pQTL associations of the genome-wide meta-analysis for protein LAYN.** The horizontal dashed line corresponds to the genome-wide significance threshold of $p = 5 \times 10^{-8}$. The variants in the cis-regulatory region (±500kb of the coding gene region) of the protein-coding gene are highlighted in green.

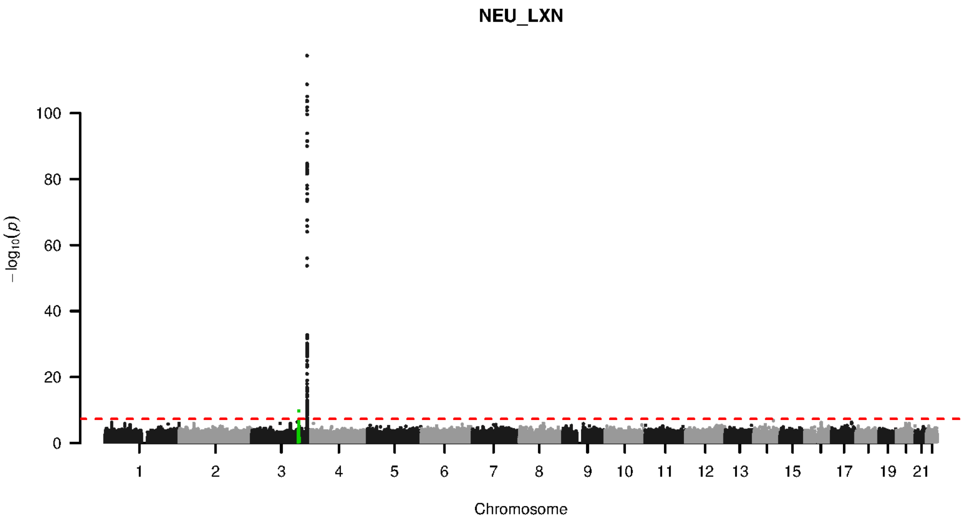


**Supplementary Figure 9-49. Manhattan plot and pQTL associations of the genome-wide meta-analysis for protein LXN.** The horizontal dashed line corresponds to the genome-wide significance threshold of $p = 5 \times 10^{-8}$. The variants in the cis-regulatory region (±500kb of the coding gene region) of the protein-coding gene are highlighted in green.

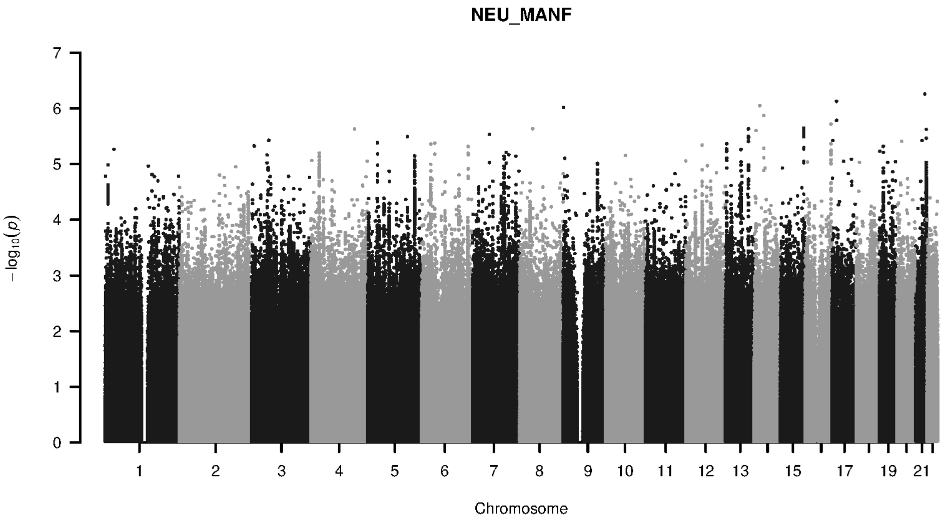


**Supplementary Figure 9-50. Manhattan plot and pQTL associations of the genome-wide meta-analysis for protein MANF.** The horizontal dashed line corresponds to the genome-wide significance threshold of $p = 5 \times 10^{-8}$. The variants in the cis-regulatory region (±500kb of the coding gene region) of the protein-coding gene are highlighted in green.

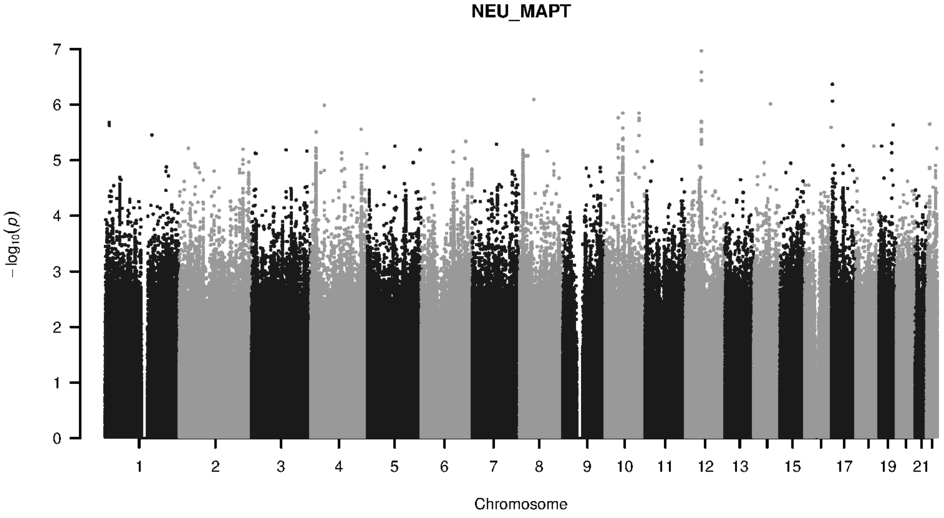


**Supplementary Figure 9-51. Manhattan plot and pQTL associations of the genome-wide meta-analysis for protein MAPT.** The horizontal dashed line corresponds to the genome-wide significance threshold of $p = 5 \times 10^{-8}$. The variants in the cis-regulatory region (±500kb of the coding gene region) of the protein-coding gene are highlighted in green.

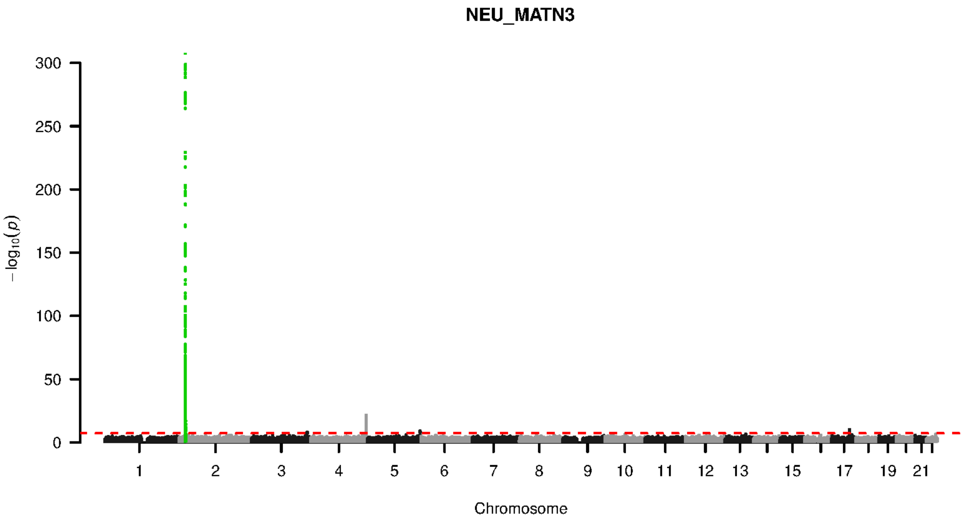


**Supplementary Figure 9-52. Manhattan plot and pQTL associations of the genome-wide meta-analysis for protein MATN3.** The horizontal dashed line corresponds to the genome-wide significance threshold of $p = 5 \times 10^{-8}$. The variants in the cis-regulatory region (±500kb of the coding gene region) of the protein-coding gene are highlighted in green.

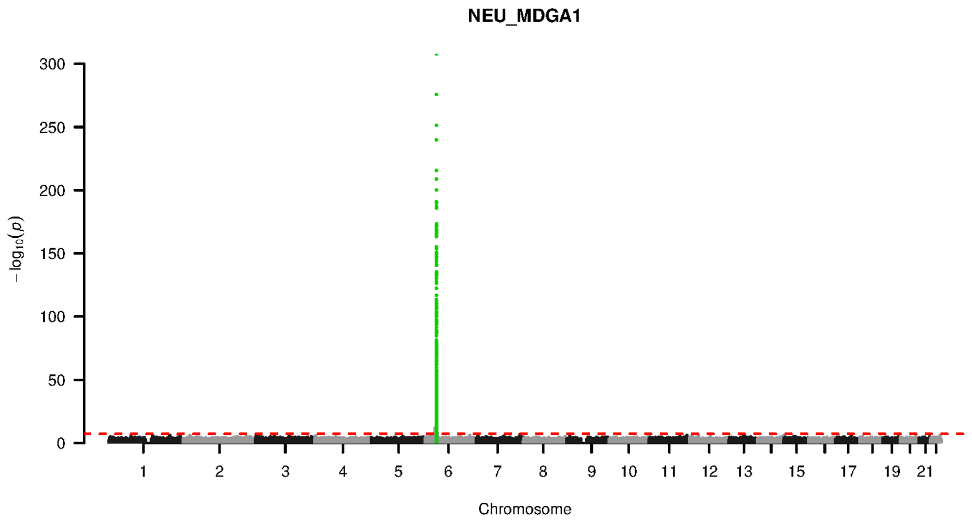


**Supplementary Figure 9-53. Manhattan plot and pQTL associations of the genome-wide meta-analysis for protein MDGA1.** The horizontal dashed line corresponds to the genome-wide significance threshold of $p = 5 \times 10^{-8}$. The variants in the cis-regulatory region (±500kb of the coding gene region) of the protein-coding gene are highlighted in green.

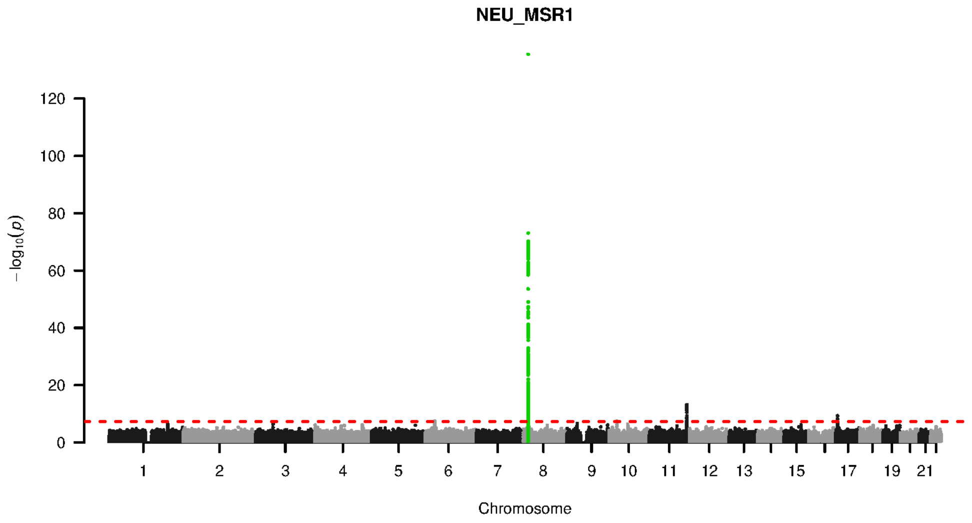


**Supplementary Figure 9-54. Manhattan plot and pQTL associations of the genome-wide meta-analysis for protein MSR1.** The horizontal dashed line corresponds to the genome-wide significance threshold of $p = 5 \times 10^{-8}$. The variants in the cis-regulatory region (±500kb of the coding gene region) of the protein-coding gene are highlighted in green.

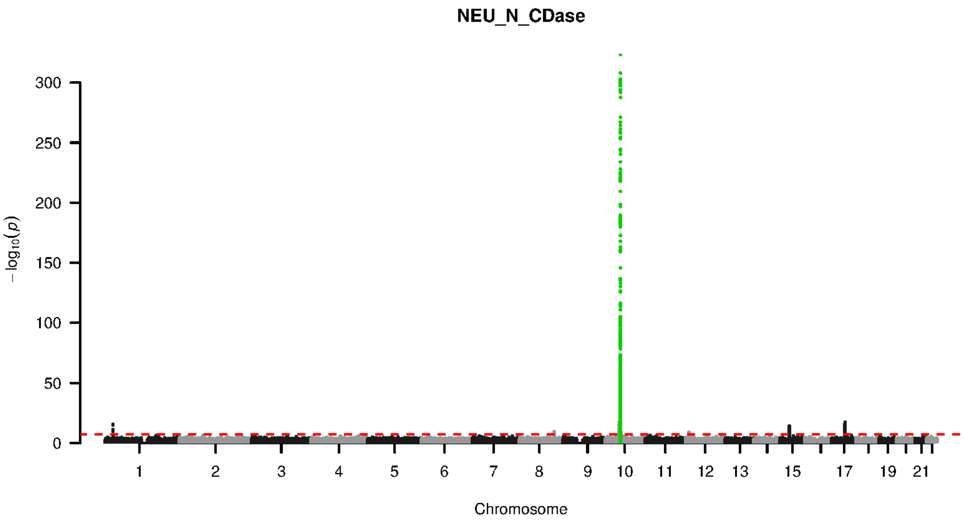


**Supplementary Figure 9-55. Manhattan plot and pQTL associations of the genome-wide meta-analysis for protein N CDASE.** The horizontal dashed line corresponds to the genome-wide significance threshold of $p = 5 \times 10^{-8}$. The variants in the cis-regulatory region (±500kb of the coding gene region) of the protein-coding gene are highlighted in green.

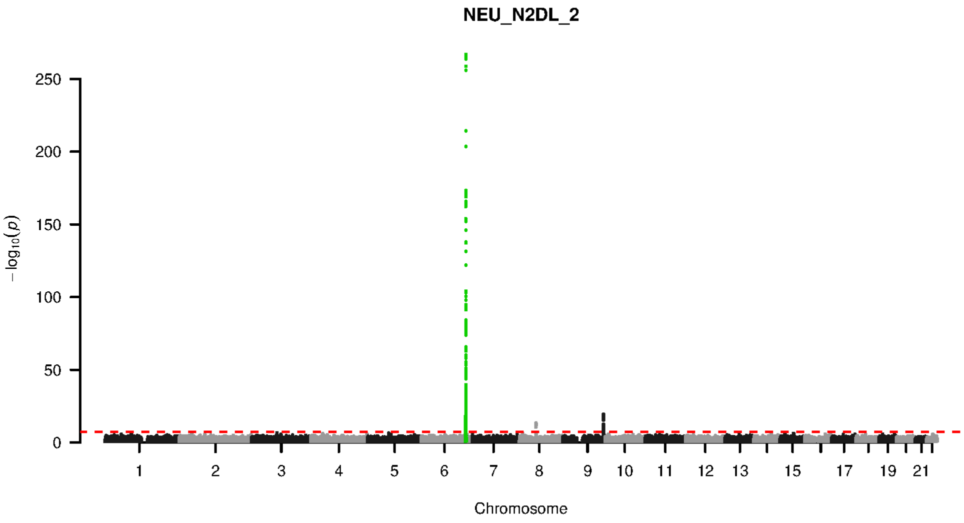


**Supplementary Figure 9-56. Manhattan plot and pQTL associations of the genome-wide meta-analysis for protein N2DL 2.** The horizontal dashed line corresponds to the genome-wide significance threshold of $p = 5 \times 10^{-8}$. The variants in the cis-regulatory region (±500kb of the coding gene region) of the protein-coding gene are highlighted in green.

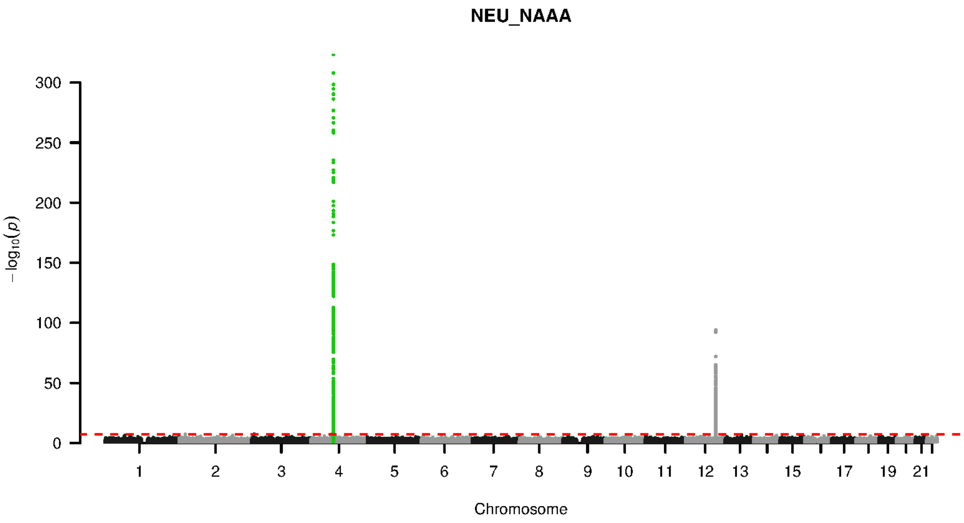


**Supplementary Figure 9-57. Manhattan plot and pQTL associations of the genome-wide meta-analysis for protein NAAA.** The horizontal dashed line corresponds to the genome-wide significance threshold of $p = 5 \times 10^{-8}$. The variants in the cis-regulatory region (±500kb of the coding gene region) of the protein-coding gene are highlighted in green.

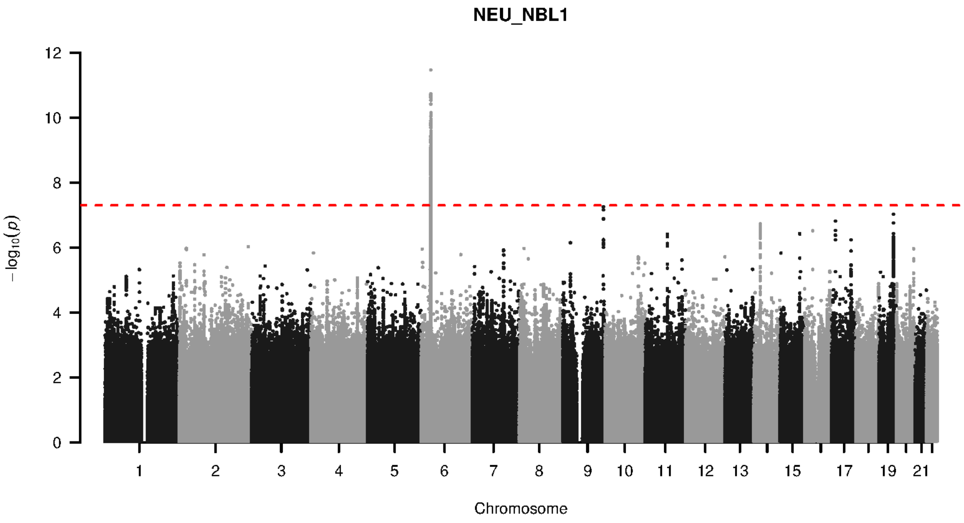


**Supplementary Figure 9-58. Manhattan plot and pQTL associations of the genome-wide meta-analysis for protein NBL1.** The horizontal dashed line corresponds to the genome-wide significance threshold of $p = 5 \times 10^{-8}$. The variants in the cis-regulatory region (±500kb of the coding gene region) of the protein-coding gene are highlighted in green.

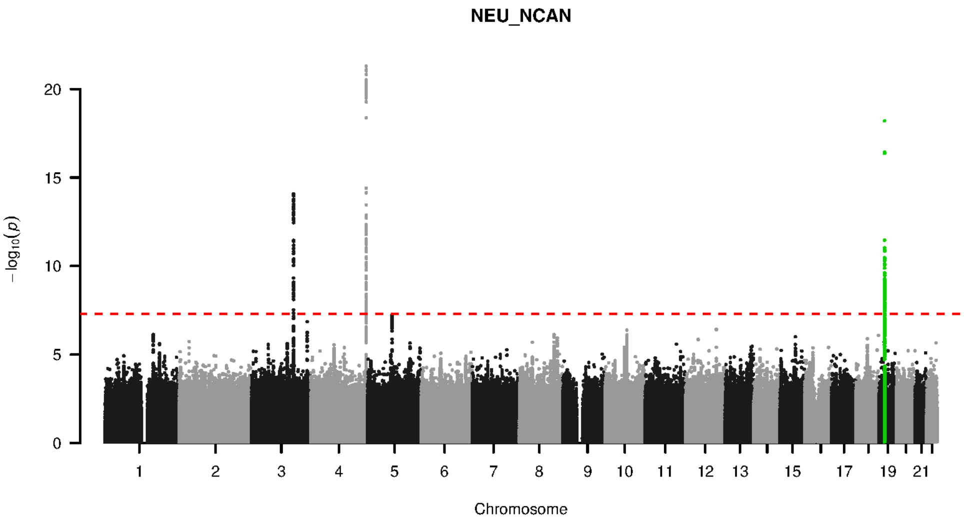


**Supplementary Figure 9-59. Manhattan plot and pQTL associations of the genome-wide meta-analysis for protein NCAN.** The horizontal dashed line corresponds to the genome-wide significance threshold of $p = 5 \times 10^{-8}$. The variants in the cis-regulatory region (±500kb of the coding gene region) of the protein-coding gene are highlighted in green.

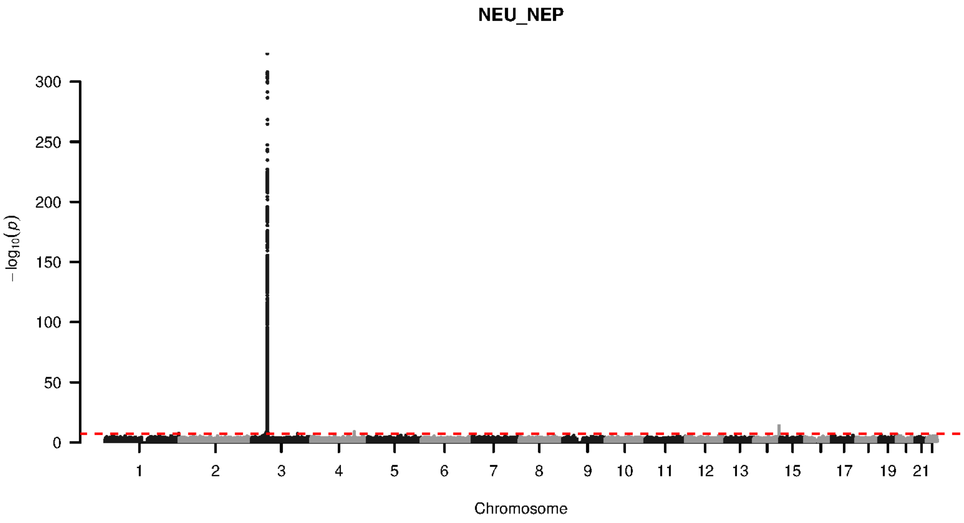


**Supplementary Figure 9-60. Manhattan plot and pQTL associations of the genome-wide meta-analysis for protein NEP.** The horizontal dashed line corresponds to the genome-wide significance threshold of $p = 5 \times 10^{-8}$. The variants in the cis-regulatory region (±500kb of the coding gene region) of the protein-coding gene are highlighted in green.

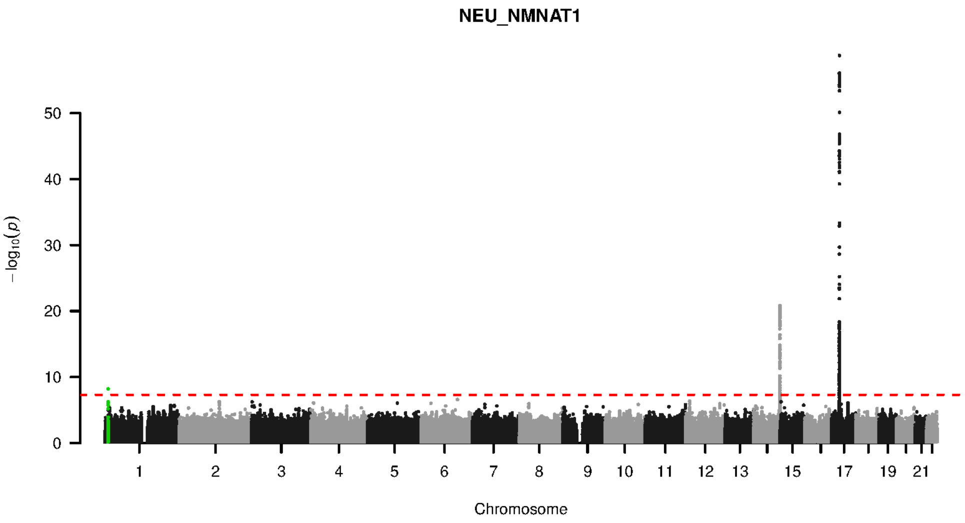


**Supplementary Figure 9-61. Manhattan plot and pQTL associations of the genome-wide meta-analysis for protein NMNAT1.** The horizontal dashed line corresponds to the genome-wide significance threshold of $p = 5 \times 10^{-8}$. The variants in the cis-regulatory region (±500kb of the coding gene region) of the protein-coding gene are highlighted in green.

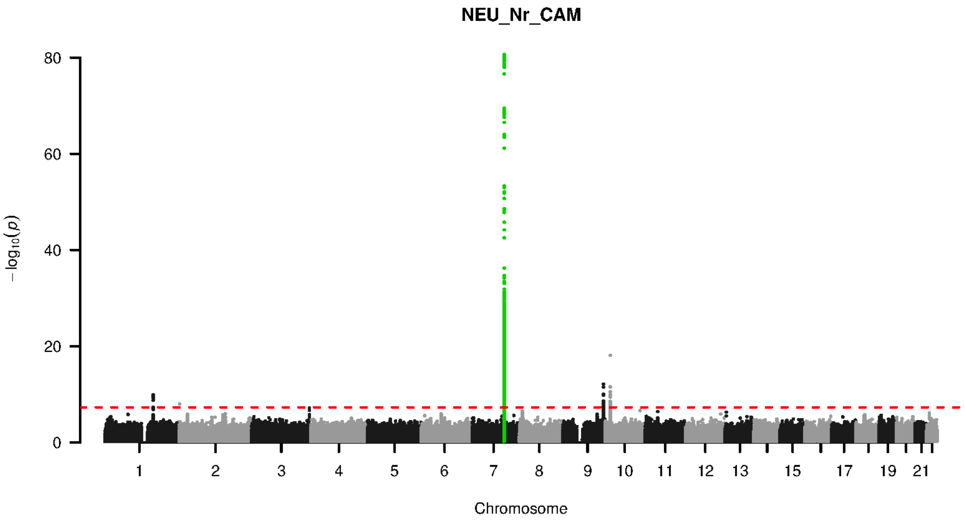


**Supplementary Figure 9-62. Manhattan plot and pQTL associations of the genome-wide meta-analysis for protein NR CAM.** The horizontal dashed line corresponds to the genome-wide significance threshold of $p = 5 \times 10^{-8}$. The variants in the cis-regulatory region (±500kb of the coding gene region) of the protein-coding gene are highlighted in green.

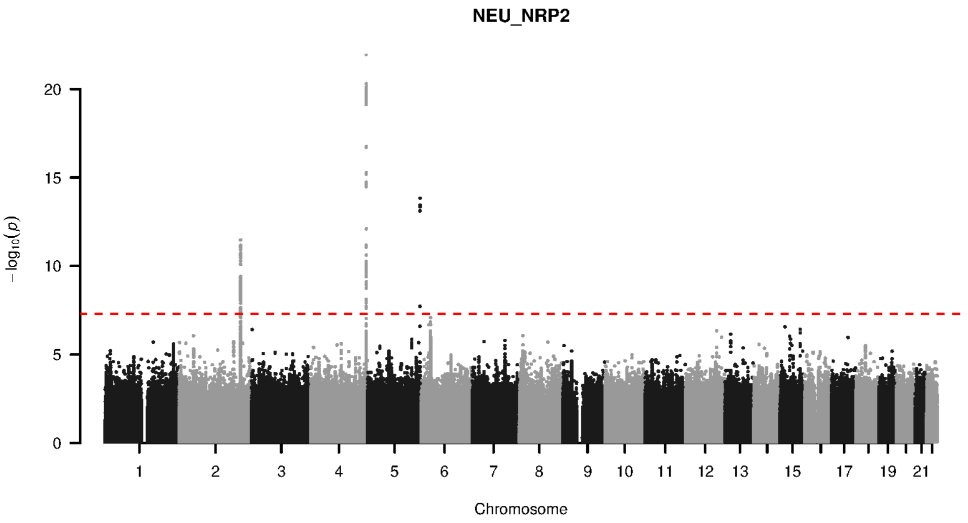


**Supplementary Figure 9-63. Manhattan plot and pQTL associations of the genome-wide meta-analysis for protein NRP2.** The horizontal dashed line corresponds to the genome-wide significance threshold of $p = 5 \times 10^{-8}$. The variants in the cis-regulatory region ($\pm$500kb of the coding gene region) of the protein-coding gene are highlighted in green.

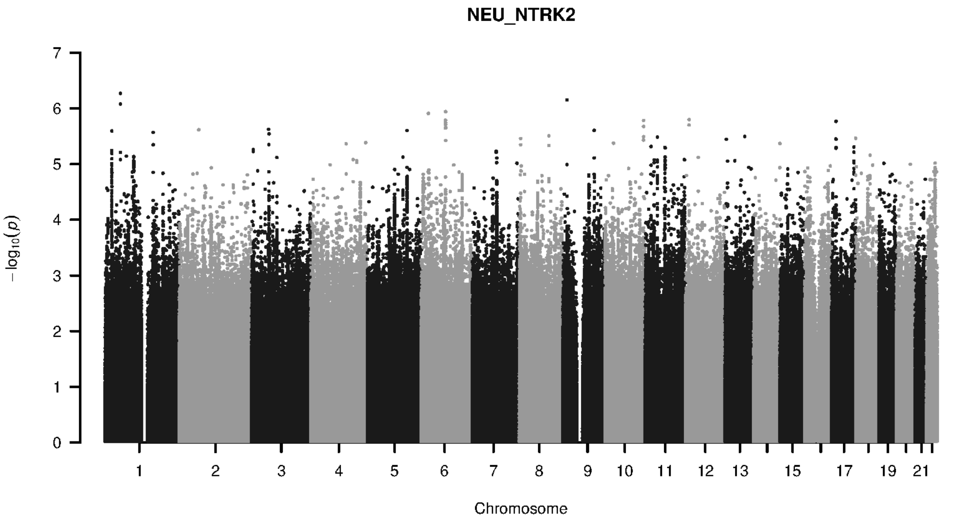


**Supplementary Figure 9-64. Manhattan plot and pQTL associations of the genome-wide meta-analysis for protein NTRK2.** The horizontal dashed line corresponds to the genome-wide significance threshold of $p = 5 \times 10^{-8}$. The variants in the cis-regulatory region ($\pm$500kb of the coding gene region) of the protein-coding gene are highlighted in green.

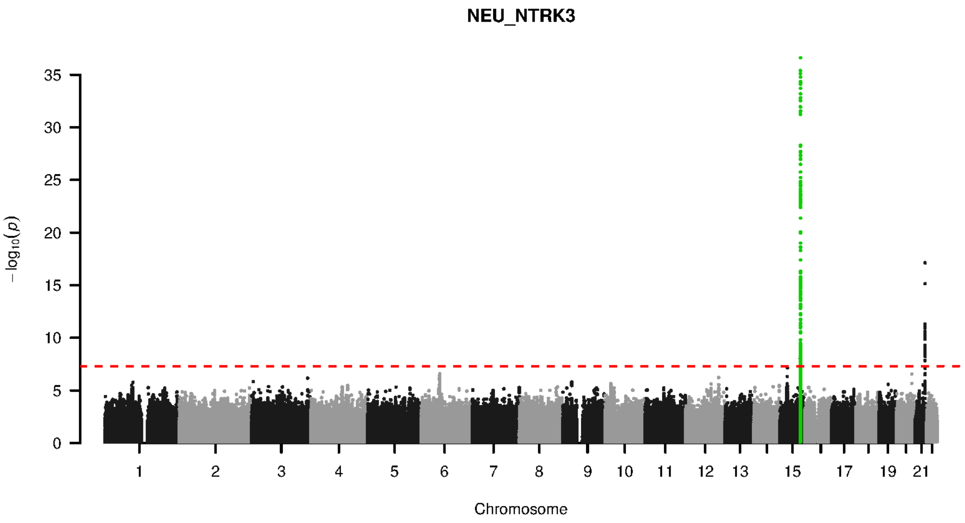


**Supplementary Figure 9-65. Manhattan plot and pQTL associations of the genome-wide meta-analysis for protein NTRK3.** The horizontal dashed line corresponds to the genome-wide significance threshold of $p = 5 \times 10^{-8}$. The variants in the cis-regulatory region (±500kb of the coding gene region) of the protein-coding gene are highlighted in green.

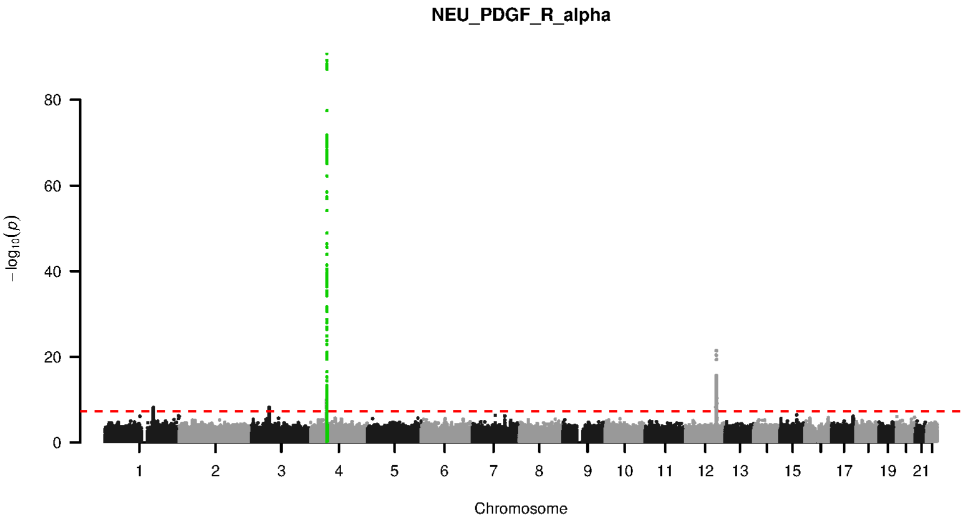


**Supplementary Figure 9-66. Manhattan plot and pQTL associations of the genome-wide meta-analysis for protein PDGF R ALPHA.** The horizontal dashed line corresponds to the genome-wide significance threshold of $p = 5 \times 10^{-8}$. The variants in the cis-regulatory region (±500kb of the coding gene region) of the protein-coding gene are highlighted in green.

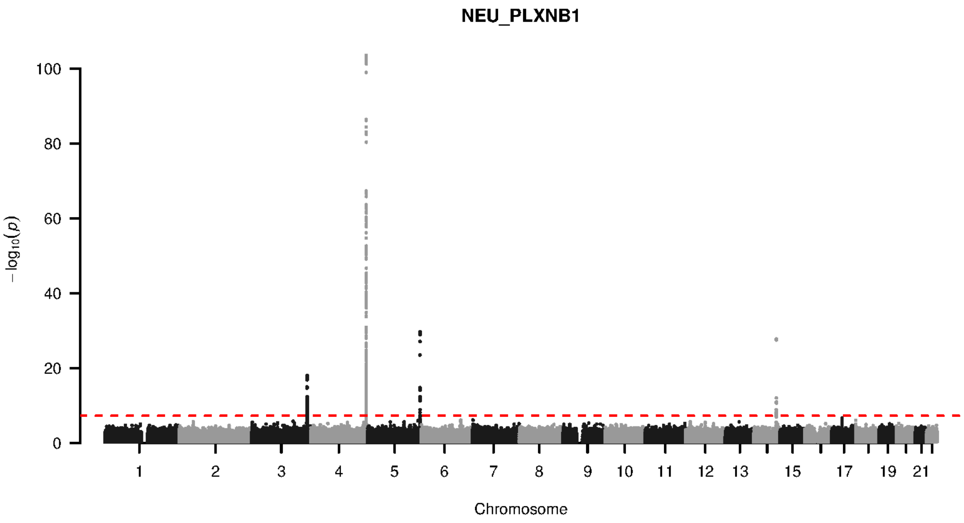


**Supplementary Figure 9-67. Manhattan plot and pQTL associations of the genome-wide meta-analysis for protein PLXNB1.** The horizontal dashed line corresponds to the genome-wide significance threshold of $p = 5 \times 10^{-8}$. The variants in the cis-regulatory region (±500kb of the coding gene region) of the protein-coding gene are highlighted in green.

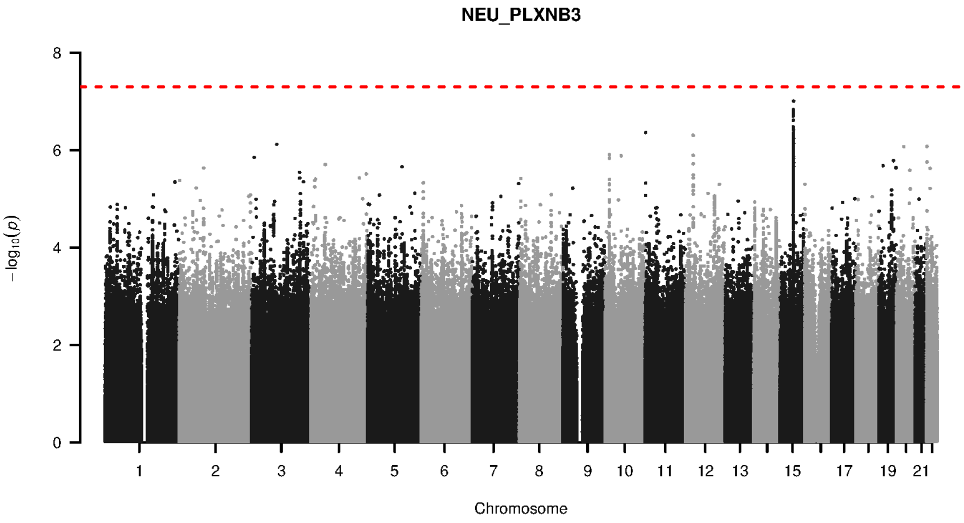


**Supplementary Figure 9-68. Manhattan plot and pQTL associations of the genome-wide meta-analysis for protein PLXNB3.** The horizontal dashed line corresponds to the genome-wide significance threshold of $p = 5 \times 10^{-8}$. The variants in the cis-regulatory region (±500kb of the coding gene region) of the protein-coding gene are highlighted in green.

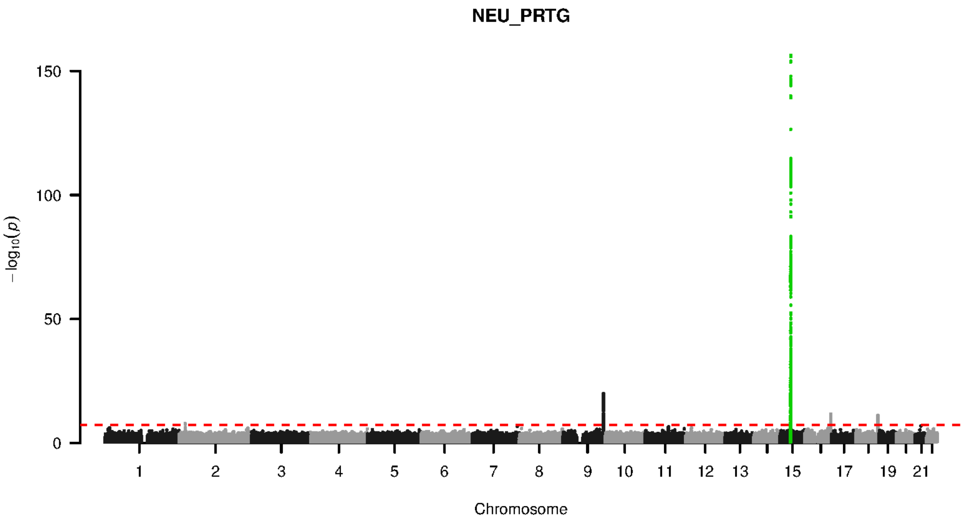


**Supplementary Figure 9-69. Manhattan plot and pQTL associations of the genome-wide meta-analysis for protein PRTG.** The horizontal dashed line corresponds to the genome-wide significance threshold of $p = 5 \times 10^{-8}$. The variants in the cis-regulatory region (±500kb of the coding gene region) of the protein-coding gene are highlighted in green.

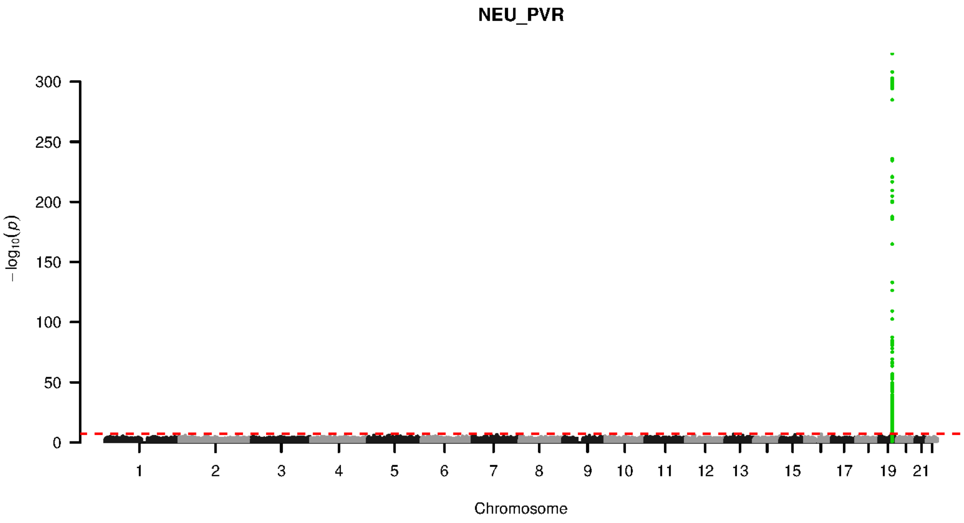


**Supplementary Figure 9-70. Manhattan plot and pQTL associations of the genome-wide meta-analysis for protein PVR.** The horizontal dashed line corresponds to the genome-wide significance threshold of $p = 5 \times 10^{-8}$. The variants in the cis-regulatory region (±500kb of the coding gene region) of the protein-coding gene are highlighted in green.

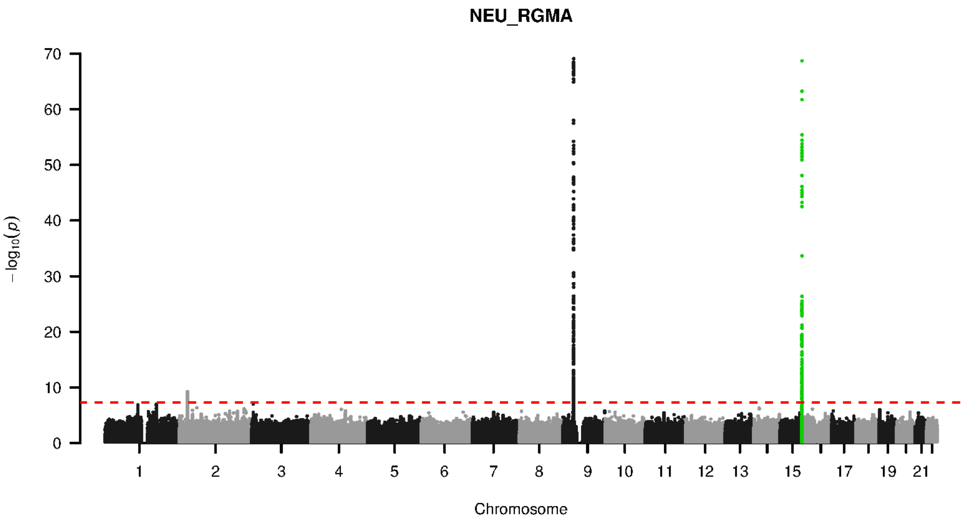


**Supplementary Figure 9-71. Manhattan plot and pQTL associations of the genome-wide meta-analysis for protein RGMA.** The horizontal dashed line corresponds to the genome-wide significance threshold of $p = 5 \times 10^{-8}$. The variants in the cis-regulatory region (±500kb of the coding gene region) of the protein-coding gene are highlighted in green.

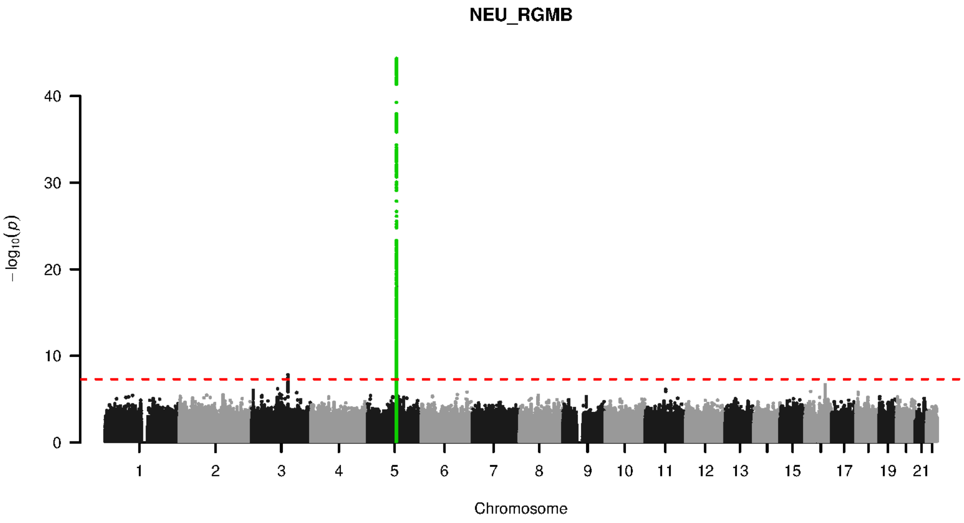


**Supplementary Figure 9-72. Manhattan plot and pQTL associations of the genome-wide meta-analysis for protein RGMB.** The horizontal dashed line corresponds to the genome-wide significance threshold of $p = 5 \times 10^{-8}$. The variants in the cis-regulatory region (±500kb of the coding gene region) of the protein-coding gene are highlighted in green.

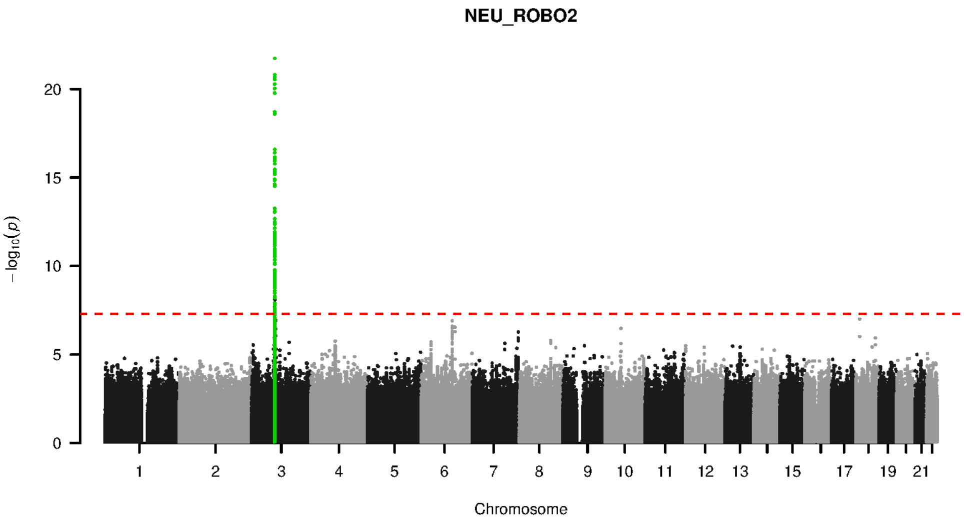


**Supplementary Figure 9-73. Manhattan plot and pQTL associations of the genome-wide meta-analysis for protein ROBO2.** The horizontal dashed line corresponds to the genome-wide significance threshold of $p = 5 \times 10^{-8}$. The variants in the cis-regulatory region (±500kb of the coding gene region) of the protein-coding gene are highlighted in green.

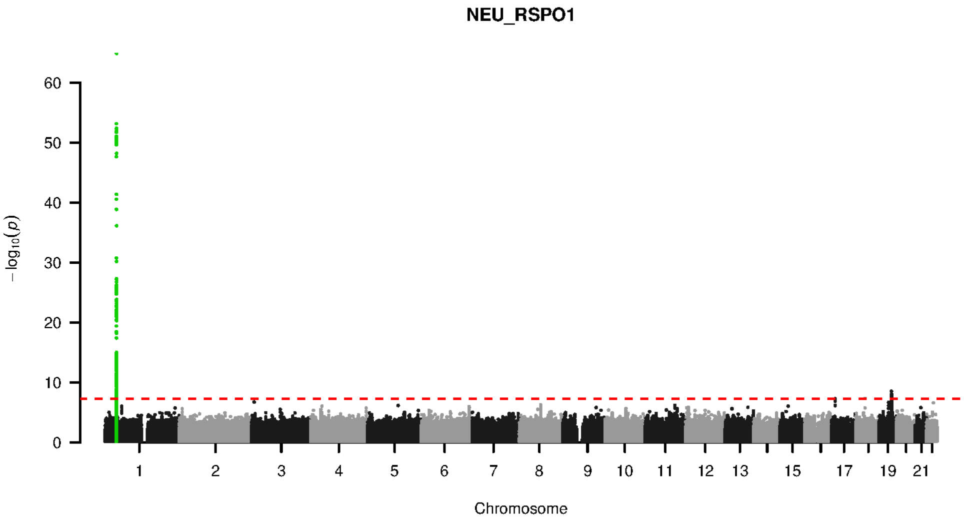


**Supplementary Figure 9-74. Manhattan plot and pQTL associations of the genome-wide meta-analysis for protein RSPO1.** The horizontal dashed line corresponds to the genome-wide significance threshold of $p = 5 \times 10^{-8}$. The variants in the cis-regulatory region (±500kb of the coding gene region) of the protein-coding gene are highlighted in green.

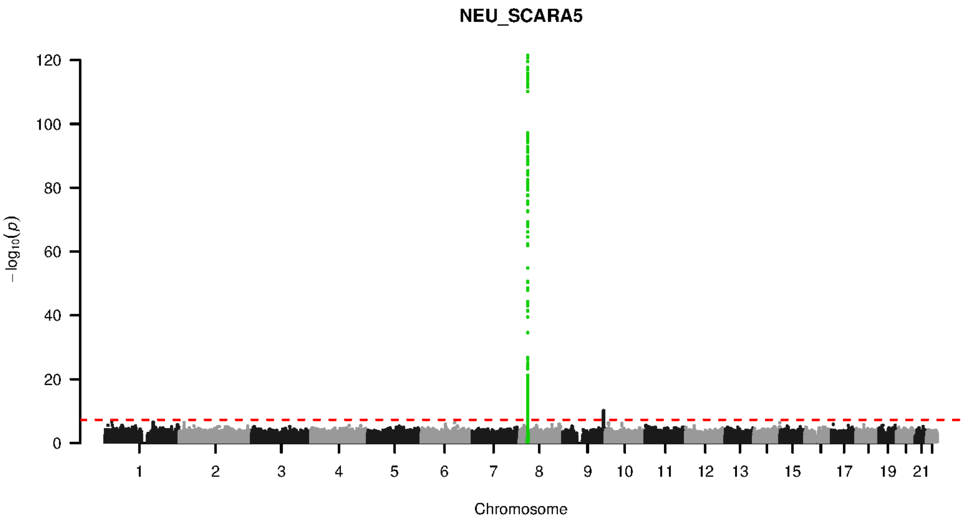


**Supplementary Figure 9-75. Manhattan plot and pQTL associations of the genome-wide meta-analysis for protein SCARA5.** The horizontal dashed line corresponds to the genome-wide significance threshold of $p = 5 \times 10^{-8}$. The variants in the cis-regulatory region (±500kb of the coding gene region) of the protein-coding gene are highlighted in green.

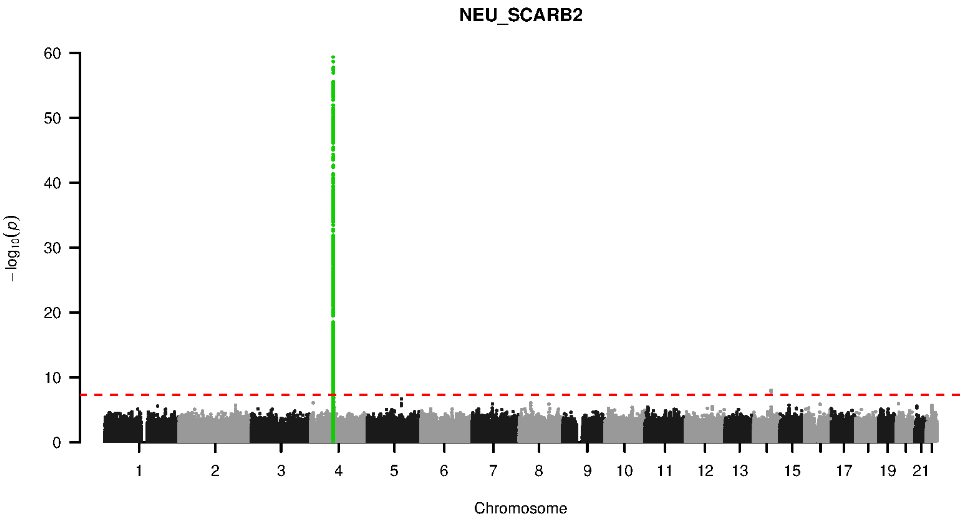


**Supplementary Figure 9-76. Manhattan plot and pQTL associations of the genome-wide meta-analysis for protein SCARB2.** The horizontal dashed line corresponds to the genome-wide significance threshold of $p = 5 \times 10^{-8}$. The variants in the cis-regulatory region (±500kb of the coding gene region) of the protein-coding gene are highlighted in green.

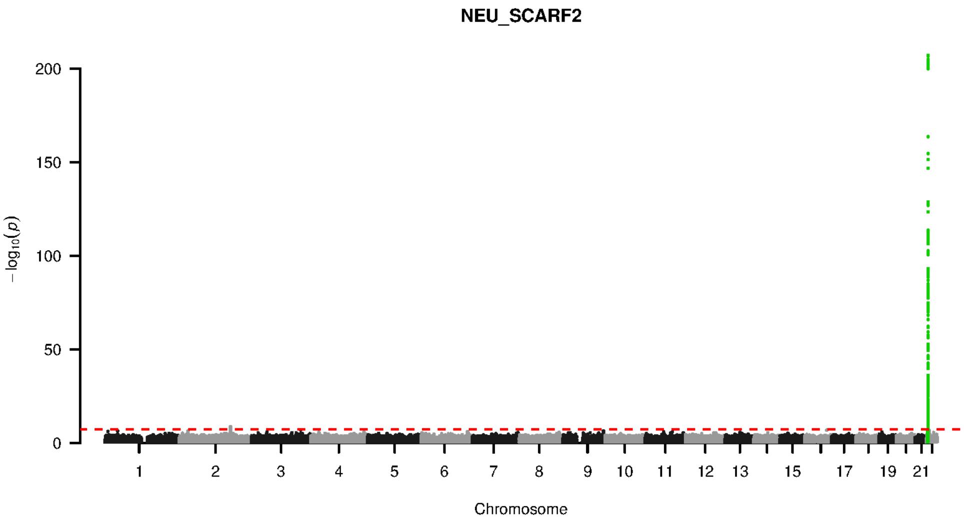


**Supplementary Figure 9-77. Manhattan plot and pQTL associations of the genome-wide meta-analysis for protein SCARF2.** The horizontal dashed line corresponds to the genome-wide significance threshold of $p = 5 \times 10^{-8}$. The variants in the cis-regulatory region (±500kb of the coding gene region) of the protein-coding gene are highlighted in green.

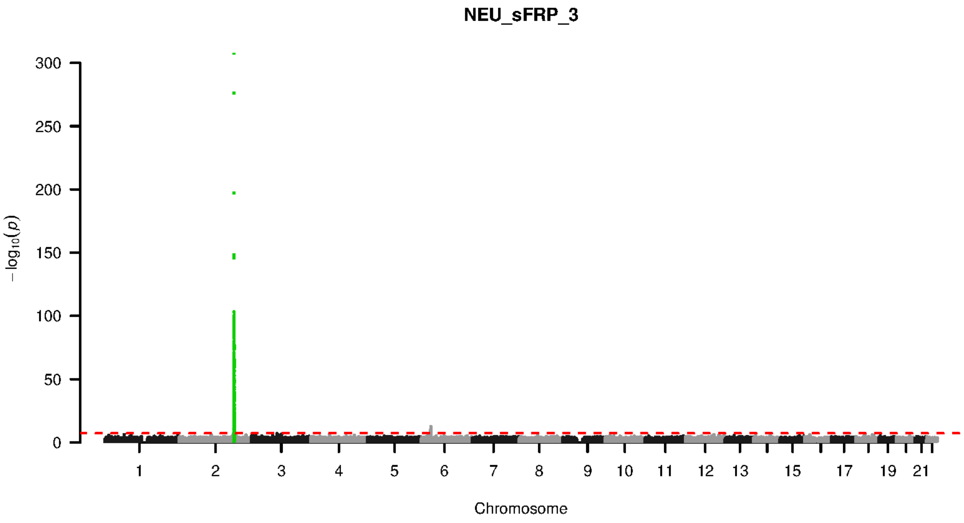


**Supplementary Figure 9-78. Manhattan plot and pQTL associations of the genome-wide meta-analysis for protein SFRP 3.** The horizontal dashed line corresponds to the genome-wide significance threshold of $p = 5 \times 10^{-8}$. The variants in the cis-regulatory region (±500kb of the coding gene region) of the protein-coding gene are highlighted in green.

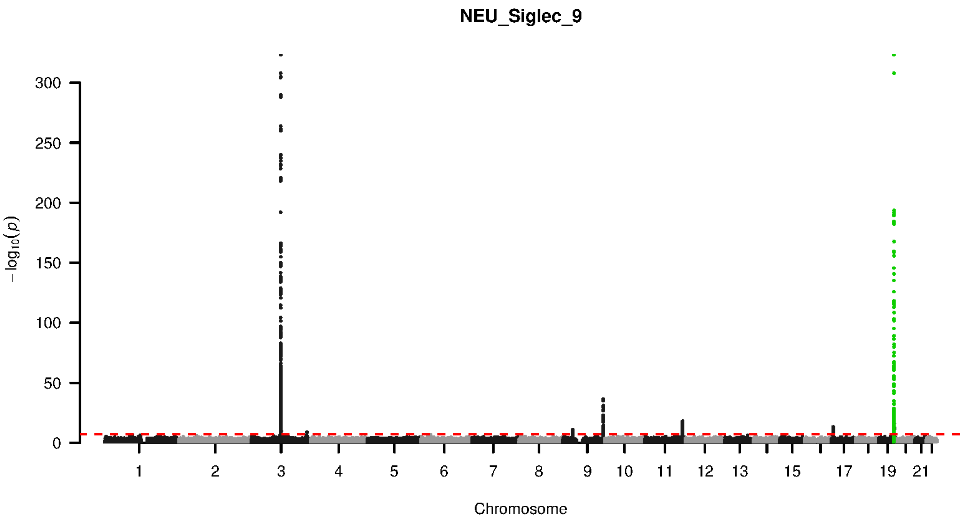


**Supplementary Figure 9-79. Manhattan plot and pQTL associations of the genome-wide meta-analysis for protein SIGLEC 9.** The horizontal dashed line corresponds to the genome-wide significance threshold of $p = 5 \times 10^{-8}$. The variants in the cis-regulatory region (±500kb of the coding gene region) of the protein-coding gene are highlighted in green.

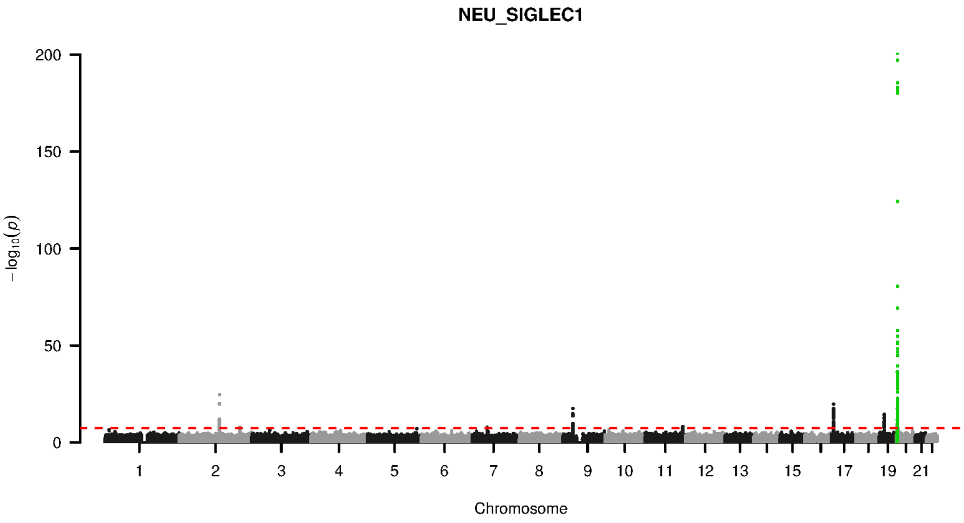


**Supplementary Figure 9-80. Manhattan plot and pQTL associations of the genome-wide meta-analysis for protein SIGLEC1.** The horizontal dashed line corresponds to the genome-wide significance threshold of $p = 5 \times 10^{-8}$. The variants in the cis-regulatory region (±500kb of the coding gene region) of the protein-coding gene are highlighted in green.

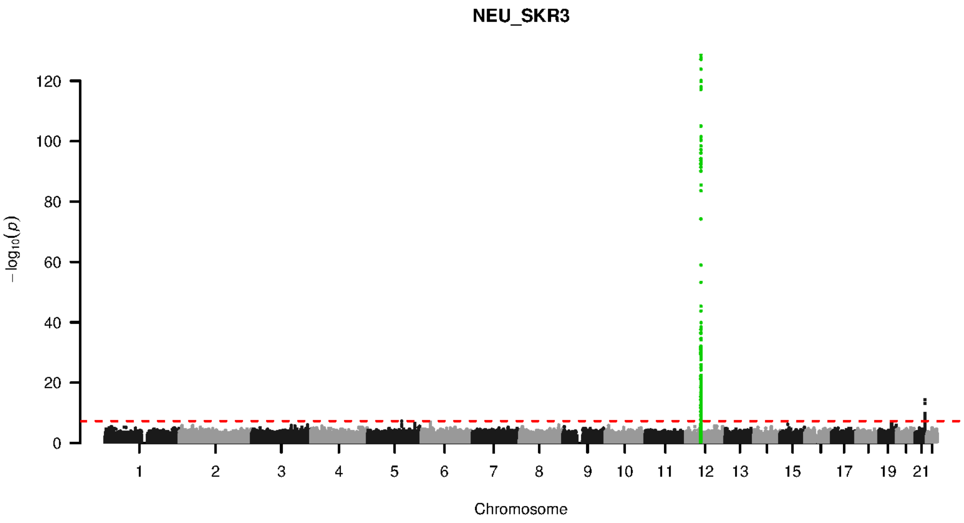


**Supplementary Figure 9-81. Manhattan plot and pQTL associations of the genome-wide meta-analysis for protein SKR3.** The horizontal dashed line corresponds to the genome-wide significance threshold of $p = 5 \times 10^{-8}$. The variants in the cis-regulatory region (±500kb of the coding gene region) of the protein-coding gene are highlighted in green.

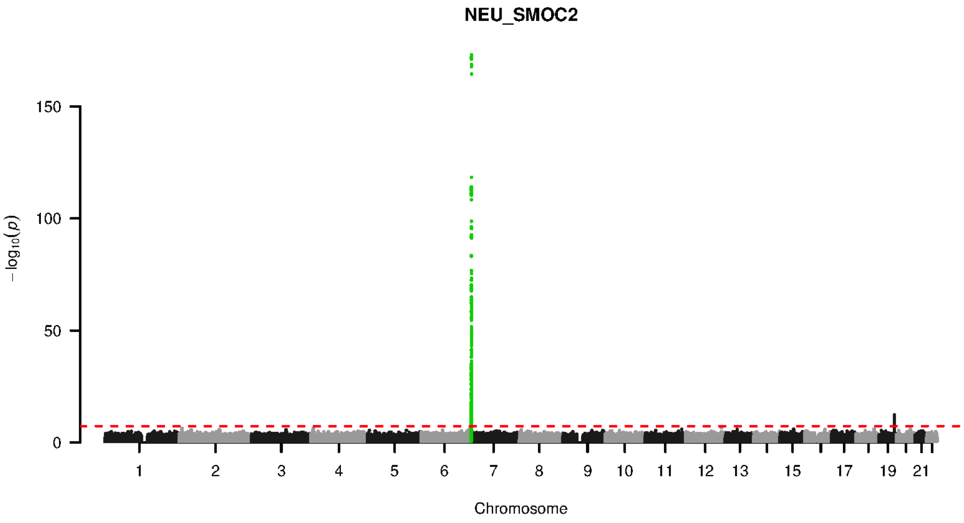


**Supplementary Figure 9-82. Manhattan plot and pQTL associations of the genome-wide meta-analysis for protein SMOC2.** The horizontal dashed line corresponds to the genome-wide significance threshold of $p = 5 \times 10^{-8}$. The variants in the cis-regulatory region (±500kb of the coding gene region) of the protein-coding gene are highlighted in green.

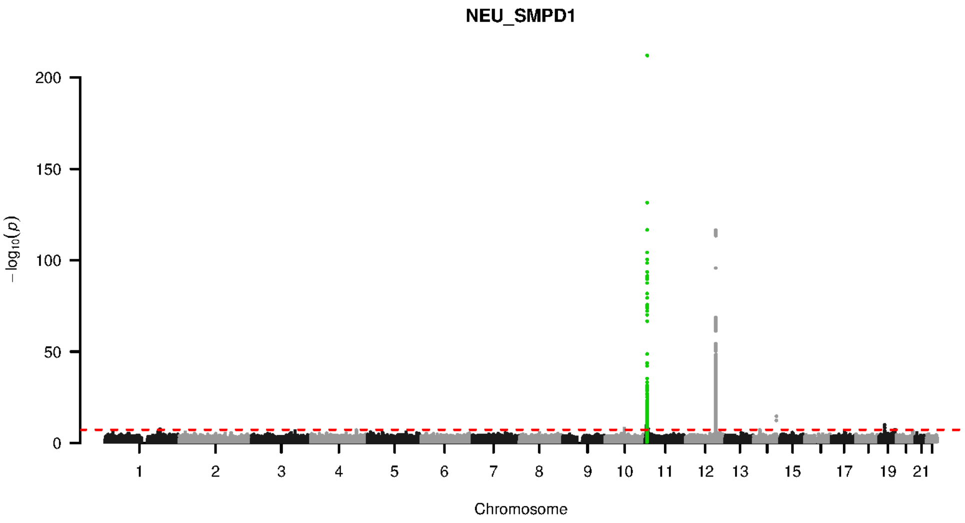


**Supplementary Figure 9-83. Manhattan plot and pQTL associations of the genome-wide meta-analysis for protein SMPD1.** The horizontal dashed line corresponds to the genome-wide significance threshold of $p = 5 \times 10^{-8}$. The variants in the cis-regulatory region (±500kb of the coding gene region) of the protein-coding gene are highlighted in green.

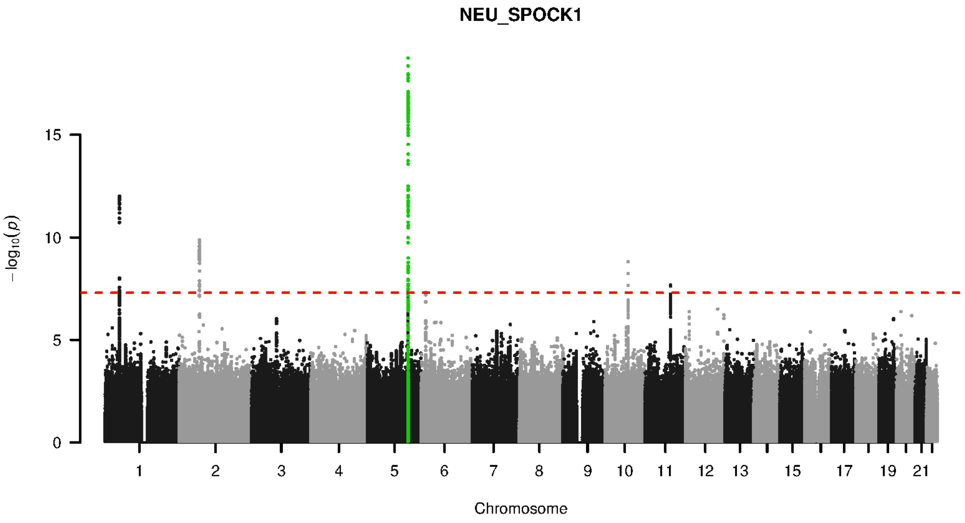


**Supplementary Figure 9-84. Manhattan plot and pQTL associations of the genome-wide meta-analysis for protein SPOCK1.** The horizontal dashed line corresponds to the genome-wide significance threshold of $p = 5 \times 10^{-8}$. The variants in the cis-regulatory region (±500kb of the coding gene region) of the protein-coding gene are highlighted in green.

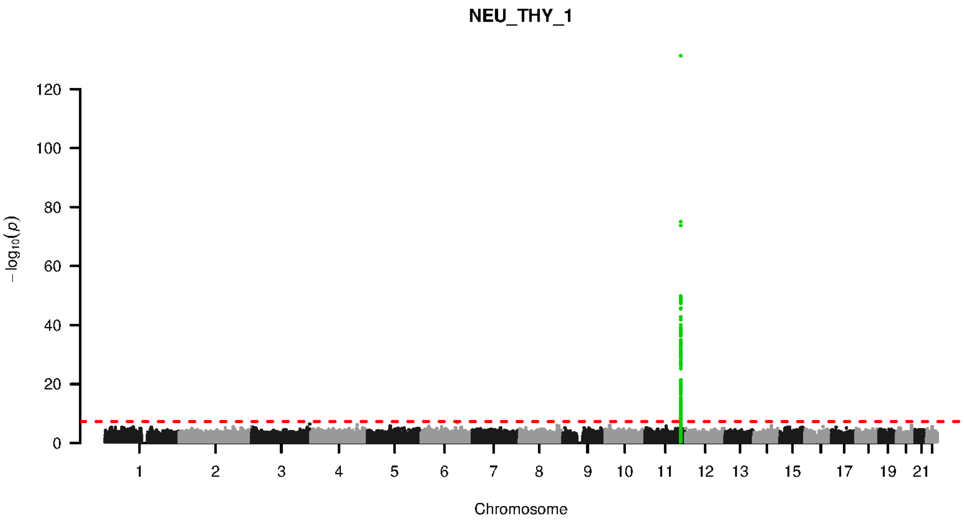


**Supplementary Figure 9-85. Manhattan plot and pQTL associations of the genome-wide meta-analysis for protein THY 1.** The horizontal dashed line corresponds to the genome-wide significance threshold of $p = 5 \times 10^{-8}$. The variants in the cis-regulatory region (±500kb of the coding gene region) of the protein-coding gene are highlighted in green.

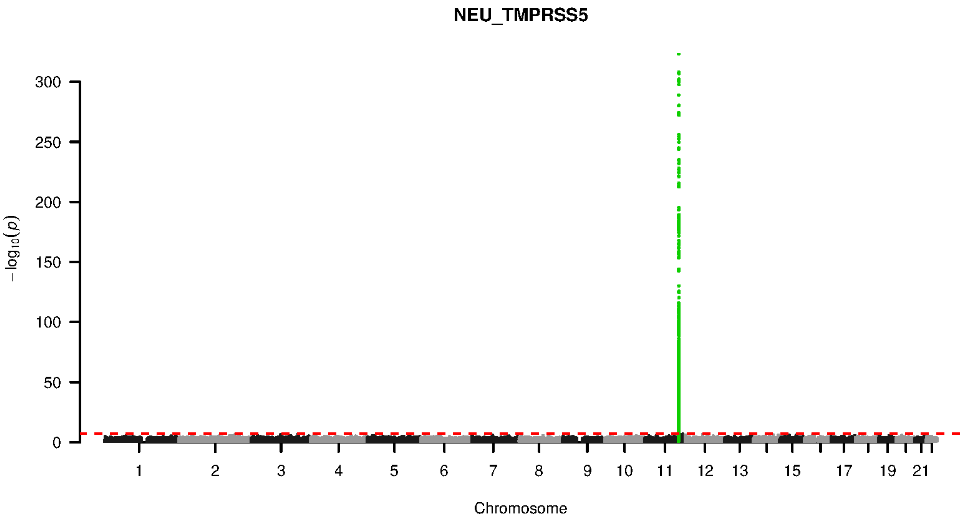


**Supplementary Figure 9-86. Manhattan plot and pQTL associations of the genome-wide meta-analysis for protein TMPRSS5.** The horizontal dashed line corresponds to the genome-wide significance threshold of $p = 5 \times 10^{-8}$. The variants in the cis-regulatory region (±500kb of the coding gene region) of the protein-coding gene are highlighted in green.

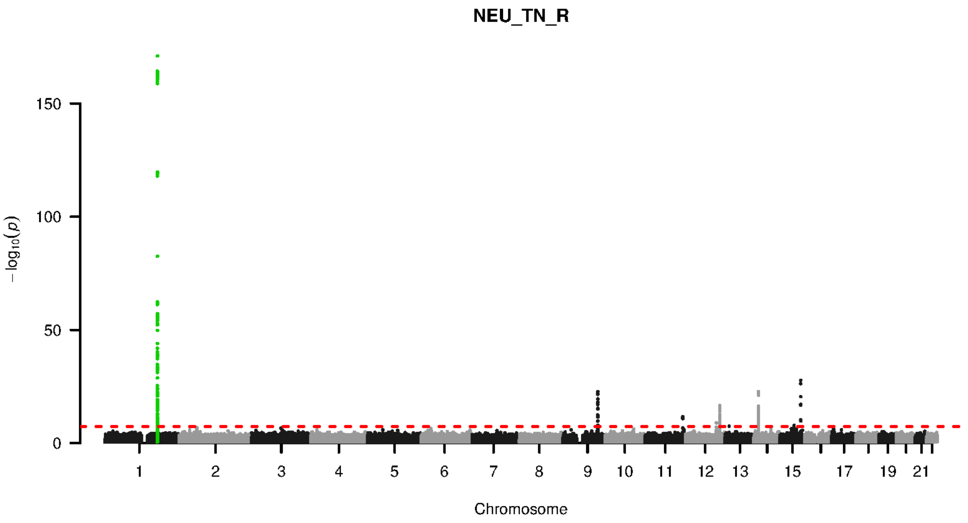


**Supplementary Figure 9-87. Manhattan plot and pQTL associations of the genome-wide meta-analysis for protein TN R.** The horizontal dashed line corresponds to the genome-wide significance threshold of $p = 5 \times 10^{-8}$. The variants in the cis-regulatory region (±500kb of the coding gene region) of the protein-coding gene are highlighted in green.

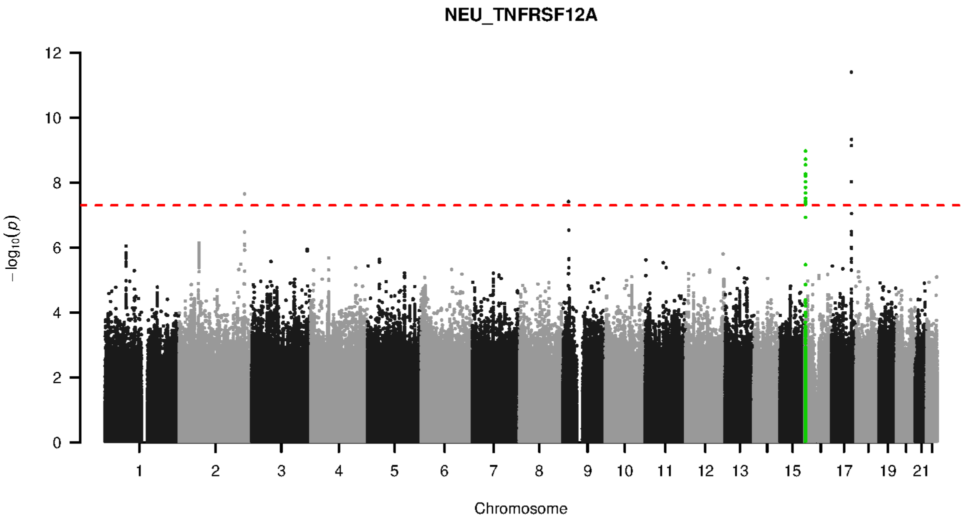


**Supplementary Figure 9-88. Manhattan plot and pQTL associations of the genome-wide meta-analysis for protein TNFRSF12A.** The horizontal dashed line corresponds to the genome-wide significance threshold of $p = 5 \times 10^{-8}$. The variants in the cis-regulatory region (±500kb of the coding gene region) of the protein-coding gene are highlighted in green.

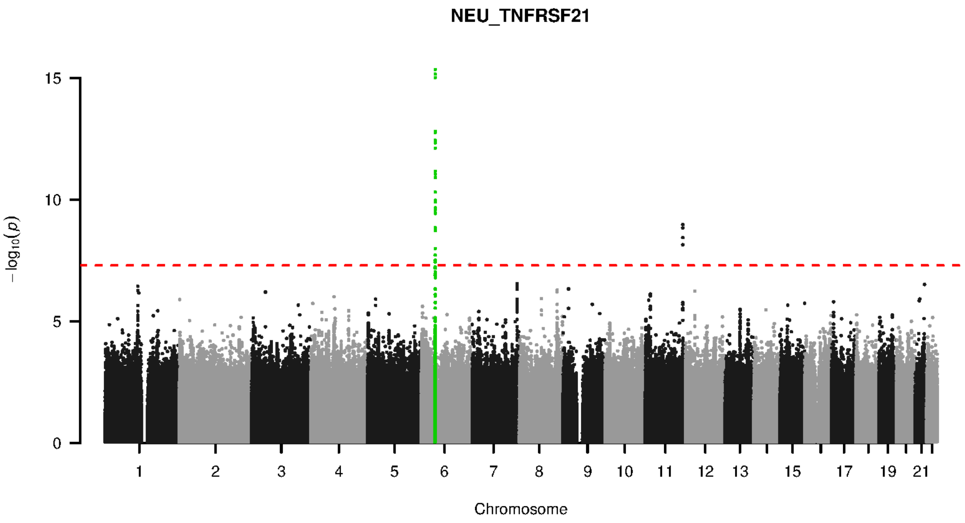


**Supplementary Figure 9-89. Manhattan plot and pQTL associations of the genome-wide meta-analysis for protein TNFRSF21.** The horizontal dashed line corresponds to the genome-wide significance threshold of $p = 5 \times 10^{-8}$. The variants in the cis-regulatory region (±500kb of the coding gene region) of the protein-coding gene are highlighted in green.

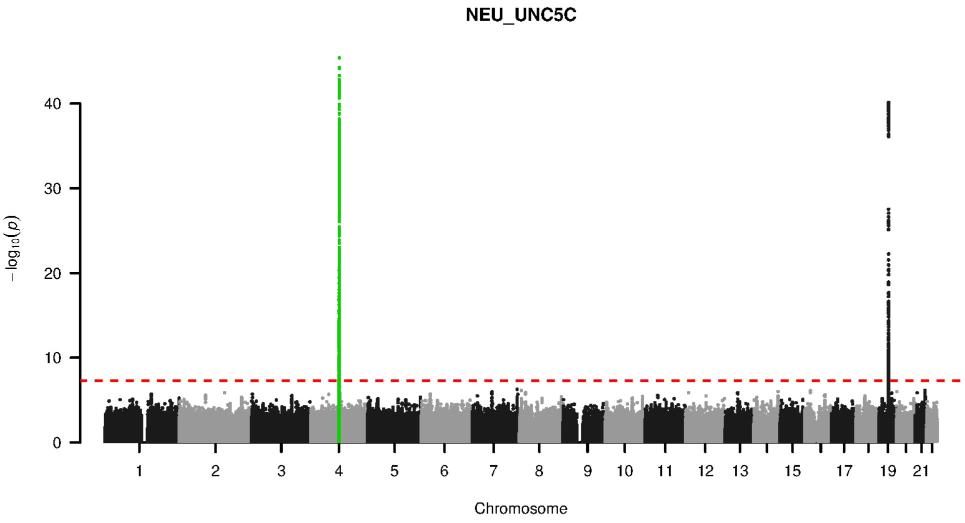


**Supplementary Figure 9-90. Manhattan plot and pQTL associations of the genome-wide meta-analysis for protein UNC5C.** The horizontal dashed line corresponds to the genome-wide significance threshold of $p = 5 \times 10^{-8}$. The variants in the cis-regulatory region (±500kb of the coding gene region) of the protein-coding gene are highlighted in green.

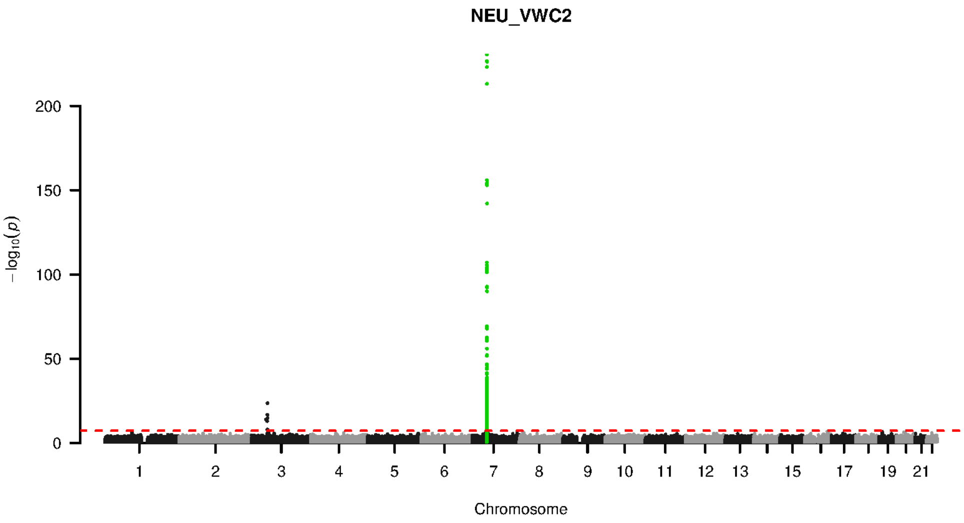


**Supplementary Figure 9-91. Manhattan plot and pQTL associations of the genome-wide meta-analysis for protein VWC2.** The horizontal dashed line corresponds to the genome-wide significance threshold of $p = 5 \times 10^{-8}$. The variants in the cis-regulatory region (±500kb of the coding gene region) of the protein-coding gene are highlighted in green.

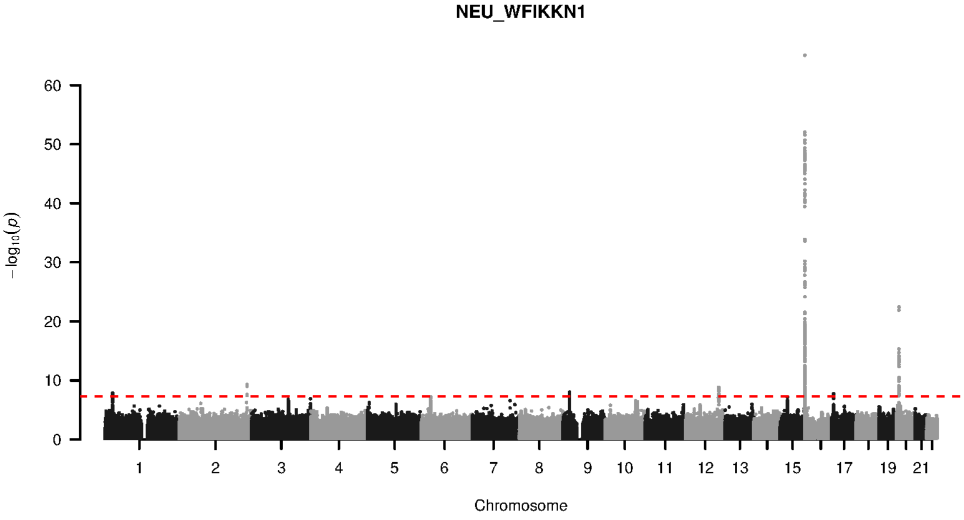


**Supplementary Figure 9-92. Manhattan plot and pQTL associations of the genome-wide meta-analysis for protein WFIKKN1.** The horizontal dashed line corresponds to the genome-wide significance threshold of $p = 5 \times 10^{-8}$. The variants in the cis-regulatory region (±500kb of the coding gene region) of the protein-coding gene are highlighted in green.

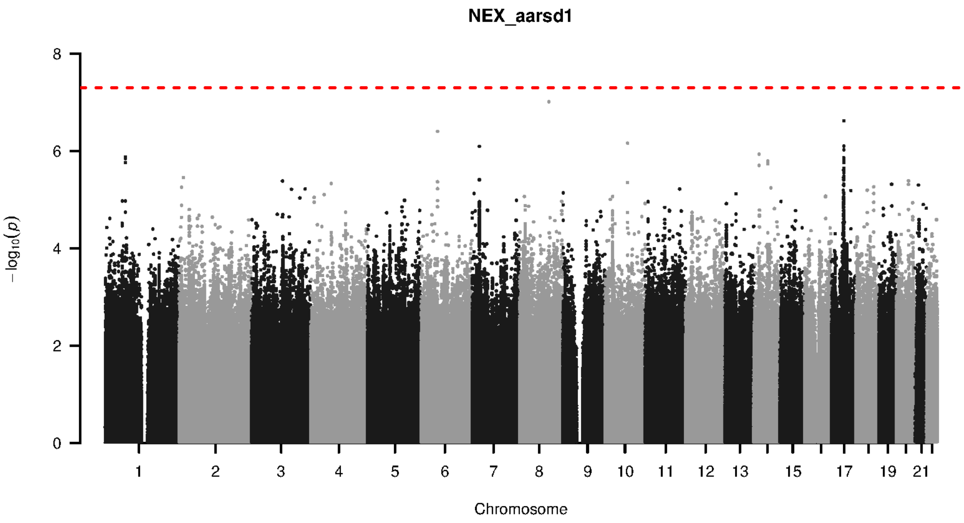


**Supplementary Figure 9-93. Manhattan plot and pQTL associations of the genome-wide meta-analysis for protein AARSD1.** The horizontal dashed line corresponds to the genome-wide significance threshold of $p = 5 \times 10^{-8}$. The variants in the cis-regulatory region (±500kb of the coding gene region) of the protein-coding gene are highlighted in green.

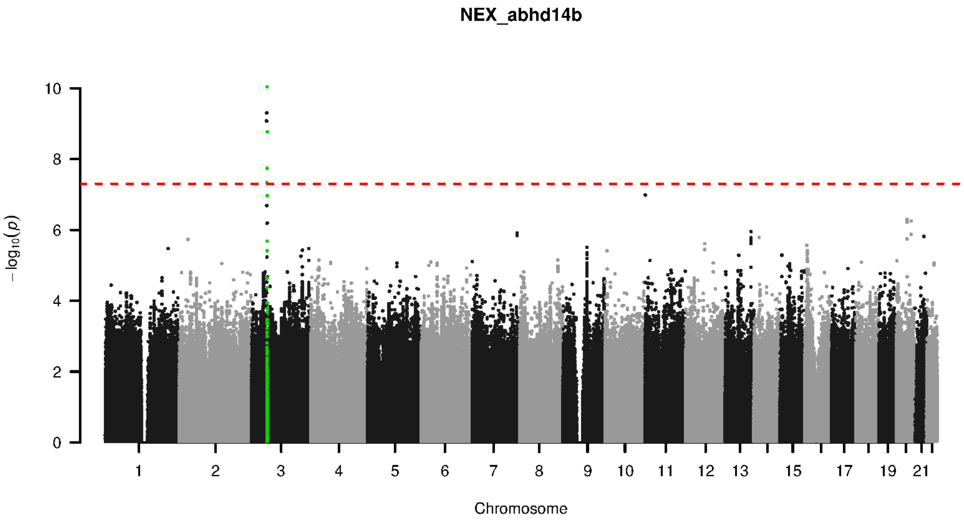


**Supplementary Figure 9-94. Manhattan plot and pQTL associations of the genome-wide meta-analysis for protein ABHD14B.** The horizontal dashed line corresponds to the genome-wide significance threshold of $p = 5 \times 10^{-8}$. The variants in the cis-regulatory region (±500kb of the coding gene region) of the protein-coding gene are highlighted in green.

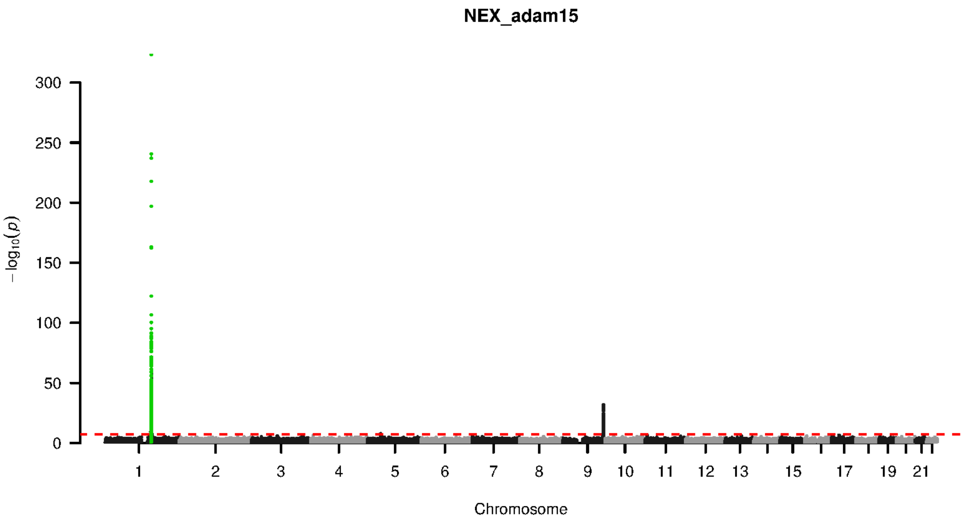


**Supplementary Figure 9-95. Manhattan plot and pQTL associations of the genome-wide meta-analysis for protein ADAM15.** The horizontal dashed line corresponds to the genome-wide significance threshold of $p = 5 \times 10^{-8}$. The variants in the cis-regulatory region (±500kb of the coding gene region) of the protein-coding gene are highlighted in green.

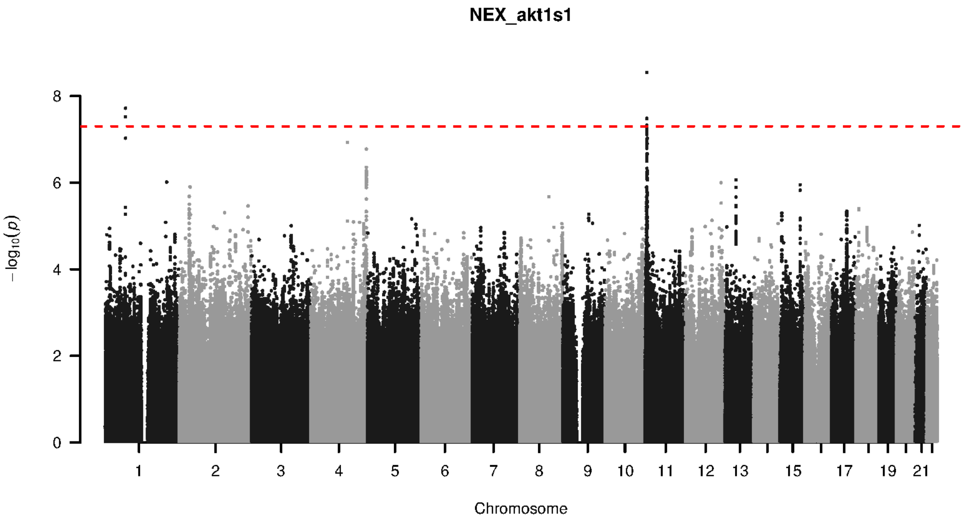


**Supplementary Figure 9-96. Manhattan plot and pQTL associations of the genome-wide meta-analysis for protein AKT1S1.** The horizontal dashed line corresponds to the genome-wide significance threshold of $p = 5 \times 10^{-8}$. The variants in the cis-regulatory region (±500kb of the coding gene region) of the protein-coding gene are highlighted in green.

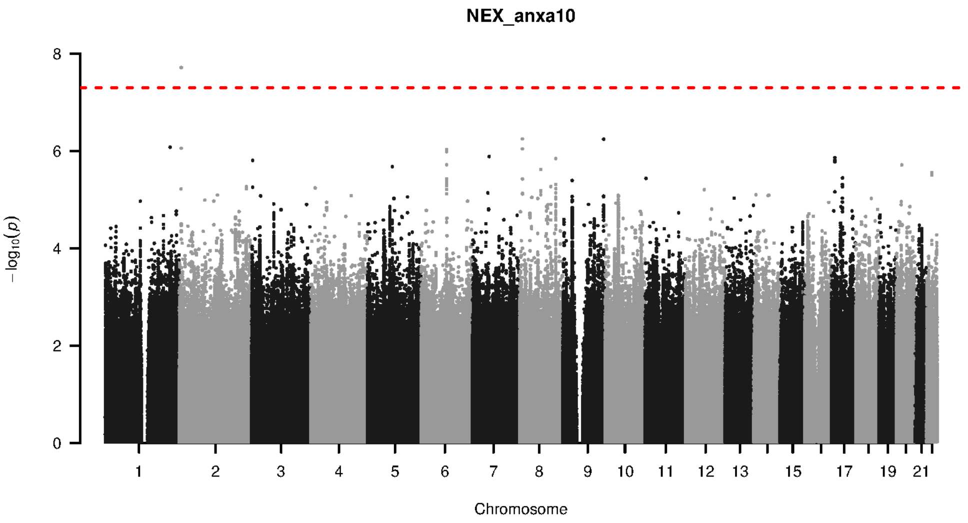


**Supplementary Figure 9-97. Manhattan plot and pQTL associations of the genome-wide meta-analysis for protein ANXA10.** The horizontal dashed line corresponds to the genome-wide significance threshold of $p = 5 \times 10^{-8}$. The variants in the cis-regulatory region (±500kb of the coding gene region) of the protein-coding gene are highlighted in green.

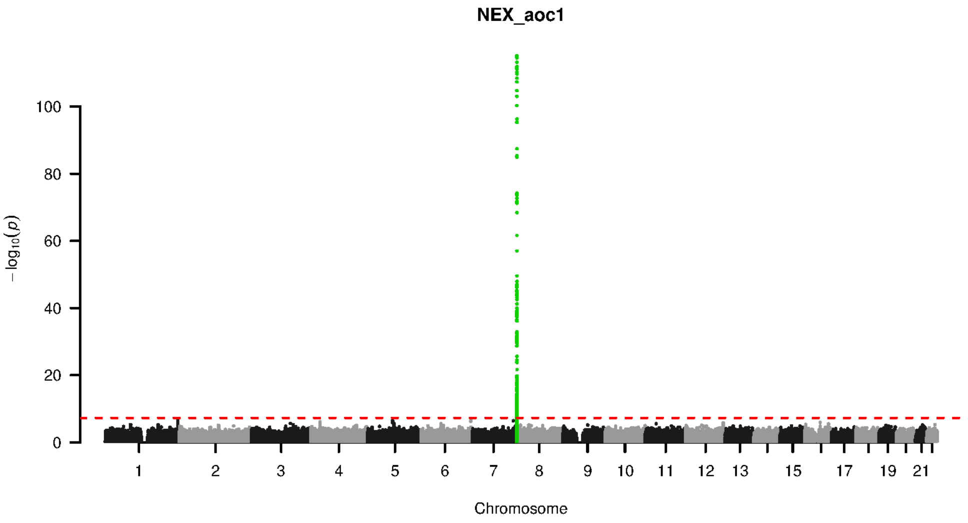


**Supplementary Figure 9-98. Manhattan plot and pQTL associations of the genome-wide meta-analysis for protein AOC1.** The horizontal dashed line corresponds to the genome-wide significance threshold of $p = 5 \times 10^{-8}$. The variants in the cis-regulatory region (±500kb of the coding gene region) of the protein-coding gene are highlighted in green.

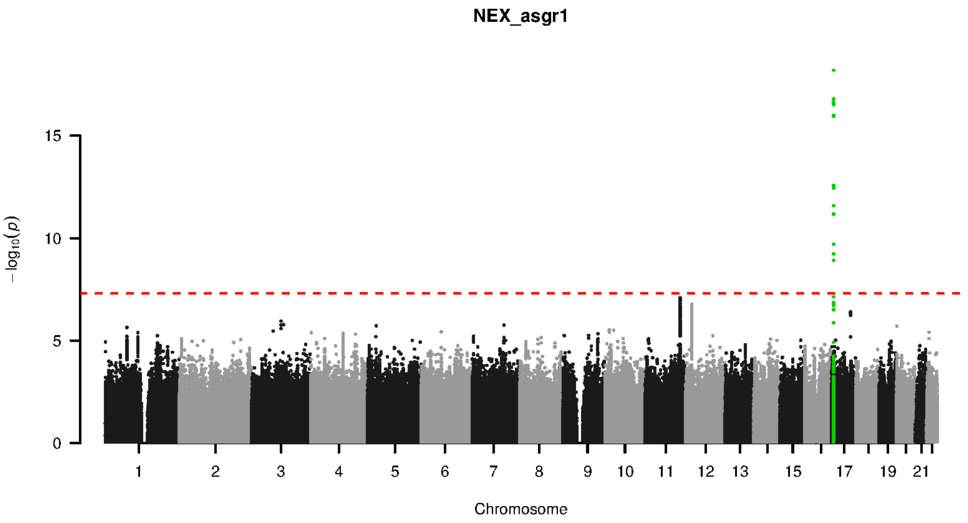


**Supplementary Figure 9-99. Manhattan plot and pQTL associations of the genome-wide meta-analysis for protein ASGR1.** The horizontal dashed line corresponds to the genome-wide significance threshold of $p = 5 \times 10^{-8}$. The variants in the cis-regulatory region (±500kb of the coding gene region) of the protein-coding gene are highlighted in green.

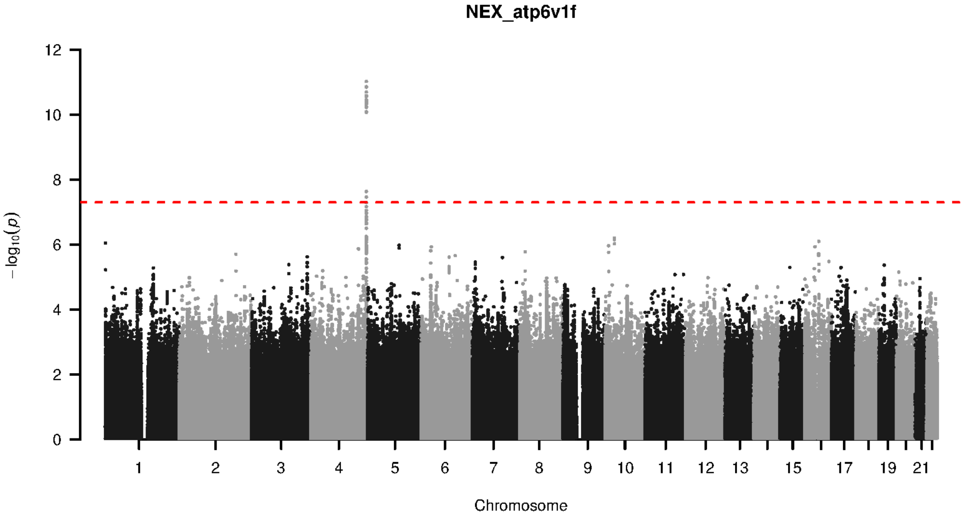


**Supplementary Figure 9-100. Manhattan plot and pQTL associations of the genome-wide meta-analysis for protein ATP6V1F.** The horizontal dashed line corresponds to the genome-wide significance threshold of $p = 5 \times 10^{-8}$. The variants in the cis-regulatory region (±500kb of the coding gene region) of the protein-coding gene are highlighted in green.

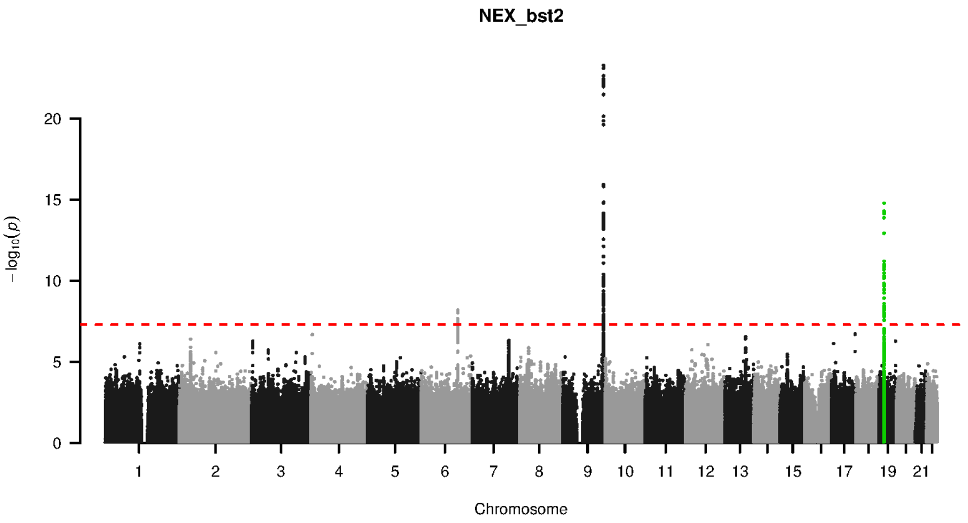


**Supplementary Figure 9-101. Manhattan plot and pQTL associations of the genome-wide meta-analysis for protein BST2.** The horizontal dashed line corresponds to the genome-wide significance threshold of $p = 5 \times 10^{-8}$. The variants in the cis-regulatory region (±500kb of the coding gene region) of the protein-coding gene are highlighted in green.

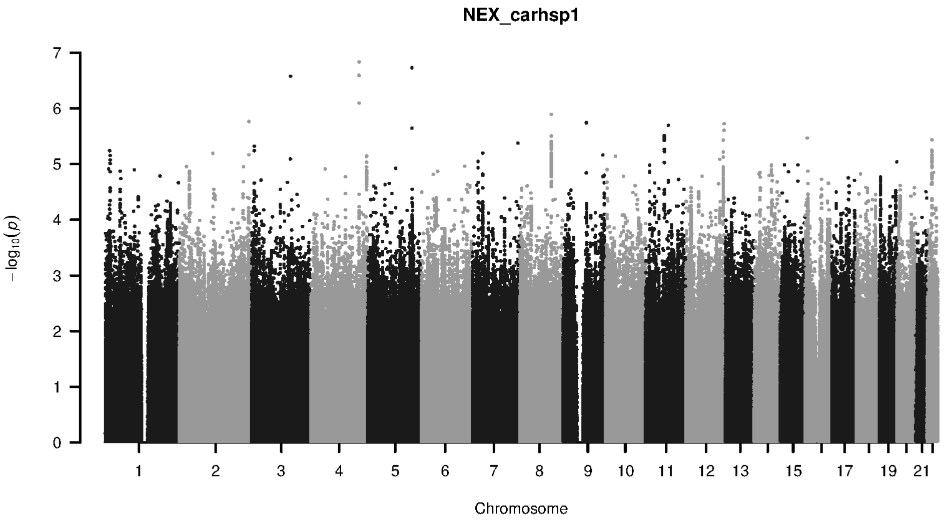


**Supplementary Figure 9-102. Manhattan plot and pQTL associations of the genome-wide meta-analysis for protein CARHSP1.** The horizontal dashed line corresponds to the genome-wide significance threshold of $p = 5 \times 10^{-8}$. The variants in the cis-regulatory region (±500kb of the coding gene region) of the protein-coding gene are highlighted in green.

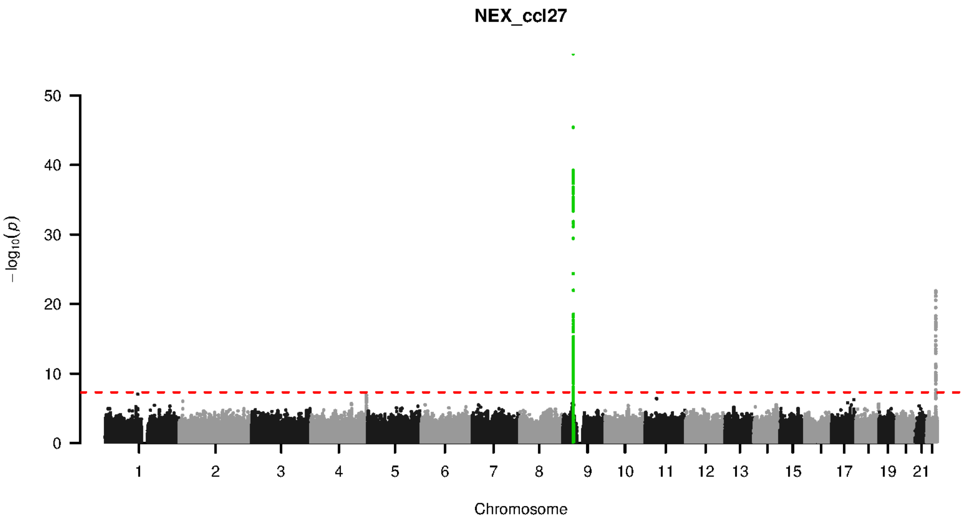


**Supplementary Figure 9-103. Manhattan plot and pQTL associations of the genome-wide meta-analysis for protein CCL27.** The horizontal dashed line corresponds to the genome-wide significance threshold of $p = 5 \times 10^{-8}$. The variants in the cis-regulatory region (±500kb of the coding gene region) of the protein-coding gene are highlighted in green.

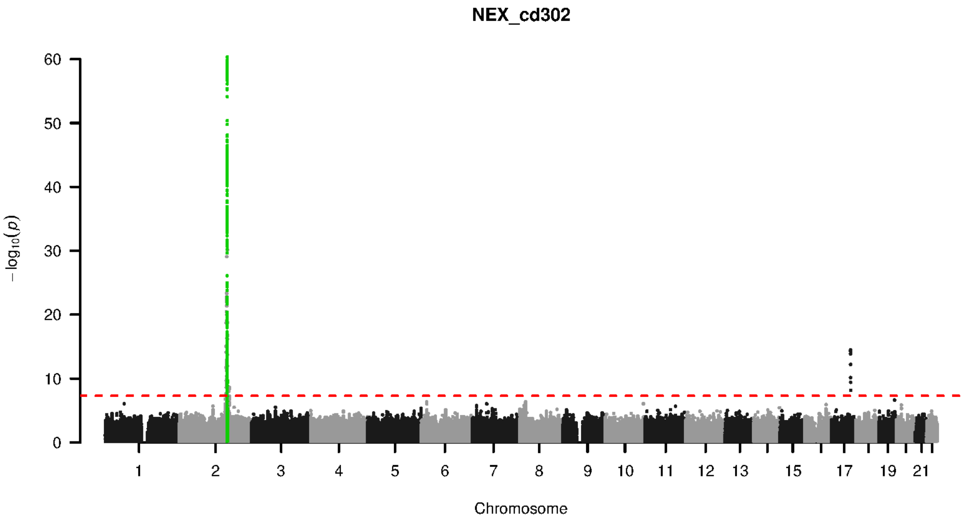


**Supplementary Figure 9-104. Manhattan plot and pQTL associations of the genome-wide meta-analysis for protein CD302.** The horizontal dashed line corresponds to the genome-wide significance threshold of $p = 5 \times 10^{-8}$. The variants in the cis-regulatory region (±500kb of the coding gene region) of the protein-coding gene are highlighted in green.

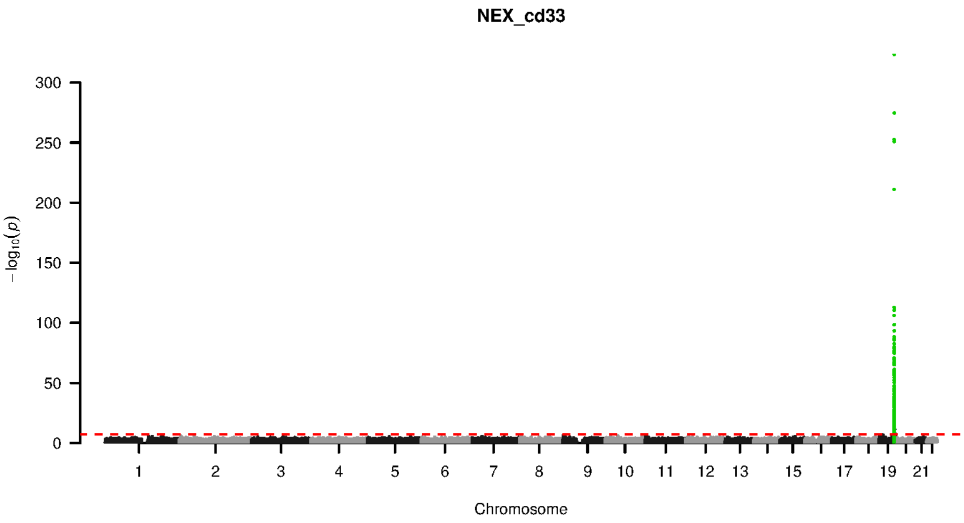


**Supplementary Figure 9-105. Manhattan plot and pQTL associations of the genome-wide meta-analysis for protein CD33.** The horizontal dashed line corresponds to the genome-wide significance threshold of $p = 5 \times 10^{-8}$. The variants in the cis-regulatory region (±500kb of the coding gene region) of the protein-coding gene are highlighted in green.

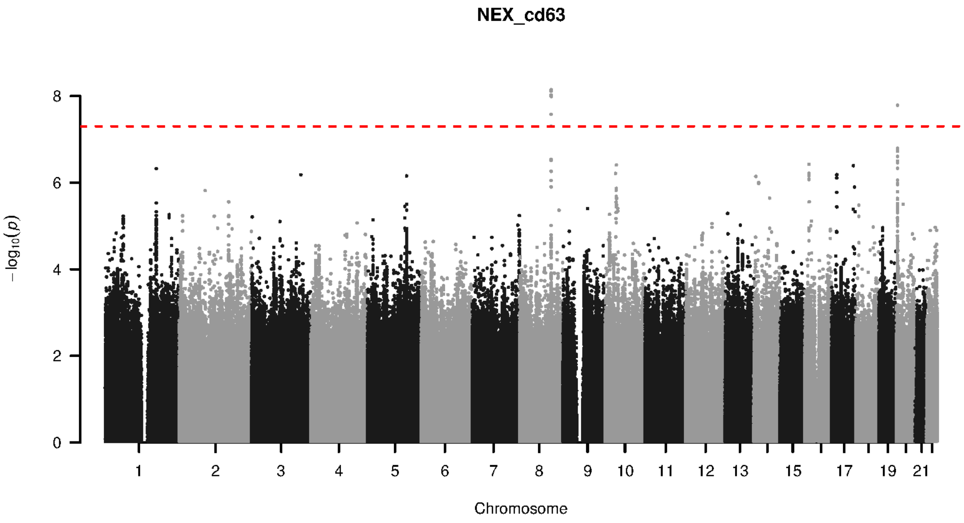


**Supplementary Figure 9-106. Manhattan plot and pQTL associations of the genome-wide meta-analysis for protein CD63.** The horizontal dashed line corresponds to the genome-wide significance threshold of $p = 5 \times 10^{-8}$. The variants in the cis-regulatory region (±500kb of the coding gene region) of the protein-coding gene are highlighted in green.

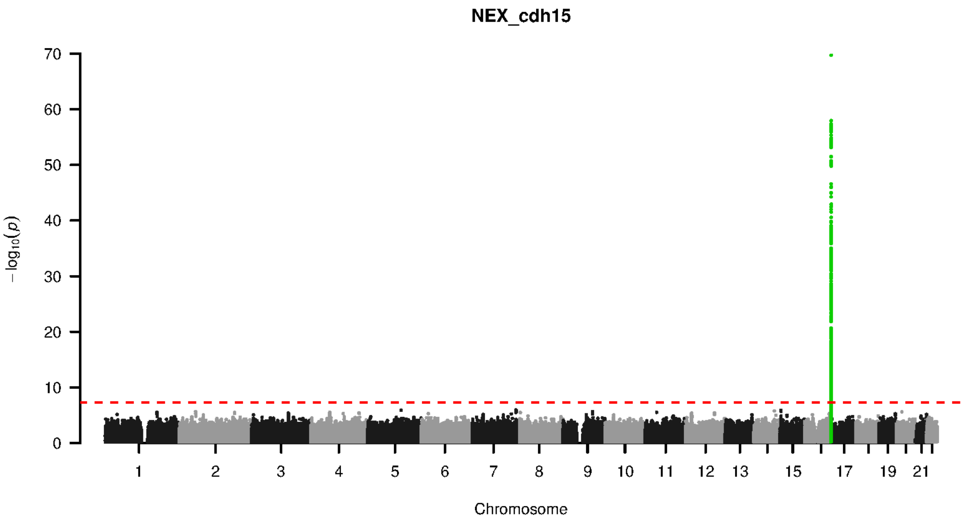


**Supplementary Figure 9-107. Manhattan plot and pQTL associations of the genome-wide meta-analysis for protein CDH15.** The horizontal dashed line corresponds to the genome-wide significance threshold of $p = 5 \times 10^{-8}$. The variants in the cis-regulatory region (±500kb of the coding gene region) of the protein-coding gene are highlighted in green.

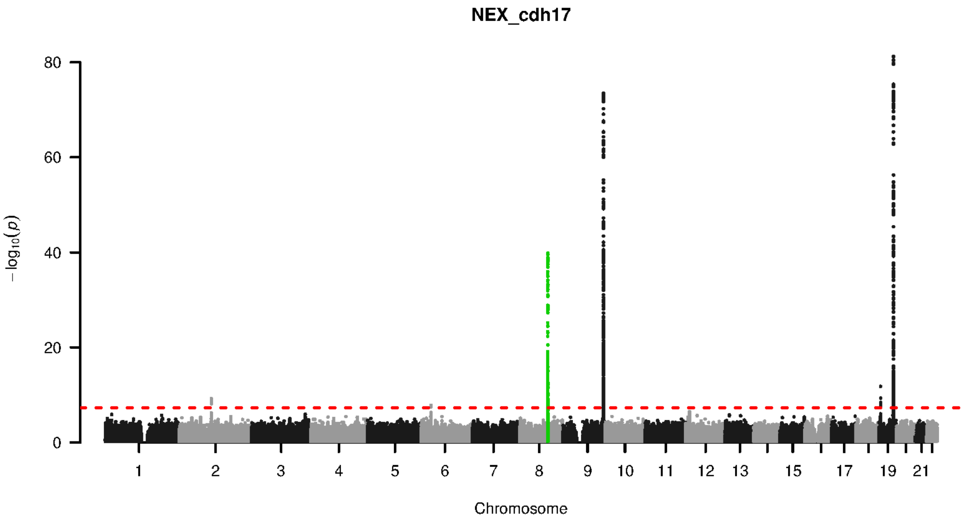


**Supplementary Figure 9-108. Manhattan plot and pQTL associations of the genome-wide meta-analysis for protein CDH17.** The horizontal dashed line corresponds to the genome-wide significance threshold of $p = 5 \times 10^{-8}$. The variants in the cis-regulatory region (±500kb of the coding gene region) of the protein-coding gene are highlighted in green.

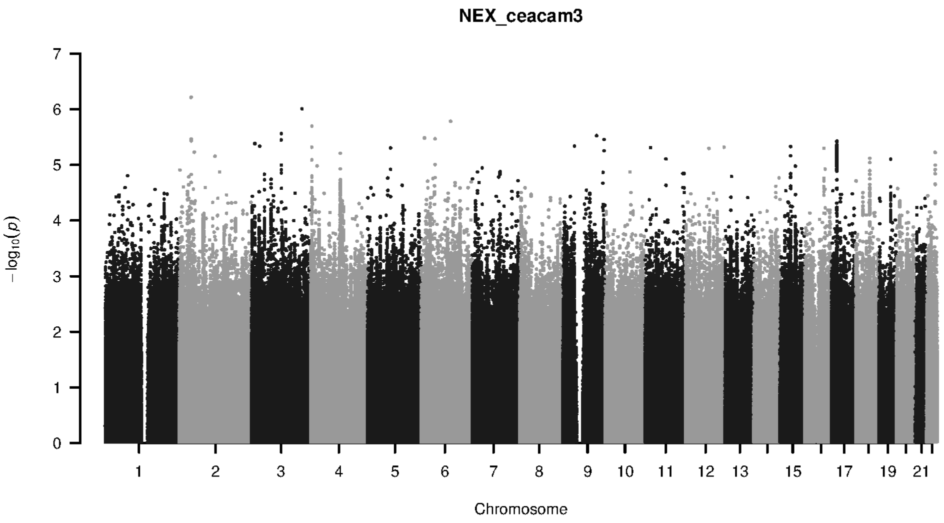


**Supplementary Figure 9-109. Manhattan plot and pQTL associations of the genome-wide meta-analysis for protein CEACAM3.** The horizontal dashed line corresponds to the genome-wide significance threshold of $p = 5 \times 10^{-8}$. The variants in the cis-regulatory region (±500kb of the coding gene region) of the protein-coding gene are highlighted in green.

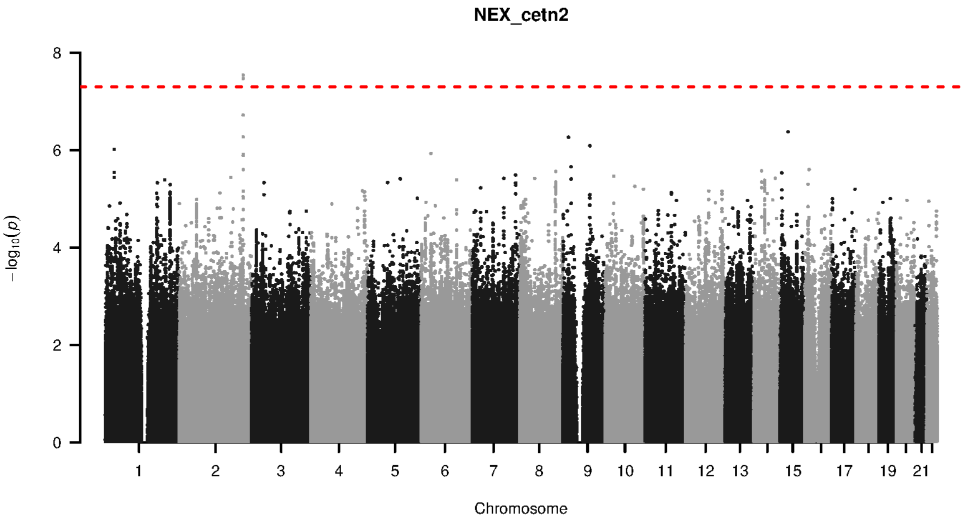


**Supplementary Figure 9-110. Manhattan plot and pQTL associations of the genome-wide meta-analysis for protein CETN2.** The horizontal dashed line corresponds to the genome-wide significance threshold of $p = 5 \times 10^{-8}$. The variants in the cis-regulatory region (±500kb of the coding gene region) of the protein-coding gene are highlighted in green.

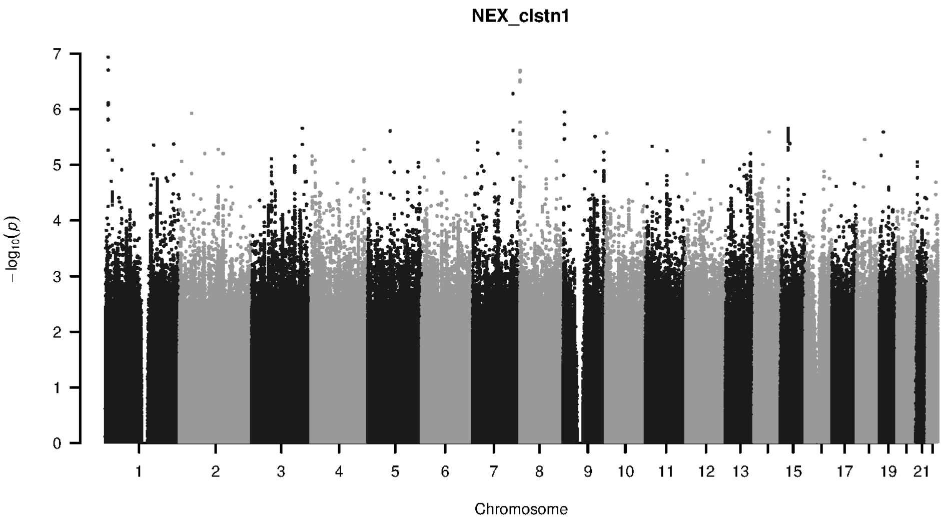


**Supplementary Figure 9-111. Manhattan plot and pQTL associations of the genome-wide meta-analysis for protein CLSTN1.** The horizontal dashed line corresponds to the genome-wide significance threshold of $p = 5 \times 10^{-8}$. The variants in the cis-regulatory region ($\pm$500kb of the coding gene region) of the protein-coding gene are highlighted in green.

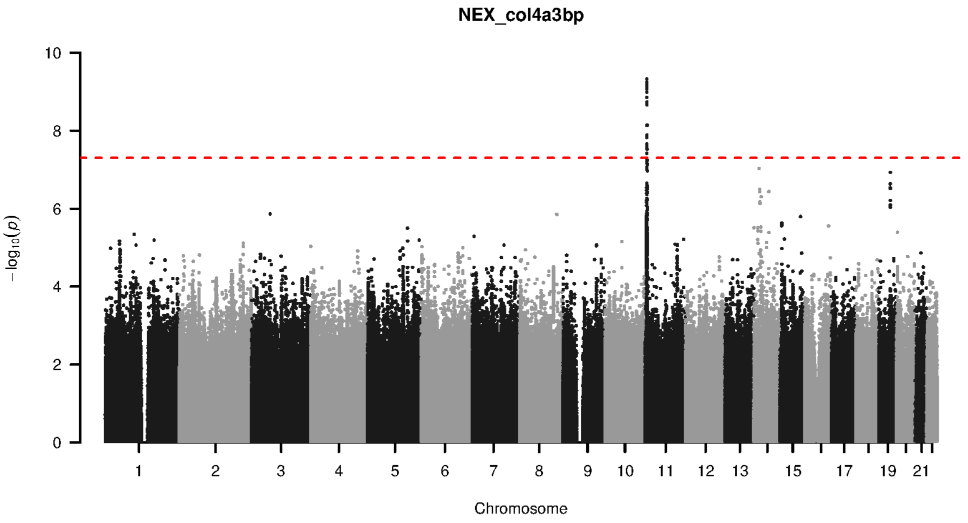


**Supplementary Figure 9-112. Manhattan plot and pQTL associations of the genome-wide meta-analysis for protein COL4A3BP.** The horizontal dashed line corresponds to the genome-wide significance threshold of $p = 5 \times 10^{-8}$. The variants in the cis-regulatory region ($\pm$500kb of the coding gene region) of the protein-coding gene are highlighted in green.

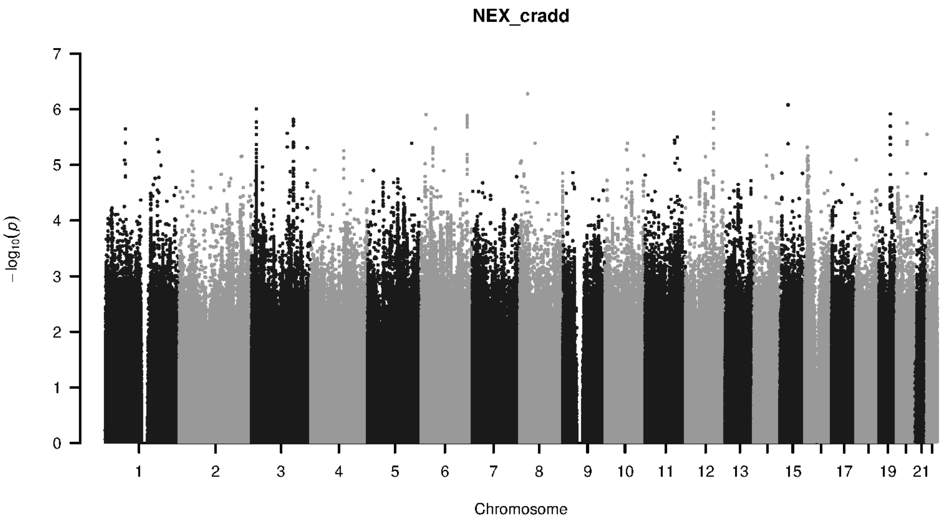


**Supplementary Figure 9-113. Manhattan plot and pQTL associations of the genome-wide meta-analysis for protein CRADD.** The horizontal dashed line corresponds to the genome-wide significance threshold of $p = 5 \times 10^{-8}$. The variants in the cis-regulatory region (±500kb of the coding gene region) of the protein-coding gene are highlighted in green.

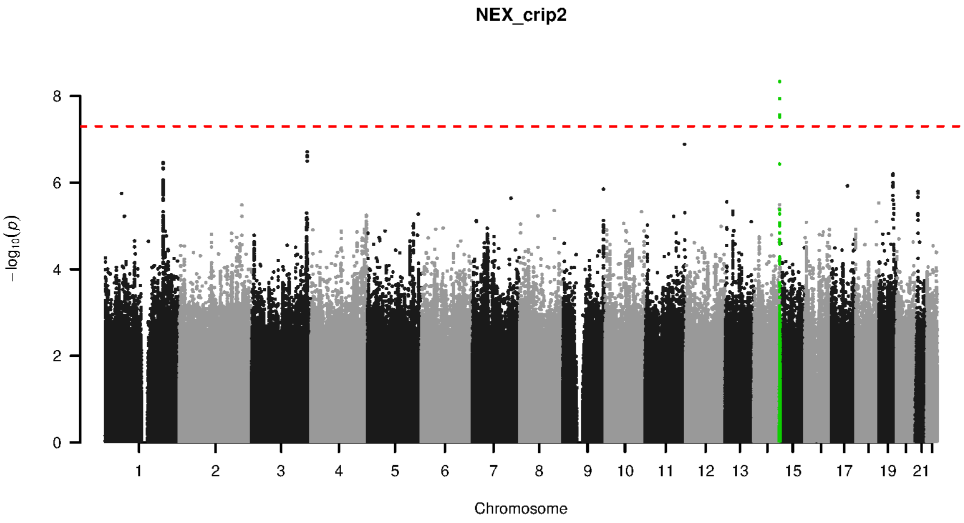


**Supplementary Figure 9-114. Manhattan plot and pQTL associations of the genome-wide meta-analysis for protein CRIP2.** The horizontal dashed line corresponds to the genome-wide significance threshold of $p = 5 \times 10^{-8}$. The variants in the cis-regulatory region (±500kb of the coding gene region) of the protein-coding gene are highlighted in green.

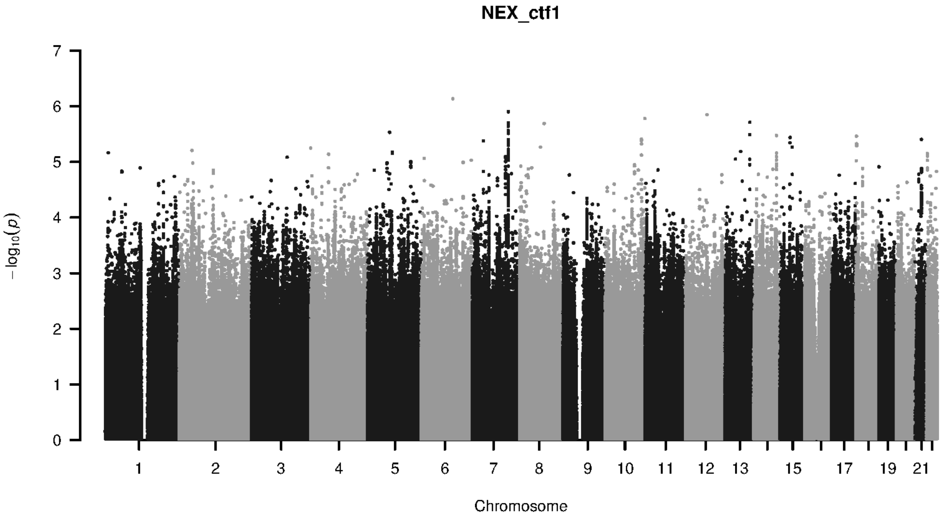


**Supplementary Figure 9-115. Manhattan plot and pQTL associations of the genome-wide meta-analysis for protein CTF1.** The horizontal dashed line corresponds to the genome-wide significance threshold of $p = 5 \times 10^{-8}$. The variants in the cis-regulatory region (±500kb of the coding gene region) of the protein-coding gene are highlighted in green.

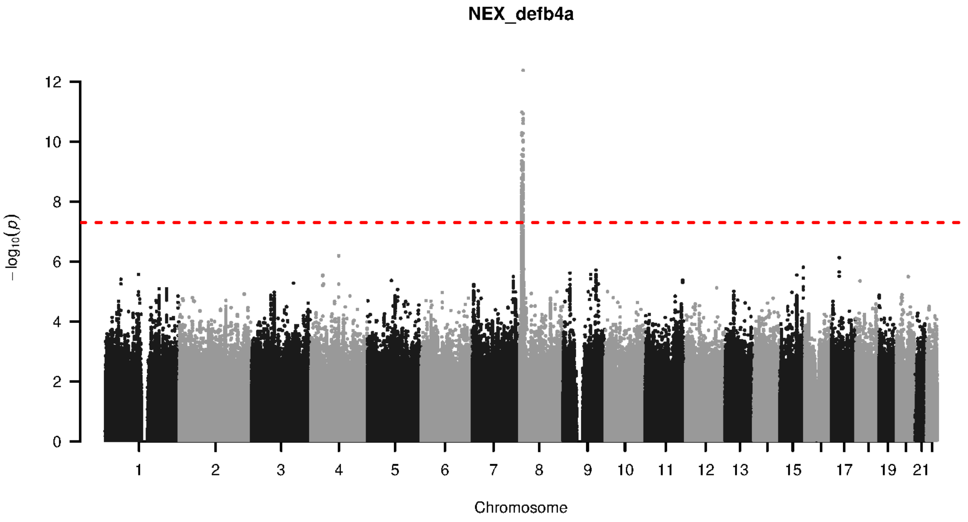


**Supplementary Figure 9-116. Manhattan plot and pQTL associations of the genome-wide meta-analysis for protein DEFB4A.** The horizontal dashed line corresponds to the genome-wide significance threshold of $p = 5 \times 10^{-8}$. The variants in the cis-regulatory region (±500kb of the coding gene region) of the protein-coding gene are highlighted in green.

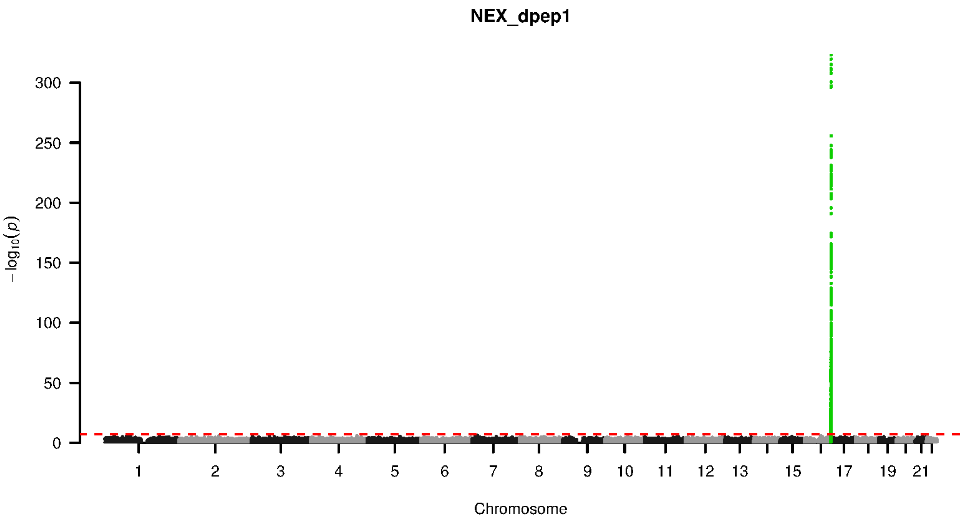


**Supplementary Figure 9-117. Manhattan plot and pQTL associations of the genome-wide meta-analysis for protein DPEP1.** The horizontal dashed line corresponds to the genome-wide significance threshold of $p = 5 \times 10^{-8}$. The variants in the cis-regulatory region (±500kb of the coding gene region) of the protein-coding gene are highlighted in green.

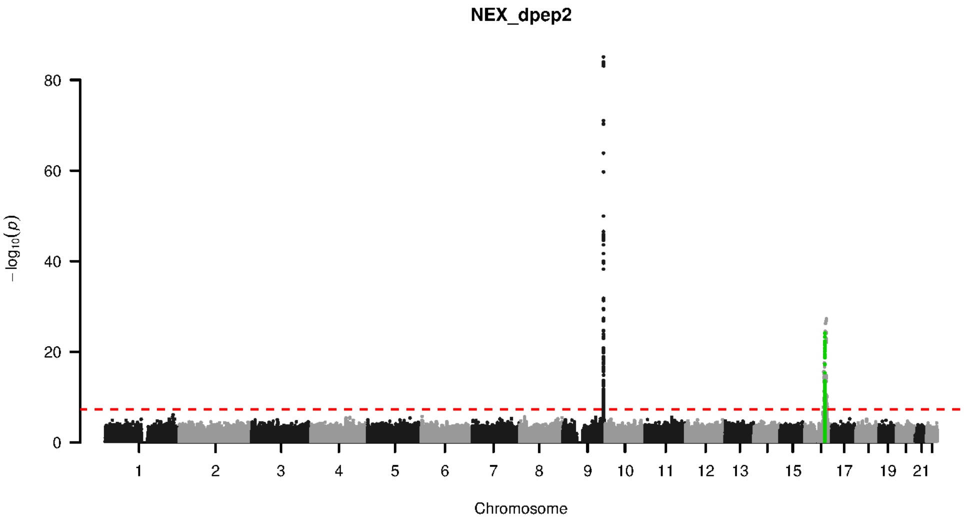


**Supplementary Figure 9-118. Manhattan plot and pQTL associations of the genome-wide meta-analysis for protein DPEP2.** The horizontal dashed line corresponds to the genome-wide significance threshold of $p = 5 \times 10^{-8}$. The variants in the cis-regulatory region (±500kb of the coding gene region) of the protein-coding gene are highlighted in green.

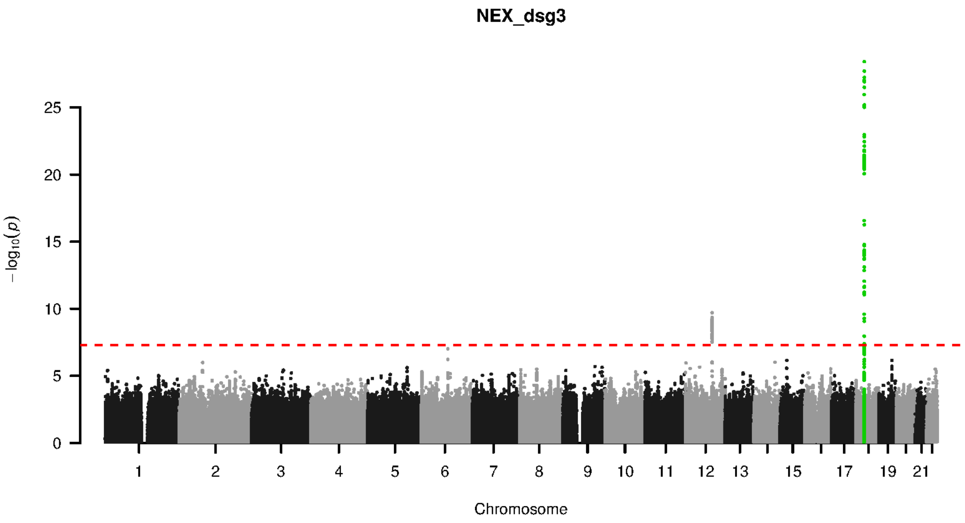


**Supplementary Figure 9-119. Manhattan plot and pQTL associations of the genome-wide meta-analysis for protein DSG3.** The horizontal dashed line corresponds to the genome-wide significance threshold of $p = 5 \times 10^{-8}$. The variants in the cis-regulatory region (±500kb of the coding gene region) of the protein-coding gene are highlighted in green.

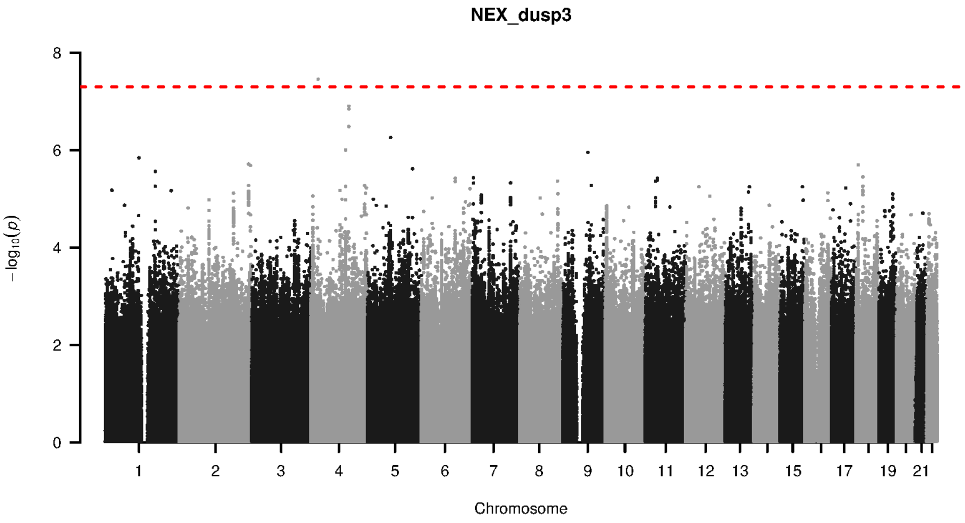


**Supplementary Figure 9-120. Manhattan plot and pQTL associations of the genome-wide meta-analysis for protein DUSP3.** The horizontal dashed line corresponds to the genome-wide significance threshold of $p = 5 \times 10^{-8}$. The variants in the cis-regulatory region (±500kb of the coding gene region) of the protein-coding gene are highlighted in green.

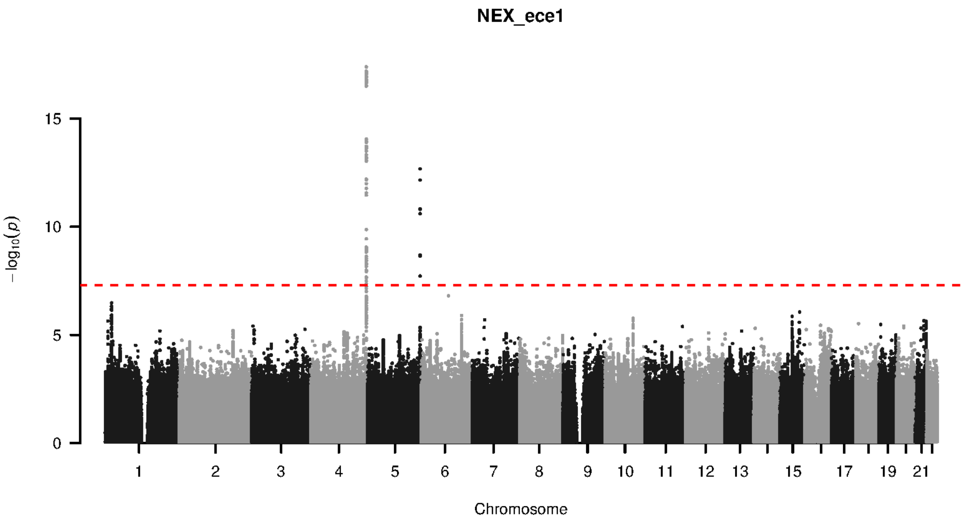


**Supplementary Figure 9-121. Manhattan plot and pQTL associations of the genome-wide meta-analysis for protein ECE1.** The horizontal dashed line corresponds to the genome-wide significance threshold of $p = 5 \times 10^{-8}$. The variants in the cis-regulatory region (±500kb of the coding gene region) of the protein-coding gene are highlighted in green.

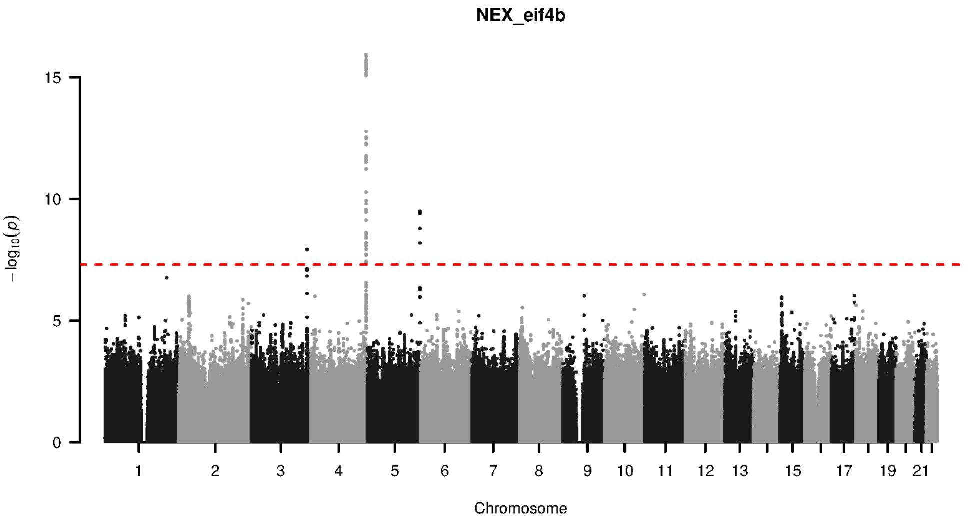


**Supplementary Figure 9-122. Manhattan plot and pQTL associations of the genome-wide meta-analysis for protein EIF4B.** The horizontal dashed line corresponds to the genome-wide significance threshold of $p = 5 \times 10^{-8}$. The variants in the cis-regulatory region (±500kb of the coding gene region) of the protein-coding gene are highlighted in green.

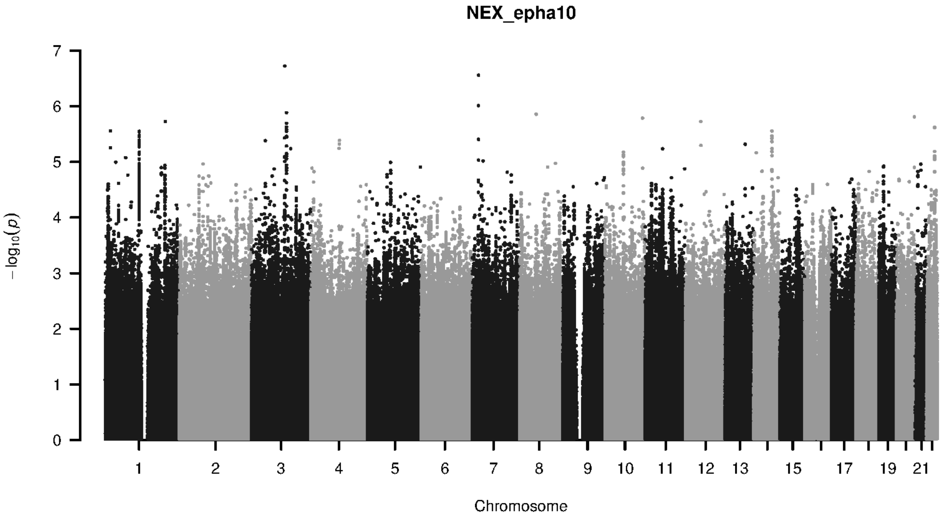


**Supplementary Figure 9-123. Manhattan plot and pQTL associations of the genome-wide meta-analysis for protein EPHA10.** The horizontal dashed line corresponds to the genome-wide significance threshold of $p = 5 \times 10^{-8}$. The variants in the cis-regulatory region ($\pm$500kb of the coding gene region) of the protein-coding gene are highlighted in green.

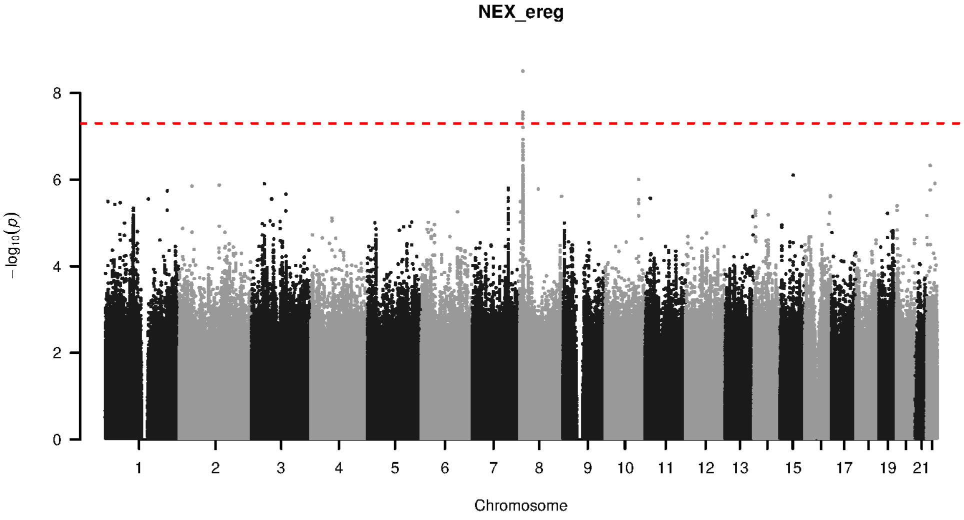


**Supplementary Figure 9-124. Manhattan plot and pQTL associations of the genome-wide meta-analysis for protein EREG.** The horizontal dashed line corresponds to the genome-wide significance threshold of $p = 5 \times 10^{-8}$. The variants in the cis-regulatory region ($\pm$500kb of the coding gene region) of the protein-coding gene are highlighted in green.

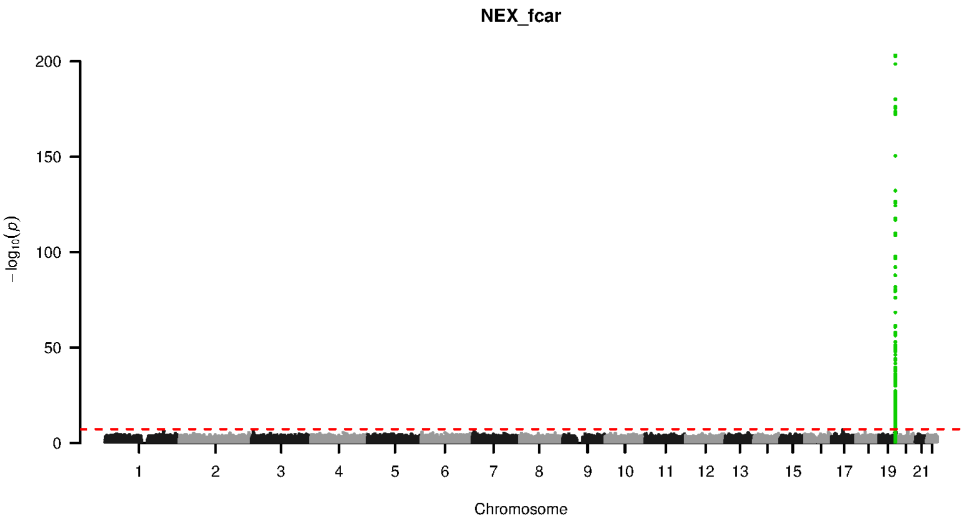


**Supplementary Figure 9-125. Manhattan plot and pQTL associations of the genome-wide meta-analysis for protein FCAR.** The horizontal dashed line corresponds to the genome-wide significance threshold of $p = 5 \times 10^{-8}$. The variants in the cis-regulatory region (±500kb of the coding gene region) of the protein-coding gene are highlighted in green.

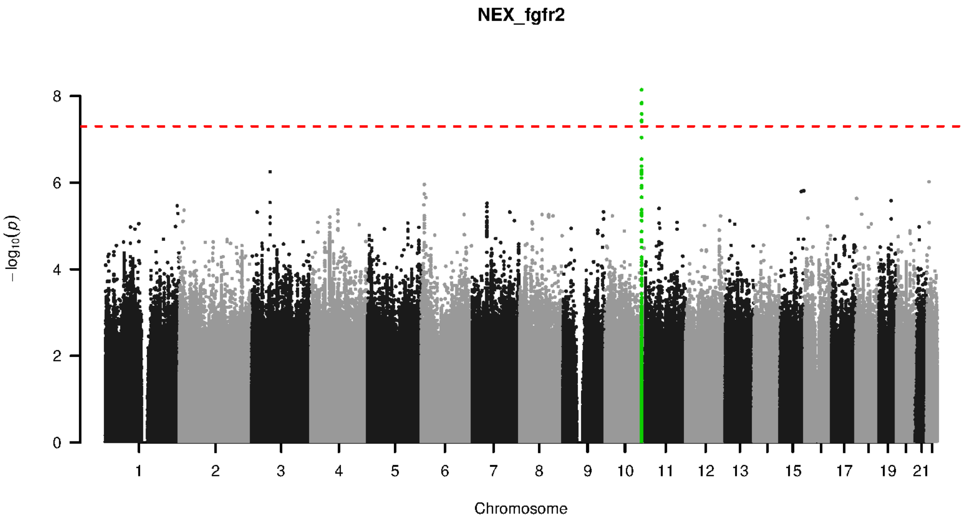


**Supplementary Figure 9-126. Manhattan plot and pQTL associations of the genome-wide meta-analysis for protein FGFR2.** The horizontal dashed line corresponds to the genome-wide significance threshold of $p = 5 \times 10^{-8}$. The variants in the cis-regulatory region (±500kb of the coding gene region) of the protein-coding gene are highlighted in green.

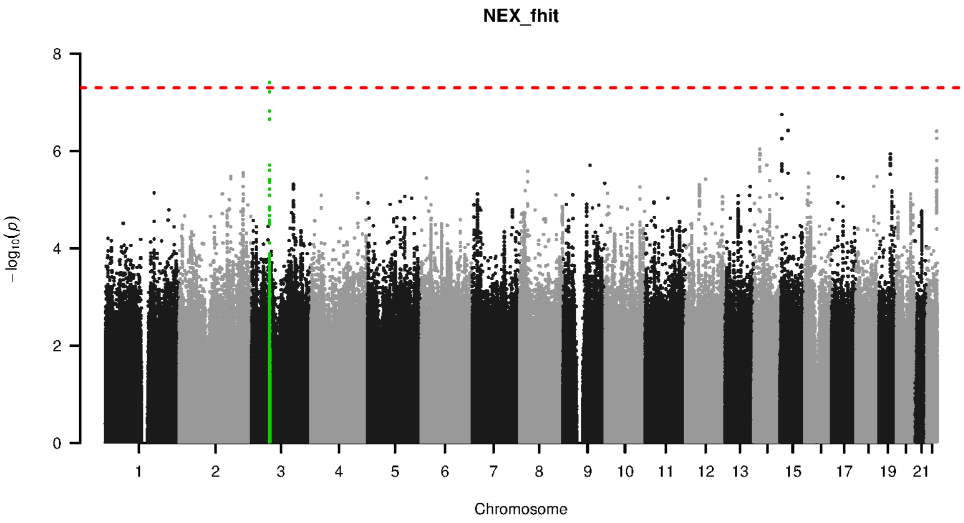


**Supplementary Figure 9-127. Manhattan plot and pQTL associations of the genome-wide meta-analysis for protein FHIT.** The horizontal dashed line corresponds to the genome-wide significance threshold of $p = 5 \times 10^{-8}$. The variants in the cis-regulatory region (±500kb of the coding gene region) of the protein-coding gene are highlighted in green.

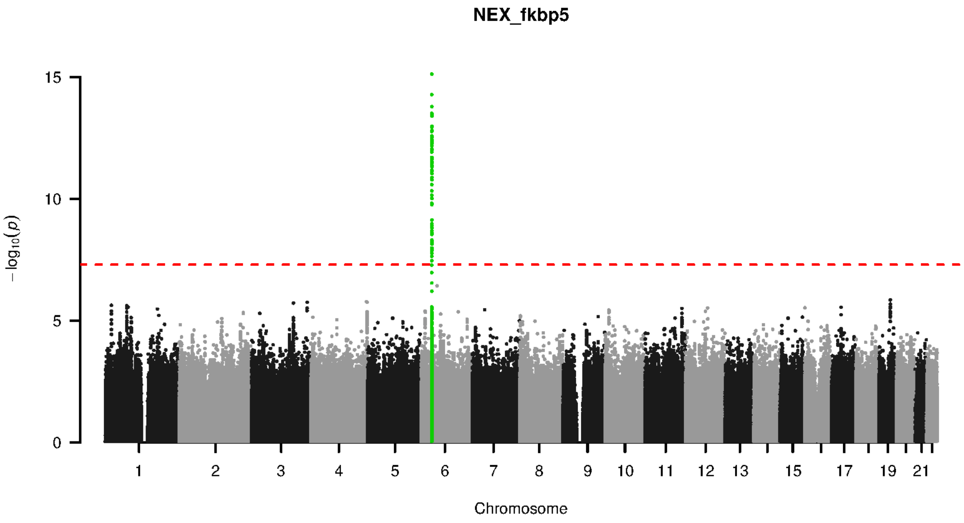


**Supplementary Figure 9-128. Manhattan plot and pQTL associations of the genome-wide meta-analysis for protein FKBP5.** The horizontal dashed line corresponds to the genome-wide significance threshold of $p = 5 \times 10^{-8}$. The variants in the cis-regulatory region (±500kb of the coding gene region) of the protein-coding gene are highlighted in green.

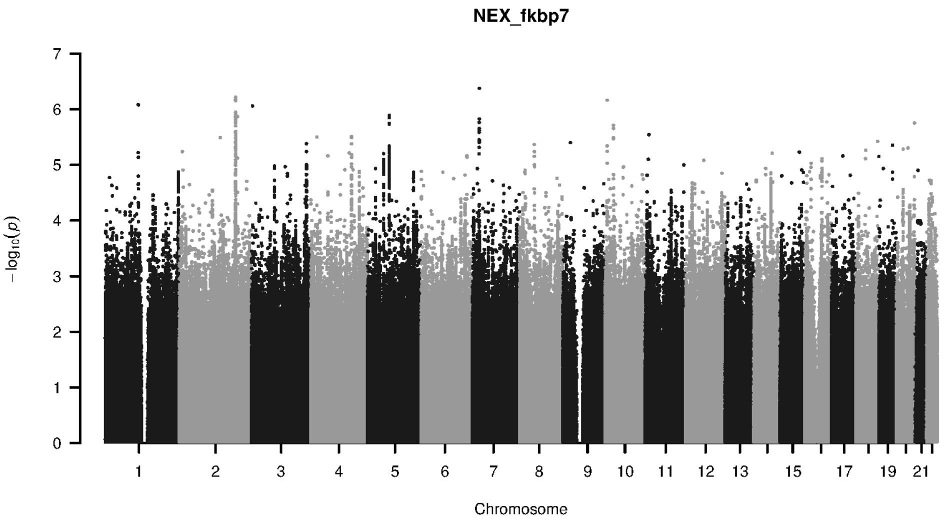


**Supplementary Figure 9-129. Manhattan plot and pQTL associations of the genome-wide meta-analysis for protein FKBP7.** The horizontal dashed line corresponds to the genome-wide significance threshold of $p = 5 \times 10^{-8}$. The variants in the cis-regulatory region (±500kb of the coding gene region) of the protein-coding gene are highlighted in green.

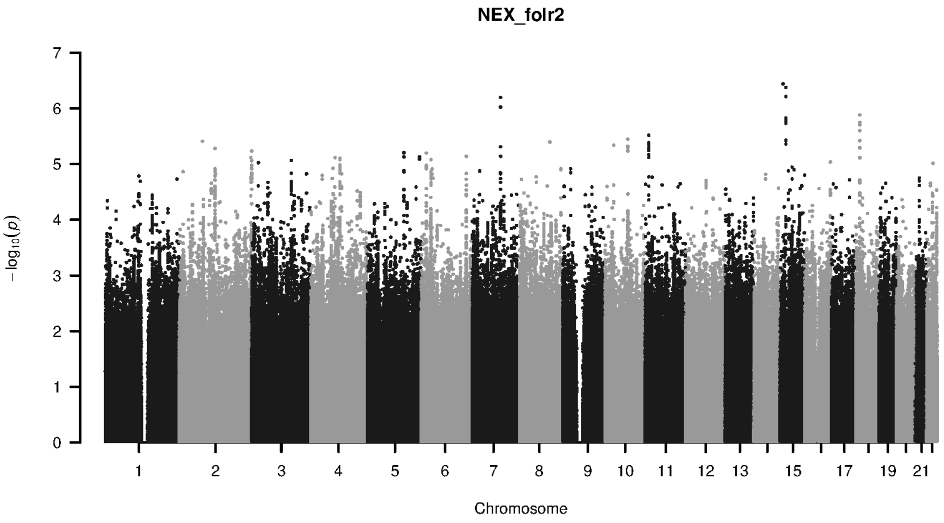


**Supplementary Figure 9-130. Manhattan plot and pQTL associations of the genome-wide meta-analysis for protein FOLR2.** The horizontal dashed line corresponds to the genome-wide significance threshold of $p = 5 \times 10^{-8}$. The variants in the cis-regulatory region (±500kb of the coding gene region) of the protein-coding gene are highlighted in green.

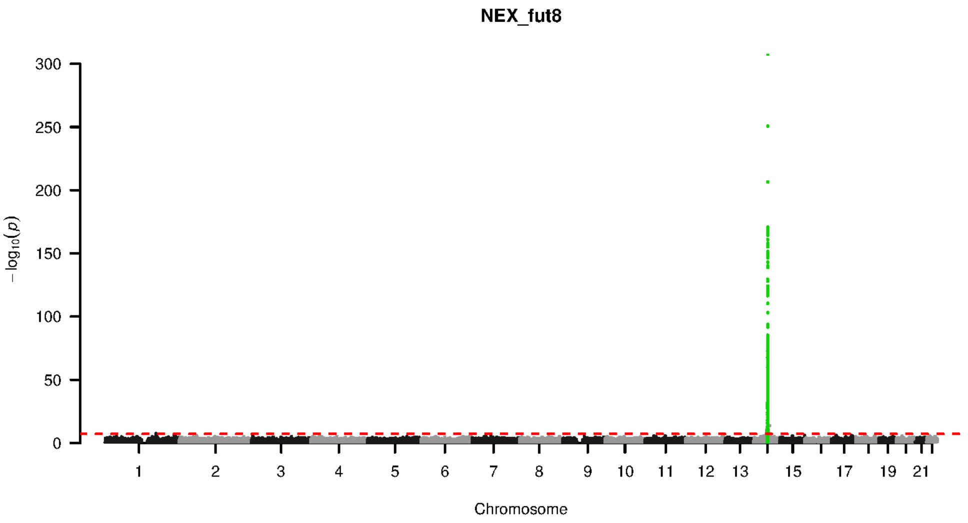


**Supplementary Figure 9-131. Manhattan plot and pQTL associations of the genome-wide meta-analysis for protein FUT8.** The horizontal dashed line corresponds to the genome-wide significance threshold of $p = 5 \times 10^{-8}$. The variants in the cis-regulatory region (±500kb of the coding gene region) of the protein-coding gene are highlighted in green.

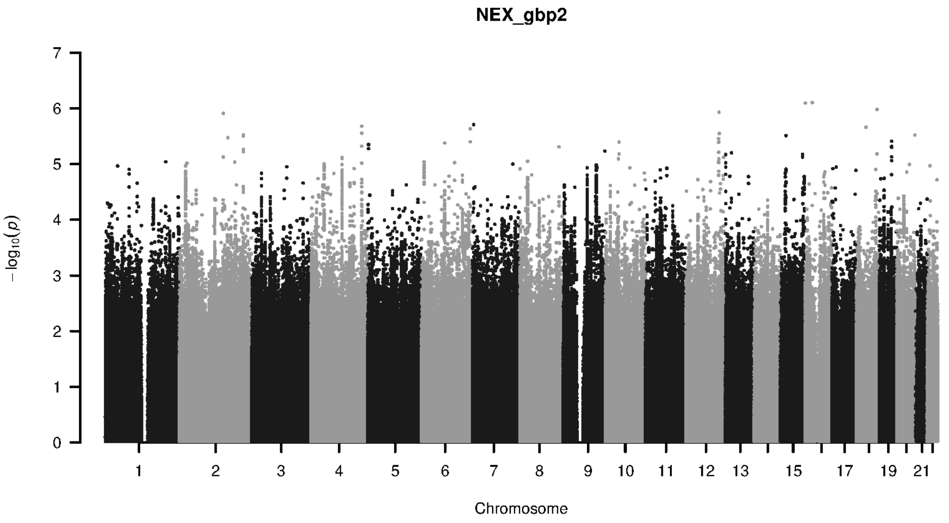


**Supplementary Figure 9-132. Manhattan plot and pQTL associations of the genome-wide meta-analysis for protein GBP2.** The horizontal dashed line corresponds to the genome-wide significance threshold of $p = 5 \times 10^{-8}$. The variants in the cis-regulatory region (±500kb of the coding gene region) of the protein-coding gene are highlighted in green.

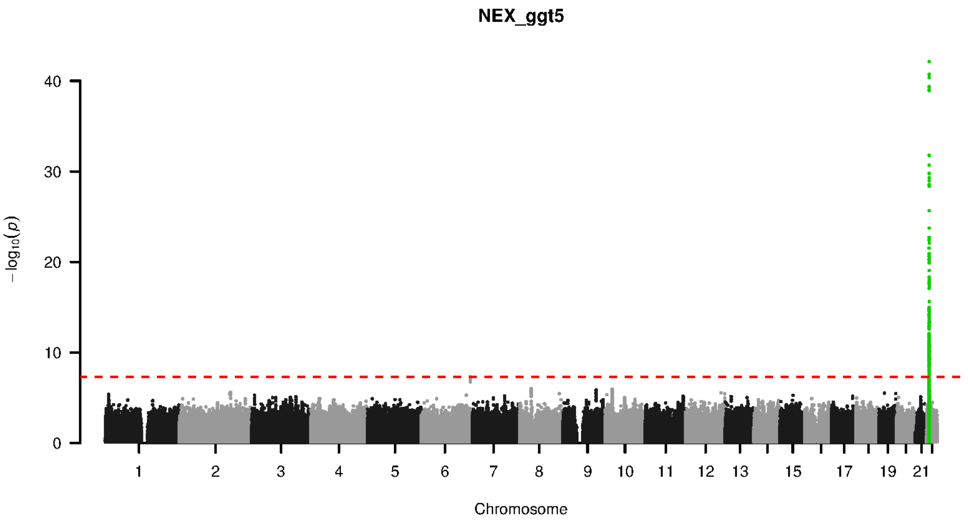


**Supplementary Figure 9-133. Manhattan plot and pQTL associations of the genome-wide meta-analysis for protein GGT5.** The horizontal dashed line corresponds to the genome-wide significance threshold of $p = 5 \times 10^{-8}$. The variants in the cis-regulatory region (±500kb of the coding gene region) of the protein-coding gene are highlighted in green.

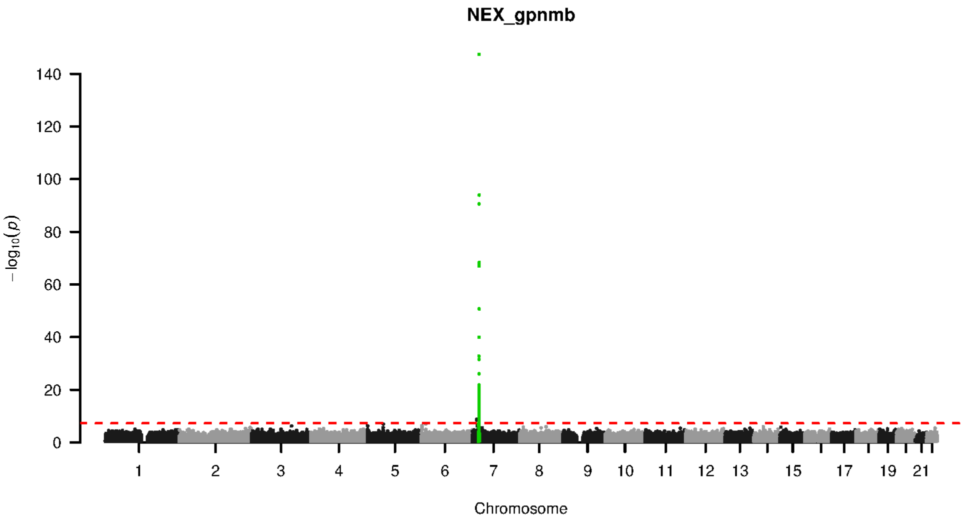


**Supplementary Figure 9-134. Manhattan plot and pQTL associations of the genome-wide meta-analysis for protein GPNMB.** The horizontal dashed line corresponds to the genome-wide significance threshold of $p = 5 \times 10^{-8}$. The variants in the cis-regulatory region (±500kb of the coding gene region) of the protein-coding gene are highlighted in green.

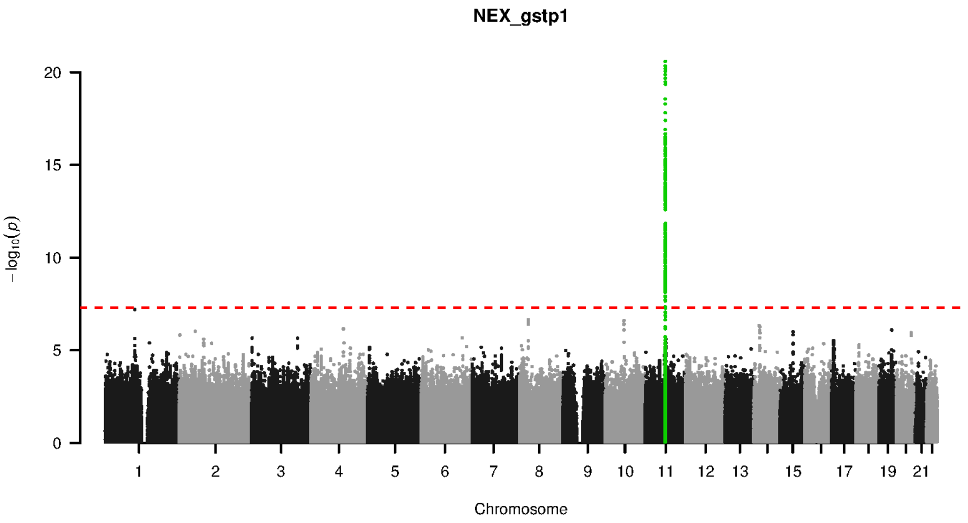


**Supplementary Figure 9-135. Manhattan plot and pQTL associations of the genome-wide meta-analysis for protein GSTP1.** The horizontal dashed line corresponds to the genome-wide significance threshold of $p = 5 \times 10^{-8}$. The variants in the cis-regulatory region ($\pm$500kb of the coding gene region) of the protein-coding gene are highlighted in green.

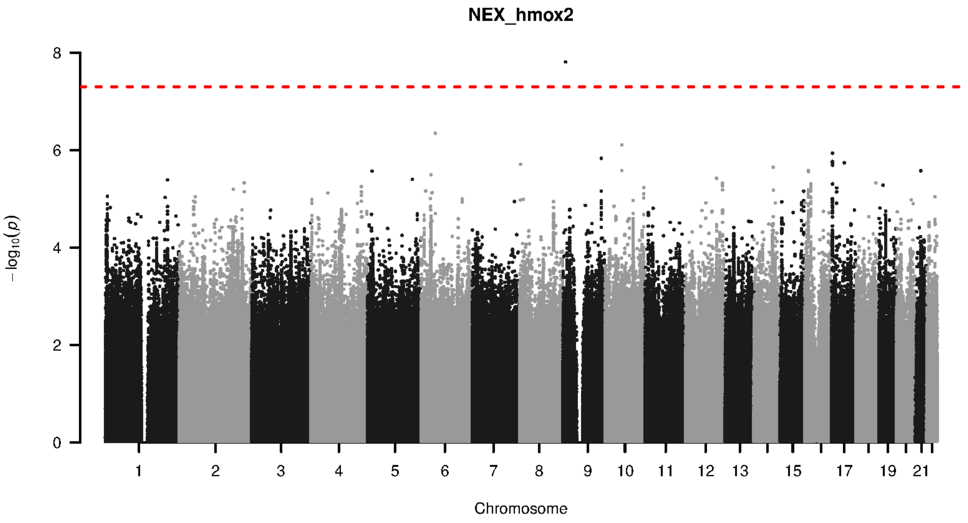


**Supplementary Figure 9-136. Manhattan plot and pQTL associations of the genome-wide meta-analysis for protein HMOX2.** The horizontal dashed line corresponds to the genome-wide significance threshold of $p = 5 \times 10^{-8}$. The variants in the cis-regulatory region ($\pm$500kb of the coding gene region) of the protein-coding gene are highlighted in green.

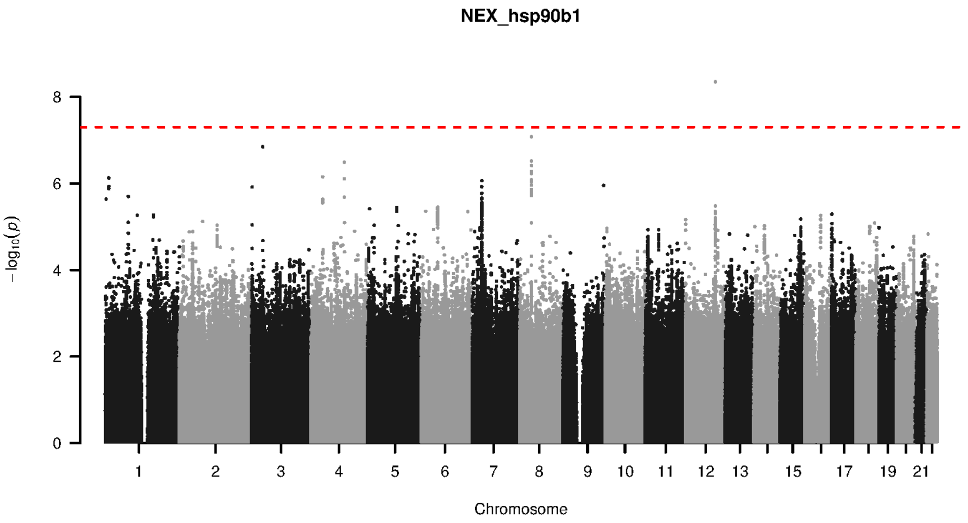


**Supplementary Figure 9-137. Manhattan plot and pQTL associations of the genome-wide meta-analysis for protein HSP90B1.** The horizontal dashed line corresponds to the genome-wide significance threshold of $p = 5 \times 10^{-8}$. The variants in the cis-regulatory region (±500kb of the coding gene region) of the protein-coding gene are highlighted in green.

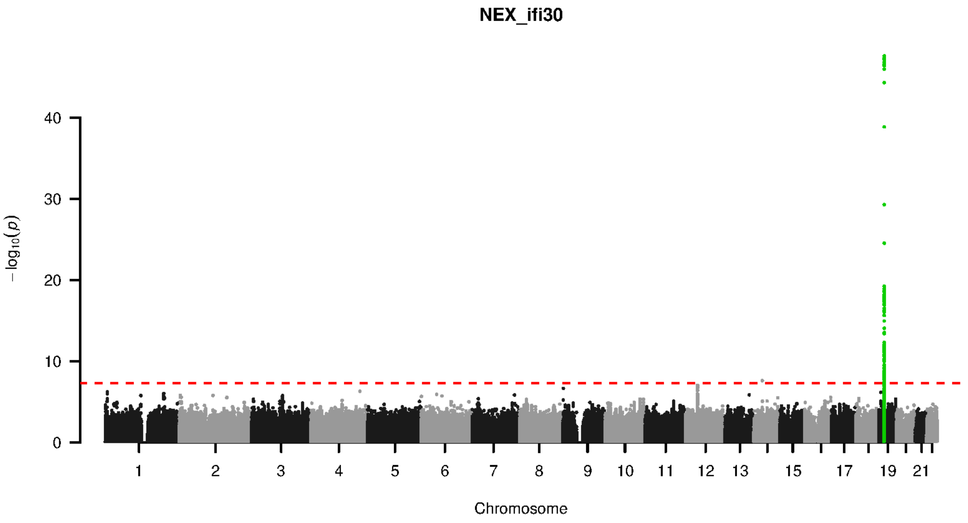


**Supplementary Figure 9-138. Manhattan plot and pQTL associations of the genome-wide meta-analysis for protein IFI30.** The horizontal dashed line corresponds to the genome-wide significance threshold of $p = 5 \times 10^{-8}$. The variants in the cis-regulatory region (±500kb of the coding gene region) of the protein-coding gene are highlighted in green.

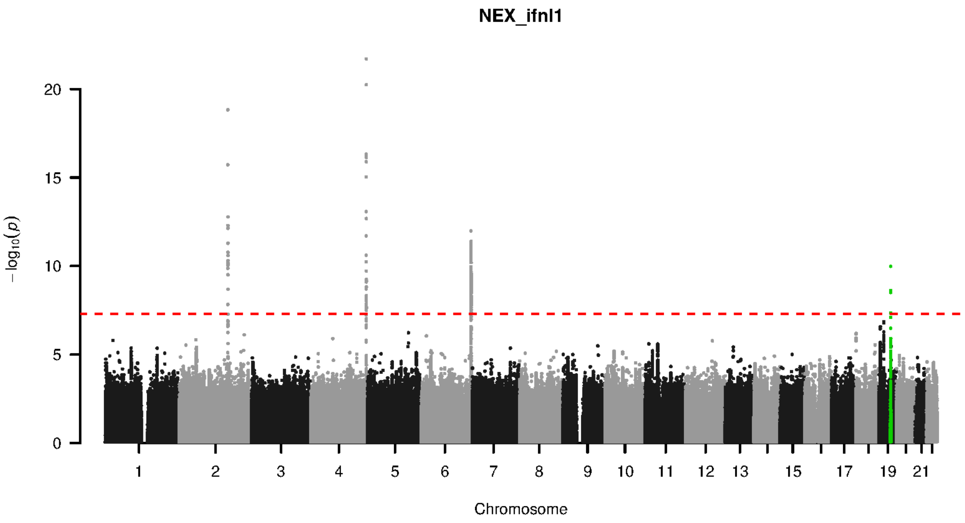


**Supplementary Figure 9-139. Manhattan plot and pQTL associations of the genome-wide meta-analysis for protein IFNL1.** The horizontal dashed line corresponds to the genome-wide significance threshold of $p = 5 \times 10^{-8}$. The variants in the cis-regulatory region (±500kb of the coding gene region) of the protein-coding gene are highlighted in green.

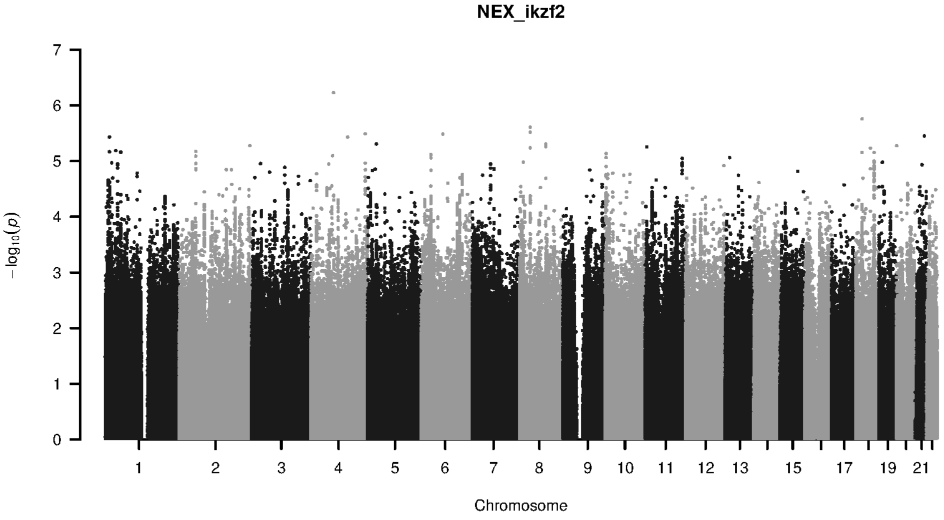


**Supplementary Figure 9-140. Manhattan plot and pQTL associations of the genome-wide meta-analysis for protein IKZF2.** The horizontal dashed line corresponds to the genome-wide significance threshold of $p = 5 \times 10^{-8}$. The variants in the cis-regulatory region (±500kb of the coding gene region) of the protein-coding gene are highlighted in green.

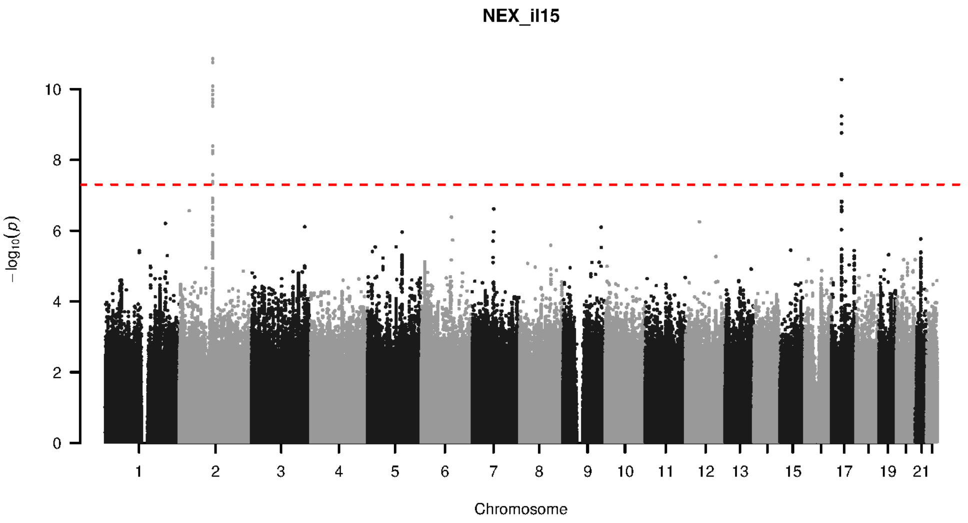


**Supplementary Figure 9-141. Manhattan plot and pQTL associations of the genome-wide meta-analysis for protein IL15.** The horizontal dashed line corresponds to the genome-wide significance threshold of $p = 5 \times 10^{-8}$. The variants in the cis-regulatory region (±500kb of the coding gene region) of the protein-coding gene are highlighted in green.

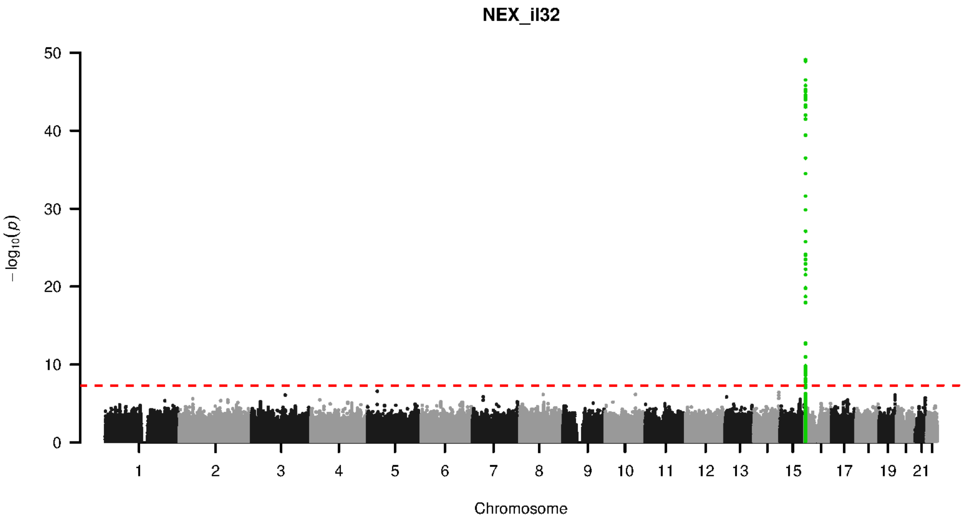


**Supplementary Figure 9-142. Manhattan plot and pQTL associations of the genome-wide meta-analysis for protein IL32.** The horizontal dashed line corresponds to the genome-wide significance threshold of $p = 5 \times 10^{-8}$. The variants in the cis-regulatory region (±500kb of the coding gene region) of the protein-coding gene are highlighted in green.

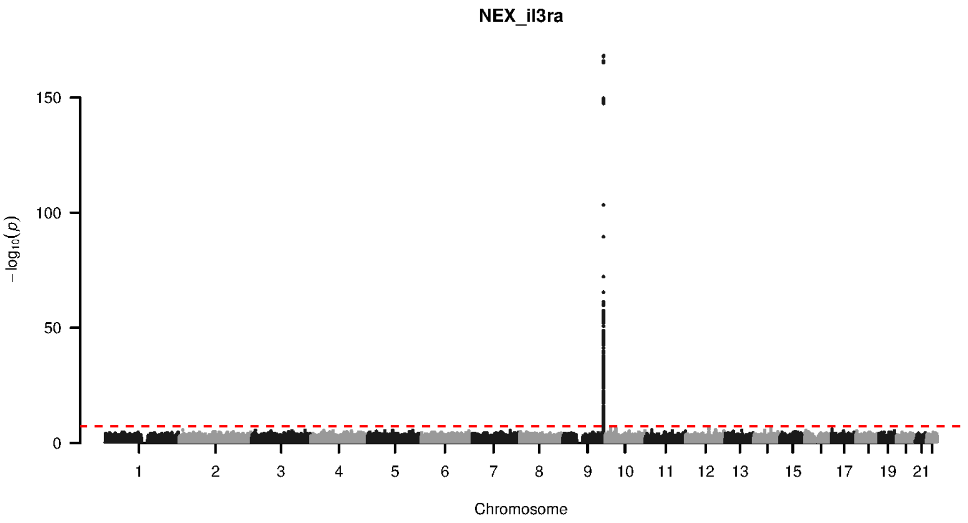


**Supplementary Figure 9-143. Manhattan plot and pQTL associations of the genome-wide meta-analysis for protein IL3RA.** The horizontal dashed line corresponds to the genome-wide significance threshold of $p = 5 \times 10^{-8}$. The variants in the cis-regulatory region (±500kb of the coding gene region) of the protein-coding gene are highlighted in green.

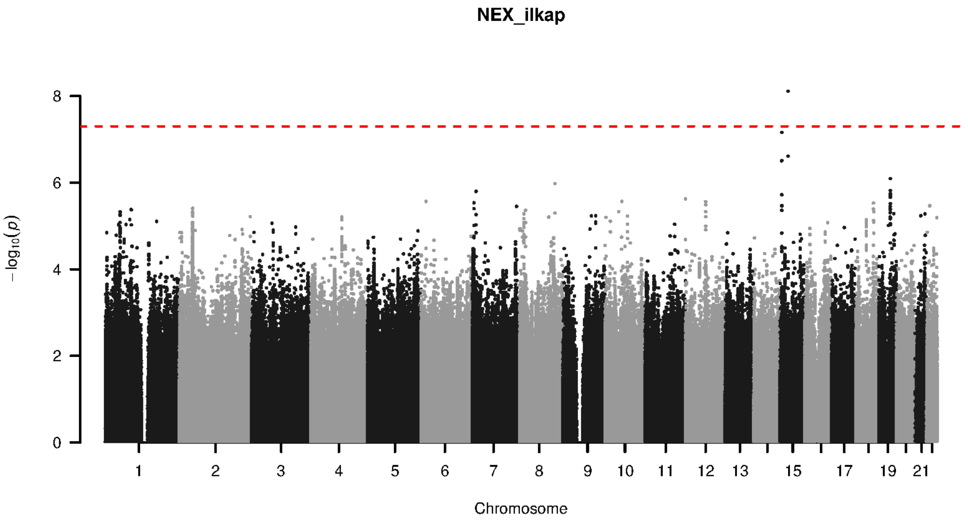


**Supplementary Figure 9-144. Manhattan plot and pQTL associations of the genome-wide meta-analysis for protein ILKAP.** The horizontal dashed line corresponds to the genome-wide significance threshold of $p = 5 \times 10^{-8}$. The variants in the cis-regulatory region (±500kb of the coding gene region) of the protein-coding gene are highlighted in green.

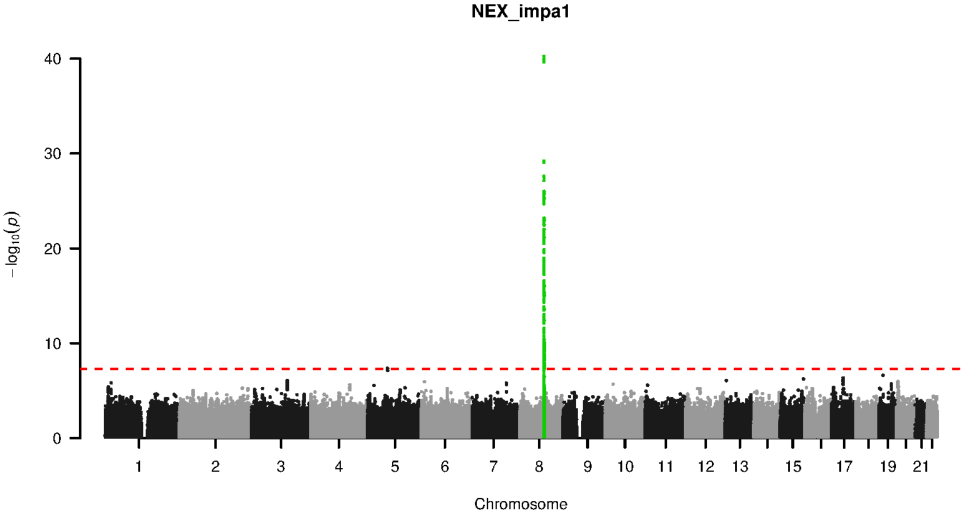


**Supplementary Figure 9-145. Manhattan plot and pQTL associations of the genome-wide meta-analysis for protein IMPA1.** The horizontal dashed line corresponds to the genome-wide significance threshold of $p = 5 \times 10^{-8}$. The variants in the cis-regulatory region (±500kb of the coding gene region) of the protein-coding gene are highlighted in green.

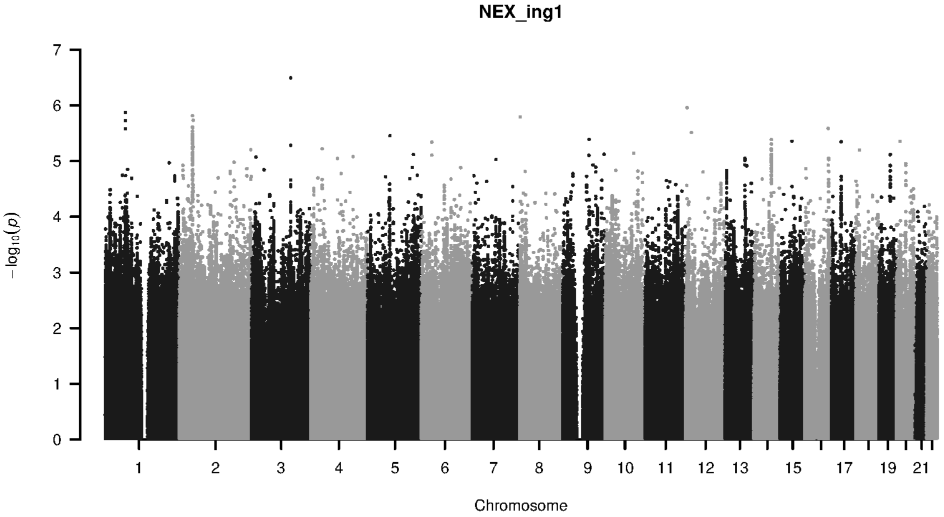


**Supplementary Figure 9-146. Manhattan plot and pQTL associations of the genome-wide meta-analysis for protein ING1.** The horizontal dashed line corresponds to the genome-wide significance threshold of $p = 5 \times 10^{-8}$. The variants in the cis-regulatory region (±500kb of the coding gene region) of the protein-coding gene are highlighted in green.

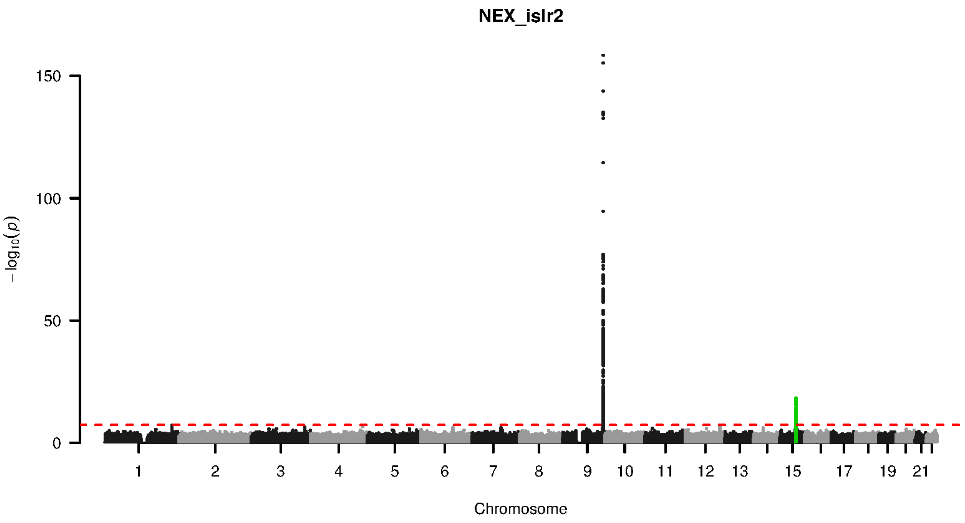


**Supplementary Figure 9-147. Manhattan plot and pQTL associations of the genome-wide meta-analysis for protein ISLR2.** The horizontal dashed line corresponds to the genome-wide significance threshold of $p = 5 \times 10^{-8}$. The variants in the cis-regulatory region ($\pm$500kb of the coding gene region) of the protein-coding gene are highlighted in green.

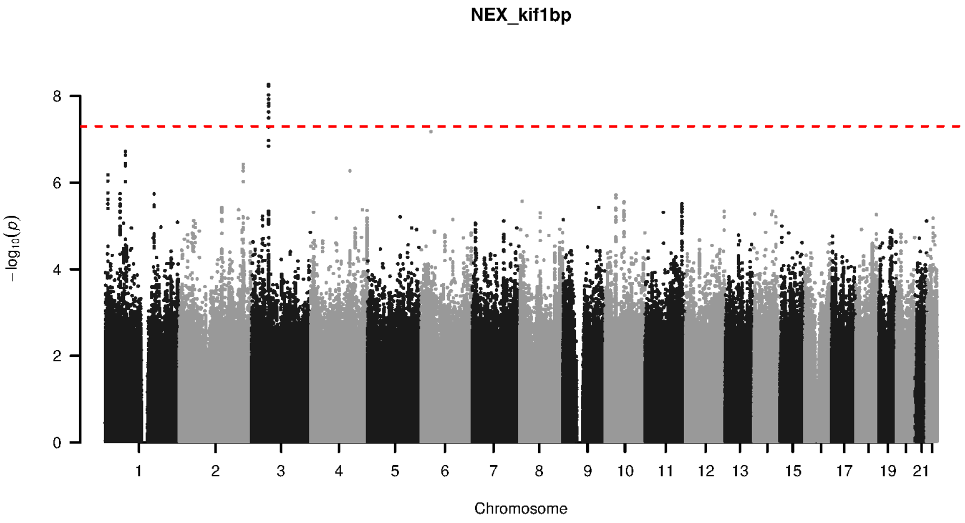


**Supplementary Figure 9-148. Manhattan plot and pQTL associations of the genome-wide meta-analysis for protein KIF1BP.** The horizontal dashed line corresponds to the genome-wide significance threshold of $p = 5 \times 10^{-8}$. The variants in the cis-regulatory region ($\pm$500kb of the coding gene region) of the protein-coding gene are highlighted in green.

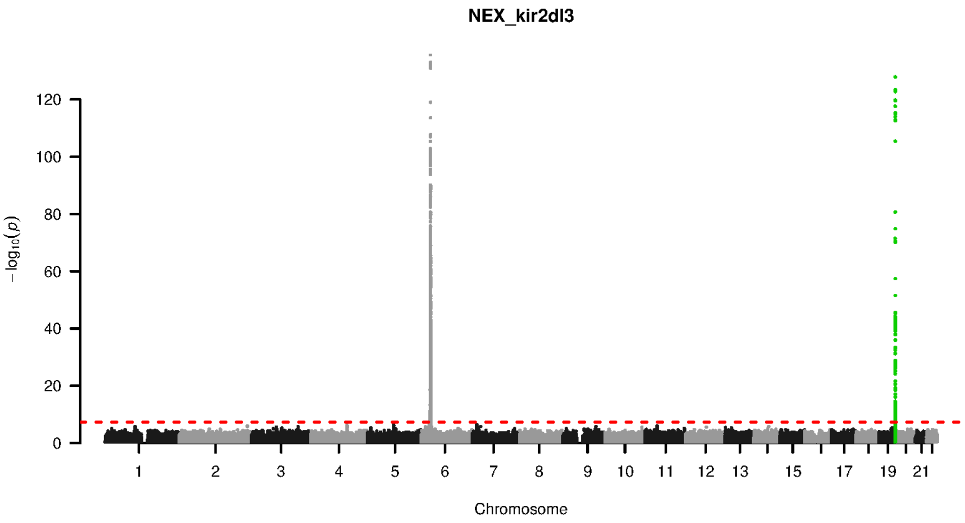


**Supplementary Figure 9-149. Manhattan plot and pQTL associations of the genome-wide meta-analysis for protein KIR2DL3.** The horizontal dashed line corresponds to the genome-wide significance threshold of $p = 5\times10^{-8}$. The variants in the cis-regulatory region (±500kb of the coding gene region) of the protein-coding gene are highlighted in green.

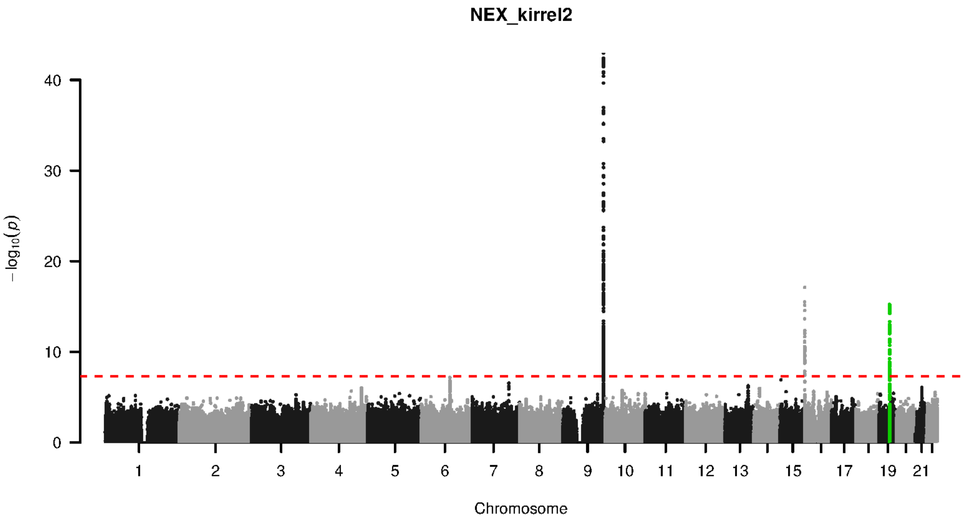


**Supplementary Figure 9-150. Manhattan plot and pQTL associations of the genome-wide meta-analysis for protein KIRREL2.** The horizontal dashed line corresponds to the genome-wide significance threshold of $p = 5\times10^{-8}$. The variants in the cis-regulatory region (±500kb of the coding gene region) of the protein-coding gene are highlighted in green.

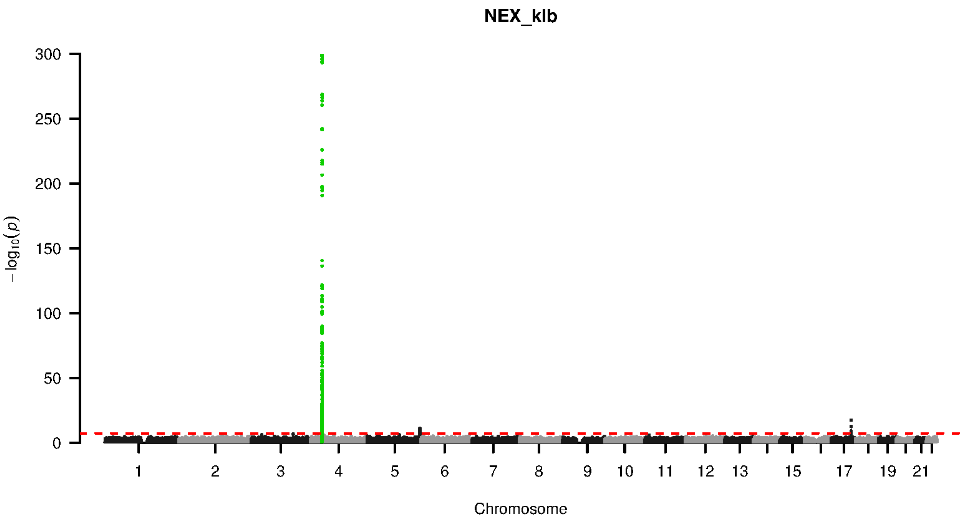


**Supplementary Figure 9-151. Manhattan plot and pQTL associations of the genome-wide meta-analysis for protein KLB.** The horizontal dashed line corresponds to the genome-wide significance threshold of $p = 5 \times 10^{-8}$. The variants in the cis-regulatory region (±500kb of the coding gene region) of the protein-coding gene are highlighted in green.

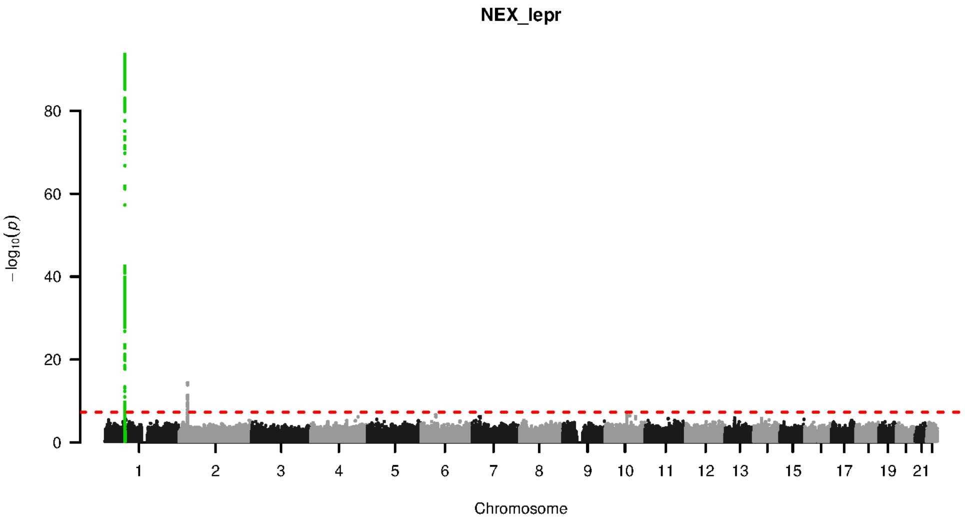


**Supplementary Figure 9-152. Manhattan plot and pQTL associations of the genome-wide meta-analysis for protein LEPR.** The horizontal dashed line corresponds to the genome-wide significance threshold of $p = 5 \times 10^{-8}$. The variants in the cis-regulatory region (±500kb of the coding gene region) of the protein-coding gene are highlighted in green.

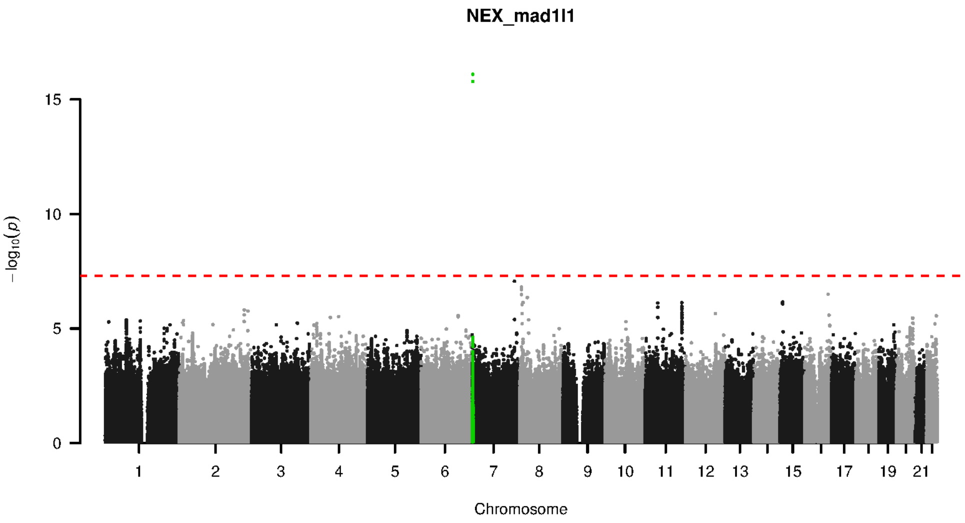


**Supplementary Figure 9-153. Manhattan plot and pQTL associations of the genome-wide meta-analysis for protein MAD1L1.** The horizontal dashed line corresponds to the genome-wide significance threshold of $p = 5 \times 10^{-8}$. The variants in the cis-regulatory region (±500kb of the coding gene region) of the protein-coding gene are highlighted in green.

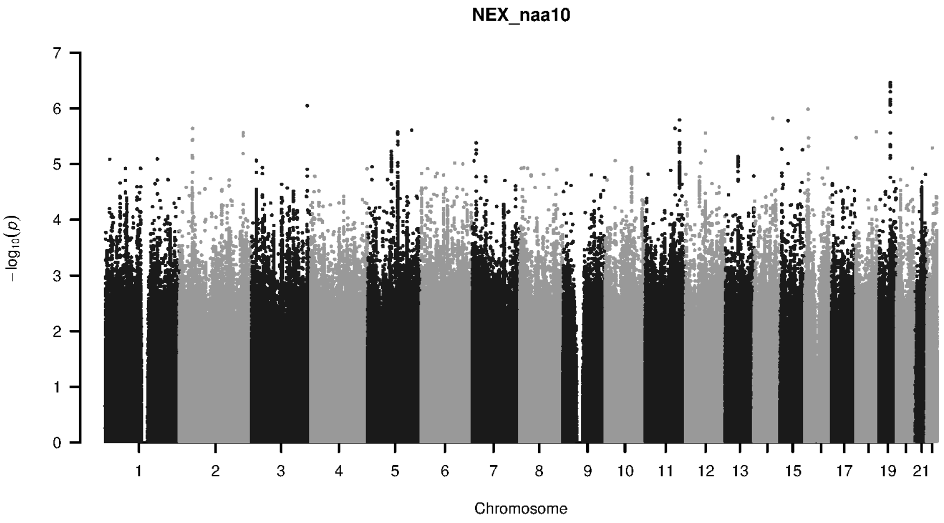


**Supplementary Figure 9-154. Manhattan plot and pQTL associations of the genome-wide meta-analysis for protein NAA10.** The horizontal dashed line corresponds to the genome-wide significance threshold of $p = 5 \times 10^{-8}$. The variants in the cis-regulatory region (±500kb of the coding gene region) of the protein-coding gene are highlighted in green.

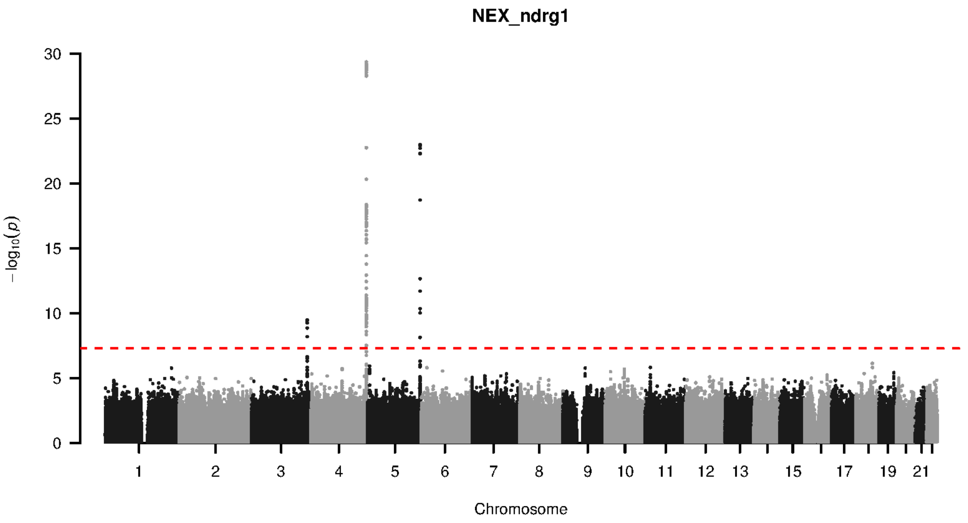


**Supplementary Figure 9-155. Manhattan plot and pQTL associations of the genome-wide meta-analysis for protein NDRG1.** The horizontal dashed line corresponds to the genome-wide significance threshold of $p = 5 \times 10^{-8}$. The variants in the cis-regulatory region (±500kb of the coding gene region) of the protein-coding gene are highlighted in green.

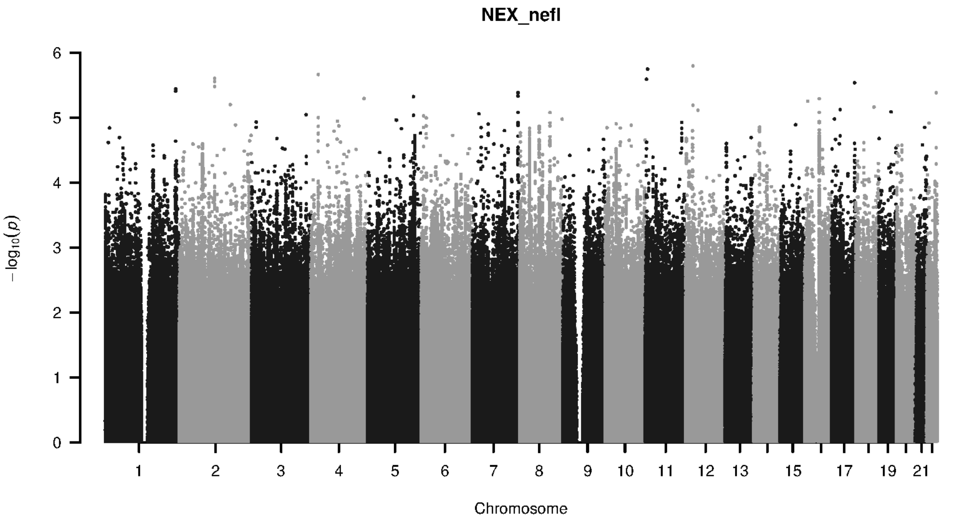


**Supplementary Figure 9-156. Manhattan plot and pQTL associations of the genome-wide meta-analysis for protein NEFL.** The horizontal dashed line corresponds to the genome-wide significance threshold of $p = 5 \times 10^{-8}$. The variants in the cis-regulatory region (±500kb of the coding gene region) of the protein-coding gene are highlighted in green.

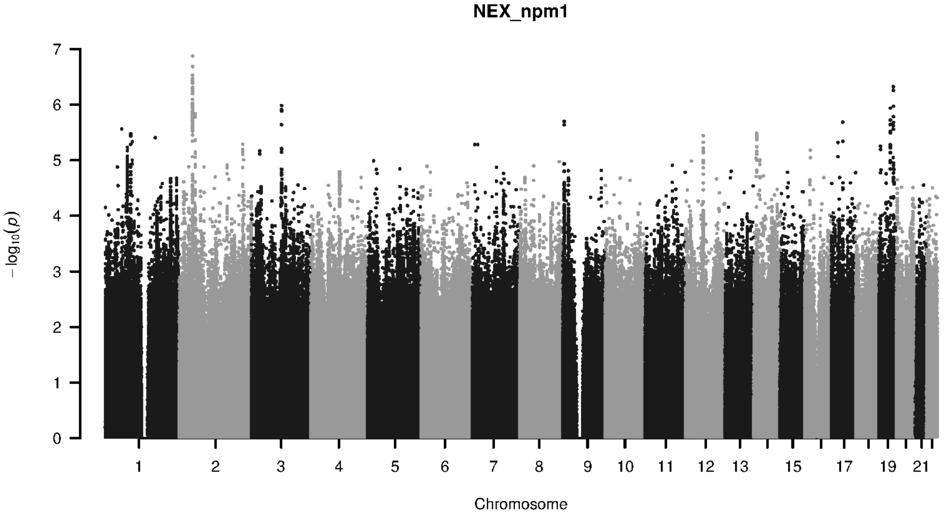


**Supplementary Figure 9-157. Manhattan plot and pQTL associations of the genome-wide meta-analysis for protein NPM1.** The horizontal dashed line corresponds to the genome-wide significance threshold of $p = 5 \times 10^{-8}$. The variants in the cis-regulatory region (±500kb of the coding gene region) of the protein-coding gene are highlighted in green.

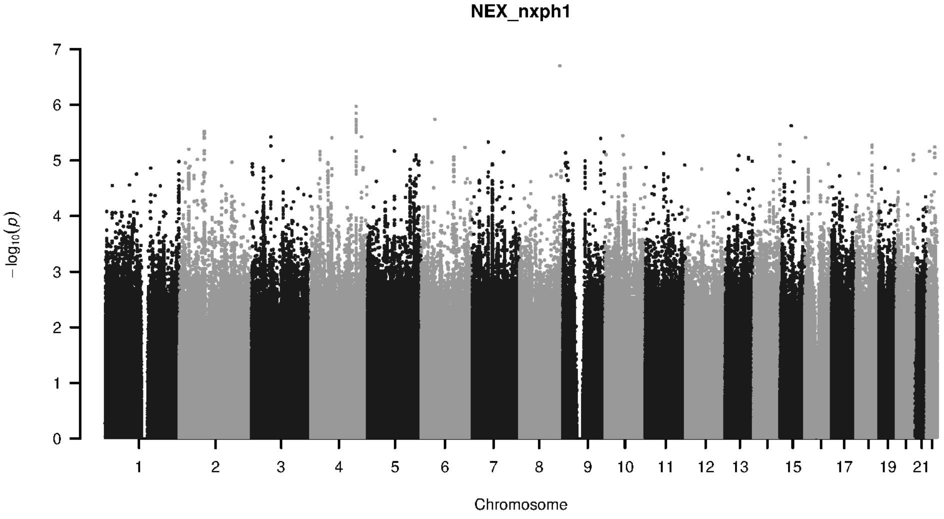


**Supplementary Figure 9-158. Manhattan plot and pQTL associations of the genome-wide meta-analysis for protein NXPH1.** The horizontal dashed line corresponds to the genome-wide significance threshold of $p = 5 \times 10^{-8}$. The variants in the cis-regulatory region (±500kb of the coding gene region) of the protein-coding gene are highlighted in green.

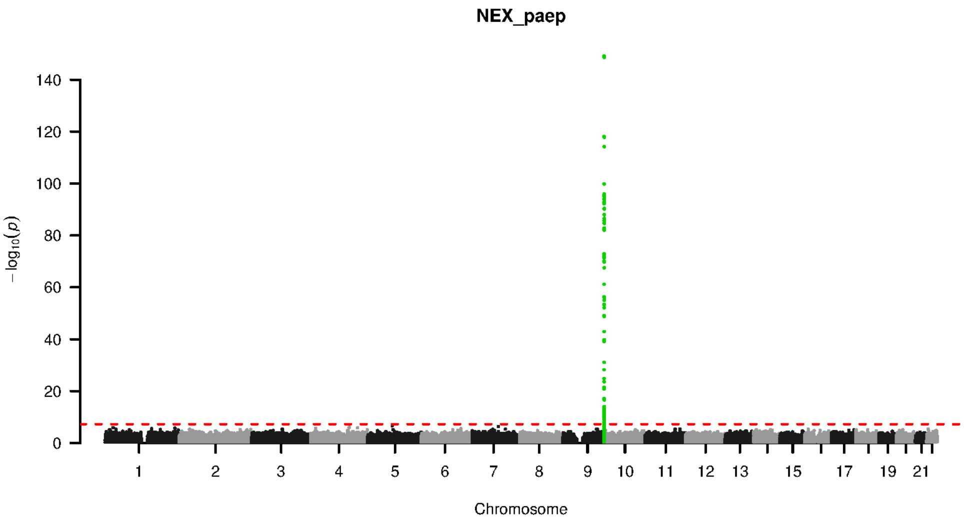


**Supplementary Figure 9-159. Manhattan plot and pQTL associations of the genome-wide meta-analysis for protein PAEP.** The horizontal dashed line corresponds to the genome-wide significance threshold of $p = 5 \times 10^{-8}$. The variants in the cis-regulatory region (±500kb of the coding gene region) of the protein-coding gene are highlighted in green.

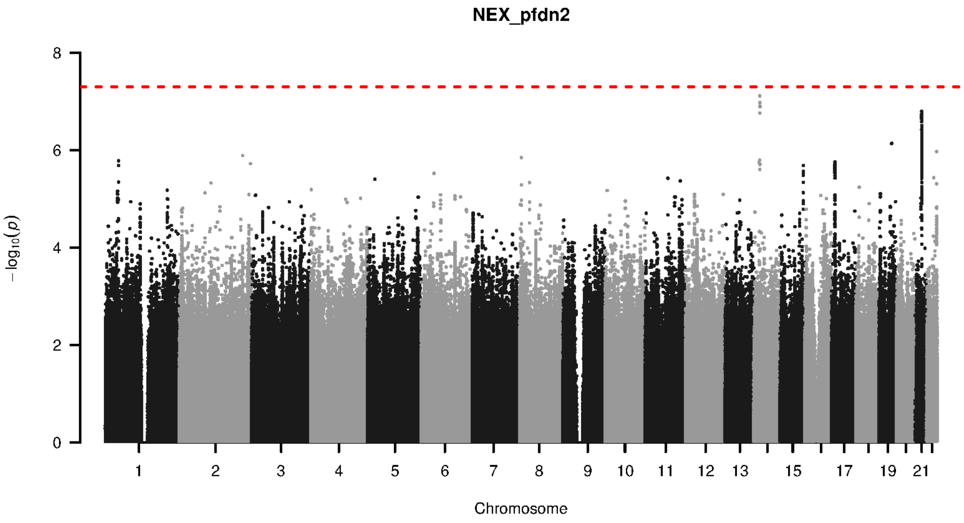


**Supplementary Figure 9-160. Manhattan plot and pQTL associations of the genome-wide meta-analysis for protein PFDN2.** The horizontal dashed line corresponds to the genome-wide significance threshold of $p = 5 \times 10^{-8}$. The variants in the cis-regulatory region (±500kb of the coding gene region) of the protein-coding gene are highlighted in green.

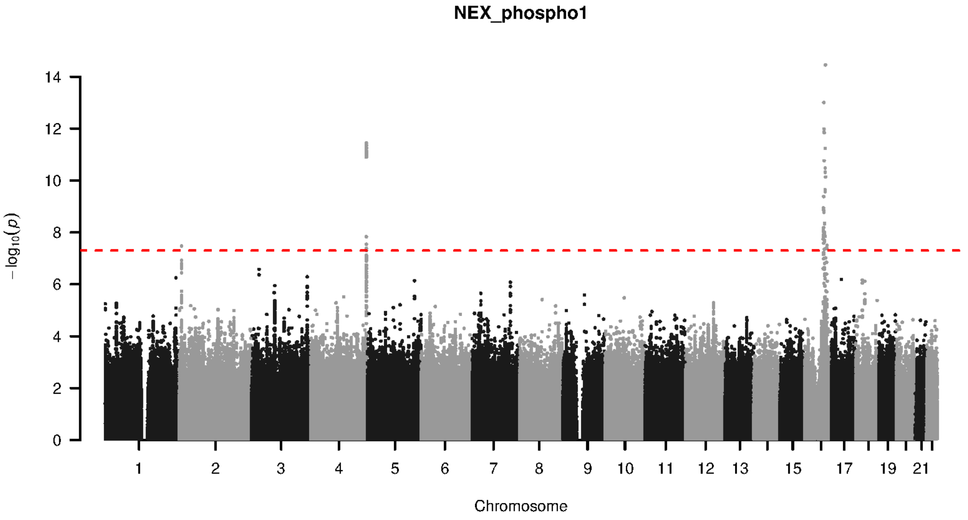


**Supplementary Figure 9-161. Manhattan plot and pQTL associations of the genome-wide meta-analysis for protein PHOSPHO1.** The horizontal dashed line corresponds to the genome-wide significance threshold of $p = 5 \times 10^{-8}$. The variants in the cis-regulatory region (±500kb of the coding gene region) of the protein-coding gene are highlighted in green.

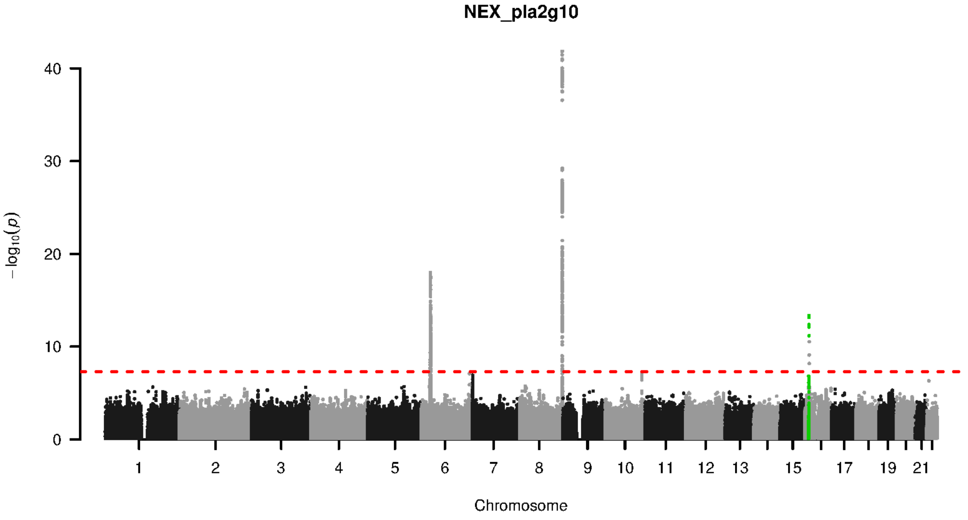


**Supplementary Figure 9-162. Manhattan plot and pQTL associations of the genome-wide meta-analysis for protein PLA2G10.** The horizontal dashed line corresponds to the genome-wide significance threshold of $p = 5 \times 10^{-8}$. The variants in the cis-regulatory region (±500kb of the coding gene region) of the protein-coding gene are highlighted in green.

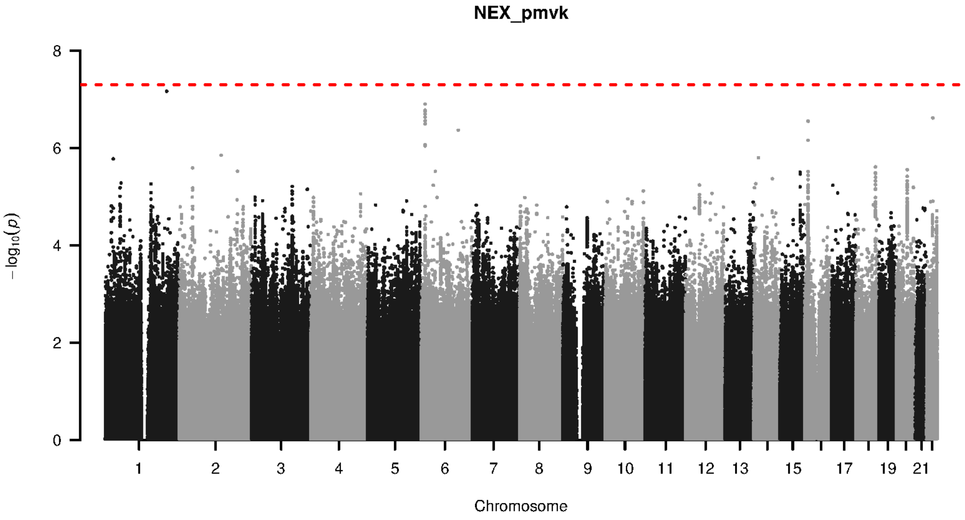


**Supplementary Figure 9-163. Manhattan plot and pQTL associations of the genome-wide meta-analysis for protein PMVK.** The horizontal dashed line corresponds to the genome-wide significance threshold of $p = 5 \times 10^{-8}$. The variants in the cis-regulatory region (±500kb of the coding gene region) of the protein-coding gene are highlighted in green.

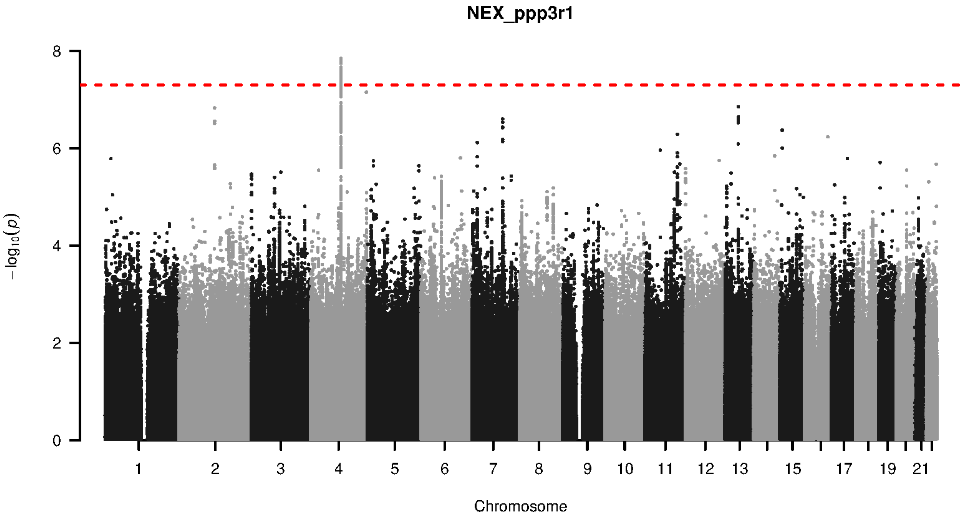


**Supplementary Figure 9-164. Manhattan plot and pQTL associations of the genome-wide meta-analysis for protein PPP3R1.** The horizontal dashed line corresponds to the genome-wide significance threshold of $p = 5 \times 10^{-8}$. The variants in the cis-regulatory region (±500kb of the coding gene region) of the protein-coding gene are highlighted in green.

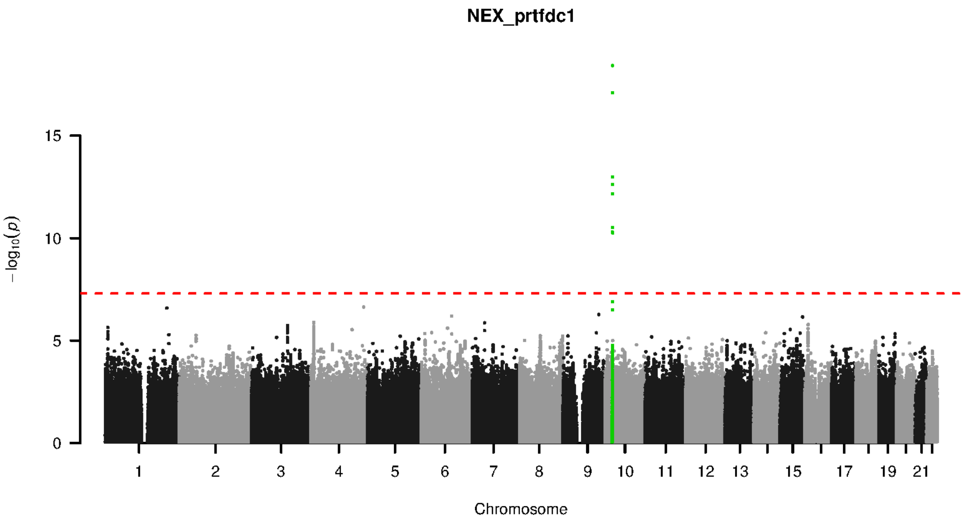


**Supplementary Figure 9-165. Manhattan plot and pQTL associations of the genome-wide meta-analysis for protein PRTFDC1.** The horizontal dashed line corresponds to the genome-wide significance threshold of $p = 5 \times 10^{-8}$. The variants in the cis-regulatory region (±500kb of the coding gene region) of the protein-coding gene are highlighted in green.

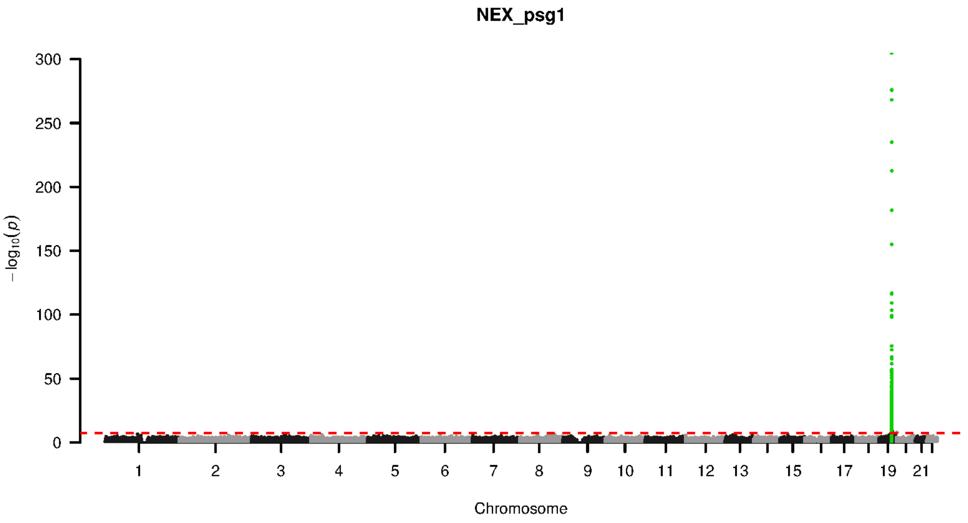


**Supplementary Figure 9-166. Manhattan plot and pQTL associations of the genome-wide meta-analysis for protein PSG1.** The horizontal dashed line corresponds to the genome-wide significance threshold of $p = 5 \times 10^{-8}$. The variants in the cis-regulatory region (±500kb of the coding gene region) of the protein-coding gene are highlighted in green.

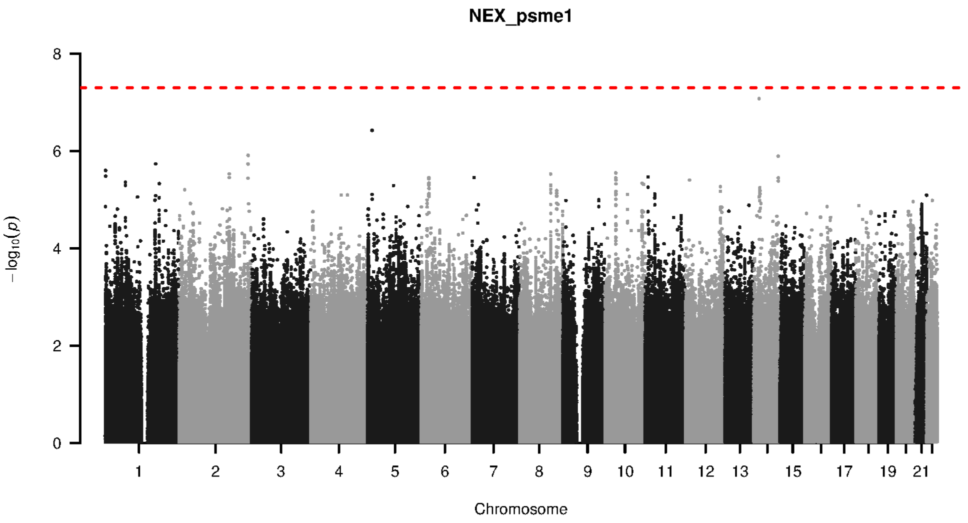


**Supplementary Figure 9-167. Manhattan plot and pQTL associations of the genome-wide meta-analysis for protein PSME1.** The horizontal dashed line corresponds to the genome-wide significance threshold of $p = 5 \times 10^{-8}$. The variants in the cis-regulatory region (±500kb of the coding gene region) of the protein-coding gene are highlighted in green.

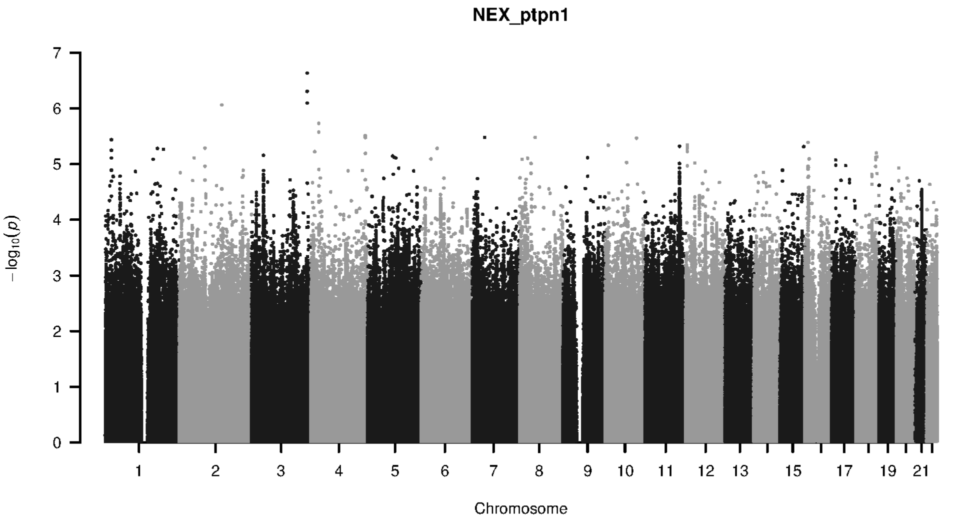


**Supplementary Figure 9-168. Manhattan plot and pQTL associations of the genome-wide meta-analysis for protein PTPN1.** The horizontal dashed line corresponds to the genome-wide significance threshold of $p = 5 \times 10^{-8}$. The variants in the cis-regulatory region (±500kb of the coding gene region) of the protein-coding gene are highlighted in green.

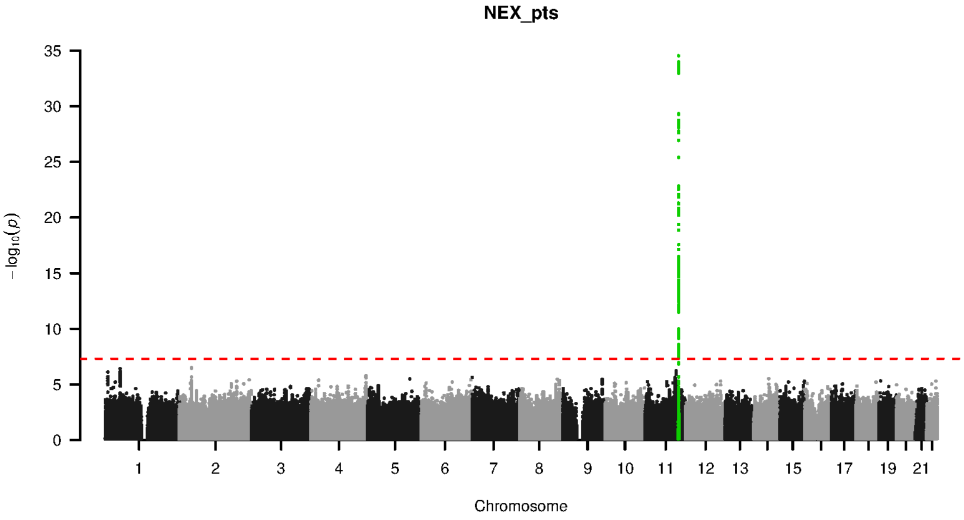


**Supplementary Figure 9-169. Manhattan plot and pQTL associations of the genome-wide meta-analysis for protein PTS.** The horizontal dashed line corresponds to the genome-wide significance threshold of $p = 5 \times 10^{-8}$. The variants in the cis-regulatory region ($\pm$500kb of the coding gene region) of the protein-coding gene are highlighted in green.

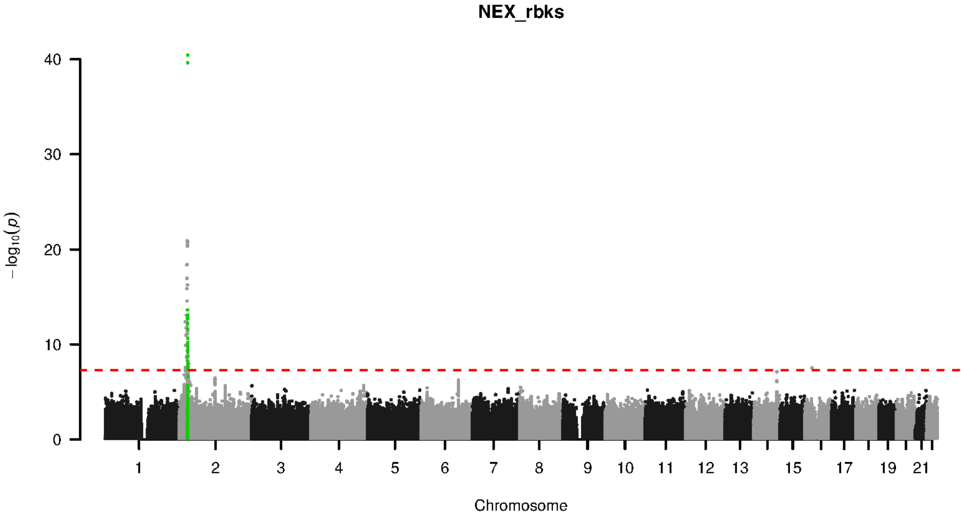


**Supplementary Figure 9-170. Manhattan plot and pQTL associations of the genome-wide meta-analysis for protein RBKS.** The horizontal dashed line corresponds to the genome-wide significance threshold of $p = 5 \times 10^{-8}$. The variants in the cis-regulatory region ($\pm$500kb of the coding gene region) of the protein-coding gene are highlighted in green.

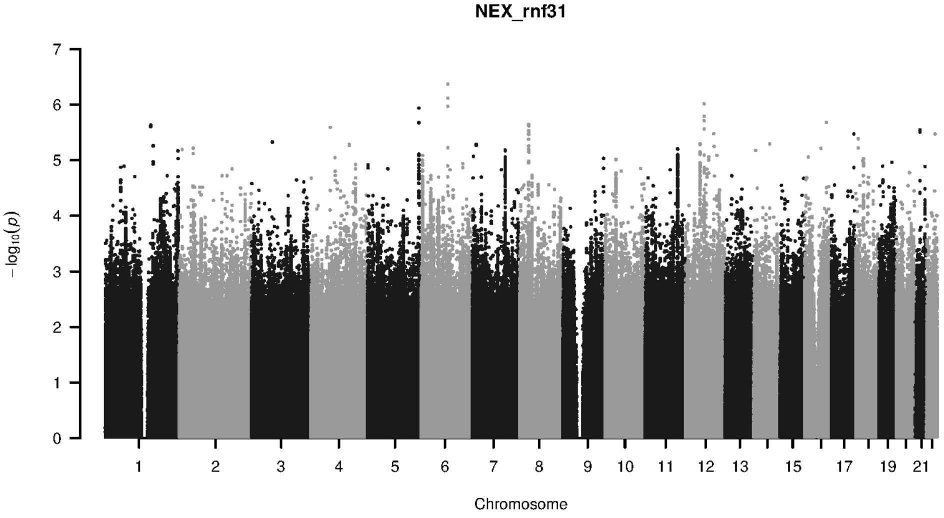


**Supplementary Figure 9-171. Manhattan plot and pQTL associations of the genome-wide meta-analysis for protein RNF31.** The horizontal dashed line corresponds to the genome-wide significance threshold of $p = 5 \times 10^{-8}$. The variants in the cis-regulatory region (±500kb of the coding gene region) of the protein-coding gene are highlighted in green.

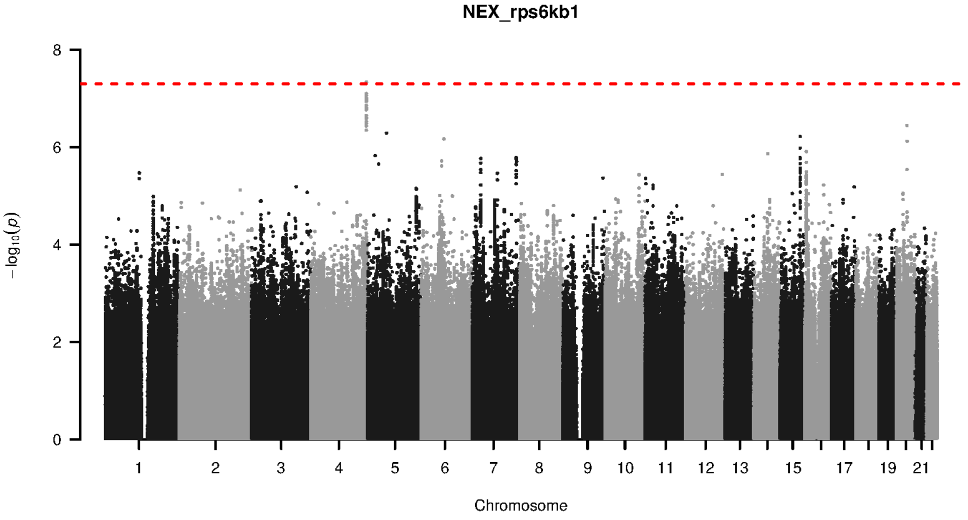


**Supplementary Figure 9-172. Manhattan plot and pQTL associations of the genome-wide meta-analysis for protein RPS6KB1.** The horizontal dashed line corresponds to the genome-wide significance threshold of $p = 5 \times 10^{-8}$. The variants in the cis-regulatory region (±500kb of the coding gene region) of the protein-coding gene are highlighted in green.

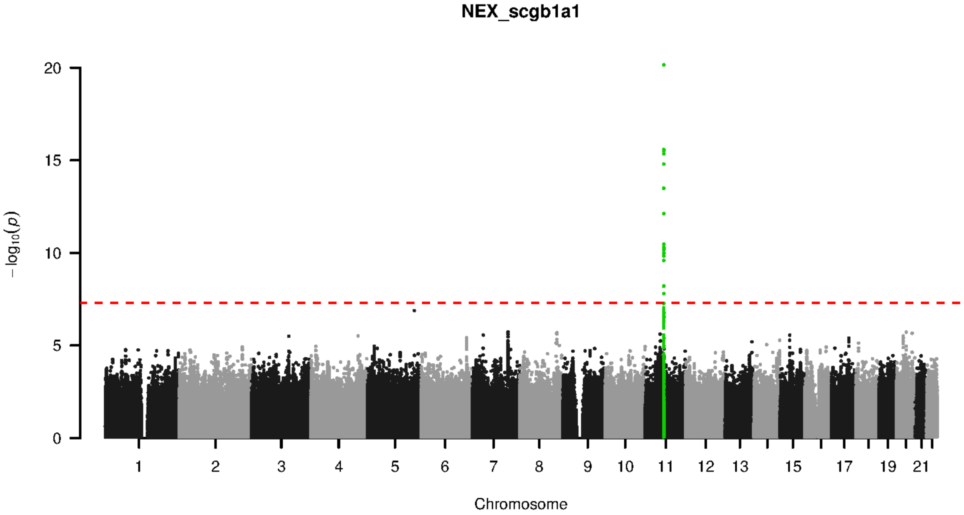


**Supplementary Figure 9-173. Manhattan plot and pQTL associations of the genome-wide meta-analysis for protein SCGB1A1.** The horizontal dashed line corresponds to the genome-wide significance threshold of $p = 5 \times 10^{-8}$. The variants in the cis-regulatory region (±500kb of the coding gene region) of the protein-coding gene are highlighted in green.

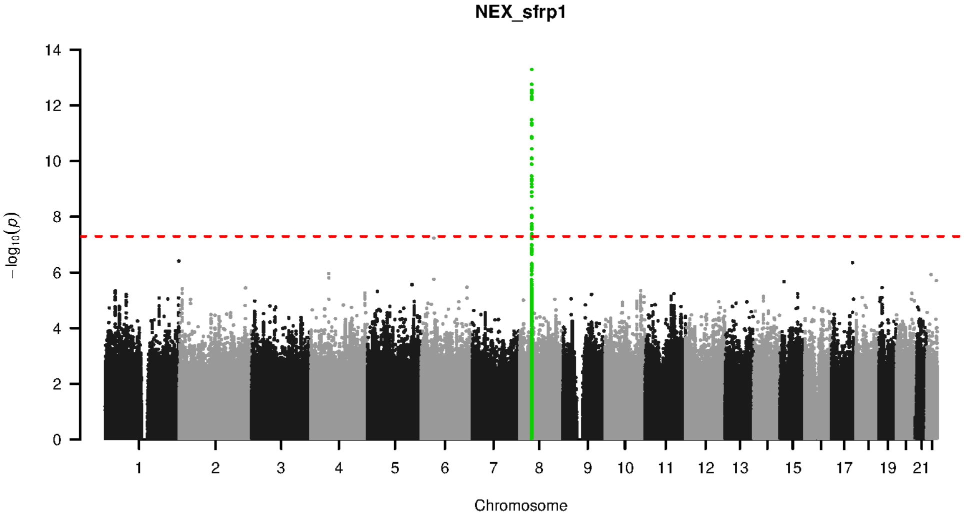


**Supplementary Figure 9-174. Manhattan plot and pQTL associations of the genome-wide meta-analysis for protein SFRP1.** The horizontal dashed line corresponds to the genome-wide significance threshold of $p = 5 \times 10^{-8}$. The variants in the cis-regulatory region (±500kb of the coding gene region) of the protein-coding gene are highlighted in green.

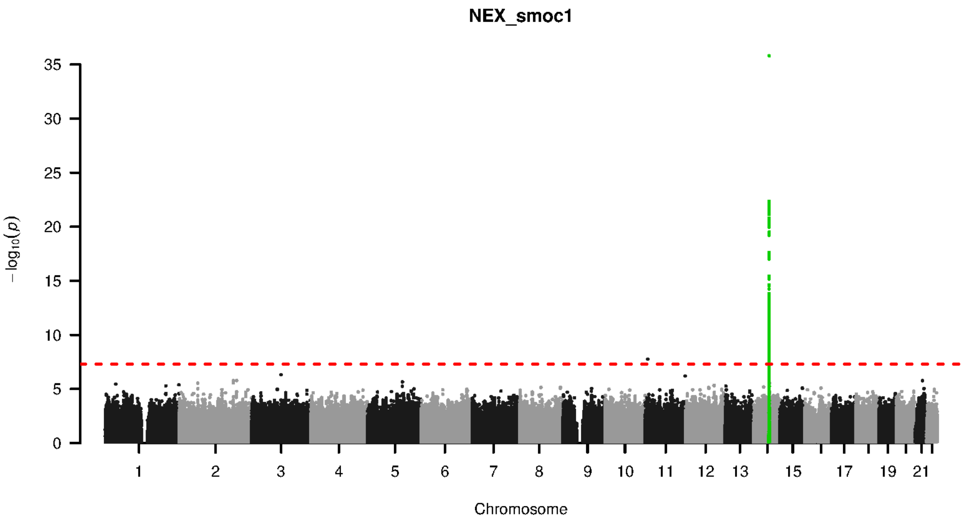


**Supplementary Figure 9-175. Manhattan plot and pQTL associations of the genome-wide meta-analysis for protein SMOC1.** The horizontal dashed line corresponds to the genome-wide significance threshold of $p = 5 \times 10^{-8}$. The variants in the cis-regulatory region (±500kb of the coding gene region) of the protein-coding gene are highlighted in green.

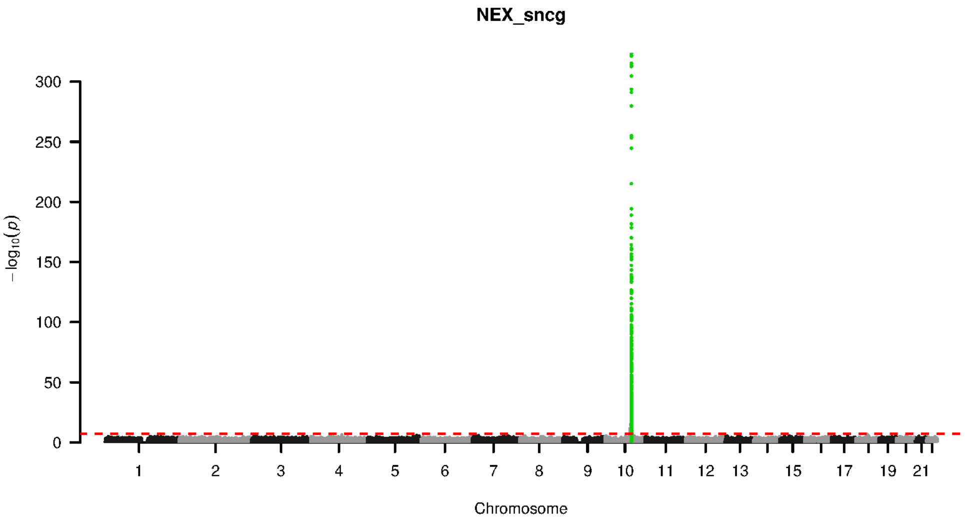


**Supplementary Figure 9-176. Manhattan plot and pQTL associations of the genome-wide meta-analysis for protein SNCG.** The horizontal dashed line corresponds to the genome-wide significance threshold of $p = 5 \times 10^{-8}$. The variants in the cis-regulatory region (±500kb of the coding gene region) of the protein-coding gene are highlighted in green.

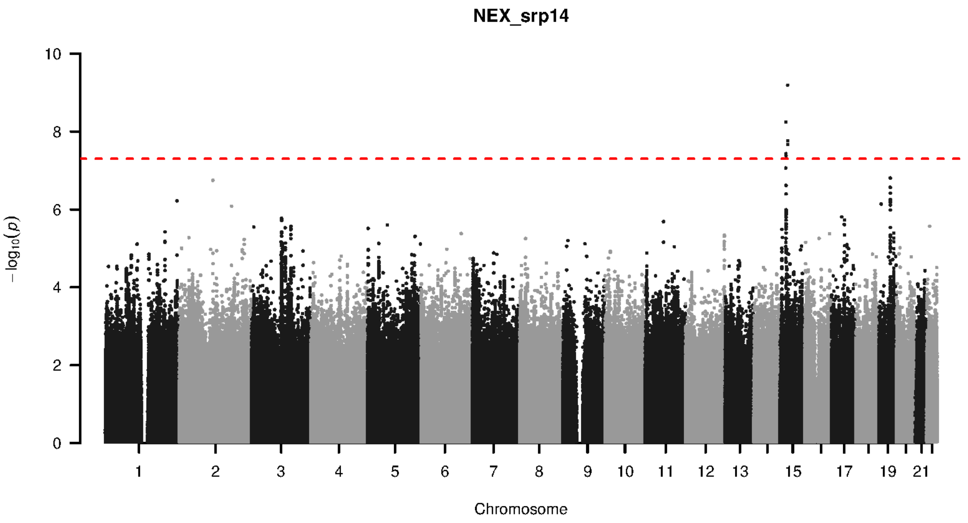


**Supplementary Figure 9-177. Manhattan plot and pQTL associations of the genome-wide meta-analysis for protein SRP14.** The horizontal dashed line corresponds to the genome-wide significance threshold of $p = 5 \times 10^{-8}$. The variants in the cis-regulatory region (±500kb of the coding gene region) of the protein-coding gene are highlighted in green.

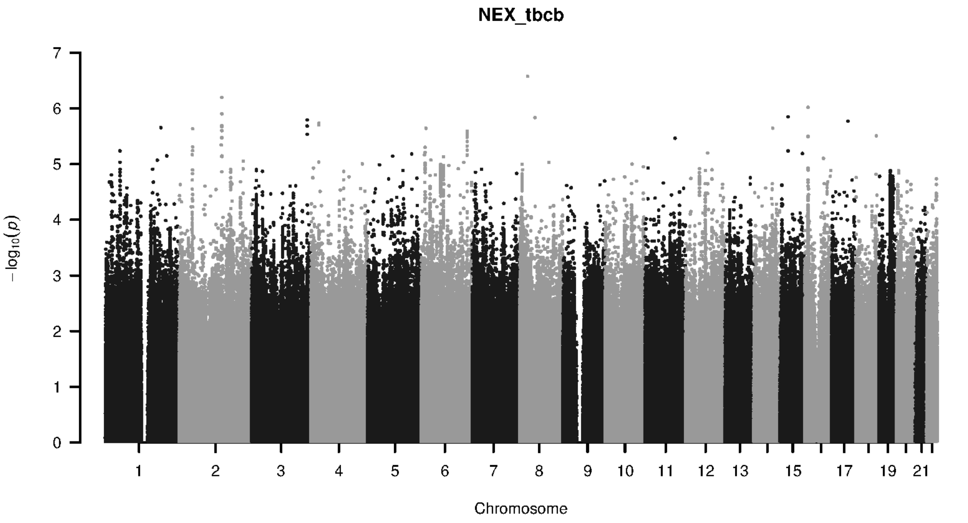


**Supplementary Figure 9-178. Manhattan plot and pQTL associations of the genome-wide meta-analysis for protein TBCB.** The horizontal dashed line corresponds to the genome-wide significance threshold of $p = 5 \times 10^{-8}$. The variants in the cis-regulatory region (±500kb of the coding gene region) of the protein-coding gene are highlighted in green.

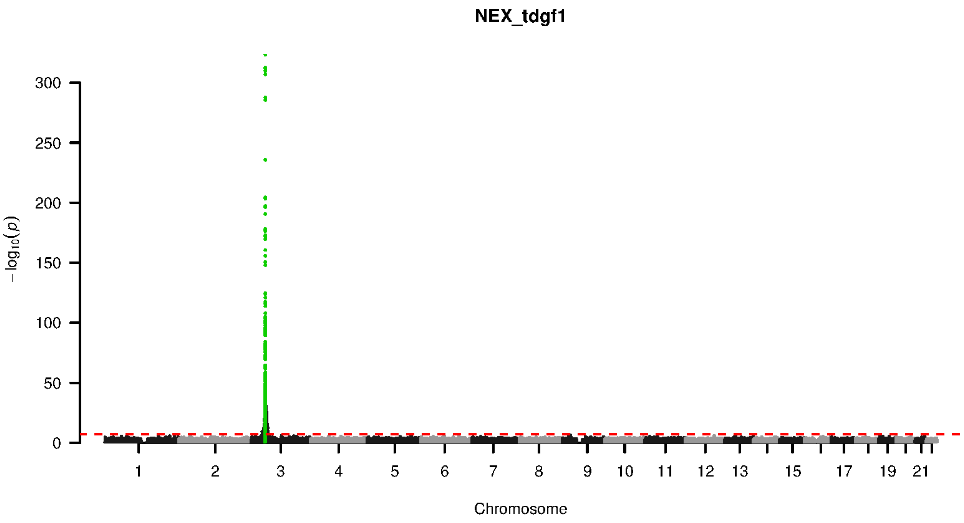


**Supplementary Figure 9-179. Manhattan plot and pQTL associations of the genome-wide meta-analysis for protein TDGF1.** The horizontal dashed line corresponds to the genome-wide significance threshold of $p = 5 \times 10^{-8}$. The variants in the cis-regulatory region (±500kb of the coding gene region) of the protein-coding gene are highlighted in green.

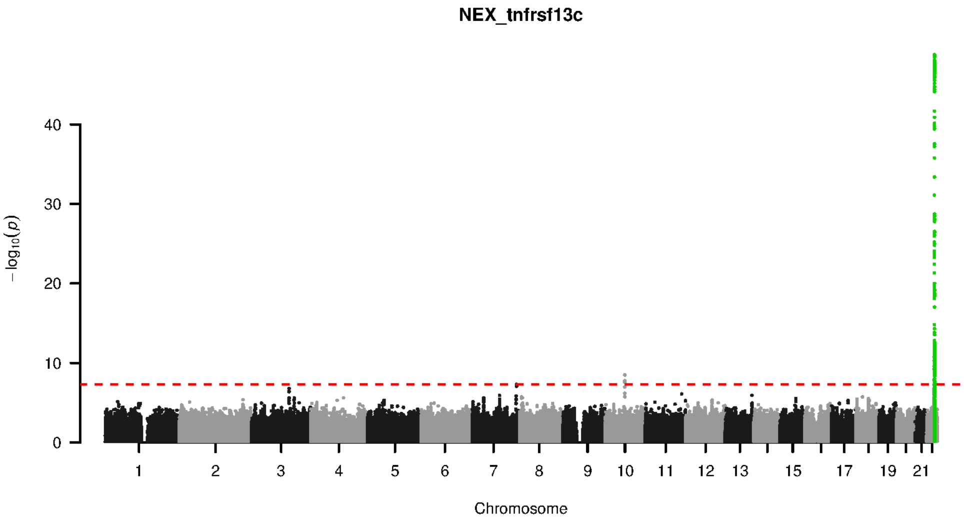


**Supplementary Figure 9-180. Manhattan plot and pQTL associations of the genome-wide meta-analysis for protein TNFRSF13C.** The horizontal dashed line corresponds to the genome-wide significance threshold of $p = 5 \times 10^{-8}$. The variants in the cis-regulatory region (±500kb of the coding gene region) of the protein-coding gene are highlighted in green.

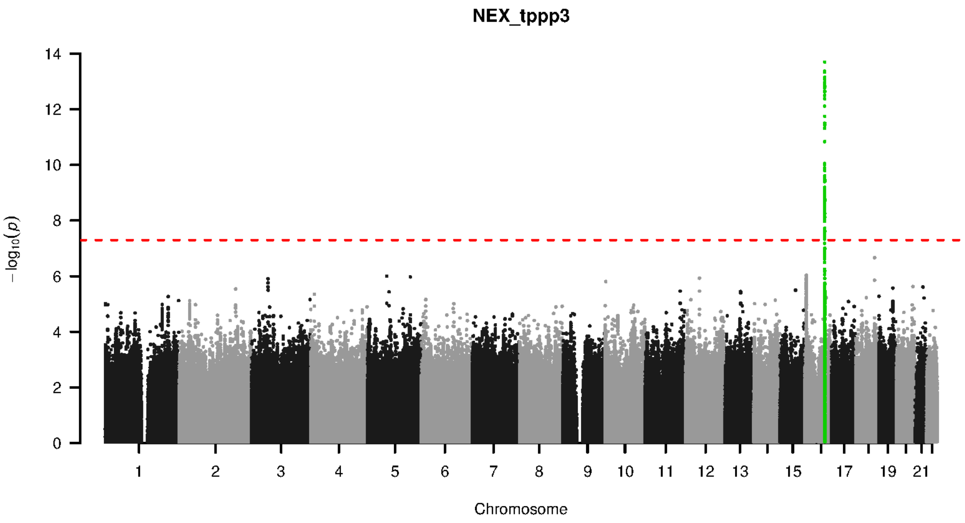


**Supplementary Figure 9-181. Manhattan plot and pQTL associations of the genome-wide meta-analysis for protein TPPP3.** The horizontal dashed line corresponds to the genome-wide significance threshold of $p = 5 \times 10^{-8}$. The variants in the cis-regulatory region (±500kb of the coding gene region) of the protein-coding gene are highlighted in green.

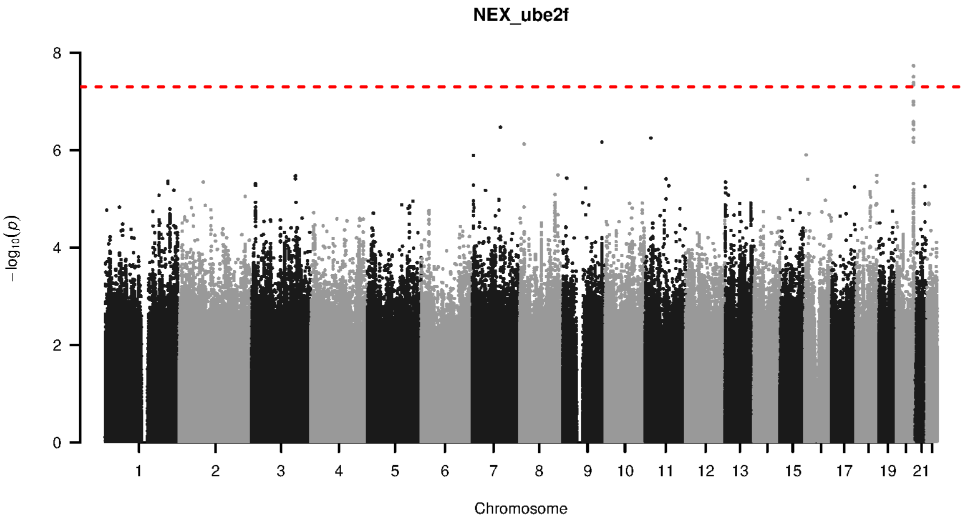


**Supplementary Figure 9-182. Manhattan plot and pQTL associations of the genome-wide meta-analysis for protein UBE2F.** The horizontal dashed line corresponds to the genome-wide significance threshold of $p = 5 \times 10^{-8}$. The variants in the cis-regulatory region (±500kb of the coding gene region) of the protein-coding gene are highlighted in green.

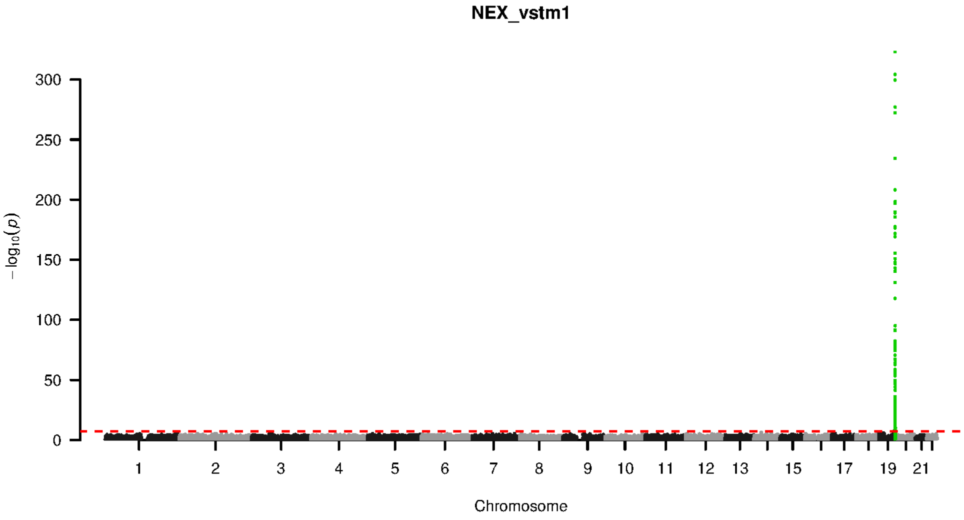


**Supplementary Figure 9-183. Manhattan plot and pQTL associations of the genome-wide meta-analysis for protein VSTM1.** The horizontal dashed line corresponds to the genome-wide significance threshold of $p = 5 \times 10^{-8}$. The variants in the cis-regulatory region (±500kb of the coding gene region) of the protein-coding gene are highlighted in green.

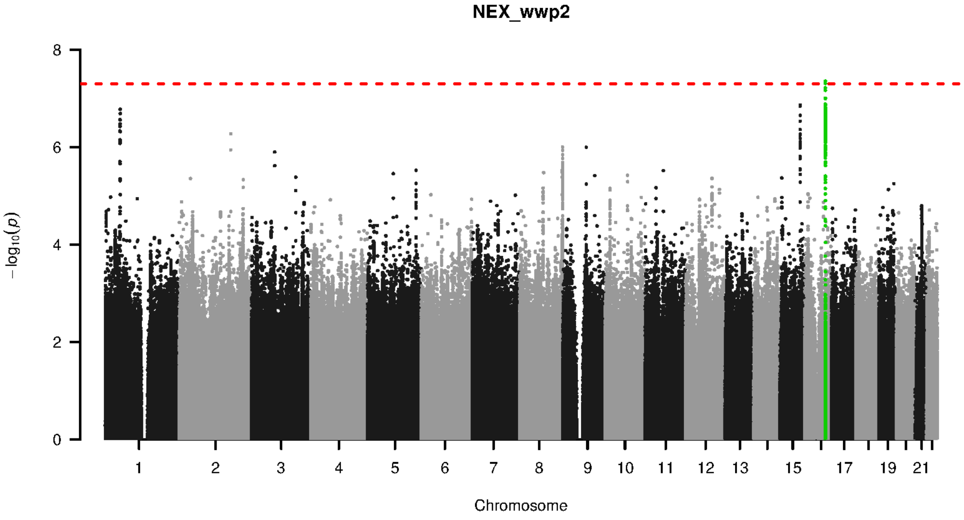


**Supplementary Figure 9-184. Manhattan plot and pQTL associations of the genome-wide meta-analysis for protein WWP2.** The horizontal dashed line corresponds to the genome-wide significance threshold of $p = 5 \times 10^{-8}$. The variants in the cis-regulatory region (±500kb of the coding gene region) of the protein-coding gene are highlighted in green.

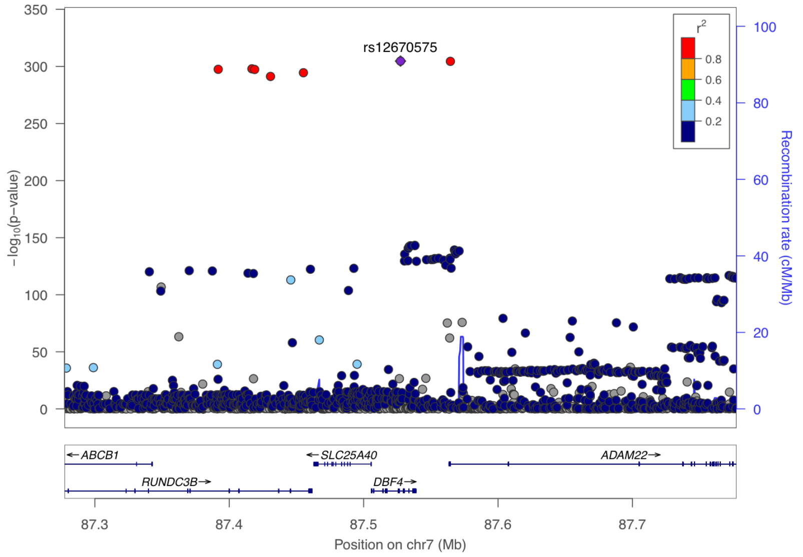


**Supplementary Figure 10-1. Visualization of the cis-pQTL association for protein ADAM 22.** The $r^2$ values are the squared linkage disequilibrium correlation coefficients between the plotted variants and the leading variant (purple dot). The middle panel marks the established associations at this locus according to the GWAS catalog ($p < 5 \times 10^{-8}$), and the bottom panel annotates the positions of the genes.

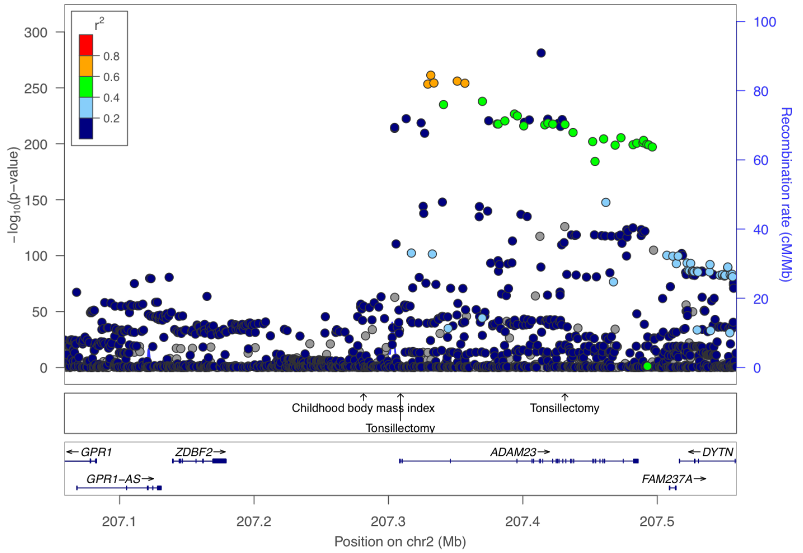


**Supplementary Figure 10-2. Visualization of the cis-pQTL association for protein ADAM 23.** The $r^2$ values are the squared linkage disequilibrium correlation coefficients between the plotted variants and the leading variant (purple dot). The middle panel marks the established associations at this locus according to the GWAS catalog ($p < 5 \times 10^{-8}$), and the bottom panel annotates the positions of the genes.

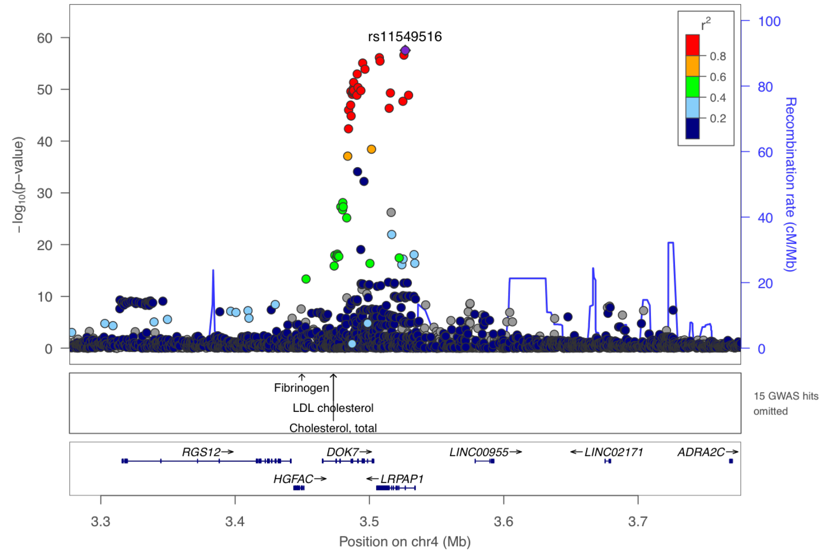


**Supplementary Figure 10-3. Visualization of the cis-pQTL association for protein ALPHA 2 MRAP.** The $r^2$ values are the squared linkage disequilibrium correlation coefficients between the plotted variants and the leading variant (purple dot). The middle panel marks the established associations at this locus according to the GWAS catalog ($p < 5 \times 10^{-8}$), and the bottom panel annotates the positions of the genes.

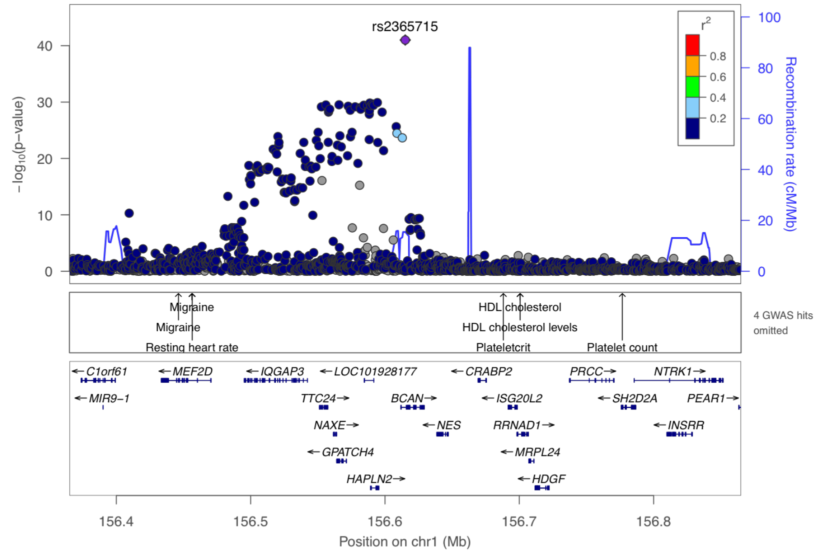


**Supplementary Figure 10-4. Visualization of the cis-pQTL association for protein BCAN.** The $r^2$ values are the squared linkage disequilibrium correlation coefficients between the plotted variants and the leading variant (purple dot). The middle panel marks the established associations at this locus according to the GWAS catalog ($p < 5 \times 10^{-8}$), and the bottom panel annotates the positions of the genes.

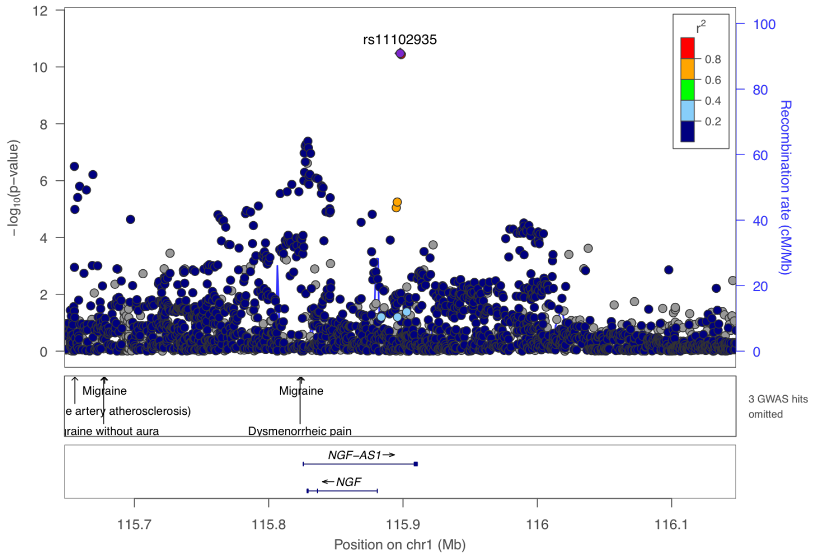


**Supplementary Figure 10-5. Visualization of the cis-pQTL association for protein BETA NGF.** The $r^2$ values are the squared linkage disequilibrium correlation coefficients between the plotted variants and the leading variant (purple dot). The middle panel marks the established associations at this locus according to the GWAS catalog ($p < 5 \times 10^{-8}$), and the bottom panel annotates the positions of the genes.

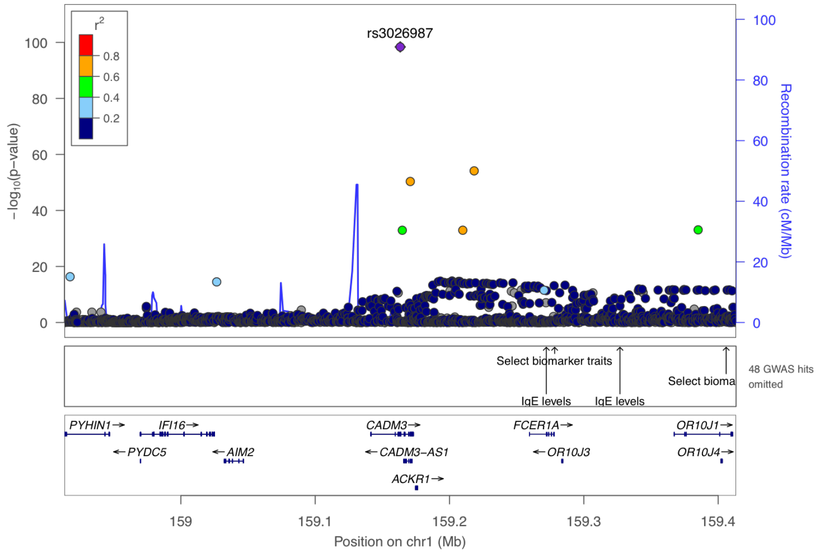


**Supplementary Figure 10-6. Visualization of the cis-pQTL association for protein CADM3.** The $r^2$ values are the squared linkage disequilibrium correlation coefficients between the plotted variants and the leading variant (purple dot). The middle panel marks the established associations at this locus according to the GWAS catalog ($p < 5 \times 10^{-8}$), and the bottom panel annotates the positions of the genes.

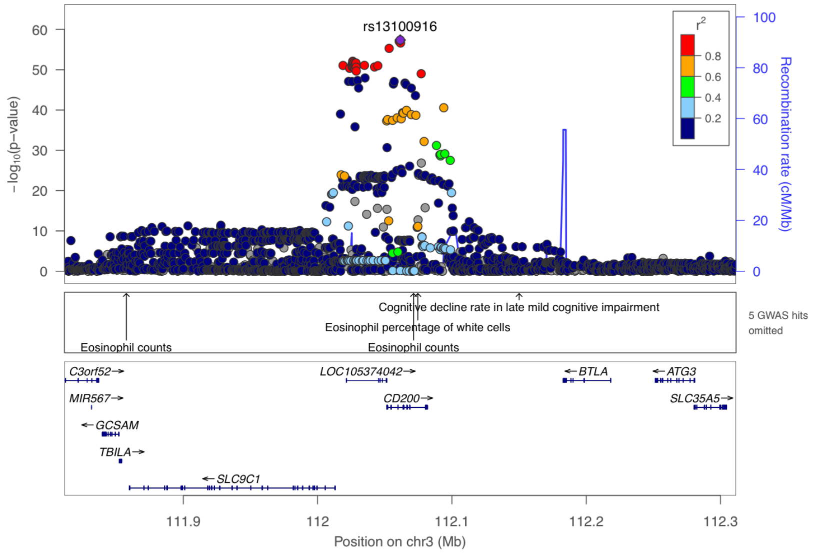


**Supplementary Figure 10-7. Visualization of the cis-pQTL association for protein CD200.** The $r^2$ values are the squared linkage disequilibrium correlation coefficients between the plotted variants and the leading variant (purple dot). The middle panel marks the established associations at this locus according to the GWAS catalog ($p < 5 \times 10^{-8}$), and the bottom panel annotates the positions of the genes.

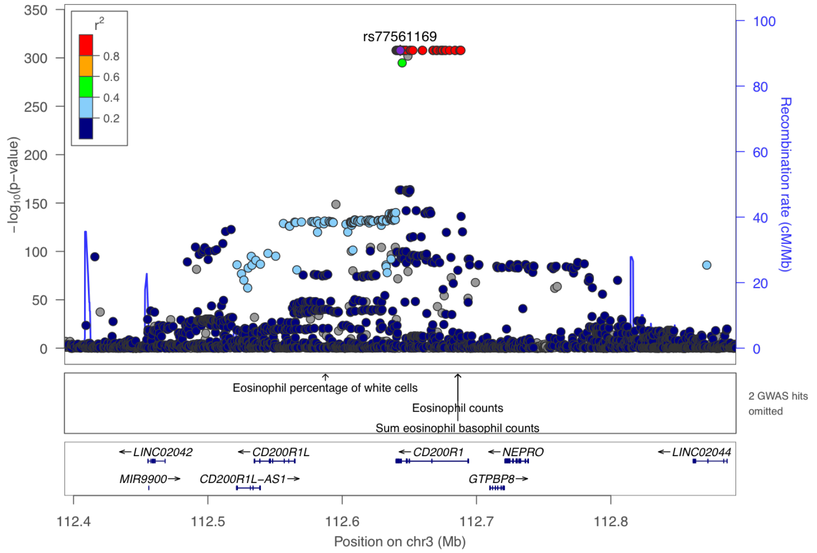


**Supplementary Figure 10-8. Visualization of the cis-pQTL association for protein CD200R1.** The $r^2$ values are the squared linkage disequilibrium correlation coefficients between the plotted variants and the leading variant (purple dot). The middle panel marks the established associations at this locus according to the GWAS catalog ($p < 5 \times 10^{-8}$), and the bottom panel annotates the positions of the genes.

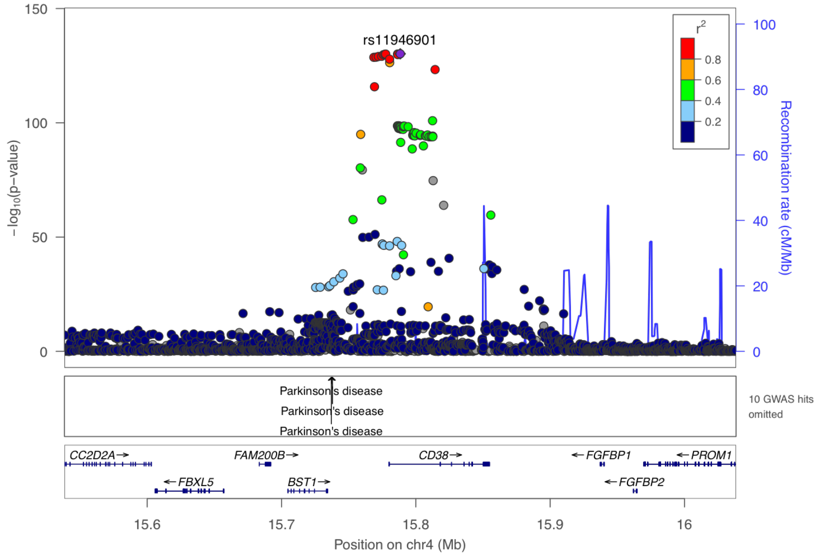


**Supplementary Figure 10-9. Visualization of the cis-pQTL association for protein CD38.** The $r^2$ values are the squared linkage disequilibrium correlation coefficients between the plotted variants and the leading variant (purple dot). The middle panel marks the established associations at this locus according to the GWAS catalog ($p < 5 \times 10^{-8}$), and the bottom panel annotates the positions of the genes.

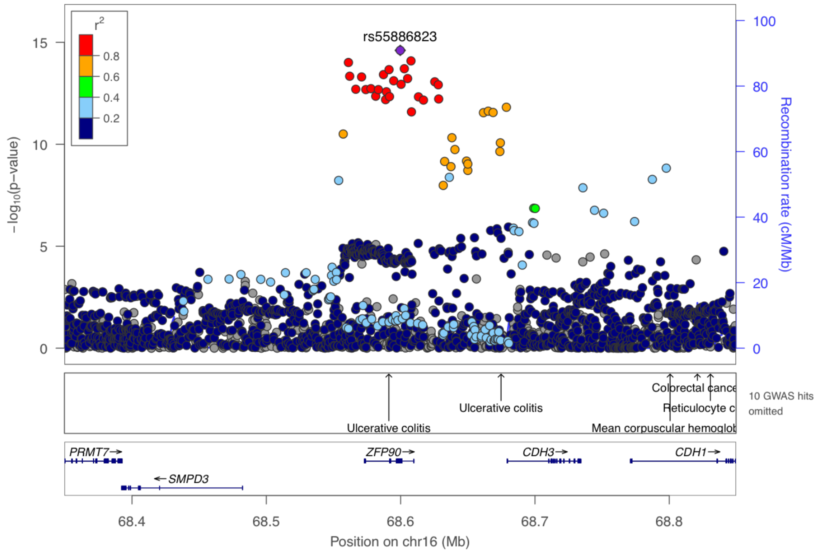


**Supplementary Figure 10-10. Visualization of the cis-pQTL association for protein CDH3.** The $r^2$ values are the squared linkage disequilibrium correlation coefficients between the plotted variants and the leading variant (purple dot). The middle panel marks the established associations at this locus according to the GWAS catalog ($p < 5 \times 10^{-8}$), and the bottom panel annotates the positions of the genes.

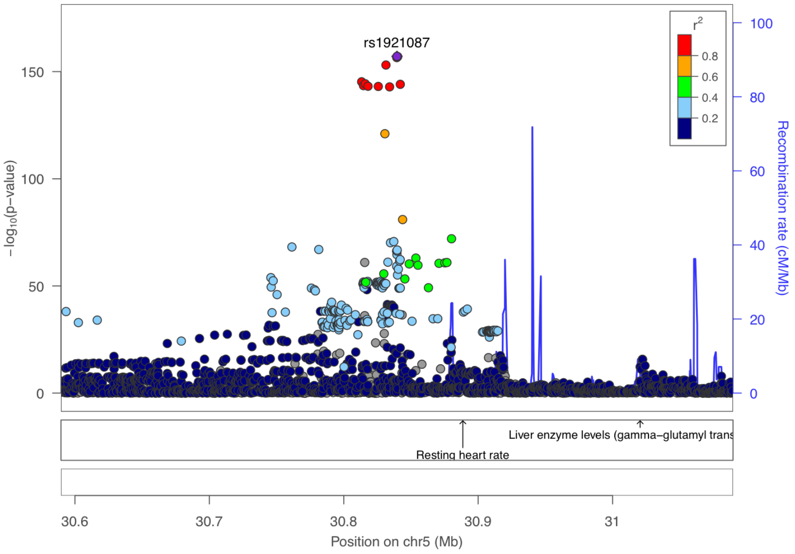


**Supplementary Figure 10-11. Visualization of the cis-pQTL association for protein CDH6.** The $r^2$ values are the squared linkage disequilibrium correlation coefficients between the plotted variants and the leading variant (purple dot). The middle panel marks the established associations at this locus according to the GWAS catalog ($p < 5 \times 10^{-8}$), and the bottom panel annotates the positions of the genes.

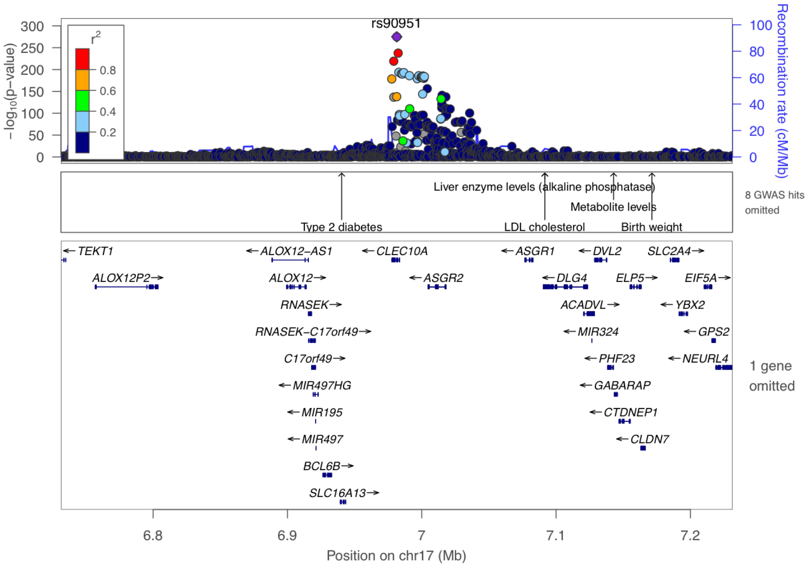


**Supplementary Figure 10-12. Visualization of the cis-pQTL association for protein CLEC10A.** The $r^2$ values are the squared linkage disequilibrium correlation coefficients between the plotted variants and the leading variant (purple dot). The middle panel marks the established associations at this locus according to the GWAS catalog ($p < 5 \times 10^{-8}$), and the bottom panel annotates the positions of the genes.

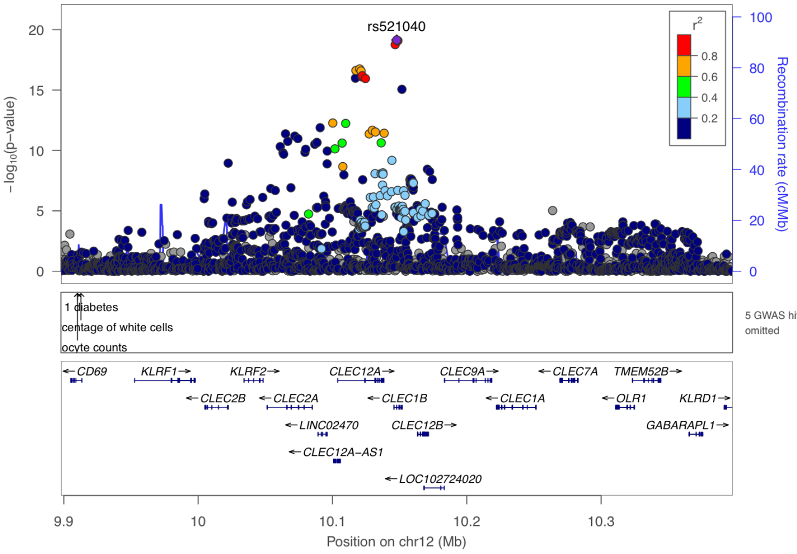


**Supplementary Figure 10-13. Visualization of the cis-pQTL association for protein CLEC1B.** The $r^2$ values are the squared linkage disequilibrium correlation coefficients between the plotted variants and the leading variant (purple dot). The middle panel marks the established associations at this locus according to the GWAS catalog ($p < 5 \times 10^{-8}$), and the bottom panel annotates the positions of the genes.

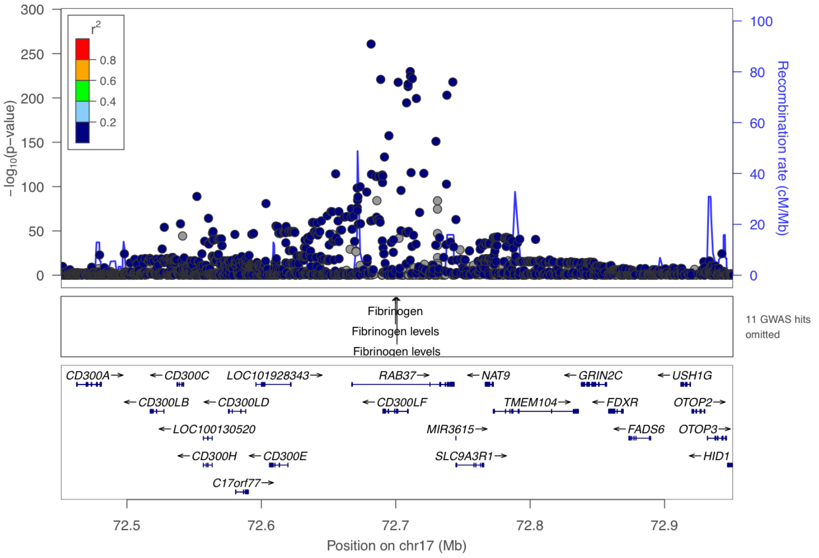


**Supplementary Figure 10-14. Visualization of the cis-pQTL association for protein CLM 1.** The $r^2$ values are the squared linkage disequilibrium correlation coefficients between the plotted variants and the leading variant (purple dot). The middle panel marks the established associations at this locus according to the GWAS catalog ($p < 5 \times 10^{-8}$), and the bottom panel annotates the positions of the genes.

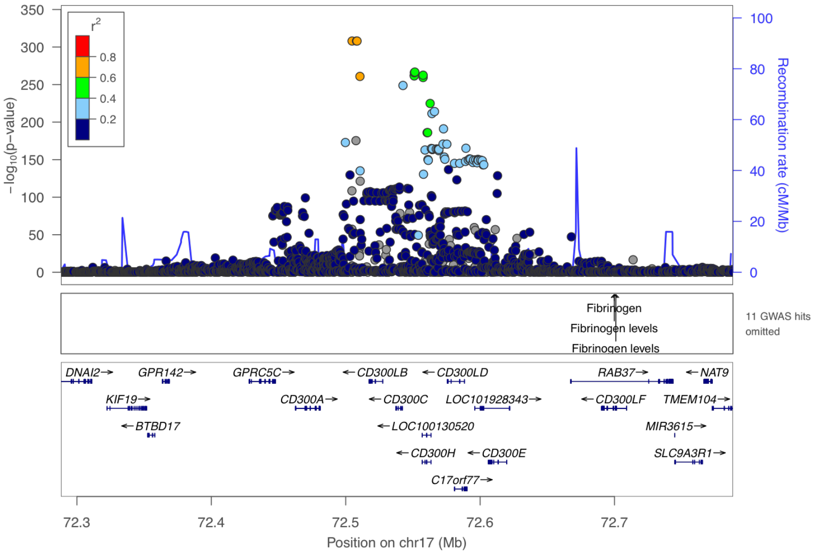


**Supplementary Figure 10-15. Visualization of the cis-pQTL association for protein CLM 6.** The $r^2$ values are the squared linkage disequilibrium correlation coefficients between the plotted variants and the leading variant (purple dot). The middle panel marks the established associations at this locus according to the GWAS catalog ($p < 5 \times 10^{-8}$), and the bottom panel annotates the positions of the genes.

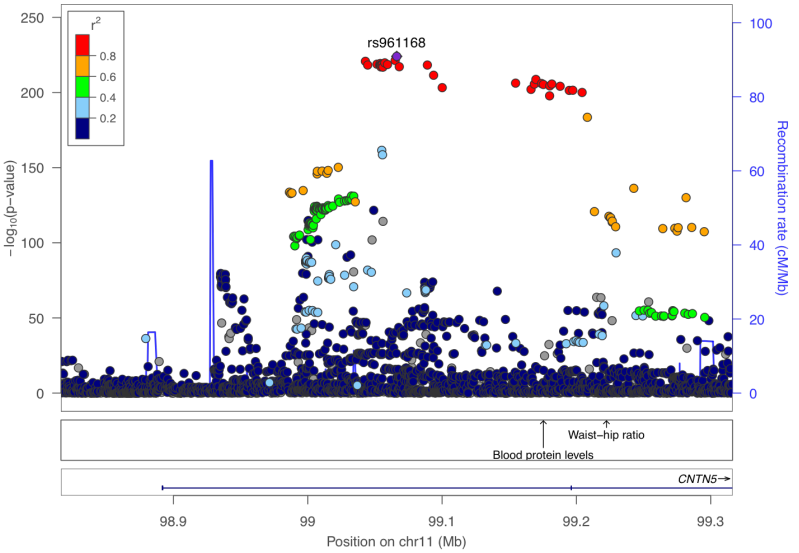


**Supplementary Figure 10-16. Visualization of the cis-pQTL association for protein CNTN5.** The $r^2$ values are the squared linkage disequilibrium correlation coefficients between the plotted variants and the leading variant (purple dot). The middle panel marks the established associations at this locus according to the GWAS catalog ($p < 5 \times 10^{-8}$), and the bottom panel annotates the positions of the genes.

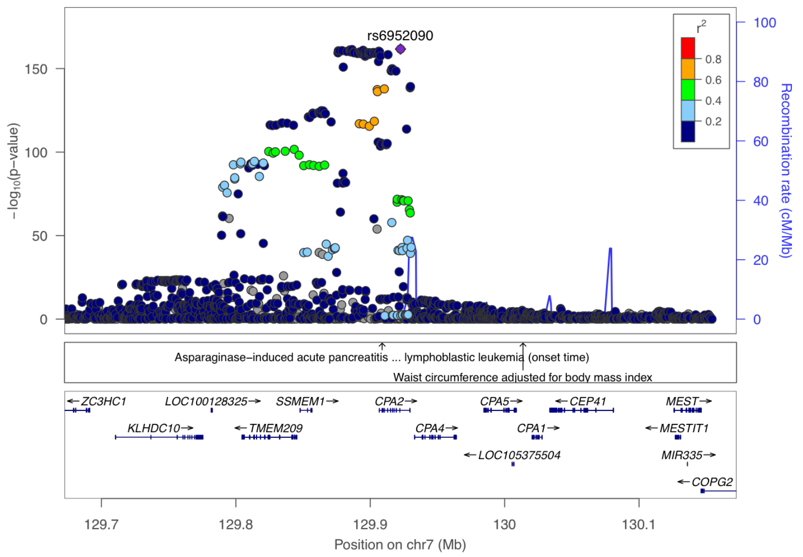


**Supplementary Figure 10-17. Visualization of the cis-pQTL association for protein CPA2.** The $r^2$ values are the squared linkage disequilibrium correlation coefficients between the plotted variants and the leading variant (purple dot). The middle panel marks the established associations at this locus according to the GWAS catalog ($p < 5 \times 10^{-8}$), and the bottom panel annotates the positions of the genes.

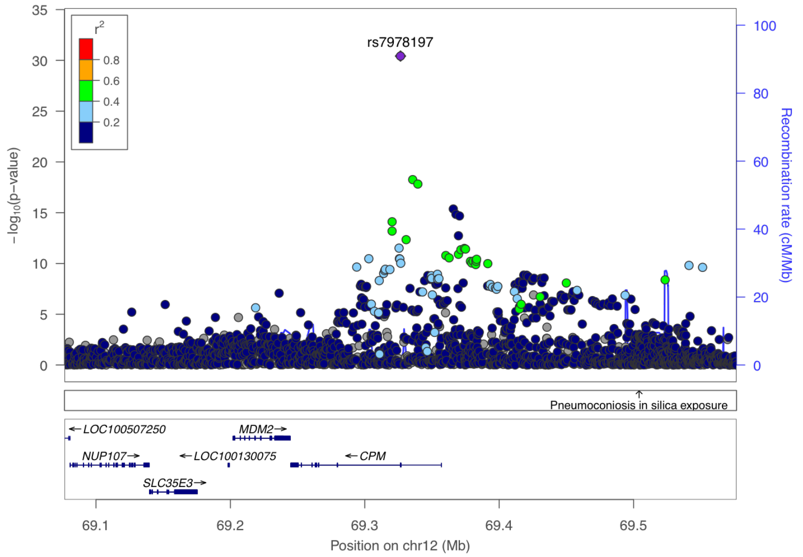


**Supplementary Figure 10-18. Visualization of the cis-pQTL association for protein CPM.** The $r^2$ values are the squared linkage disequilibrium correlation coefficients between the plotted variants and the leading variant (purple dot). The middle panel marks the established associations at this locus according to the GWAS catalog ($p < 5 \times 10^{-8}$), and the bottom panel annotates the positions of the genes.

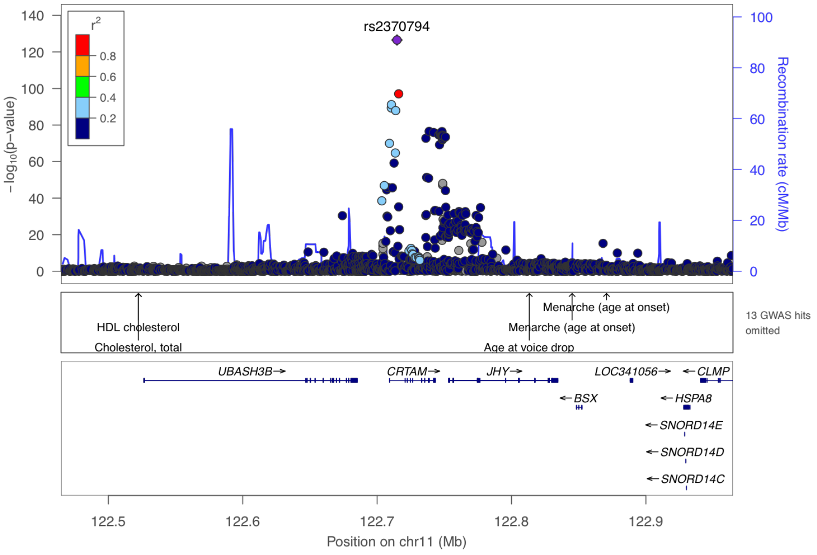


**Supplementary Figure 10-19. Visualization of the cis-pQTL association for protein CRTAM.** The $r^2$ values are the squared linkage disequilibrium correlation coefficients between the plotted variants and the leading variant (purple dot). The middle panel marks the established associations at this locus according to the GWAS catalog ($p < 5 \times 10^{-8}$), and the bottom panel annotates the positions of the genes.

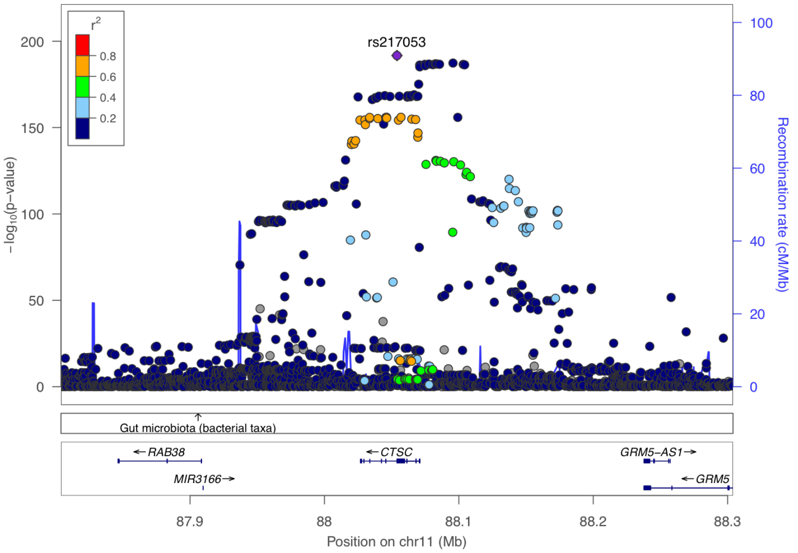


**Supplementary Figure 10-20. Visualization of the cis-pQTL association for protein CTSC.** The $r^2$ values are the squared linkage disequilibrium correlation coefficients between the plotted variants and the leading variant (purple dot). The middle panel marks the established associations at this locus according to the GWAS catalog ($p < 5 \times 10^{-8}$), and the bottom panel annotates the positions of the genes.

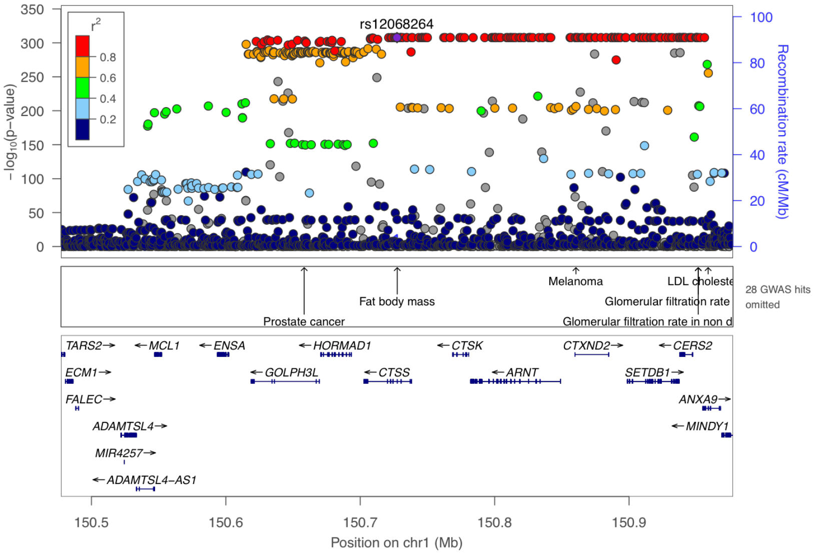


**Supplementary Figure 10-21. Visualization of the cis-pQTL association for protein CTSS.** The $r^2$ values are the squared linkage disequilibrium correlation coefficients between the plotted variants and the leading variant (purple dot). The middle panel marks the established associations at this locus according to the GWAS catalog ($p < 5 \times 10^{-8}$), and the bottom panel annotates the positions of the genes.

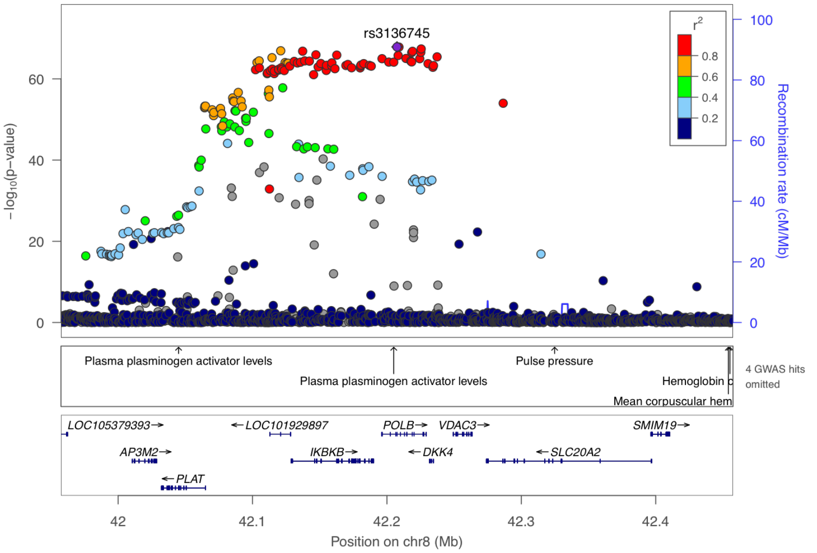


**Supplementary Figure 10-22. Visualization of the cis-pQTL association for protein DKK 4.** The $r^2$ values are the squared linkage disequilibrium correlation coefficients between the plotted variants and the leading variant (purple dot). The middle panel marks the established associations at this locus according to the GWAS catalog ($p < 5 \times 10^{-8}$), and the bottom panel annotates the positions of the genes.

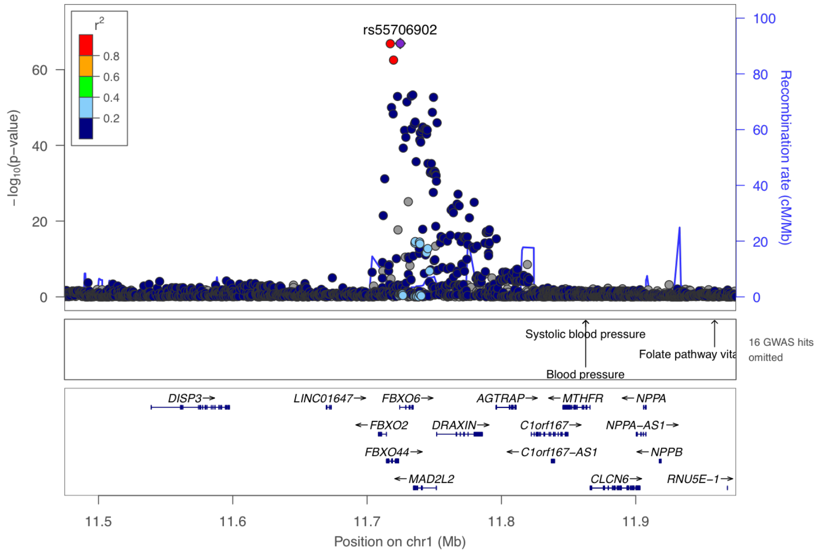


**Supplementary Figure 10-23. Visualization of the cis-pQTL association for protein DRAXIN.** The $r^2$ values are the squared linkage disequilibrium correlation coefficients between the plotted variants and the leading variant (purple dot). The middle panel marks the established associations at this locus according to the GWAS catalog ($p < 5 \times 10^{-8}$), and the bottom panel annotates the positions of the genes.

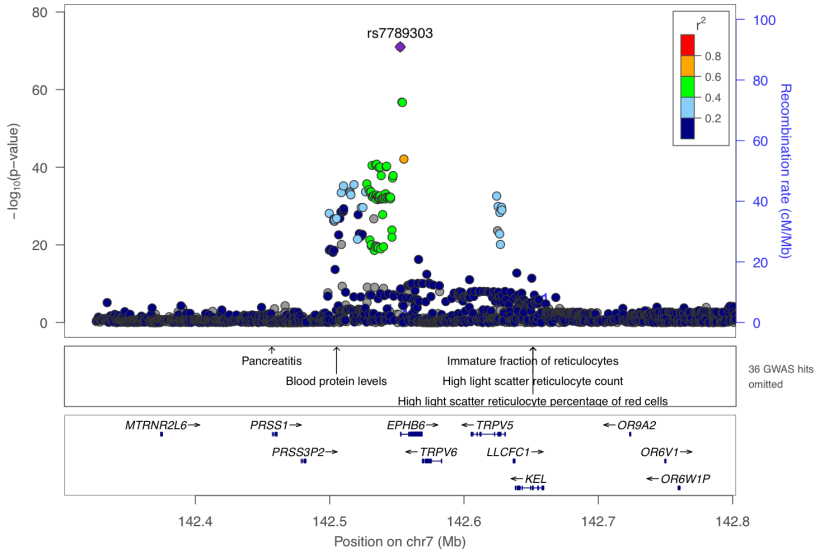


**Supplementary Figure 10-24. Visualization of the cis-pQTL association for protein EPHB6.** The $r^2$ values are the squared linkage disequilibrium correlation coefficients between the plotted variants and the leading variant (purple dot). The middle panel marks the established associations at this locus according to the GWAS catalog ($p < 5 \times 10^{-8}$), and the bottom panel annotates the positions of the genes.

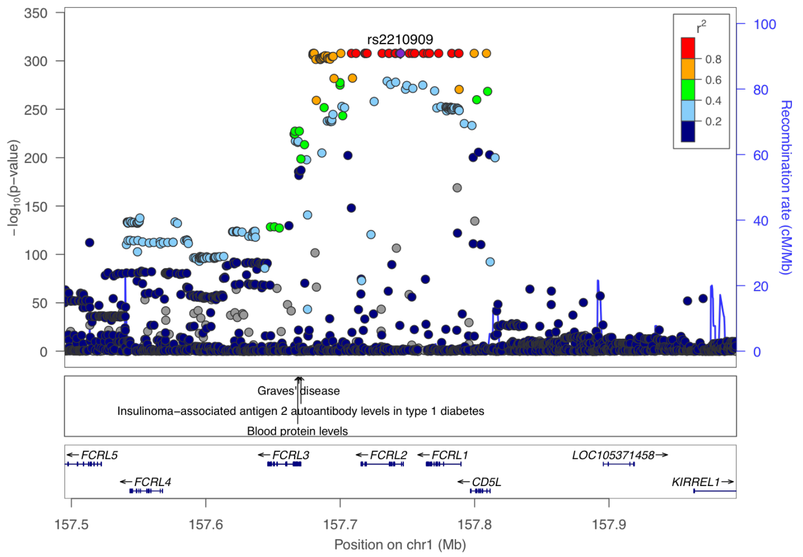


**Supplementary Figure 10-25. Visualization of the cis-pQTL association for protein FCRL2.** The $r^2$ values are the squared linkage disequilibrium correlation coefficients between the plotted variants and the leading variant (purple dot). The middle panel marks the established associations at this locus according to the GWAS catalog ($p < 5 \times 10^{-8}$), and the bottom panel annotates the positions of the genes.

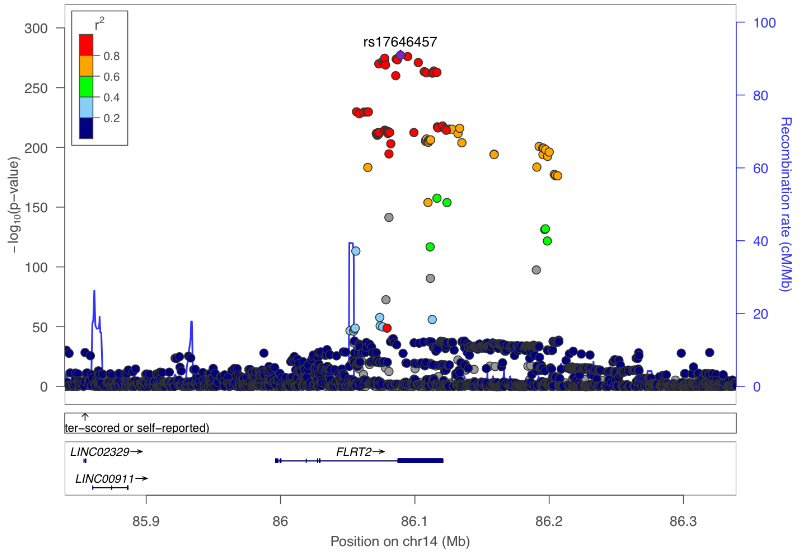


**Supplementary Figure 10-26. Visualization of the cis-pQTL association for protein FLRT2.** The $r^2$ values are the squared linkage disequilibrium correlation coefficients between the plotted variants and the leading variant (purple dot). The middle panel marks the established associations at this locus according to the GWAS catalog ($p < 5 \times 10^{-8}$), and the bottom panel annotates the positions of the genes.

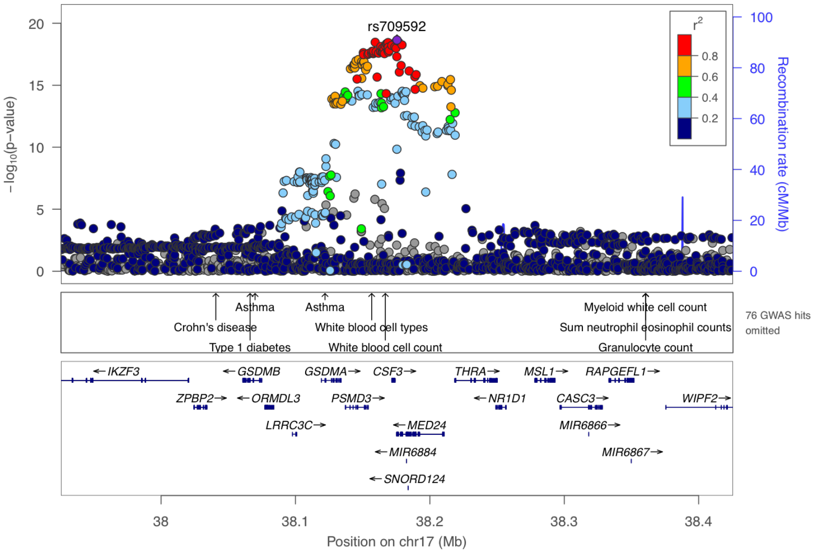


**Supplementary Figure 10-27. Visualization of the cis-pQTL association for protein G CSF.** The $r^2$ values are the squared linkage disequilibrium correlation coefficients between the plotted variants and the leading variant (purple dot). The middle panel marks the established associations at this locus according to the GWAS catalog ($p < 5 \times 10^{-8}$), and the bottom panel annotates the positions of the genes.

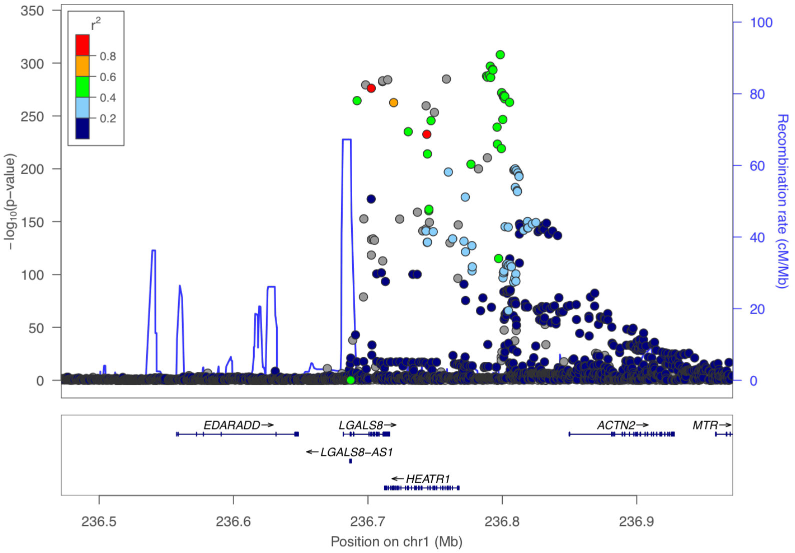


**Supplementary Figure 10-28. Visualization of the cis-pQTL association for protein GAL 8.** The $r^2$ values are the squared linkage disequilibrium correlation coefficients between the plotted variants and the leading variant (purple dot). The middle panel marks the established associations at this locus according to the GWAS catalog ($p < 5 \times 10^{-8}$), and the bottom panel annotates the positions of the genes.

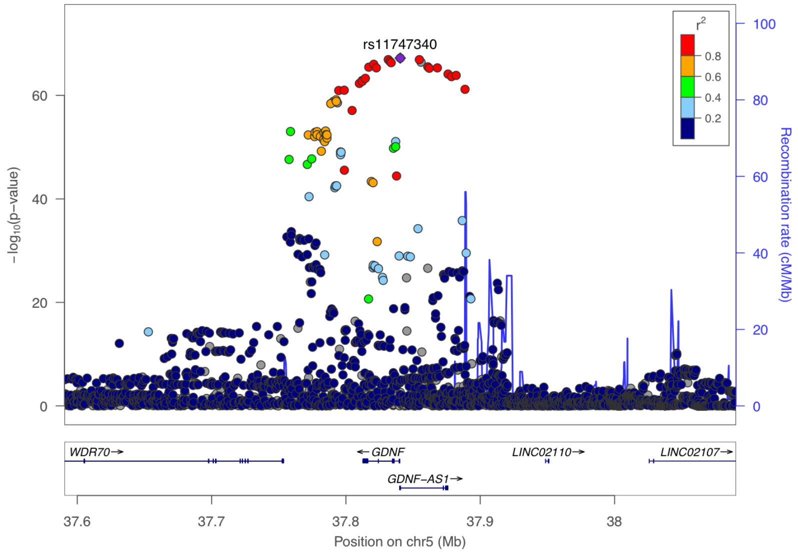


**Supplementary Figure 10-29. Visualization of the cis-pQTL association for protein GDNF.** The $r^2$ values are the squared linkage disequilibrium correlation coefficients between the plotted variants and the leading variant (purple dot). The middle panel marks the established associations at this locus according to the GWAS catalog ($p < 5 \times 10^{-8}$), and the bottom panel annotates the positions of the genes.

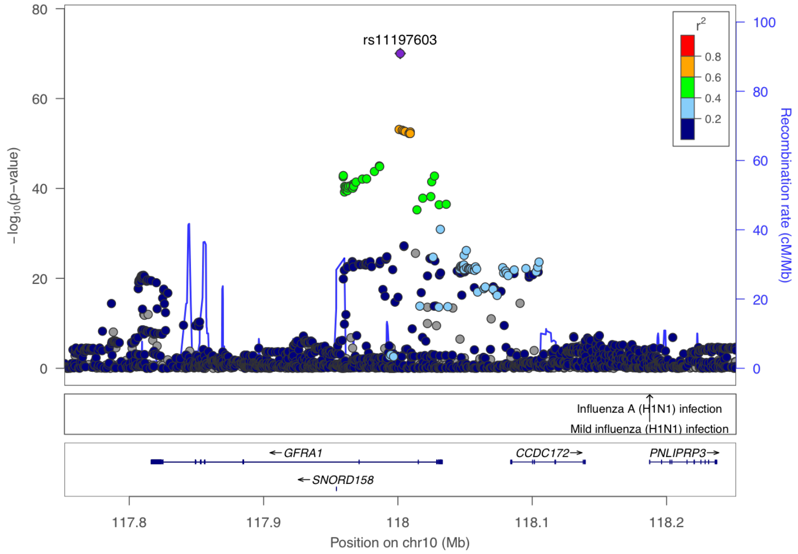


**Supplementary Figure 10-30. Visualization of the cis-pQTL association for protein GFR ALPHA 1.** The $r^2$ values are the squared linkage disequilibrium correlation coefficients between the plotted variants and the leading variant (purple dot). The middle panel marks the established associations at this locus according to the GWAS catalog ($p < 5 \times 10^{-8}$), and the bottom panel annotates the positions of the genes.

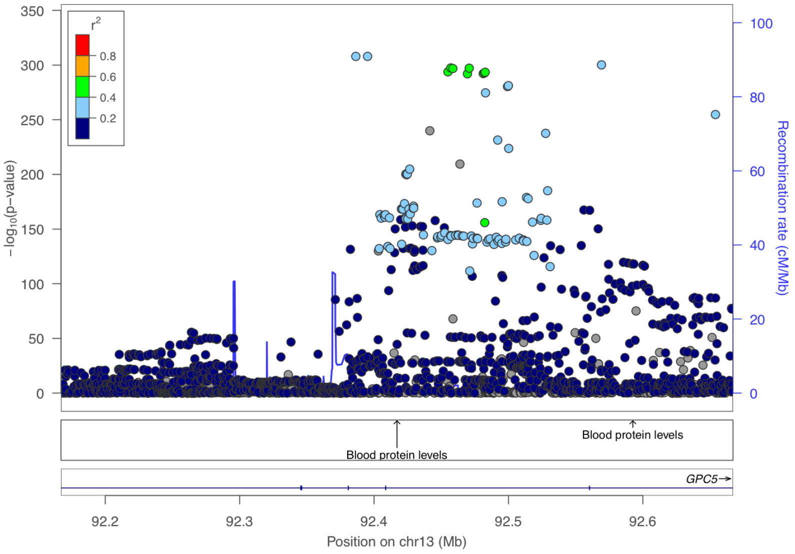


**Supplementary Figure 10-31. Visualization of the cis-pQTL association for protein GPC5.** The $r^2$ values are the squared linkage disequilibrium correlation coefficients between the plotted variants and the leading variant (purple dot). The middle panel marks the established associations at this locus according to the GWAS catalog ($p < 5 \times 10^{-8}$), and the bottom panel annotates the positions of the genes.

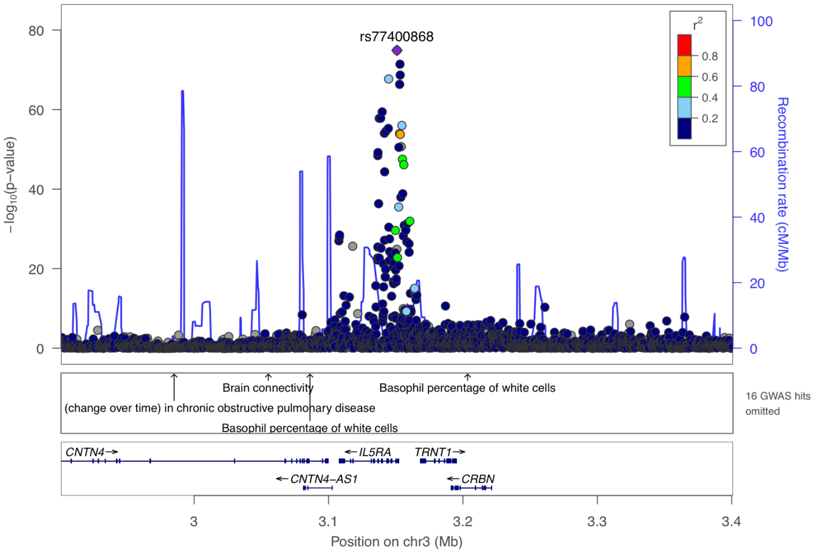


**Supplementary Figure 10-32. Visualization of the cis-pQTL association for protein IL 5R ALPHA.** The $r^2$ values are the squared linkage disequilibrium correlation coefficients between the plotted variants and the leading variant (purple dot). The middle panel marks the established associations at this locus according to the GWAS catalog ($p < 5 \times 10^{-8}$), and the bottom panel annotates the positions of the genes.

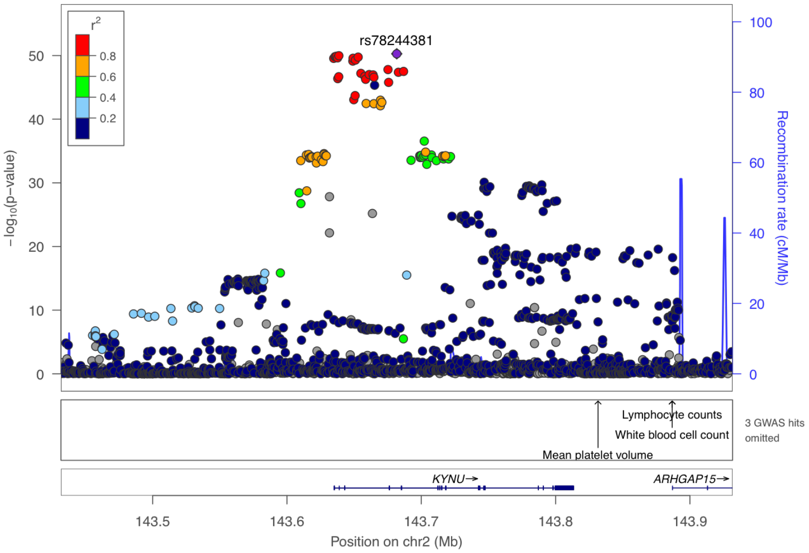


**Supplementary Figure 10-33. Visualization of the cis-pQTL association for protein KYNU.** The $r^2$ values are the squared linkage disequilibrium correlation coefficients between the plotted variants and the leading variant (purple dot). The middle panel marks the established associations at this locus according to the GWAS catalog ($p < 5 \times 10^{-8}$), and the bottom panel annotates the positions of the genes.

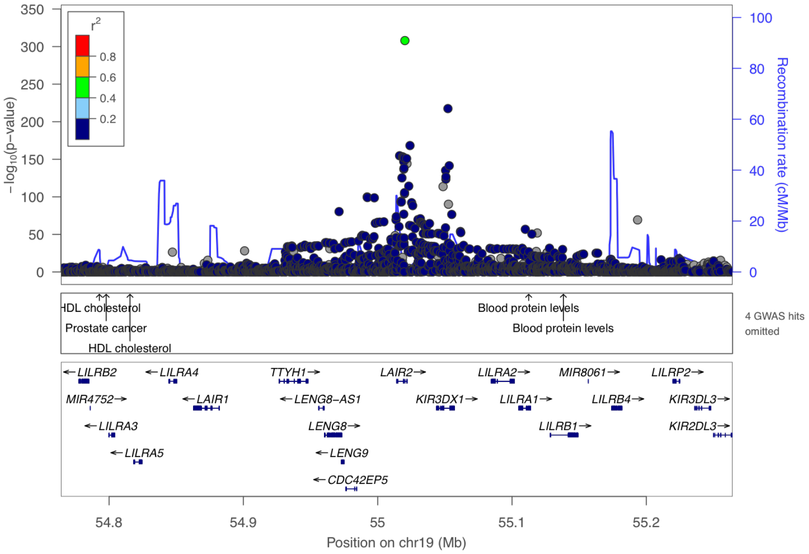


**Supplementary Figure 10-34. Visualization of the cis-pQTL association for protein LAIR 2.** The $r^2$ values are the squared linkage disequilibrium correlation coefficients between the plotted variants and the leading variant (purple dot). The middle panel marks the established associations at this locus according to the GWAS catalog ($p < 5 \times 10^{-8}$), and the bottom panel annotates the positions of the genes.

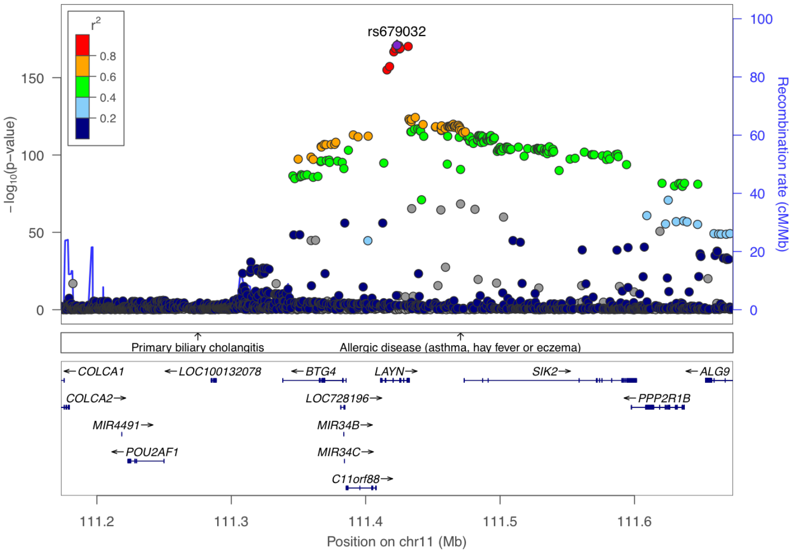


**Supplementary Figure 10-35. Visualization of the cis-pQTL association for protein LAYN.** The $r^2$ values are the squared linkage disequilibrium correlation coefficients between the plotted variants and the leading variant (purple dot). The middle panel marks the established associations at this locus according to the GWAS catalog ($p < 5 \times 10^{-8}$), and the bottom panel annotates the positions of the genes.

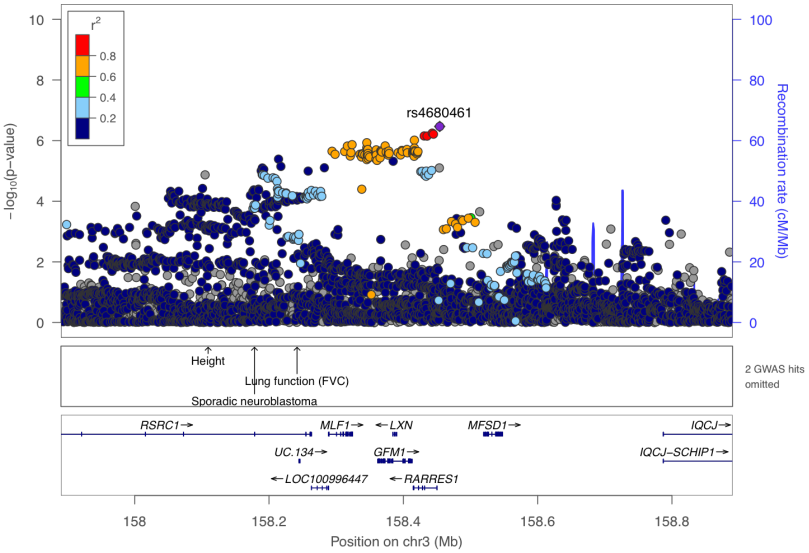


**Supplementary Figure 10-36. Visualization of the cis-pQTL association for protein LXN.** The $r^2$ values are the squared linkage disequilibrium correlation coefficients between the plotted variants and the leading variant (purple dot). The middle panel marks the established associations at this locus according to the GWAS catalog ($p < 5 \times 10^{-8}$), and the bottom panel annotates the positions of the genes.

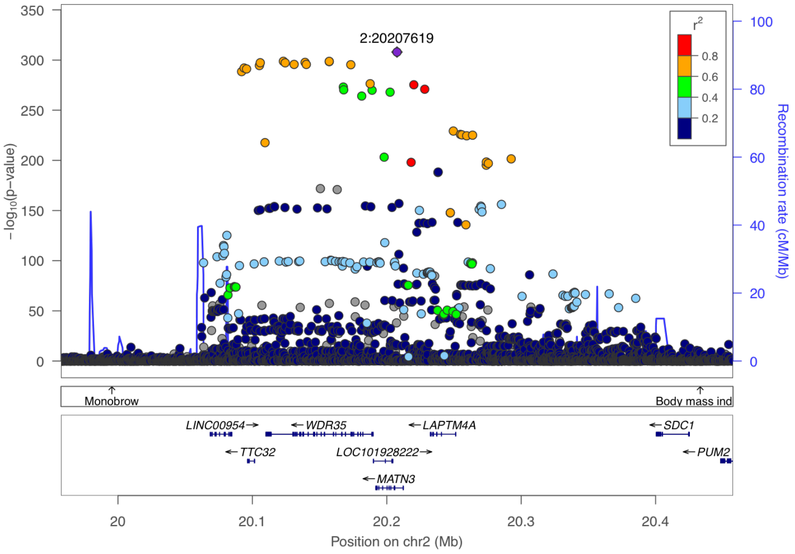


**Supplementary Figure 10-37. Visualization of the cis-pQTL association for protein MATN3.** The $r^2$ values are the squared linkage disequilibrium correlation coefficients between the plotted variants and the leading variant (purple dot). The middle panel marks the established associations at this locus according to the GWAS catalog ($p < 5 \times 10^{-8}$), and the bottom panel annotates the positions of the genes.

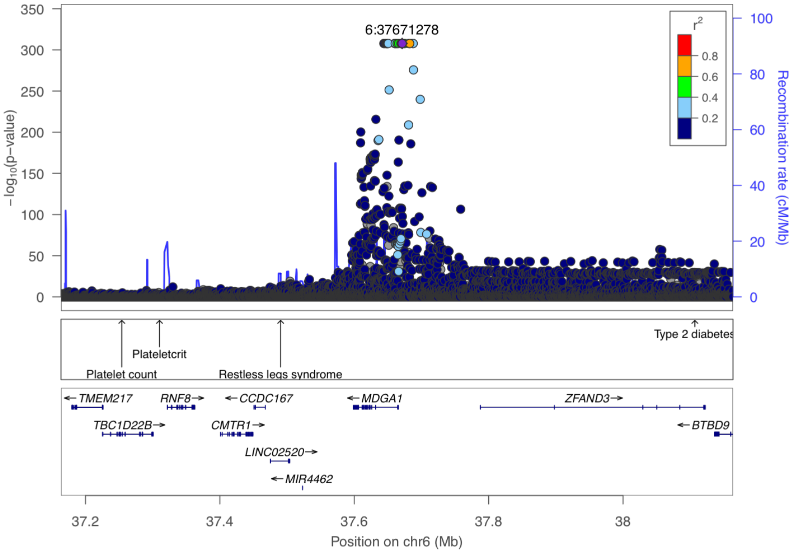


**Supplementary Figure 10-38. Visualization of the cis-pQTL association for protein MDGA1.** The $r^2$ values are the squared linkage disequilibrium correlation coefficients between the plotted variants and the leading variant (purple dot). The middle panel marks the established associations at this locus according to the GWAS catalog ($p < 5 \times 10^{-8}$), and the bottom panel annotates the positions of the genes.

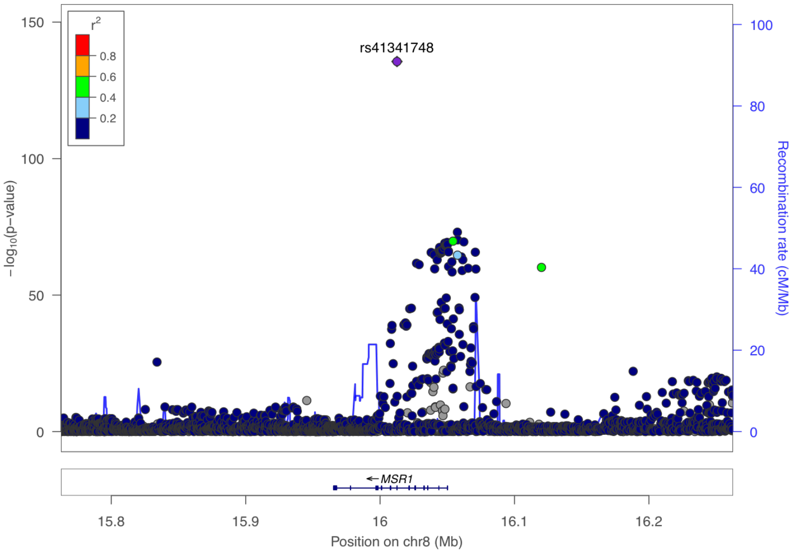


**Supplementary Figure 10-39. Visualization of the cis-pQTL association for protein MSR1.** The $r^2$ values are the squared linkage disequilibrium correlation coefficients between the plotted variants and the leading variant (purple dot). The middle panel marks the established associations at this locus according to the GWAS catalog ($p < 5 \times 10^{-8}$), and the bottom panel annotates the positions of the genes.

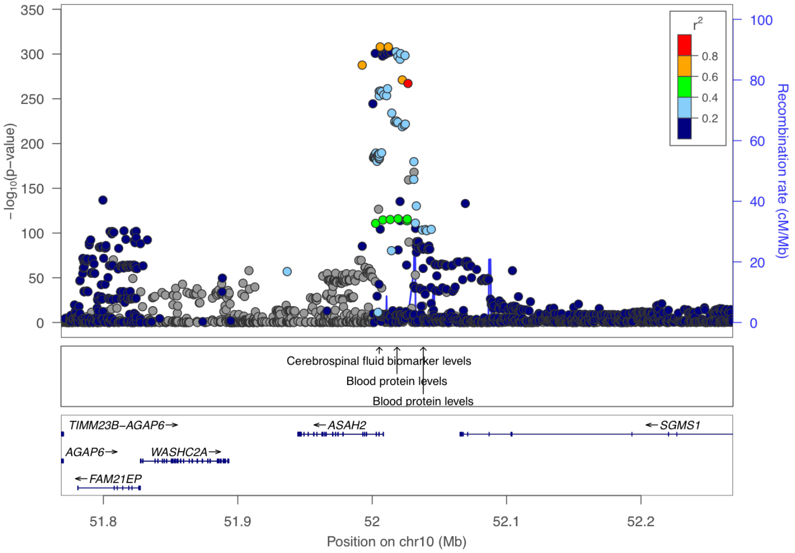


**Supplementary Figure 10-40. Visualization of the cis-pQTL association for protein N CDASE.** The $r^2$ values are the squared linkage disequilibrium correlation coefficients between the plotted variants and the leading variant (purple dot). The middle panel marks the established associations at this locus according to the GWAS catalog ($p < 5 \times 10^{-8}$), and the bottom panel annotates the positions of the genes.

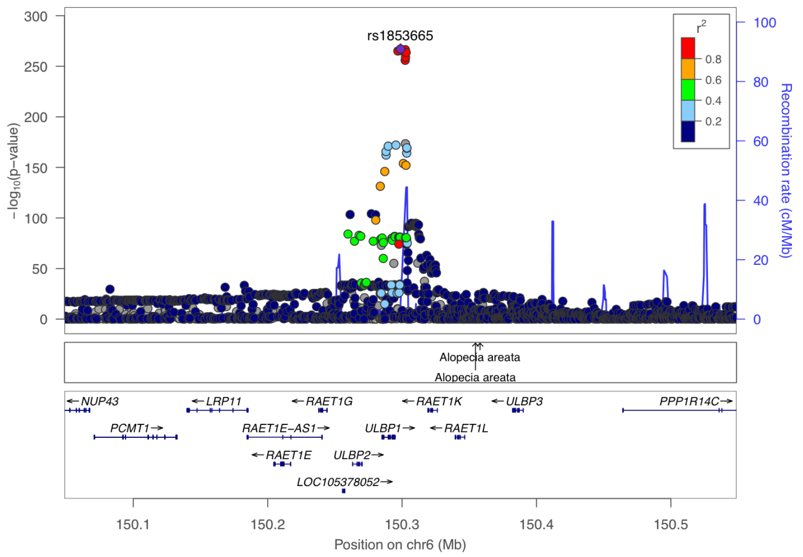


**Supplementary Figure 10-41. Visualization of the cis-pQTL association for protein N2DL 2.** The $r^2$ values are the squared linkage disequilibrium correlation coefficients between the plotted variants and the leading variant (purple dot). The middle panel marks the established associations at this locus according to the GWAS catalog ($p < 5 \times 10^{-8}$), and the bottom panel annotates the positions of the genes.

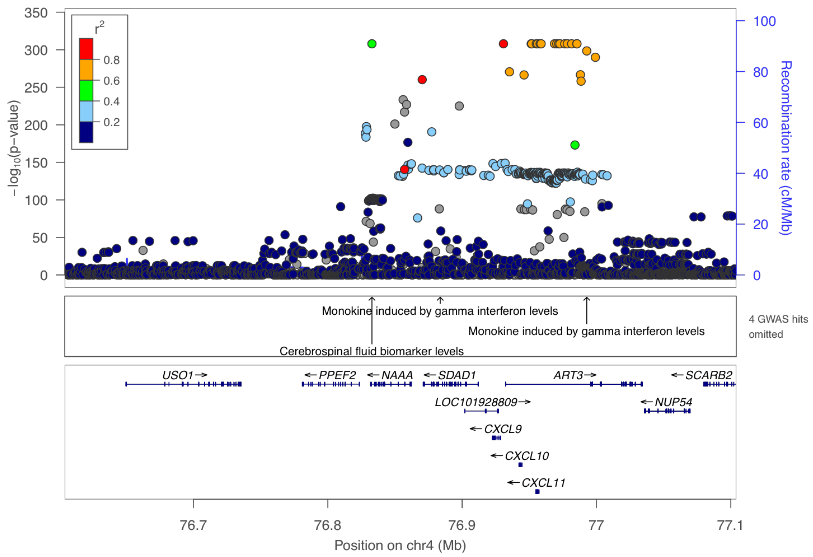


**Supplementary Figure 10-42. Visualization of the cis-pQTL association for protein NAAA.** The $r^2$ values are the squared linkage disequilibrium correlation coefficients between the plotted variants and the leading variant (purple dot). The middle panel marks the established associations at this locus according to the GWAS catalog ($p < 5 \times 10^{-8}$), and the bottom panel annotates the positions of the genes.

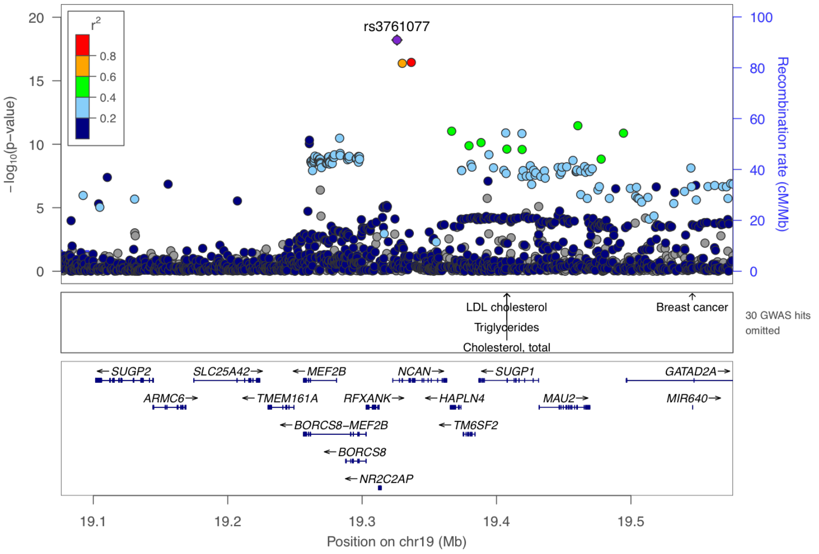


**Supplementary Figure 10-43. Visualization of the cis-pQTL association for protein NCAN.** The $r^2$ values are the squared linkage disequilibrium correlation coefficients between the plotted variants and the leading variant (purple dot). The middle panel marks the established associations at this locus according to the GWAS catalog ($p < 5 \times 10^{-8}$), and the bottom panel annotates the positions of the genes.

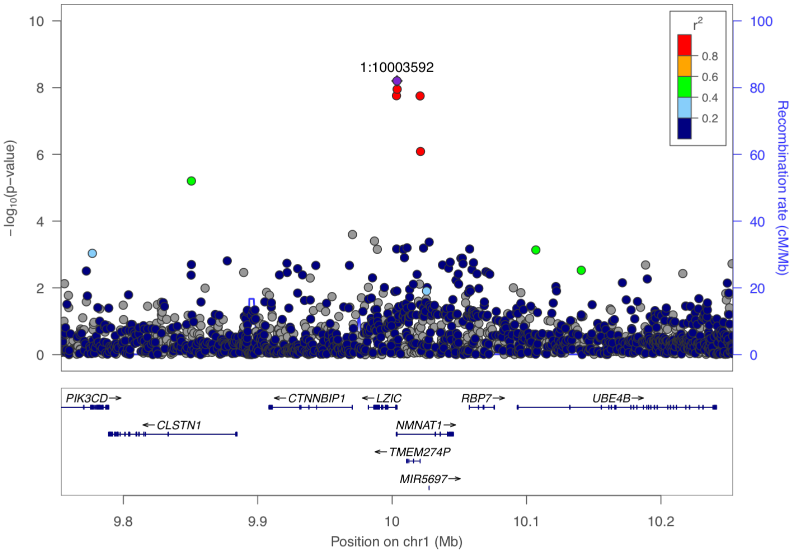


**Supplementary Figure 10-44. Visualization of the cis-pQTL association for protein NMNAT1.** The $r^2$ values are the squared linkage disequilibrium correlation coefficients between the plotted variants and the leading variant (purple dot). The middle panel marks the established associations at this locus according to the GWAS catalog ($p < 5 \times 10^{-8}$), and the bottom panel annotates the positions of the genes.

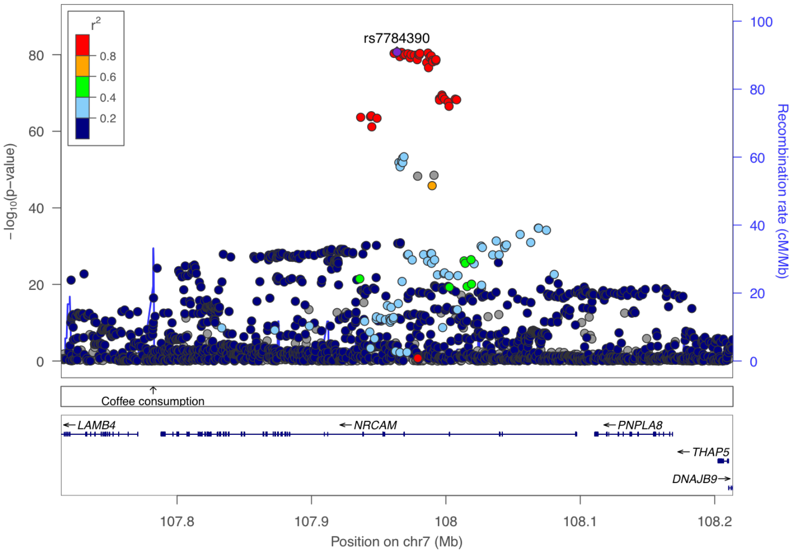


**Supplementary Figure 10-45. Visualization of the cis-pQTL association for protein NR CAM.** The $r^2$ values are the squared linkage disequilibrium correlation coefficients between the plotted variants and the leading variant (purple dot). The middle panel marks the established associations at this locus according to the GWAS catalog ($p < 5 \times 10^{-8}$), and the bottom panel annotates the positions of the genes.

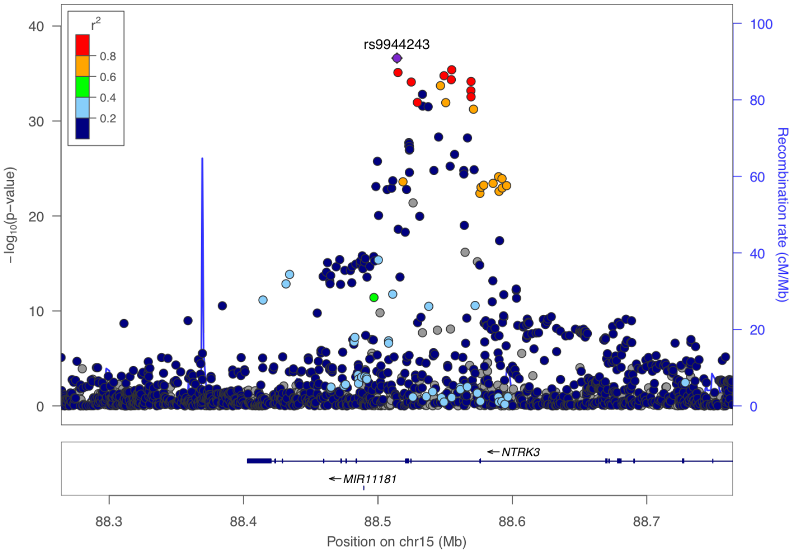


**Supplementary Figure 10-46. Visualization of the cis-pQTL association for protein NTRK3.** The $r^2$ values are the squared linkage disequilibrium correlation coefficients between the plotted variants and the leading variant (purple dot). The middle panel marks the established associations at this locus according to the GWAS catalog ($p < 5 \times 10^{-8}$), and the bottom panel annotates the positions of the genes.

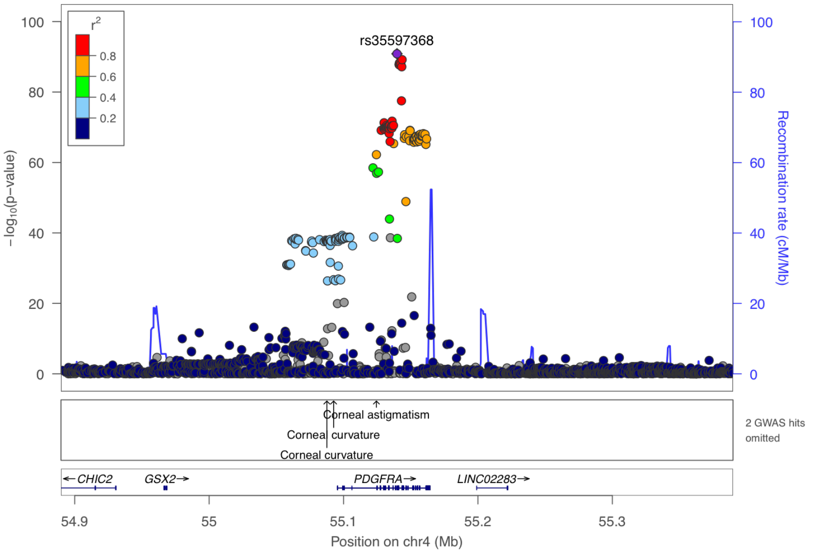


**Supplementary Figure 10-47. Visualization of the cis-pQTL association for protein PDGF R ALPHA.** The $r^2$ values are the squared linkage disequilibrium correlation coefficients between the plotted variants and the leading variant (purple dot). The middle panel marks the established associations at this locus according to the GWAS catalog ($p < 5 \times 10^{-8}$), and the bottom panel annotates the positions of the genes.

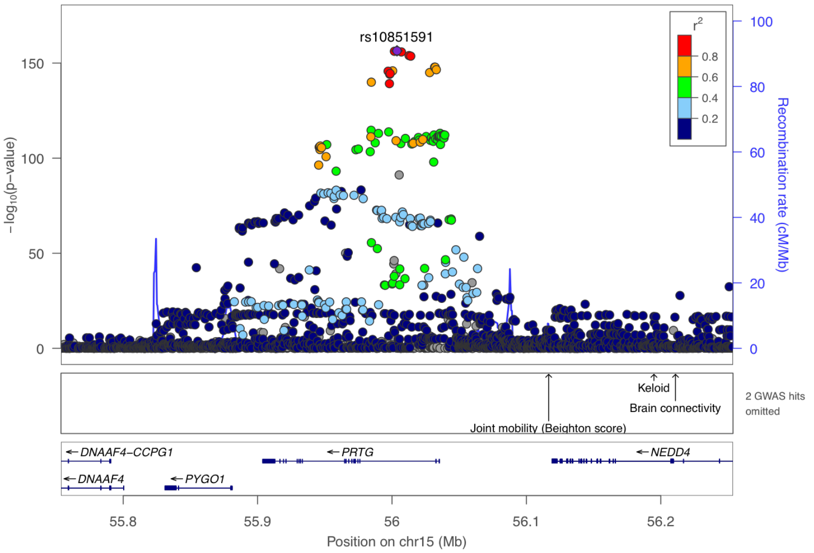


**Supplementary Figure 10-48. Visualization of the cis-pQTL association for protein PRTG.** The $r^2$ values are the squared linkage disequilibrium correlation coefficients between the plotted variants and the leading variant (purple dot). The middle panel marks the established associations at this locus according to the GWAS catalog ($p < 5 \times 10^{-8}$), and the bottom panel annotates the positions of the genes.

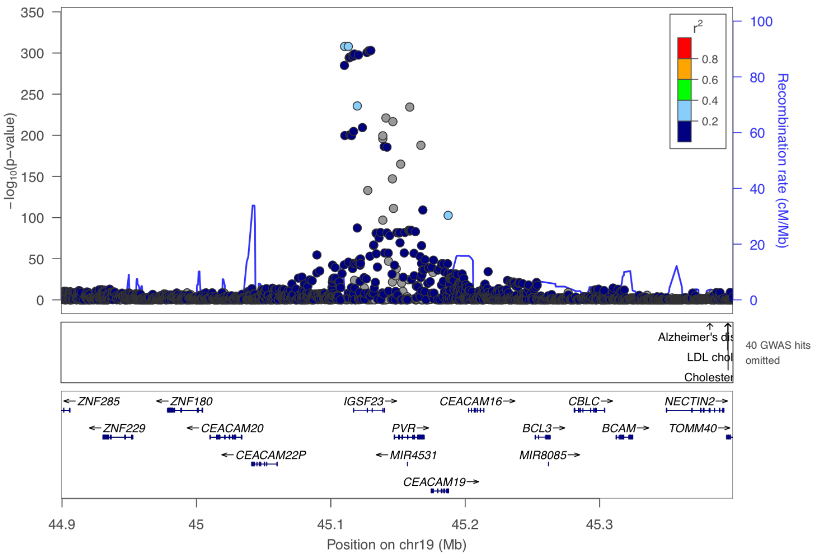


**Supplementary Figure 10-49. Visualization of the cis-pQTL association for protein PVR.** The $r^2$ values are the squared linkage disequilibrium correlation coefficients between the plotted variants and the leading variant (purple dot). The middle panel marks the established associations at this locus according to the GWAS catalog ($p < 5 \times 10^{-8}$), and the bottom panel annotates the positions of the genes.

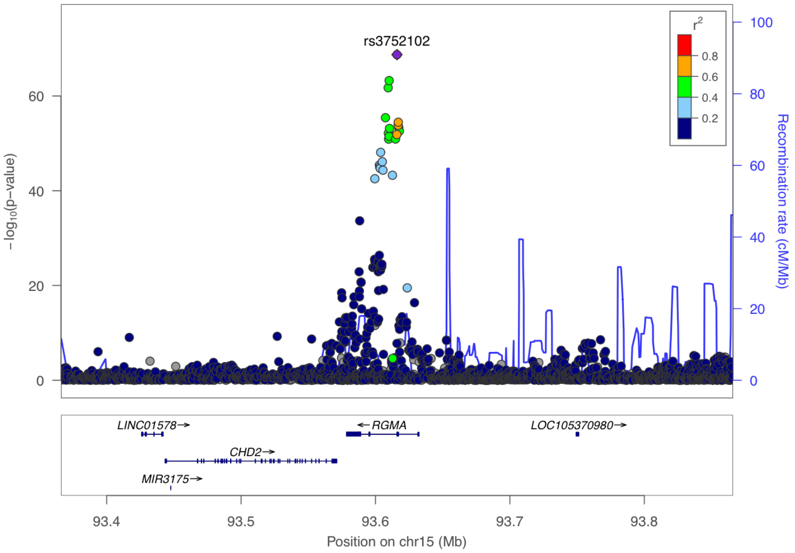


**Supplementary Figure 10-50. Visualization of the cis-pQTL association for protein RGMA.** The $r^2$ values are the squared linkage disequilibrium correlation coefficients between the plotted variants and the leading variant (purple dot). The middle panel marks the established associations at this locus according to the GWAS catalog ($p < 5 \times 10^{-8}$), and the bottom panel annotates the positions of the genes.

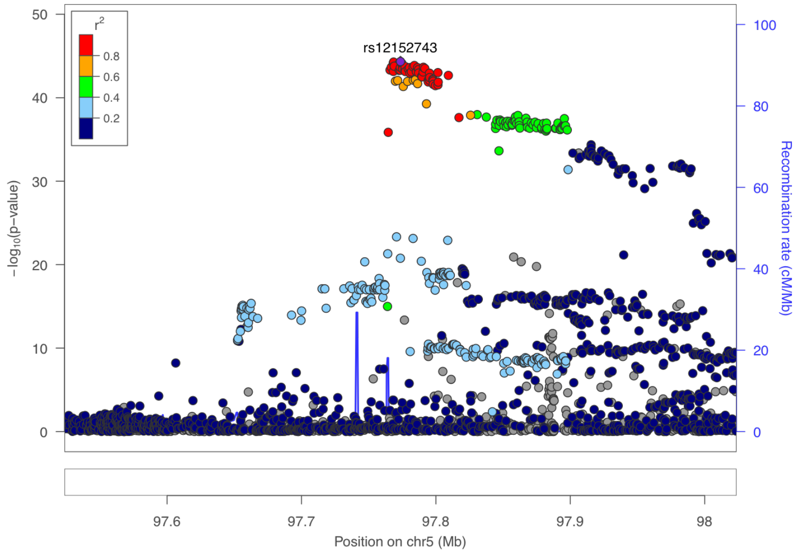


**Supplementary Figure 10-51. Visualization of the cis-pQTL association for protein RGMB.** The $r^2$ values are the squared linkage disequilibrium correlation coefficients between the plotted variants and the leading variant (purple dot). The middle panel marks the established associations at this locus according to the GWAS catalog ($p < 5 \times 10^{-8}$), and the bottom panel annotates the positions of the genes.

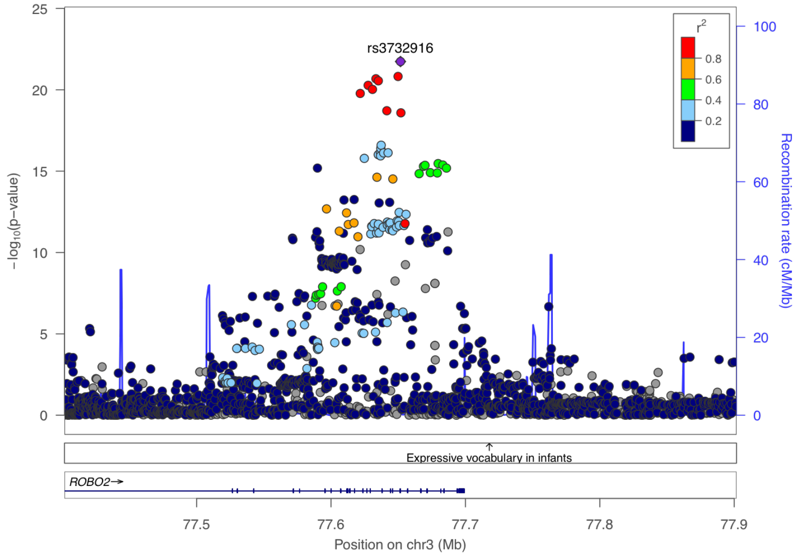


**Supplementary Figure 10-52. Visualization of the cis-pQTL association for protein ROBO2.** The $r^2$ values are the squared linkage disequilibrium correlation coefficients between the plotted variants and the leading variant (purple dot). The middle panel marks the established associations at this locus according to the GWAS catalog ($p < 5 \times 10^{-8}$), and the bottom panel annotates the positions of the genes.

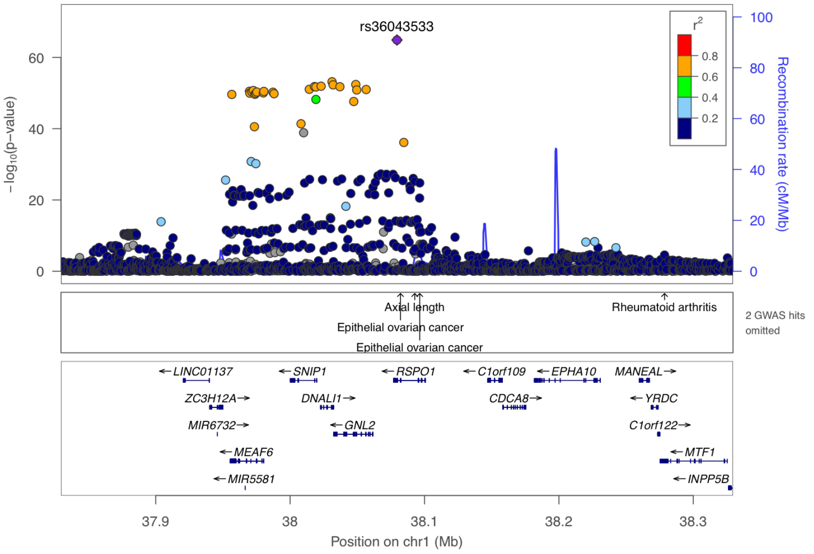


**Supplementary Figure 10-53. Visualization of the cis-pQTL association for protein RSPO1.** The $r^2$ values are the squared linkage disequilibrium correlation coefficients between the plotted variants and the leading variant (purple dot). The middle panel marks the established associations at this locus according to the GWAS catalog ($p < 5 \times 10^{-8}$), and the bottom panel annotates the positions of the genes.

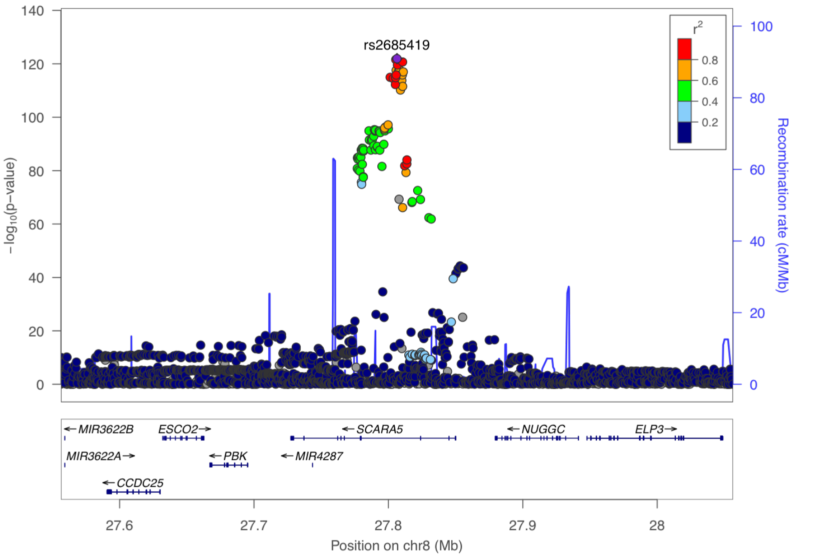


**Supplementary Figure 10-54. Visualization of the cis-pQTL association for protein SCARA5.** The $r^2$ values are the squared linkage disequilibrium correlation coefficients between the plotted variants and the leading variant (purple dot). The middle panel marks the established associations at this locus according to the GWAS catalog ($p < 5 \times 10^{-8}$), and the bottom panel annotates the positions of the genes.

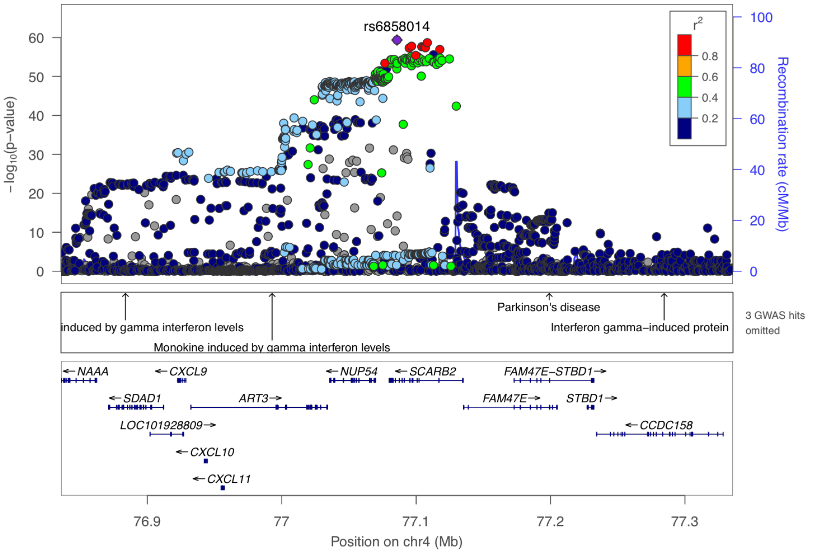


**Supplementary Figure 10-55. Visualization of the cis-pQTL association for protein SCARB2.** The $r^2$ values are the squared linkage disequilibrium correlation coefficients between the plotted variants and the leading variant (purple dot). The middle panel marks the established associations at this locus according to the GWAS catalog ($p < 5 \times 10^{-8}$), and the bottom panel annotates the positions of the genes.

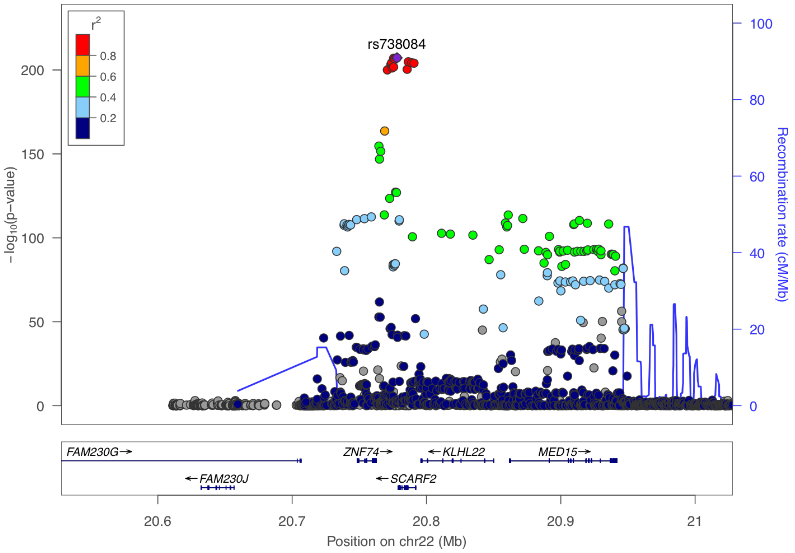


**Supplementary Figure 10-56. Visualization of the cis-pQTL association for protein SCARF2.** The $r^2$ values are the squared linkage disequilibrium correlation coefficients between the plotted variants and the leading variant (purple dot). The middle panel marks the established associations at this locus according to the GWAS catalog ($p < 5 \times 10^{-8}$), and the bottom panel annotates the positions of the genes.

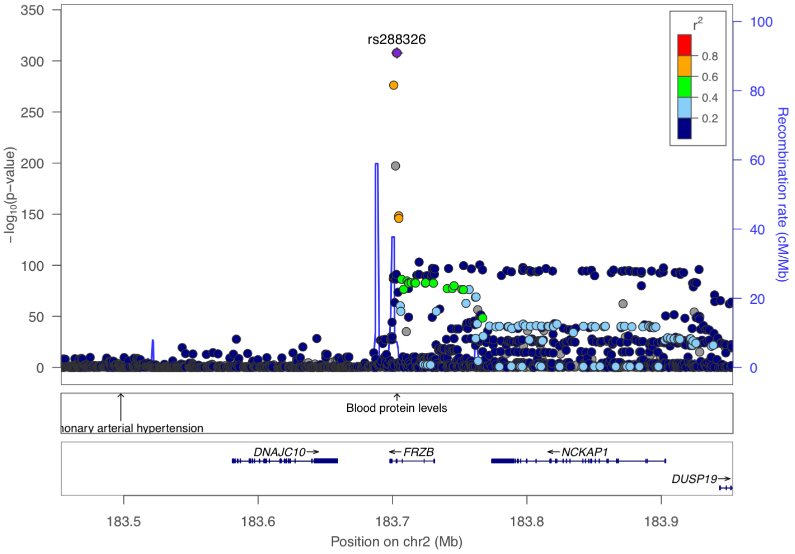


**Supplementary Figure 10-57. Visualization of the cis-pQTL association for protein SFRP 3.** The $r^2$ values are the squared linkage disequilibrium correlation coefficients between the plotted variants and the leading variant (purple dot). The middle panel marks the established associations at this locus according to the GWAS catalog ($p < 5 \times 10^{-8}$), and the bottom panel annotates the positions of the genes.

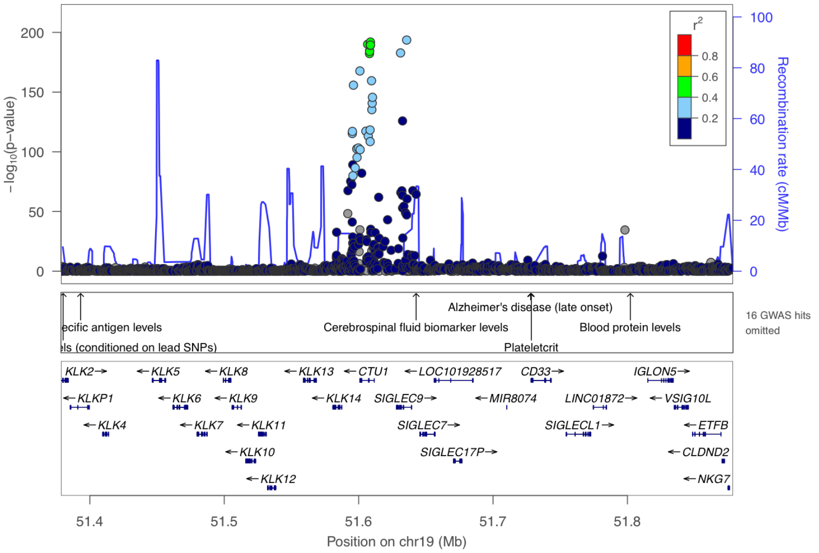


**Supplementary Figure 10-58. Visualization of the cis-pQTL association for protein SIGLEC 9.** The $r^2$ values are the squared linkage disequilibrium correlation coefficients between the plotted variants and the leading variant (purple dot). The middle panel marks the established associations at this locus according to the GWAS catalog ($p < 5 \times 10^{-8}$), and the bottom panel annotates the positions of the genes.

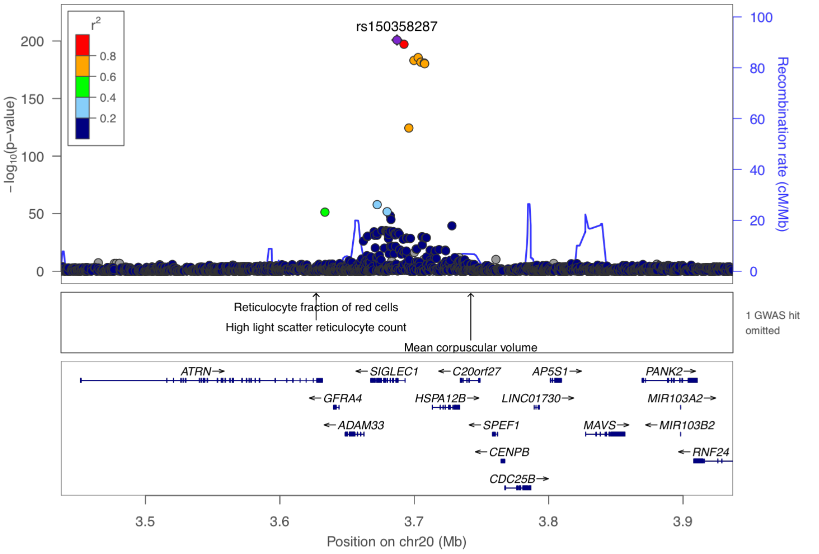


**Supplementary Figure 10-59. Visualization of the cis-pQTL association for protein SIGLEC1.** The $r^2$ values are the squared linkage disequilibrium correlation coefficients between the plotted variants and the leading variant (purple dot). The middle panel marks the established associations at this locus according to the GWAS catalog ($p < 5 \times 10^{-8}$), and the bottom panel annotates the positions of the genes.

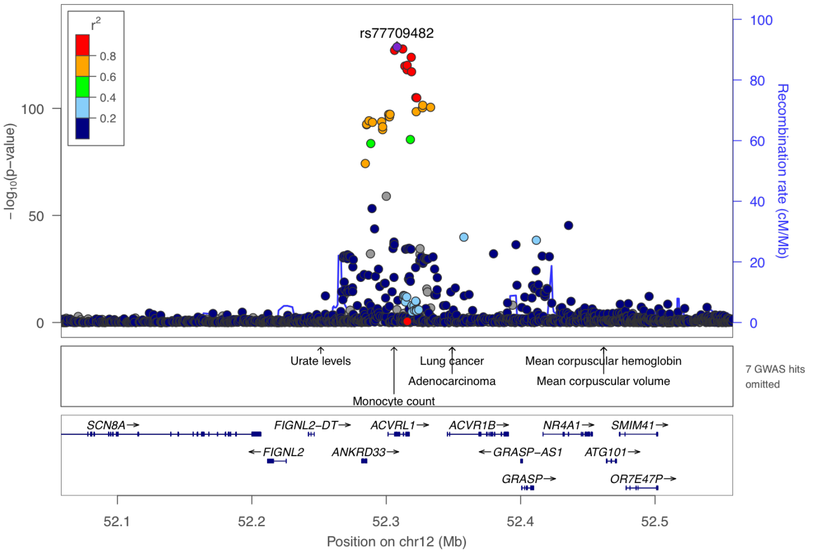


**Supplementary Figure 10-60. Visualization of the cis-pQTL association for protein SKR3.** The $r^2$ values are the squared linkage disequilibrium correlation coefficients between the plotted variants and the leading variant (purple dot). The middle panel marks the established associations at this locus according to the GWAS catalog ($p < 5 \times 10^{-8}$), and the bottom panel annotates the positions of the genes.

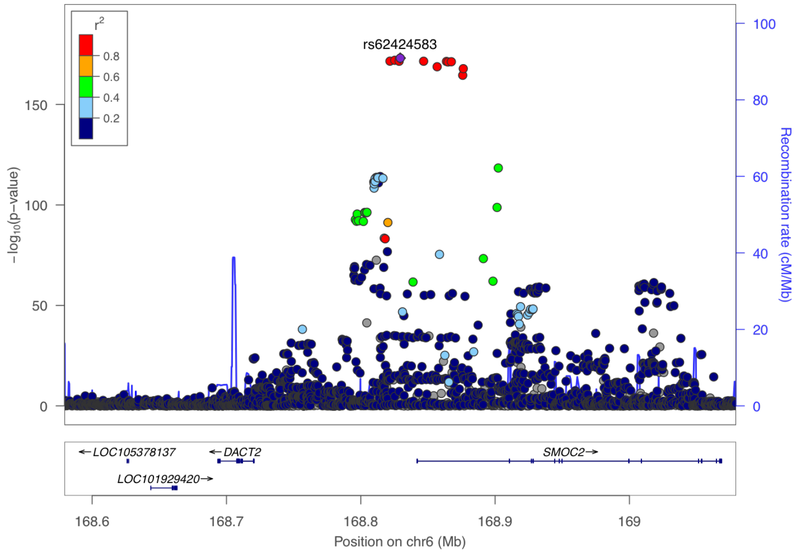


**Supplementary Figure 10-61. Visualization of the cis-pQTL association for protein SMOC2.** The $r^2$ values are the squared linkage disequilibrium correlation coefficients between the plotted variants and the leading variant (purple dot). The middle panel marks the established associations at this locus according to the GWAS catalog ($p < 5 \times 10^{-8}$), and the bottom panel annotates the positions of the genes.

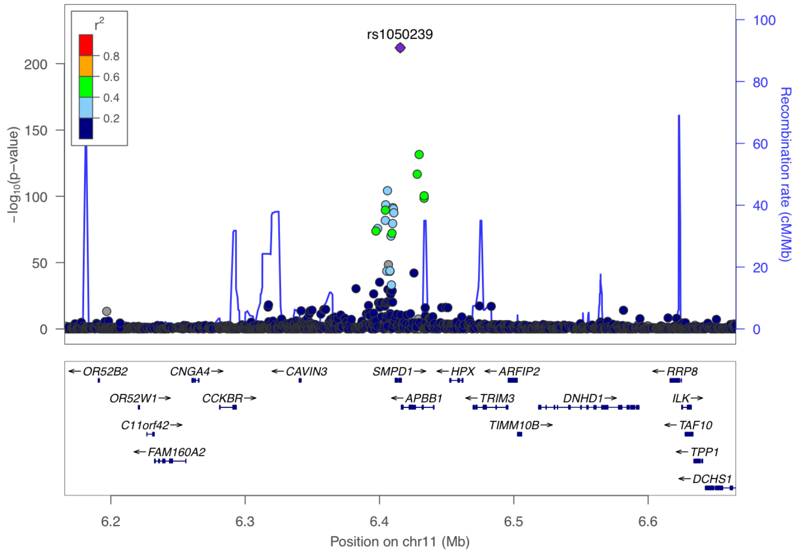


**Supplementary Figure 10-62. Visualization of the cis-pQTL association for protein SMPD1.** The $r^2$ values are the squared linkage disequilibrium correlation coefficients between the plotted variants and the leading variant (purple dot). The middle panel marks the established associations at this locus according to the GWAS catalog ($p < 5 \times 10^{-8}$), and the bottom panel annotates the positions of the genes.

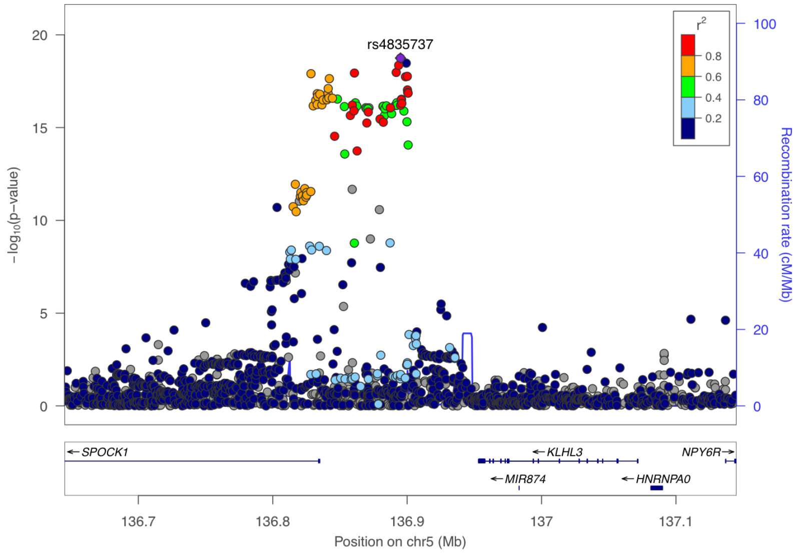


**Supplementary Figure 10-63. Visualization of the cis-pQTL association for protein SPOCK1.** The $r^2$ values are the squared linkage disequilibrium correlation coefficients between the plotted variants and the leading variant (purple dot). The middle panel marks the established associations at this locus according to the GWAS catalog ($p < 5 \times 10^{-8}$), and the bottom panel annotates the positions of the genes.

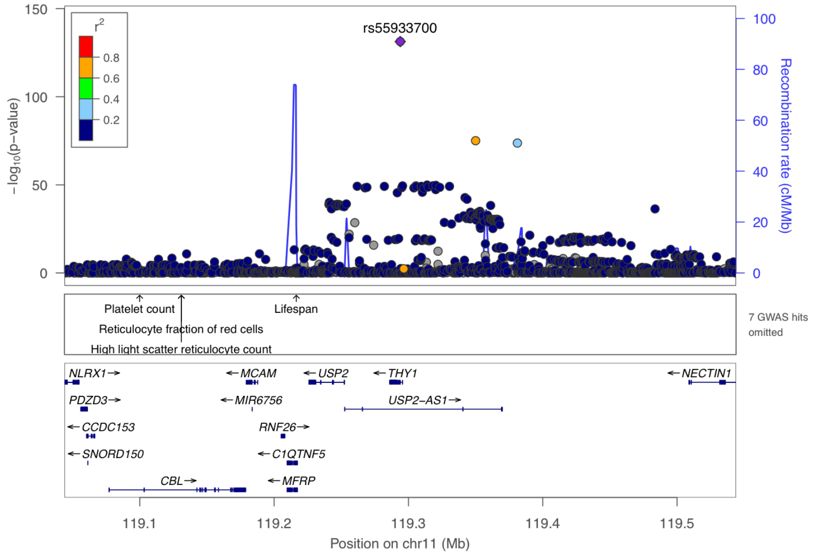


**Supplementary Figure 10-64. Visualization of the cis-pQTL association for protein THY 1.** The $r^2$ values are the squared linkage disequilibrium correlation coefficients between the plotted variants and the leading variant (purple dot). The middle panel marks the established associations at this locus according to the GWAS catalog ($p < 5 \times 10^{-8}$), and the bottom panel annotates the positions of the genes.

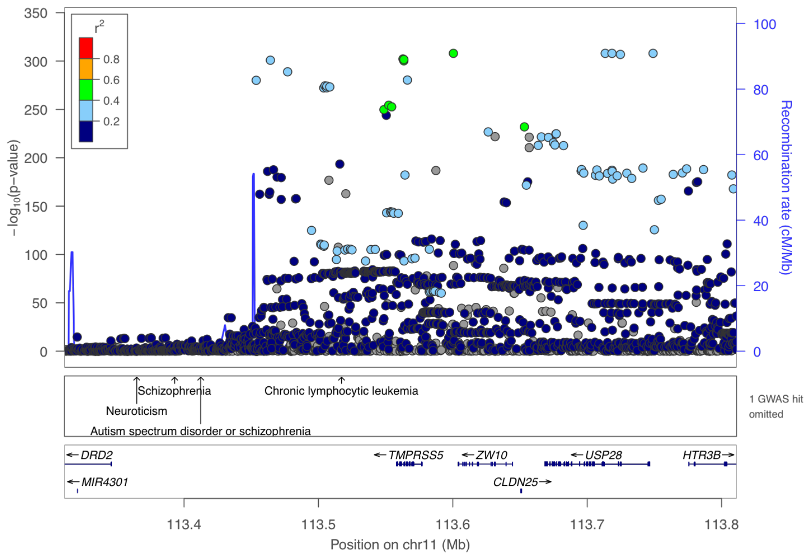


**Supplementary Figure 10-65. Visualization of the cis-pQTL association for protein TMPRSS5.** The $r^2$ values are the squared linkage disequilibrium correlation coefficients between the plotted variants and the leading variant (purple dot). The middle panel marks the established associations at this locus according to the GWAS catalog ($p < 5 \times 10^{-8}$), and the bottom panel annotates the positions of the genes.

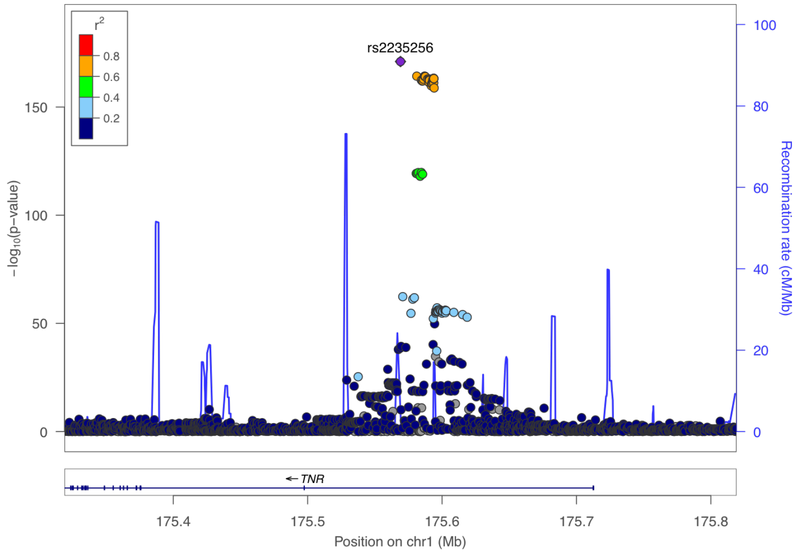


**Supplementary Figure 10-66. Visualization of the cis-pQTL association for protein TN R.** The $r^2$ values are the squared linkage disequilibrium correlation coefficients between the plotted variants and the leading variant (purple dot). The middle panel marks the established associations at this locus according to the GWAS catalog ($p < 5 \times 10^{-8}$), and the bottom panel annotates the positions of the genes.

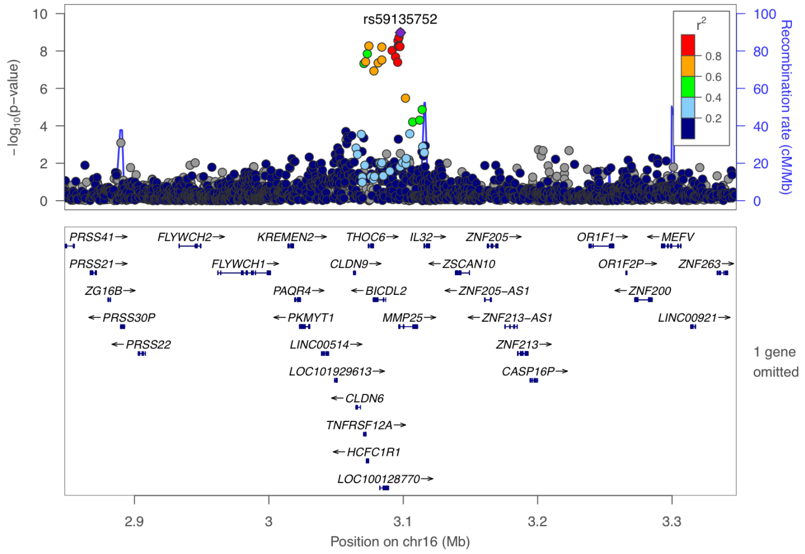


**Supplementary Figure 10-67. Visualization of the cis-pQTL association for protein TNFRSF12A.** The $r^2$ values are the squared linkage disequilibrium correlation coefficients between the plotted variants and the leading variant (purple dot). The middle panel marks the established associations at this locus according to the GWAS catalog ($p < 5 \times 10^{-8}$), and the bottom panel annotates the positions of the genes.

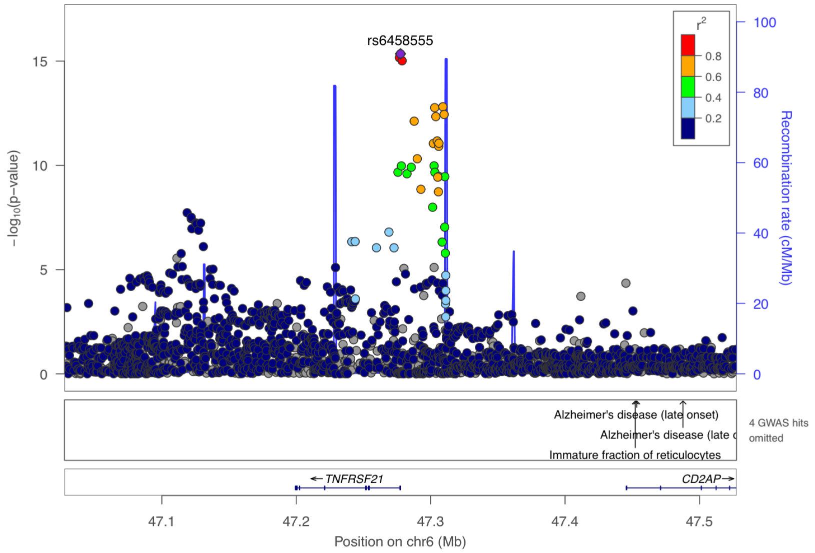


**Supplementary Figure 10-68. Visualization of the cis-pQTL association for protein TNFRSF21.** The $r^2$ values are the squared linkage disequilibrium correlation coefficients between the plotted variants and the leading variant (purple dot). The middle panel marks the established associations at this locus according to the GWAS catalog ($p < 5 \times 10^{-8}$), and the bottom panel annotates the positions of the genes.

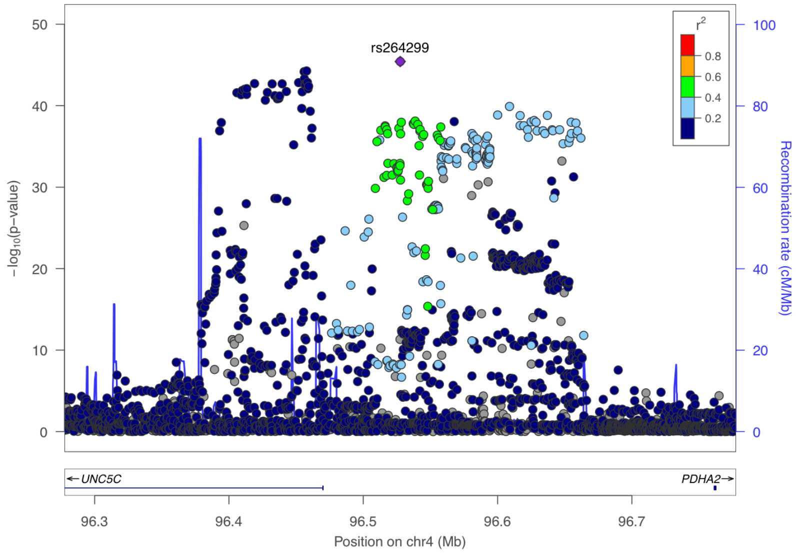


**Supplementary Figure 10-69. Visualization of the cis-pQTL association for protein UNC5C.** The $r^2$ values are the squared linkage disequilibrium correlation coefficients between the plotted variants and the leading variant (purple dot). The middle panel marks the established associations at this locus according to the GWAS catalog ($p < 5 \times 10^{-8}$), and the bottom panel annotates the positions of the genes.

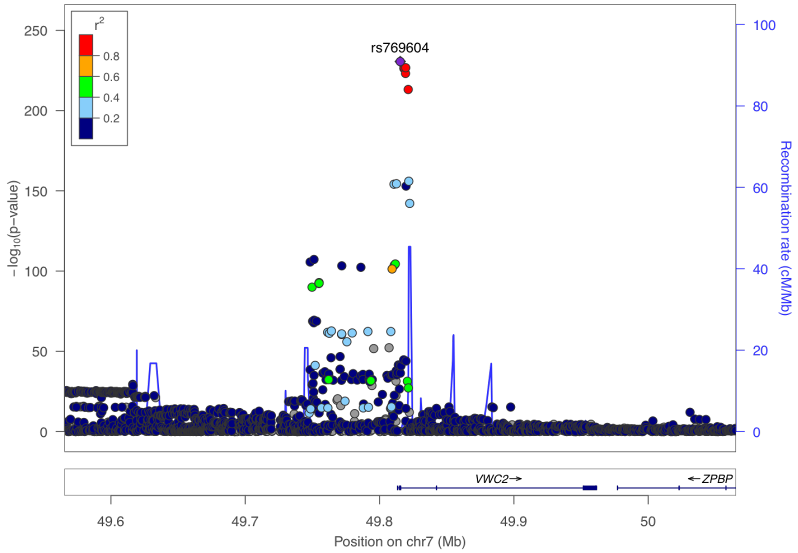


**Supplementary Figure 10-70. Visualization of the cis-pQTL association for protein VWC2.** The $r^2$ values are the squared linkage disequilibrium correlation coefficients between the plotted variants and the leading variant (purple dot). The middle panel marks the established associations at this locus according to the GWAS catalog ($p < 5 \times 10^{-8}$), and the bottom panel annotates the positions of the genes.

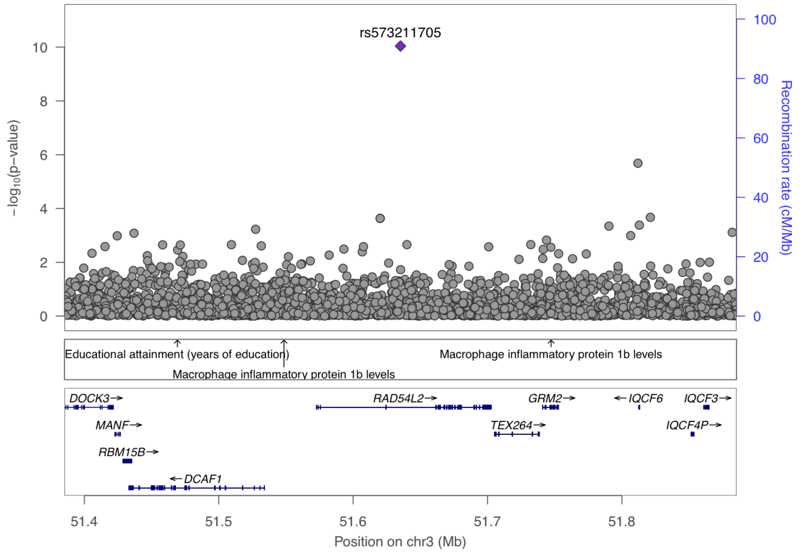


**Supplementary Figure 10-71. Visualization of the cis-pQTL association for protein ABHD14B.** The $r^2$ values are the squared linkage disequilibrium correlation coefficients between the plotted variants and the leading variant (purple dot). The middle panel marks the established associations at this locus according to the GWAS catalog ($p < 5 \times 10^{-8}$), and the bottom panel annotates the positions of the genes.

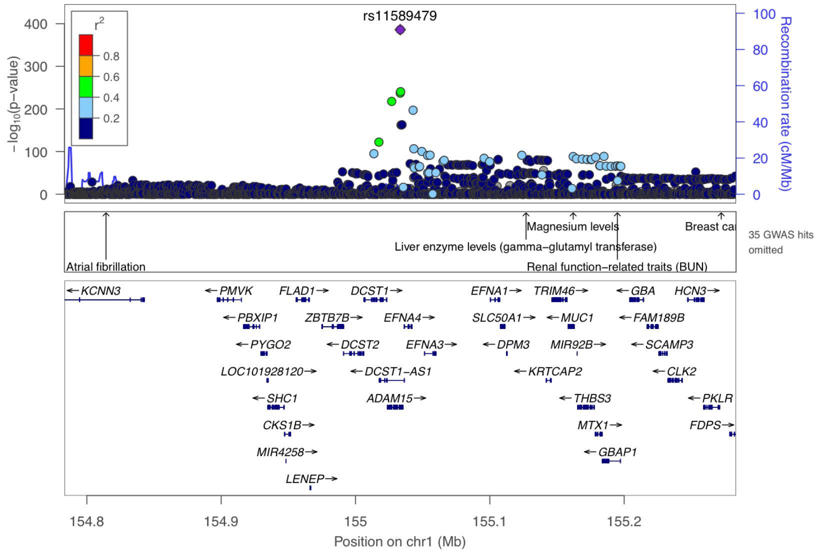


**Supplementary Figure 10-72. Visualization of the cis-pQTL association for protein ADAM15.** The $r^2$ values are the squared linkage disequilibrium correlation coefficients between the plotted variants and the leading variant (purple dot). The middle panel marks the established associations at this locus according to the GWAS catalog ($p < 5 \times 10^{-8}$), and the bottom panel annotates the positions of the genes.

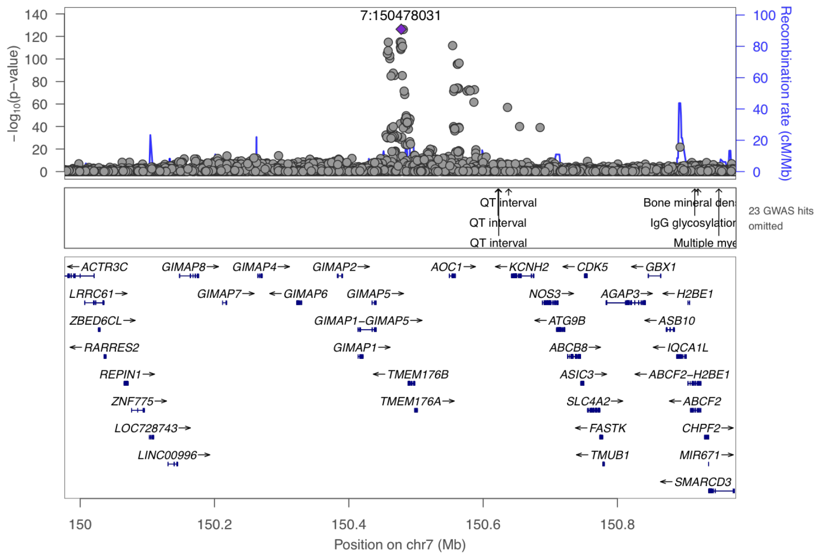


**Supplementary Figure 10-73. Visualization of the cis-pQTL association for protein AOC1.** The $r^2$ values are the squared linkage disequilibrium correlation coefficients between the plotted variants and the leading variant (purple dot). The middle panel marks the established associations at this locus according to the GWAS catalog ($p < 5 \times 10^{-8}$), and the bottom panel annotates the positions of the genes.

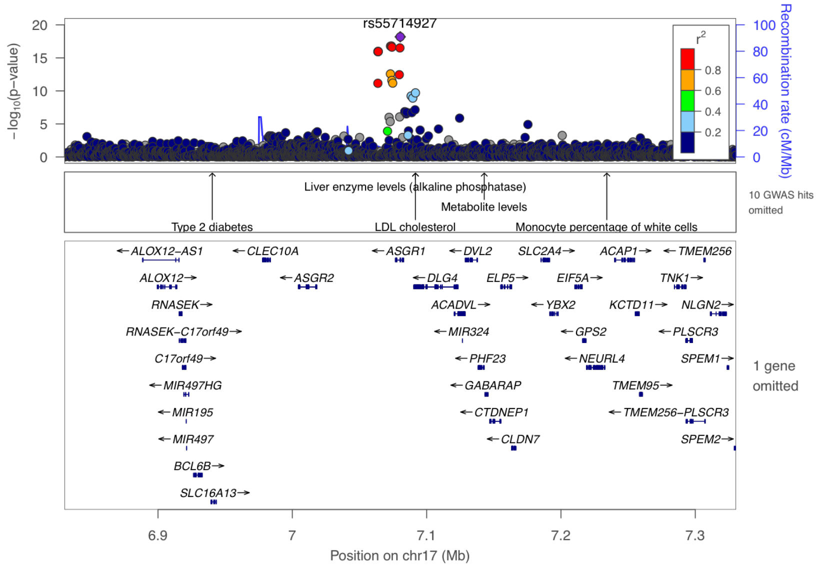


**Supplementary Figure 10-74. Visualization of the cis-pQTL association for protein ASGR1.** The $r^2$ values are the squared linkage disequilibrium correlation coefficients between the plotted variants and the leading variant (purple dot). The middle panel marks the established associations at this locus according to the GWAS catalog ($p < 5 \times 10^{-8}$), and the bottom panel annotates the positions of the genes.

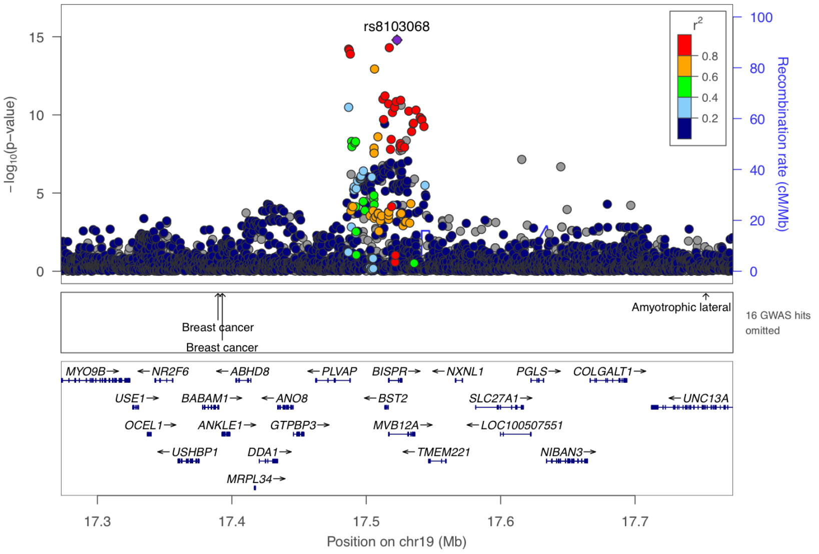


**Supplementary Figure 10-75. Visualization of the cis-pQTL association for protein BST2.** The $r^2$ values are the squared linkage disequilibrium correlation coefficients between the plotted variants and the leading variant (purple dot). The middle panel marks the established associations at this locus according to the GWAS catalog ($p < 5 \times 10^{-8}$), and the bottom panel annotates the positions of the genes.

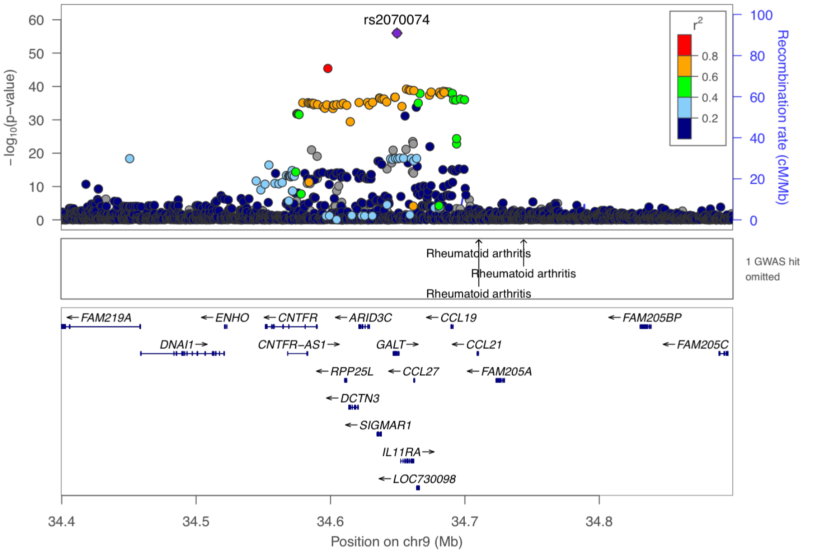


**Supplementary Figure 10-76. Visualization of the cis-pQTL association for protein CCL27.** The $r^2$ values are the squared linkage disequilibrium correlation coefficients between the plotted variants and the leading variant (purple dot). The middle panel marks the established associations at this locus according to the GWAS catalog ($p < 5 \times 10^{-8}$), and the bottom panel annotates the positions of the genes.

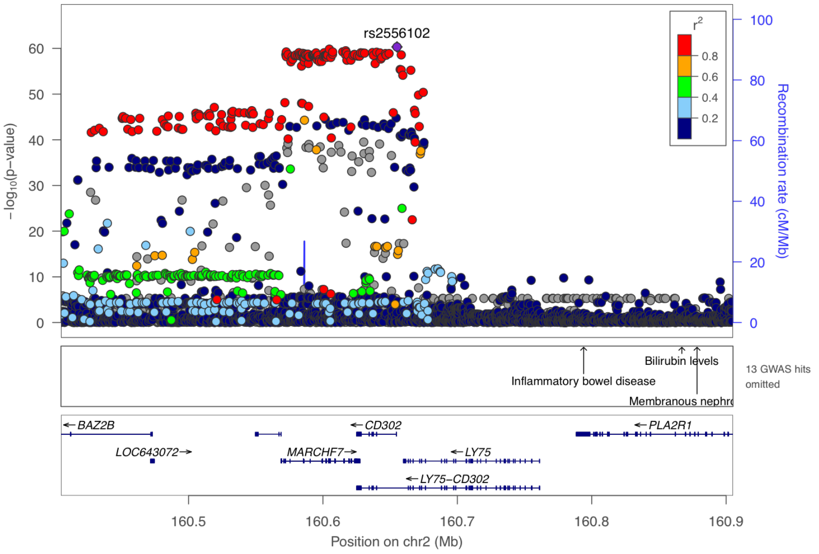


**Supplementary Figure 10-77. Visualization of the cis-pQTL association for protein CD302.** The $r^2$ values are the squared linkage disequilibrium correlation coefficients between the plotted variants and the leading variant (purple dot). The middle panel marks the established associations at this locus according to the GWAS catalog ($p < 5 \times 10^{-8}$), and the bottom panel annotates the positions of the genes.

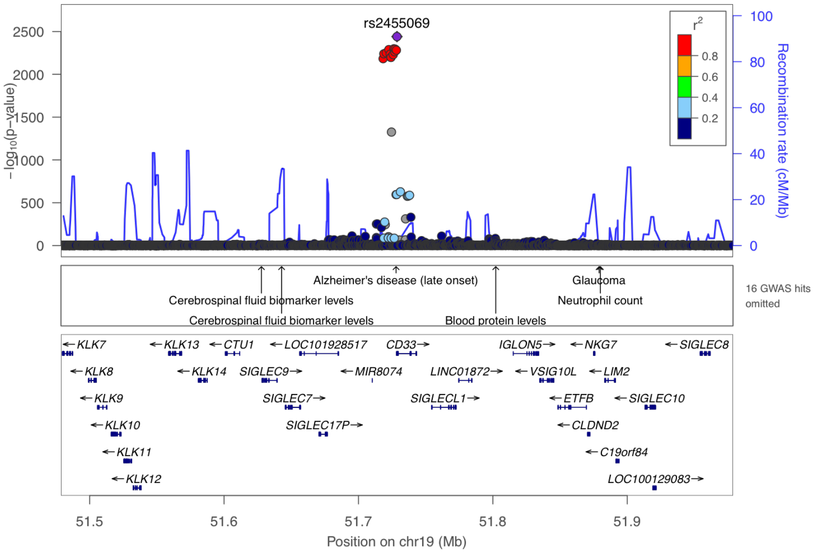


**Supplementary Figure 10-78. Visualization of the cis-pQTL association for protein CD33.** The $r^2$ values are the squared linkage disequilibrium correlation coefficients between the plotted variants and the leading variant (purple dot). The middle panel marks the established associations at this locus according to the GWAS catalog ($p < 5 \times 10^{-8}$), and the bottom panel annotates the positions of the genes.

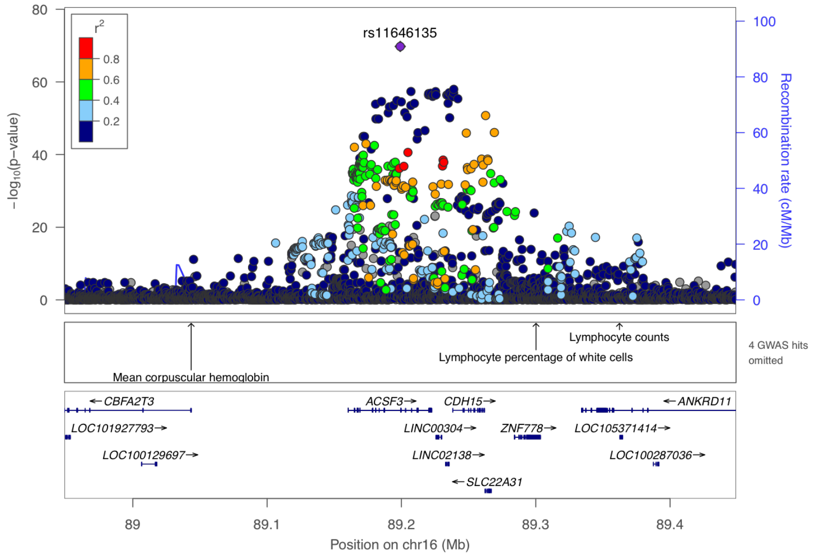


**Supplementary Figure 10-79. Visualization of the cis-pQTL association for protein CDH15.** The $r^2$ values are the squared linkage disequilibrium correlation coefficients between the plotted variants and the leading variant (purple dot). The middle panel marks the established associations at this locus according to the GWAS catalog ($p < 5 \times 10^{-8}$), and the bottom panel annotates the positions of the genes.

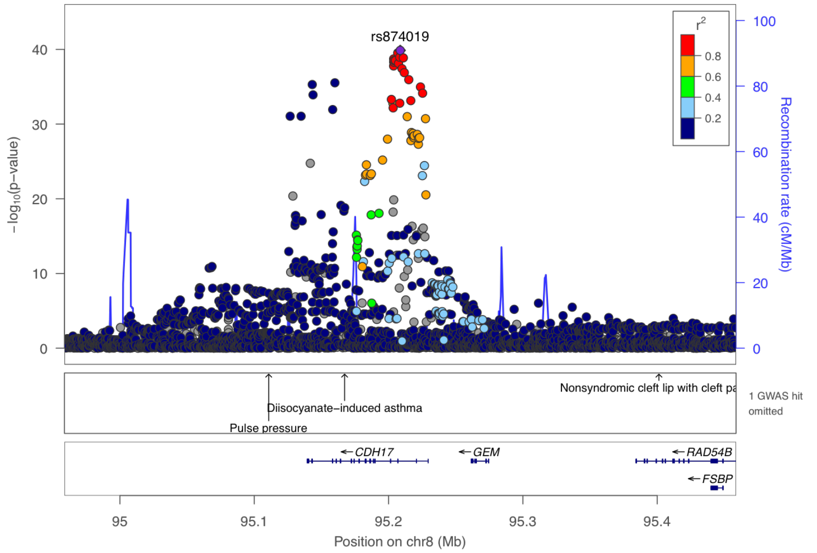


**Supplementary Figure 10-80. Visualization of the cis-pQTL association for protein CDH17.** The $r^2$ values are the squared linkage disequilibrium correlation coefficients between the plotted variants and the leading variant (purple dot). The middle panel marks the established associations at this locus according to the GWAS catalog ($p < 5 \times 10^{-8}$), and the bottom panel annotates the positions of the genes.

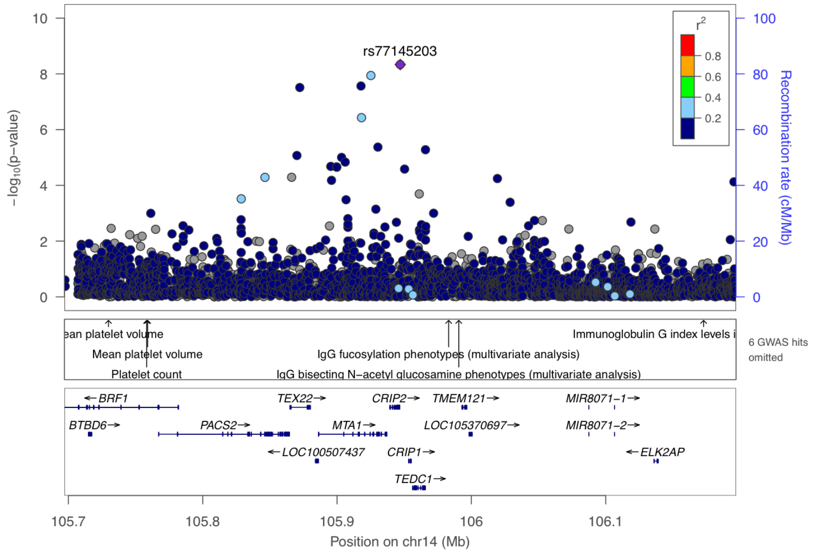


**Supplementary Figure 10-81. Visualization of the cis-pQTL association for protein CRIP2.** The $r^2$ values are the squared linkage disequilibrium correlation coefficients between the plotted variants and the leading variant (purple dot). The middle panel marks the established associations at this locus according to the GWAS catalog ($p < 5 \times 10^{-8}$), and the bottom panel annotates the positions of the genes.

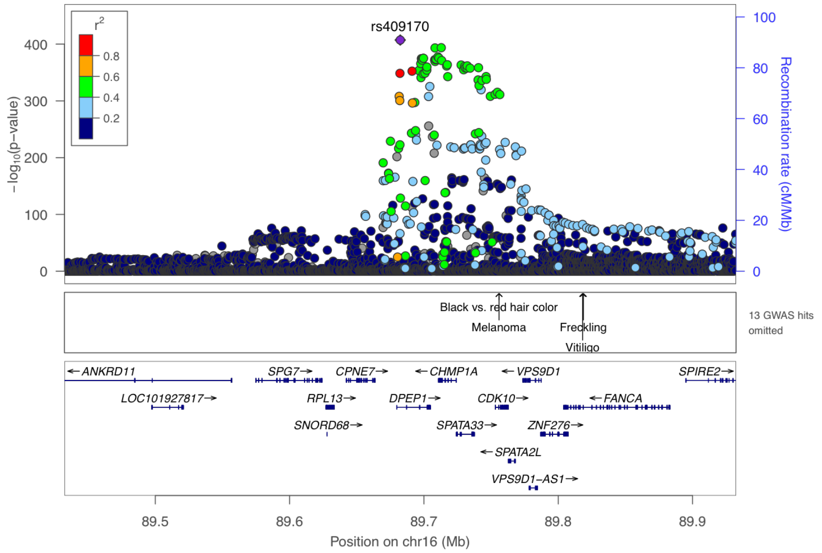


**Supplementary Figure 10-82. Visualization of the cis-pQTL association for protein DPEP1.** The $r^2$ values are the squared linkage disequilibrium correlation coefficients between the plotted variants and the leading variant (purple dot). The middle panel marks the established associations at this locus according to the GWAS catalog ($p < 5 \times 10^{-8}$), and the bottom panel annotates the positions of the genes.

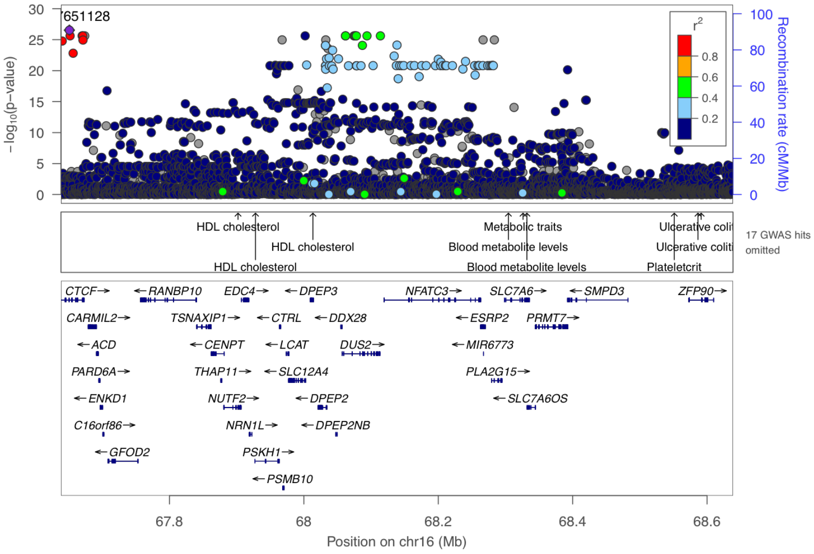


**Supplementary Figure 10-83. Visualization of the cis-pQTL association for protein DPEP2.** The $r^2$ values are the squared linkage disequilibrium correlation coefficients between the plotted variants and the leading variant (purple dot). The middle panel marks the established associations at this locus according to the GWAS catalog ($p < 5 \times 10^{-8}$), and the bottom panel annotates the positions of the genes.

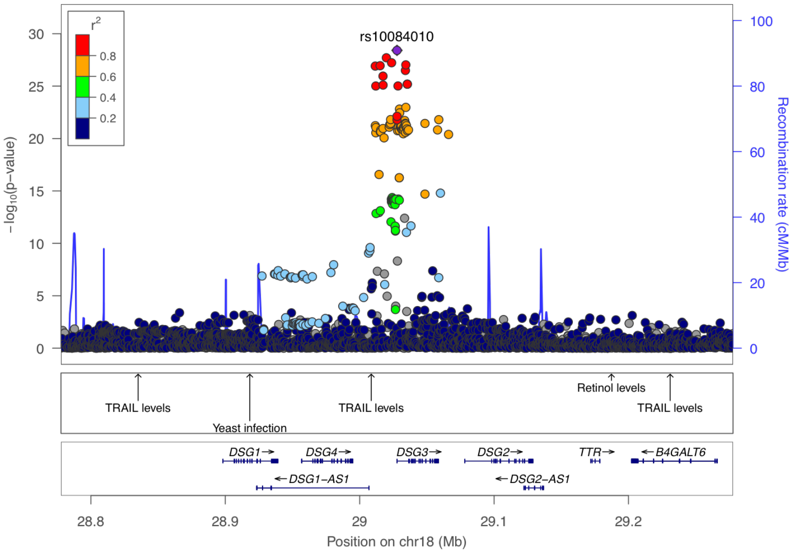


**Supplementary Figure 10-84. Visualization of the cis-pQTL association for protein DSG3.** The $r^2$ values are the squared linkage disequilibrium correlation coefficients between the plotted variants and the leading variant (purple dot). The middle panel marks the established associations at this locus according to the GWAS catalog ($p < 5 \times 10^{-8}$), and the bottom panel annotates the positions of the genes.

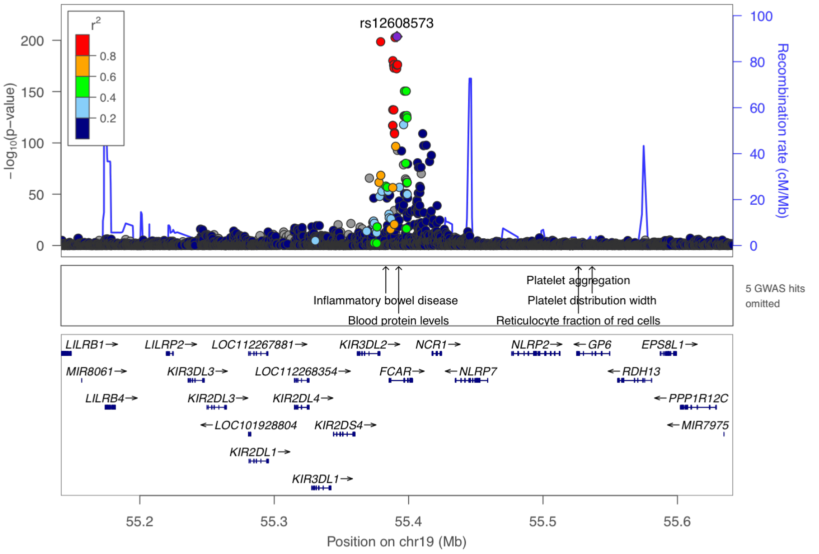


**Supplementary Figure 10-85. Visualization of the cis-pQTL association for protein FCAR.** The $r^2$ values are the squared linkage disequilibrium correlation coefficients between the plotted variants and the leading variant (purple dot). The middle panel marks the established associations at this locus according to the GWAS catalog ($p < 5 \times 10^{-8}$), and the bottom panel annotates the positions of the genes.

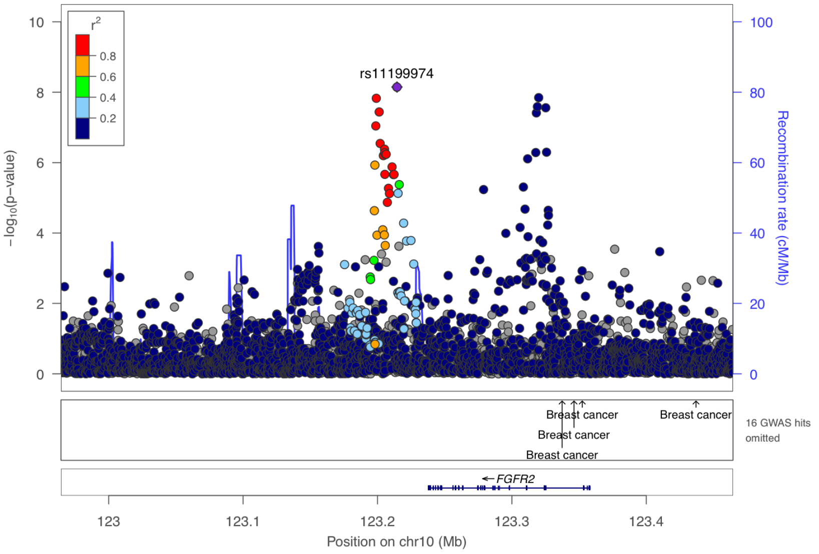


**Supplementary Figure 10-86. Visualization of the cis-pQTL association for protein FGFR2.** The $r^2$ values are the squared linkage disequilibrium correlation coefficients between the plotted variants and the leading variant (purple dot). The middle panel marks the established associations at this locus according to the GWAS catalog ($p < 5 \times 10^{-8}$), and the bottom panel annotates the positions of the genes.

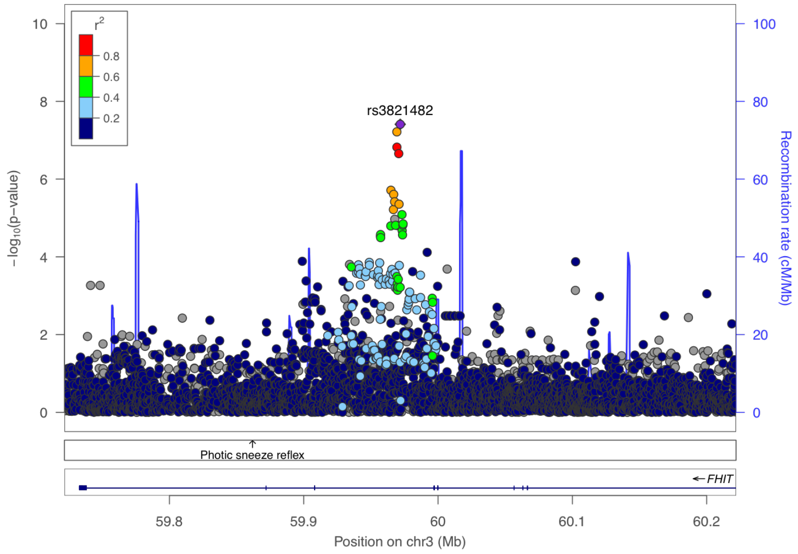


**Supplementary Figure 10-87. Visualization of the cis-pQTL association for protein FHIT.** The $r^2$ values are the squared linkage disequilibrium correlation coefficients between the plotted variants and the leading variant (purple dot). The middle panel marks the established associations at this locus according to the GWAS catalog ($p < 5 \times 10^{-8}$), and the bottom panel annotates the positions of the genes.

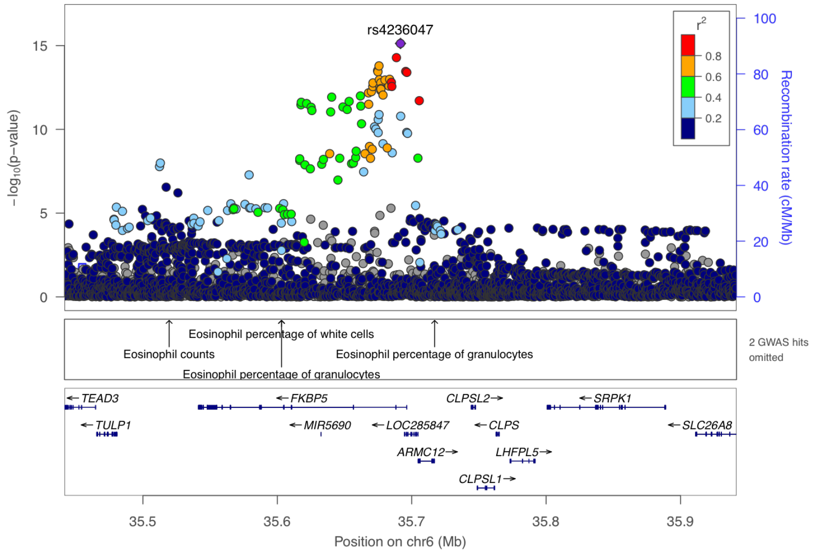


**Supplementary Figure 10-88. Visualization of the cis-pQTL association for protein FKBP5.** The $r^2$ values are the squared linkage disequilibrium correlation coefficients between the plotted variants and the leading variant (purple dot). The middle panel marks the established associations at this locus according to the GWAS catalog ($p < 5 \times 10^{-8}$), and the bottom panel annotates the positions of the genes.

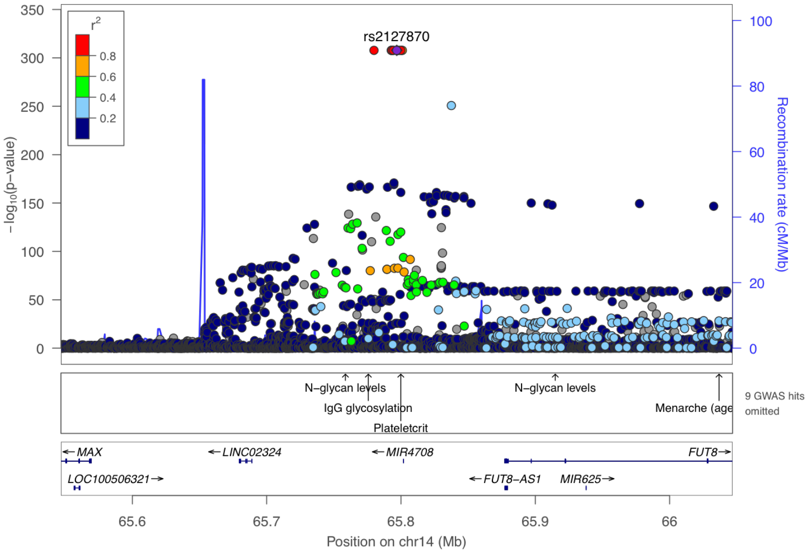


**Supplementary Figure 10-89. Visualization of the cis-pQTL association for protein FUT8.** The $r^2$ values are the squared linkage disequilibrium correlation coefficients between the plotted variants and the leading variant (purple dot). The middle panel marks the established associations at this locus according to the GWAS catalog ($p < 5 \times 10^{-8}$), and the bottom panel annotates the positions of the genes.

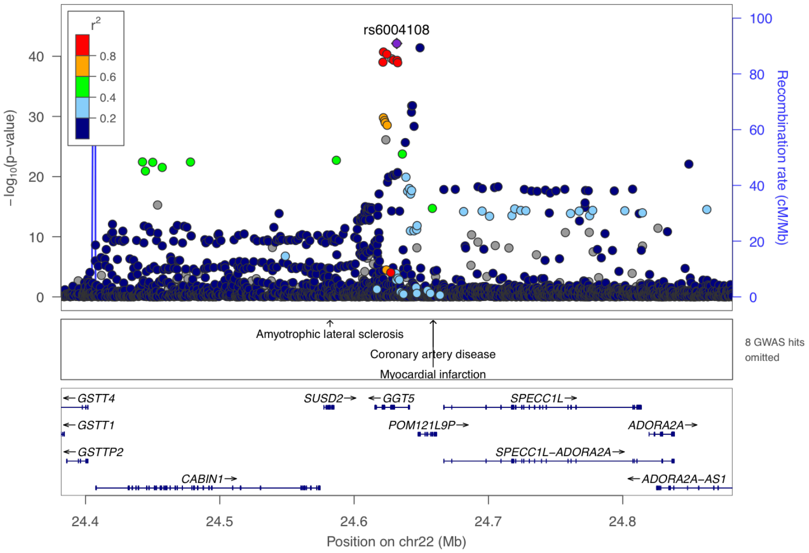


**Supplementary Figure 10-90. Visualization of the cis-pQTL association for protein GGT5.** The $r^2$ values are the squared linkage disequilibrium correlation coefficients between the plotted variants and the leading variant (purple dot). The middle panel marks the established associations at this locus according to the GWAS catalog ($p < 5 \times 10^{-8}$), and the bottom panel annotates the positions of the genes.

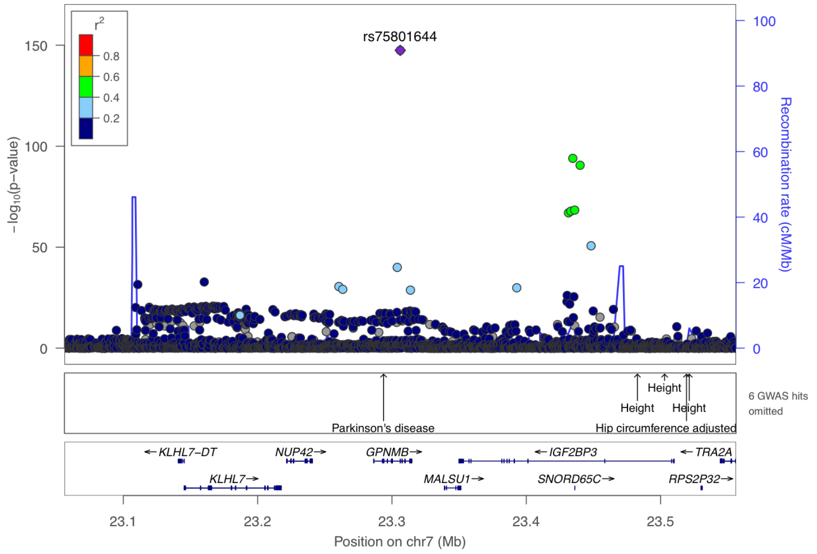


**Supplementary Figure 10-91. Visualization of the cis-pQTL association for protein GPNMB.** The $r^2$ values are the squared linkage disequilibrium correlation coefficients between the plotted variants and the leading variant (purple dot). The middle panel marks the established associations at this locus according to the GWAS catalog ($p < 5 \times 10^{-8}$), and the bottom panel annotates the positions of the genes.

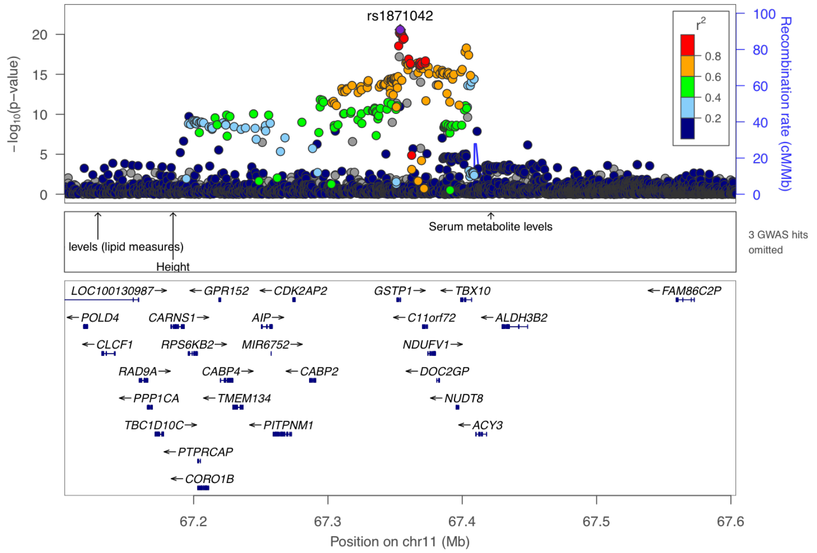


**Supplementary Figure 10-92. Visualization of the cis-pQTL association for protein GSTP1.** The $r^2$ values are the squared linkage disequilibrium correlation coefficients between the plotted variants and the leading variant (purple dot). The middle panel marks the established associations at this locus according to the GWAS catalog ($p < 5 \times 10^{-8}$), and the bottom panel annotates the positions of the genes.

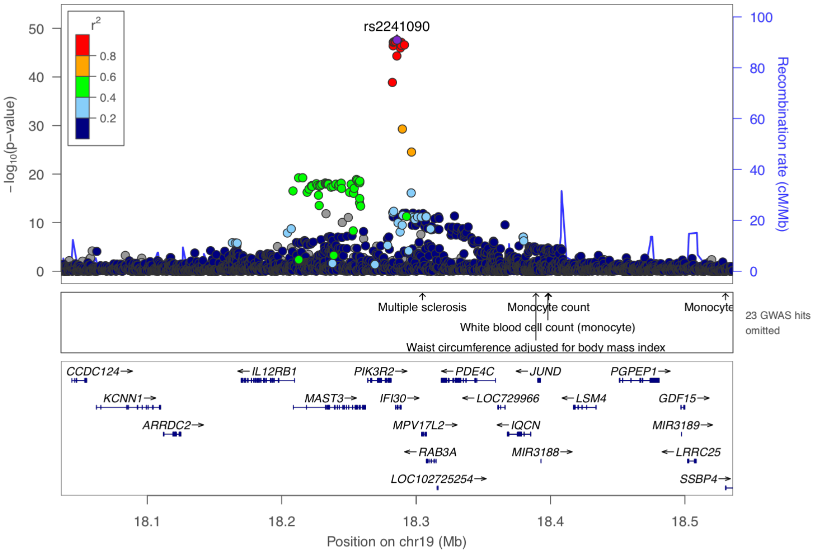


**Supplementary Figure 10-93. Visualization of the cis-pQTL association for protein IFI30.** The $r^2$ values are the squared linkage disequilibrium correlation coefficients between the plotted variants and the leading variant (purple dot). The middle panel marks the established associations at this locus according to the GWAS catalog ($p < 5 \times 10^{-8}$), and the bottom panel annotates the positions of the genes.

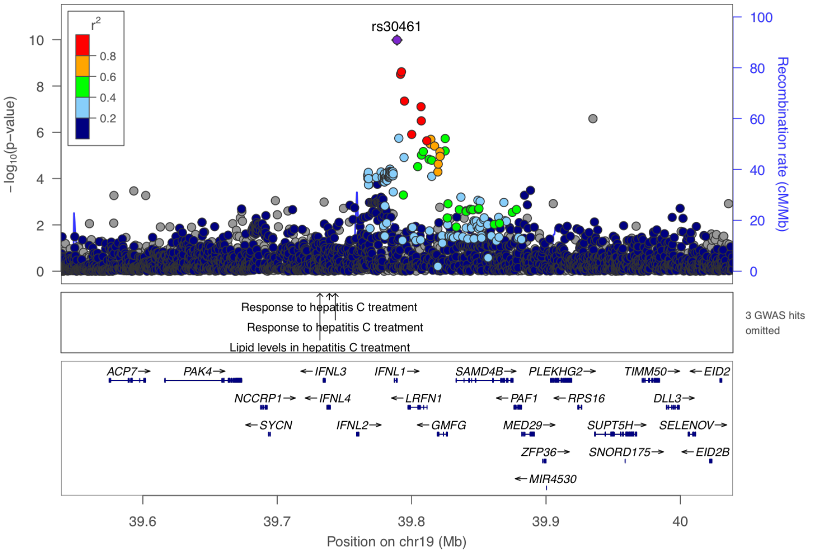


**Supplementary Figure 10-94. Visualization of the cis-pQTL association for protein IFNL1.** The $r^2$ values are the squared linkage disequilibrium correlation coefficients between the plotted variants and the leading variant (purple dot). The middle panel marks the established associations at this locus according to the GWAS catalog ($p < 5 \times 10^{-8}$), and the bottom panel annotates the positions of the genes.

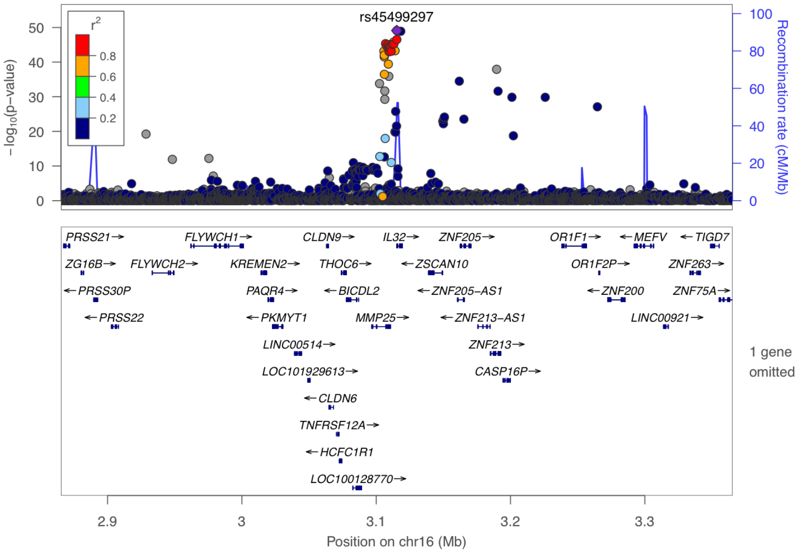


**Supplementary Figure 10-95. Visualization of the cis-pQTL association for protein IL32.** The $r^2$ values are the squared linkage disequilibrium correlation coefficients between the plotted variants and the leading variant (purple dot). The middle panel marks the established associations at this locus according to the GWAS catalog ($p < 5 \times 10^{-8}$), and the bottom panel annotates the positions of the genes.

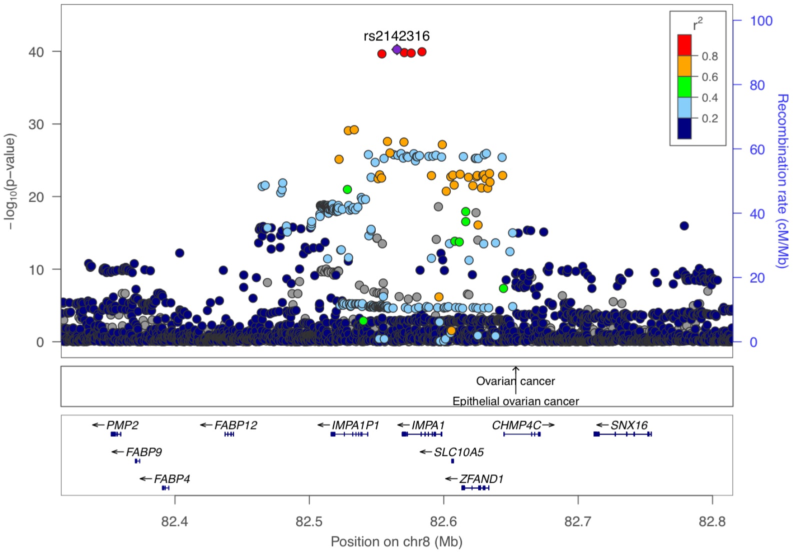


**Supplementary Figure 10-96. Visualization of the cis-pQTL association for protein IMPA1.** The $r^2$ values are the squared linkage disequilibrium correlation coefficients between the plotted variants and the leading variant (purple dot). The middle panel marks the established associations at this locus according to the GWAS catalog ($p < 5 \times 10^{-8}$), and the bottom panel annotates the positions of the genes.

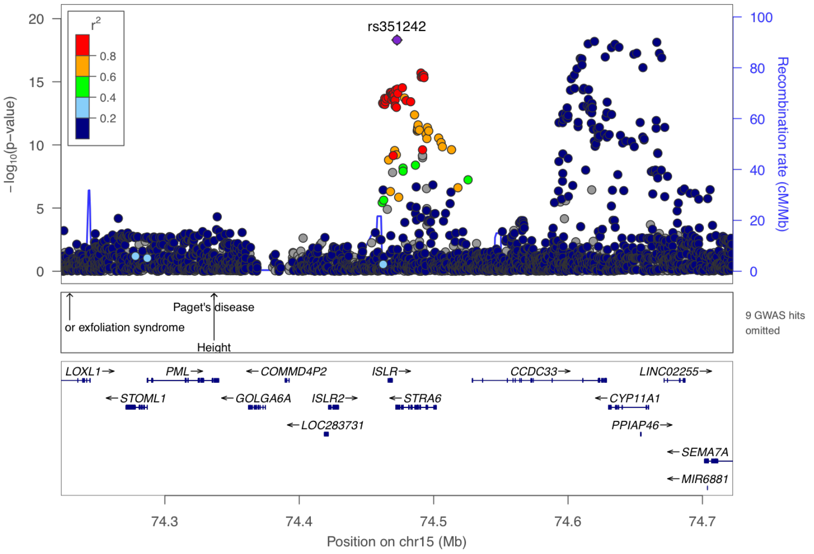


**Supplementary Figure 10-97. Visualization of the cis-pQTL association for protein ISLR2.** The $r^2$ values are the squared linkage disequilibrium correlation coefficients between the plotted variants and the leading variant (purple dot). The middle panel marks the established associations at this locus according to the GWAS catalog ($p < 5 \times 10^{-8}$), and the bottom panel annotates the positions of the genes.

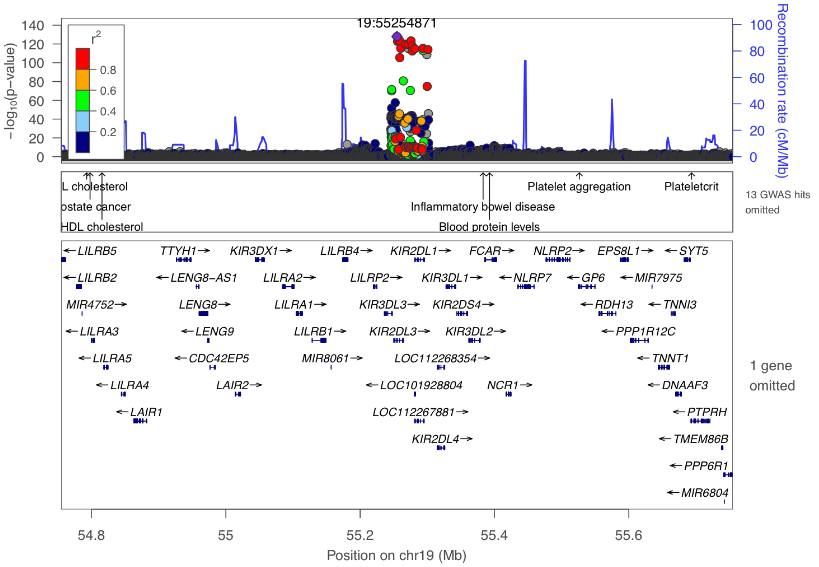


**Supplementary Figure 10-98. Visualization of the cis-pQTL association for protein KIR2DL3.** The $r^2$ values are the squared linkage disequilibrium correlation coefficients between the plotted variants and the leading variant (purple dot). The middle panel marks the established associations at this locus according to the GWAS catalog ($p < 5 \times 10^{-8}$), and the bottom panel annotates the positions of the genes.

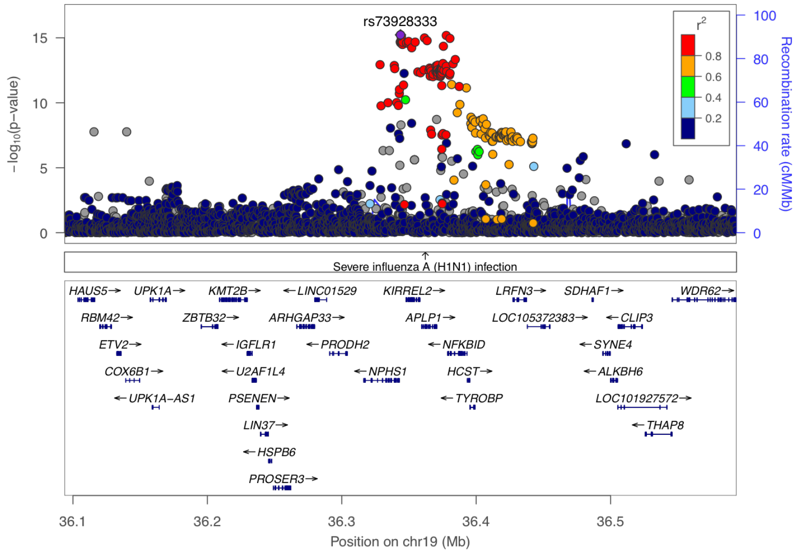


**Supplementary Figure 10-99. Visualization of the cis-pQTL association for protein KIRREL2.** The $r^2$ values are the squared linkage disequilibrium correlation coefficients between the plotted variants and the leading variant (purple dot). The middle panel marks the established associations at this locus according to the GWAS catalog ($p < 5 \times 10^{-8}$), and the bottom panel annotates the positions of the genes.

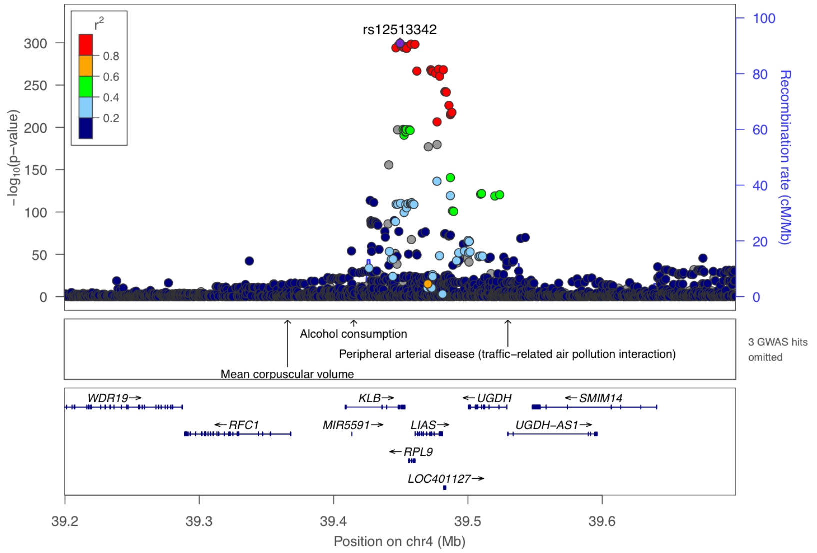


**Supplementary Figure 10-100. Visualization of the cis-pQTL association for protein KLB.** The $r^2$ values are the squared linkage disequilibrium correlation coefficients between the plotted variants and the leading variant (purple dot). The middle panel marks the established associations at this locus according to the GWAS catalog ($p < 5 \times 10^{-8}$), and the bottom panel annotates the positions of the genes.

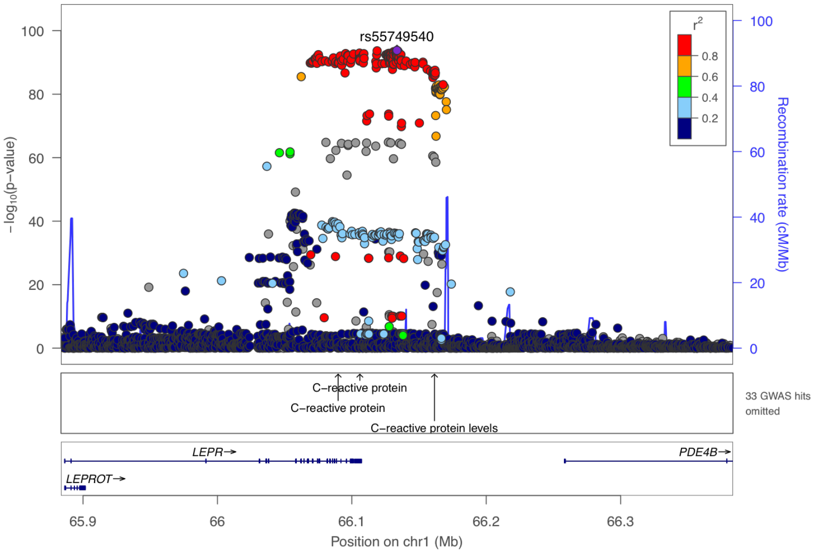


**Supplementary Figure 10-101. Visualization of the cis-pQTL association for protein LEPR.** The $r^2$ values are the squared linkage disequilibrium correlation coefficients between the plotted variants and the leading variant (purple dot). The middle panel marks the established associations at this locus according to the GWAS catalog ($p < 5 \times 10^{-8}$), and the bottom panel annotates the positions of the genes.

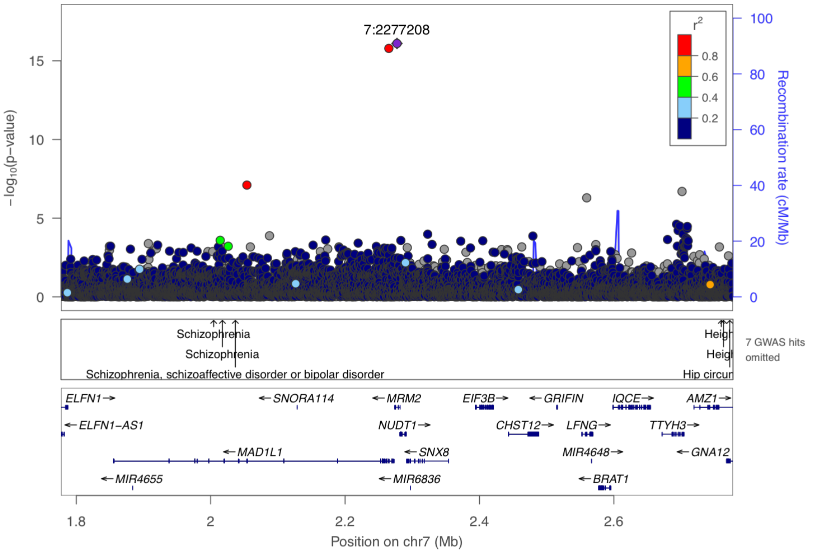


**Supplementary Figure 10-102. Visualization of the cis-pQTL association for protein MAD1L1.** The $r^2$ values are the squared linkage disequilibrium correlation coefficients between the plotted variants and the leading variant (purple dot). The middle panel marks the established associations at this locus according to the GWAS catalog ($p < 5 \times 10^{-8}$), and the bottom panel annotates the positions of the genes.

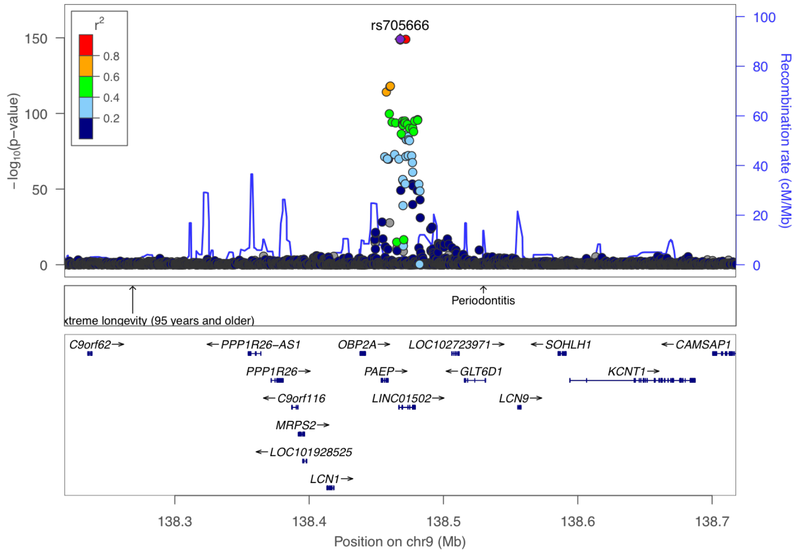


**Supplementary Figure 10-103. Visualization of the cis-pQTL association for protein PAEP.** The $r^2$ values are the squared linkage disequilibrium correlation coefficients between the plotted variants and the leading variant (purple dot). The middle panel marks the established associations at this locus according to the GWAS catalog ($p < 5 \times 10^{-8}$), and the bottom panel annotates the positions of the genes.

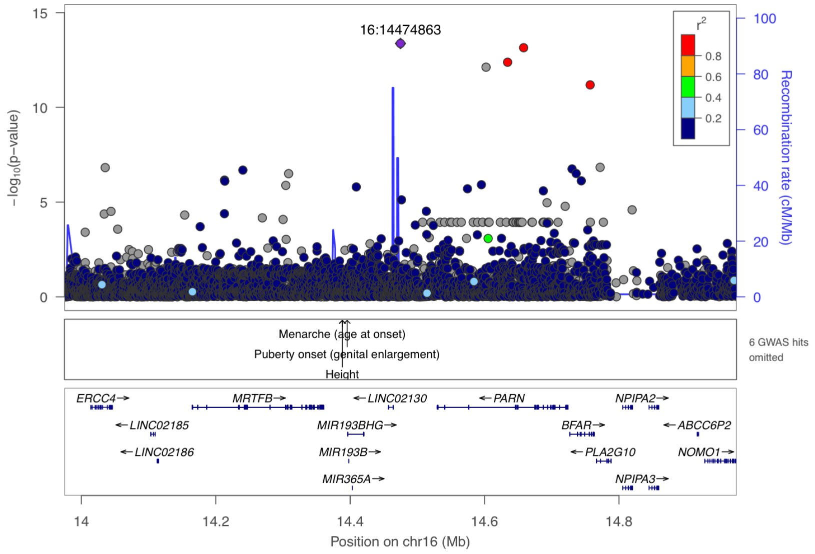


**Supplementary Figure 10-104. Visualization of the cis-pQTL association for protein PLA2G10.** The $r^2$ values are the squared linkage disequilibrium correlation coefficients between the plotted variants and the leading variant (purple dot). The middle panel marks the established associations at this locus according to the GWAS catalog ($p < 5 \times 10^{-8}$), and the bottom panel annotates the positions of the genes.

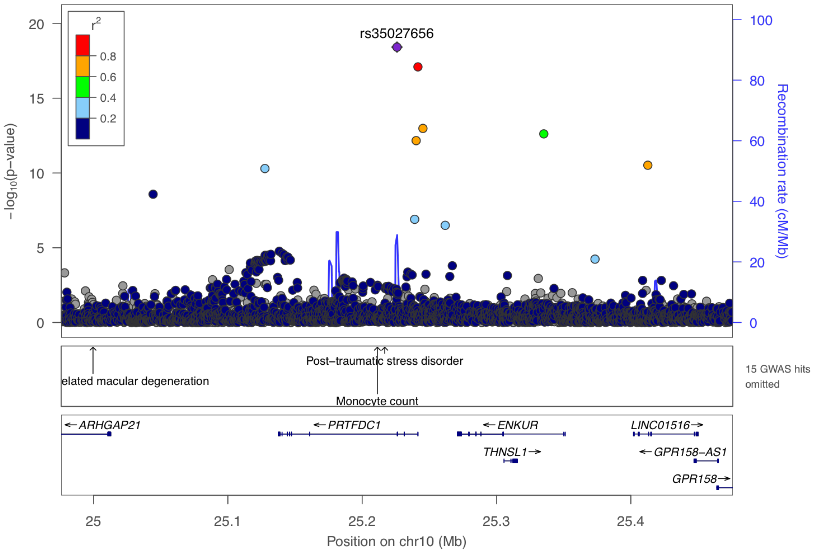


**Supplementary Figure 10-105. Visualization of the cis-pQTL association for protein PRTFDC1.** The $r^2$ values are the squared linkage disequilibrium correlation coefficients between the plotted variants and the leading variant (purple dot). The middle panel marks the established associations at this locus according to the GWAS catalog ($p < 5 \times 10^{-8}$), and the bottom panel annotates the positions of the genes.

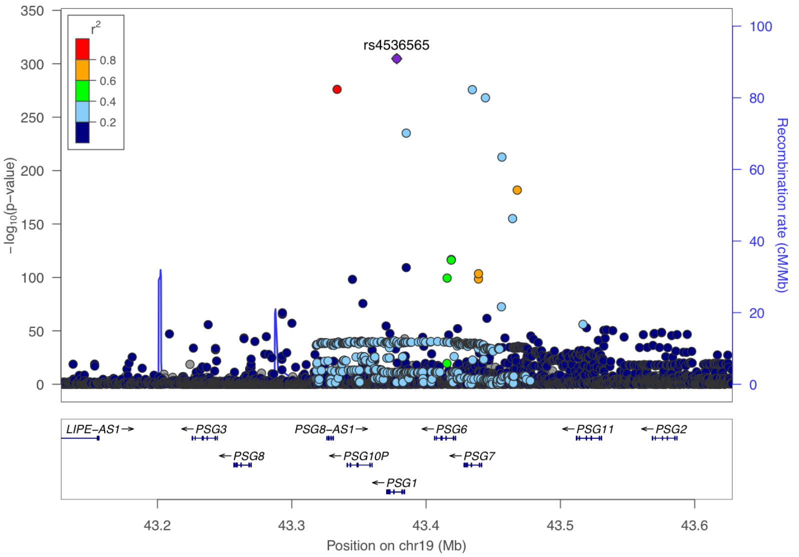


**Supplementary Figure 10-106. Visualization of the cis-pQTL association for protein PSG1.** The $r^2$ values are the squared linkage disequilibrium correlation coefficients between the plotted variants and the leading variant (purple dot). The middle panel marks the established associations at this locus according to the GWAS catalog ($p < 5 \times 10^{-8}$), and the bottom panel annotates the positions of the genes.

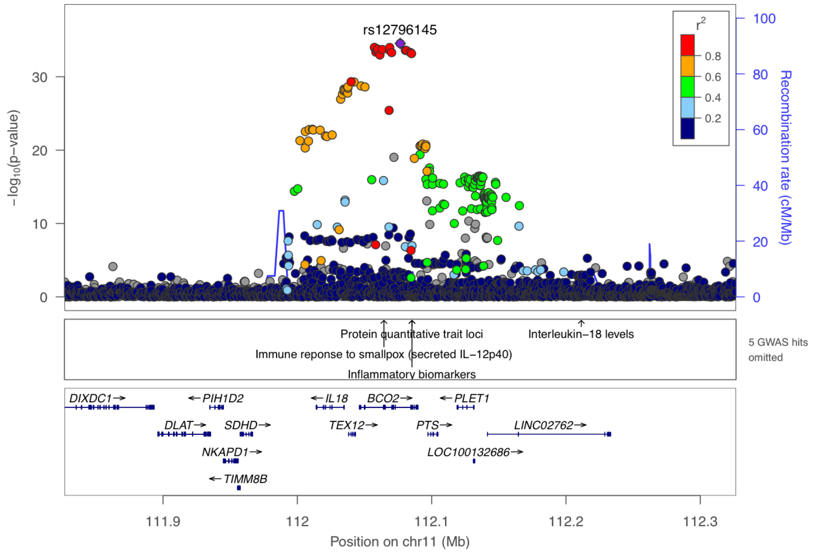


**Supplementary Figure 10-107. Visualization of the cis-pQTL association for protein PTS.** The $r^2$ values are the squared linkage disequilibrium correlation coefficients between the plotted variants and the leading variant (purple dot). The middle panel marks the established associations at this locus according to the GWAS catalog ($p < 5 \times 10^{-8}$), and the bottom panel annotates the positions of the genes.

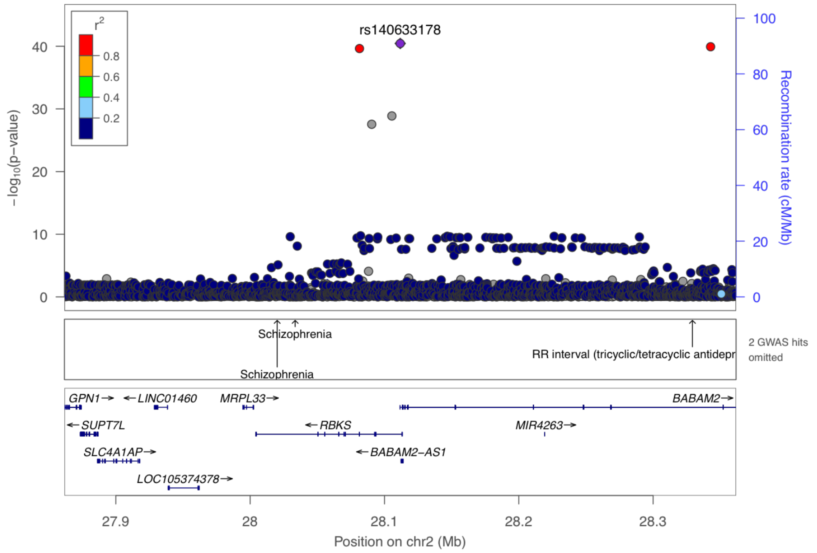


**Supplementary Figure 10-108. Visualization of the cis-pQTL association for protein RBKS.** The $r^2$ values are the squared linkage disequilibrium correlation coefficients between the plotted variants and the leading variant (purple dot). The middle panel marks the established associations at this locus according to the GWAS catalog ($p < 5 \times 10^{-8}$), and the bottom panel annotates the positions of the genes.

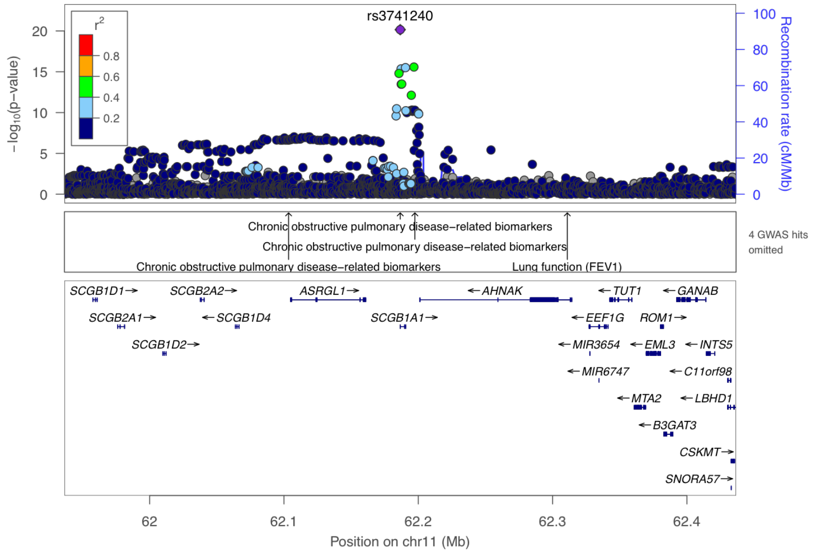


**Supplementary Figure 10-109. Visualization of the cis-pQTL association for protein SCGB1A1.** The $r^2$ values are the squared linkage disequilibrium correlation coefficients between the plotted variants and the leading variant (purple dot). The middle panel marks the established associations at this locus according to the GWAS catalog ($p < 5 \times 10^{-8}$), and the bottom panel annotates the positions of the genes.

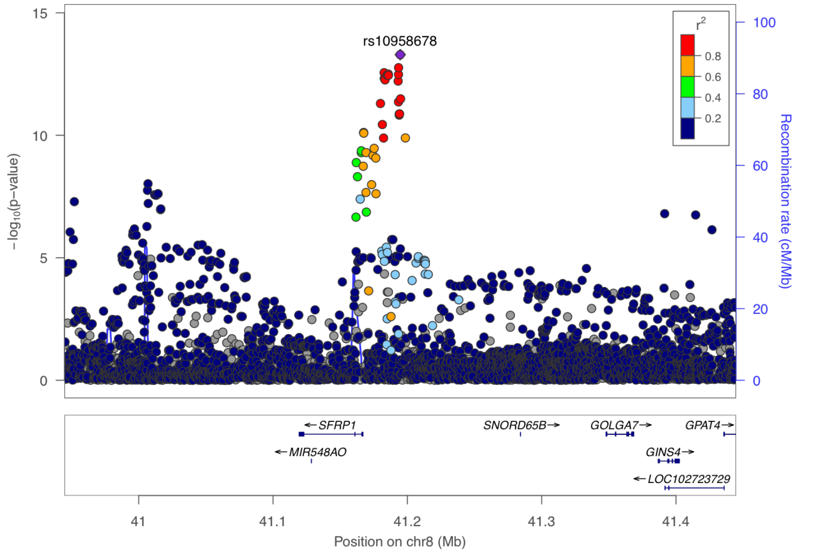


**Supplementary Figure 10-110. Visualization of the cis-pQTL association for protein SFRP1.** The $r^2$ values are the squared linkage disequilibrium correlation coefficients between the plotted variants and the leading variant (purple dot). The middle panel marks the established associations at this locus according to the GWAS catalog ($p < 5 \times 10^{-8}$), and the bottom panel annotates the positions of the genes.

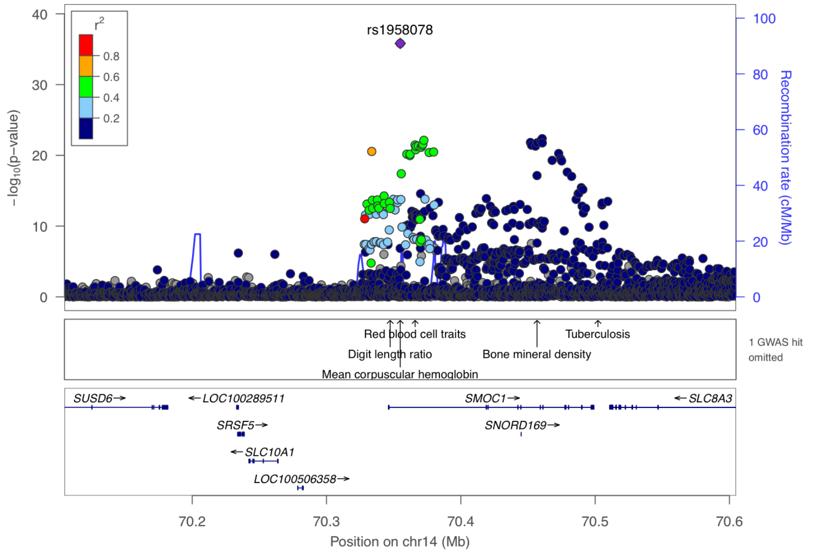


**Supplementary Figure 10-111. Visualization of the cis-pQTL association for protein SMOC1.** The $r^2$ values are the squared linkage disequilibrium correlation coefficients between the plotted variants and the leading variant (purple dot). The middle panel marks the established associations at this locus according to the GWAS catalog ($p < 5 \times 10^{-8}$), and the bottom panel annotates the positions of the genes.

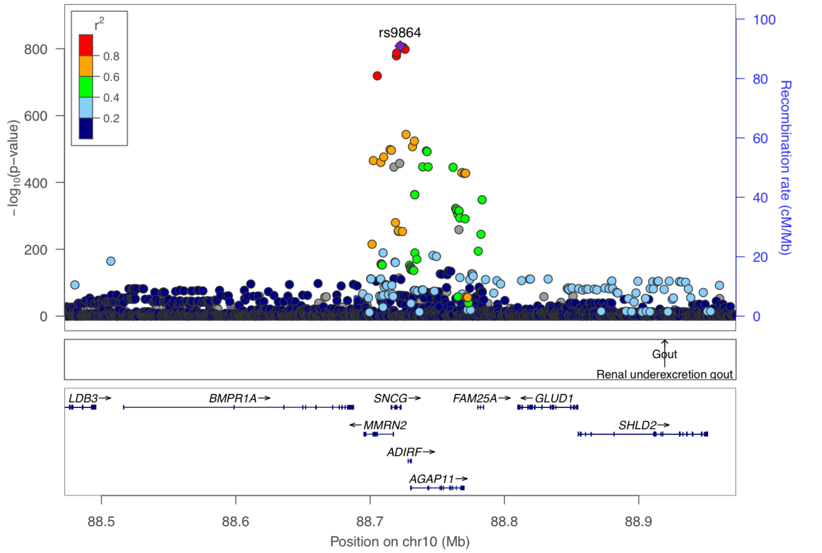


**Supplementary Figure 10-112. Visualization of the cis-pQTL association for protein SNCG.** The $r^2$ values are the squared linkage disequilibrium correlation coefficients between the plotted variants and the leading variant (purple dot). The middle panel marks the established associations at this locus according to the GWAS catalog ($p < 5 \times 10^{-8}$), and the bottom panel annotates the positions of the genes.

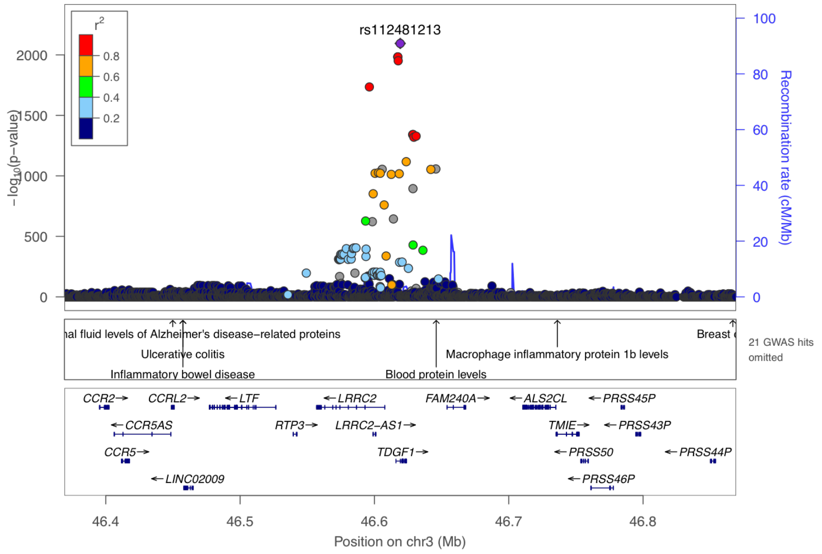


**Supplementary Figure 10-113. Visualization of the cis-pQTL association for protein TDGF1.** The $r^2$ values are the squared linkage disequilibrium correlation coefficients between the plotted variants and the leading variant (purple dot). The middle panel marks the established associations at this locus according to the GWAS catalog ($p < 5 \times 10^{-8}$), and the bottom panel annotates the positions of the genes.

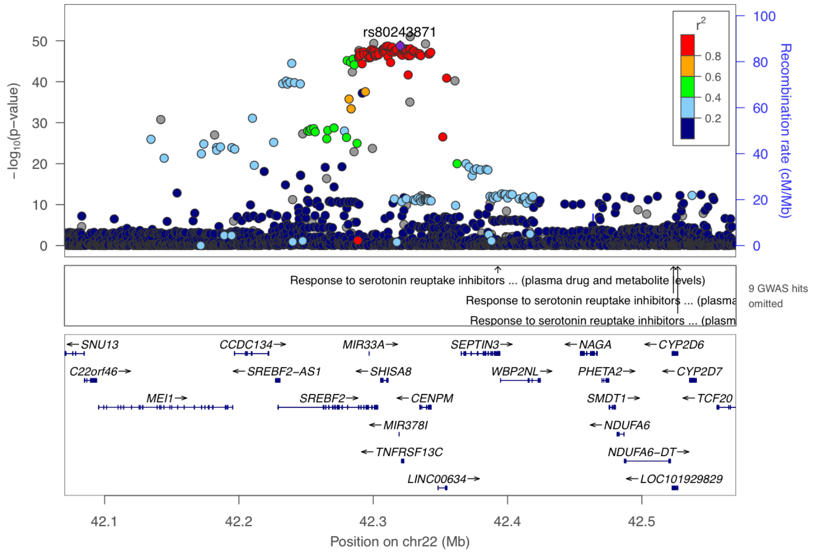


**Supplementary Figure 10-114. Visualization of the cis-pQTL association for protein TNFRSF13C.** The $r^2$ values are the squared linkage disequilibrium correlation coefficients between the plotted variants and the leading variant (purple dot). The middle panel marks the established associations at this locus according to the GWAS catalog ($p < 5 \times 10^{-8}$), and the bottom panel annotates the positions of the genes.

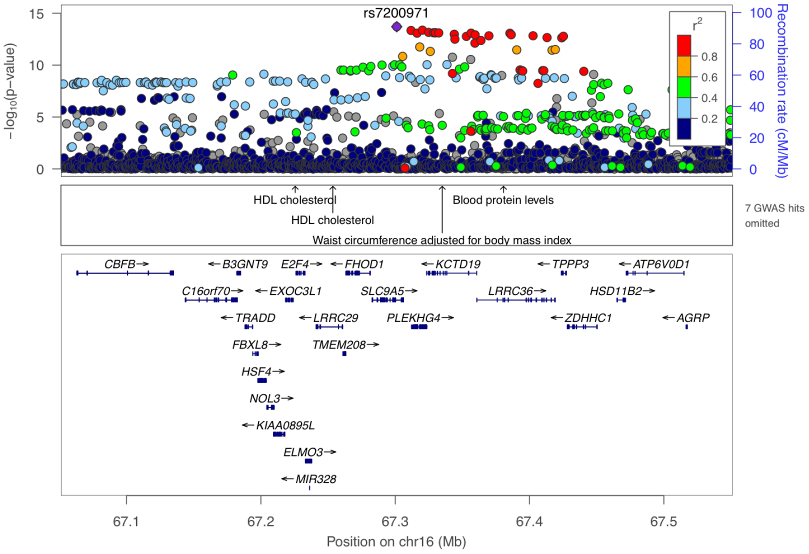


**Supplementary Figure 10-115. Visualization of the cis-pQTL association for protein TPPP3.** The $r^2$ values are the squared linkage disequilibrium correlation coefficients between the plotted variants and the leading variant (purple dot). The middle panel marks the established associations at this locus according to the GWAS catalog ($p < 5 \times 10^{-8}$), and the bottom panel annotates the positions of the genes.

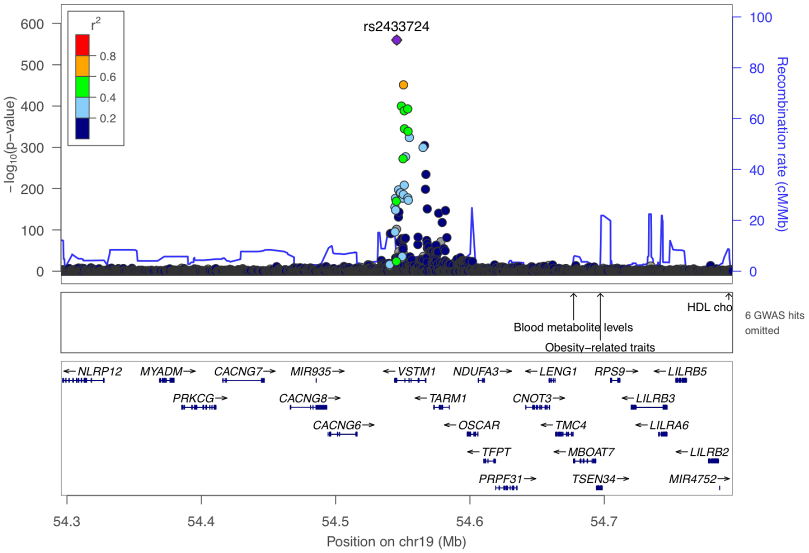


**Supplementary Figure 10-116. Visualization of the cis-pQTL association for protein VSTM1.** The $r^2$ values are the squared linkage disequilibrium correlation coefficients between the plotted variants and the leading variant (purple dot). The middle panel marks the established associations at this locus according to the GWAS catalog ($p < 5 \times 10^{-8}$), and the bottom panel annotates the positions of the genes.

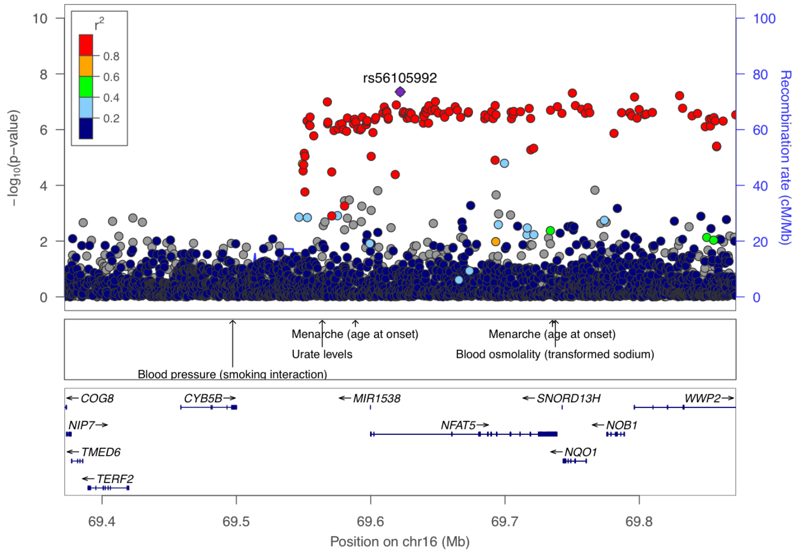


**Supplementary Figure 10-117. Visualization of the cis-pQTL association for protein WWP2.** The $r^2$ values are the squared linkage disequilibrium correlation coefficients between the plotted variants and the leading variant (purple dot). The middle panel marks the established associations at this locus according to the GWAS catalog ($p < 5 \times 10^{-8}$), and the bottom panel annotates the positions of the genes.
